# Genomic and phenomic landscape of clonal hematopoiesis in over a million ancestrally diverse participants

**DOI:** 10.1101/2022.07.29.22278015

**Authors:** Md Mesbah Uddin, Zhi Yu, Joshua S. Weinstock, Abhishek Niroula, Caitlyn Vlasschaert, Satoshi Koyama, Tetsushi Nakao, Sarah M. Urbut, Seyedeh M. Zekavat, Kaavya Paruchuri, Buu Truong, Alexander J. Silver, Taralynn M. Mack, Megan Y. Wong, Sara M. Haidermota, Romit Bhattacharya, Saman Doroodgar Jorshery, Michael A. Raddatz, Michael C. Honigberg, Whitney E. Hornsby, Martin Jinye Zhang, Jacqueline Dron, Jonathan Brett Heimlich, Vijay G. Sankaran, Gabriel K. Griffin, Christopher J. Gibson, Hailey A. Adegboye, Kelly Cho, Yan V. Sun, Saiju Pyarajan, Peter W.F. Wilson, Veterans Affairs’ Million Veteran Program, Giulio Genovese, Yaomin Xu, Michael R. Savona, Yii-Der Ida Chen, Wayne H-H Sheu, Yi-Jen Hung, Laura M. Raffield, Bertha A. Hidalgo, Myriam Fornage, Robert C. Kaplan, Keoki L. Williams, Donna K Arnett, Mike Province, Barbara A Konkle, Sharon Kardia, Patricia A. Peyser, Jennifer A. Smith, Rasika A. Mathias, Lisa R. Yanek, Lewis C. Becker, Mariza de Andrade, Dawood Darbar, Jiang He, John Blangero, Scott T. Weiss, Brian Custer, Deborah A. Meyers, Ryan L. Minster, Take Naseri, Satupaitea Viali, Braxton D. Mitchell, Kathleen C. Barnes, Susan Redline, David Schwartz, Benjamin M. Shoemaker, Nicholette D. Palmer, Barry I. Freedman, Donald W. Bowden, Christine M. Albert, Rajesh Kumar, Nicholas Smith, James Floyd, Russell Tracy, Joshua C. Bis, Bruce M. Psaty, Kent D. Taylor, Stephen S. Rich, Jerome I. Rotter, Eric Boerwinkle, Vasan S. Ramachandran, Steven Lubitz, Patrick T. Ellinor, Zhe Wang, Ruth J.F. Loos, Jeong H. Yun, Michael H. Cho, Edwin K. Silverman, Goncalo Abecasis, Charles Kooperberg, Paul L. Auer, Alexander P. Reiner, Siddhartha Jaiswal, Benjamin L. Ebert, NHLBI Trans-Omics for Precision Medicine (TOPMed) Consortium, Alexander G. Bick, Pradeep Natarajan

**Affiliations:** Program in Medical and Population Genetics and Cardiovascular Disease Initiative, Broad Institute of MIT and Harvard, Cambridge, MA 02142, USA; Heart and Vascular Institute, Mass General Brigham; Center for Genomic Medicine, Massachusetts General Hospital, Boston, MA 02114, USA; Department of Biostatistics, University of Michigan School of Public Health, Ann Arbor, MI, USA; Department of Medical Oncology, Dana-Farber Cancer Institute, Boston, MA 02115, USA; Broad Institute of MIT and Harvard, Cambridge, MA 02142, USA; Department of Laboratory Medicine, Lund University, Lund, Sweden; Department of Medicine, Queens University, Kingston, Ontario, Canada; Division of Cardiovascular Medicine, Department of Medicine, Brigham and Women’s Hospital, Boston, MA 02115, USA; Cardiology Division, Department of Medicine, Massachusetts General Hospital, Boston, MA 02114, USA; Computational Biology & Bioinformatics Program, Yale University, New Haven, CT 06510, USA; Department of Medicine, Harvard Medical School, Boston, MA, USA; Vanderbilt University School of Medicine, Nashville, TN, USA; Division of Genetic Medicine, Department of Medicine, Vanderbilt University Medical Center, Nashville, TN, USA; Program in Medical and Population Genetics, Broad Institute of MIT and Harvard, Cambridge, MA, USA; Department of Computer Science, University of Toronto, Toronto, Canada; Department of Computer Science and Electrical Engineering, Massachusetts Institute of Technology, Cambridge, MA, USA; Center for Genomic Medicine, Massachusetts General Hospital, Boston, MA 02114, USA; Department of Epidemiology, Harvard T.H. Chan School of Public Health, Boston, MA, USA; Division of Hematology/Oncology, Boston Children’s Hospital, Harvard Medical School, Boston, USA; Department of Pediatric Oncology, Dana-Farber Cancer Institute, Harvard Medical School, Boston, USA; Harvard Stem Cell Institute, Cambridge, MA USA; Department of Pathology, Dana-Farber Cancer Institute, Boston, MA 02215, USA; Division of Hematopathology, Department of Pathology, Brigham and Women’s Hospital, Boston MA 02115, USA; Epigenomics Program, Broad Institute of MIT and Harvard, Cambridge, MA, USA; Veterans Affairs Boston Healthcare System, Boston, MA, USA; Department of Medicine, Brigham and Women’s Hospital, Harvard Medical School, Boston, MA, USA; Veterans Affairs Atlanta Healthcare Systems, Decatur, GA, USA; Department of Epidemiology, Emory University Rollins School of Public Health, Atlanta, GA, USA; Department of Medicine, Emory University School of Medicine, Atlanta, GA, USA; A full list of MVP authors appears in the Supplementary Note; Stanley Center for Psychiatric Research, Broad Institute of MIT and Harvard, Cambridge, MA 02142, USA; Department of Genetics, Harvard Medical School, Boston, MA 02115, USA; Department of Biostatistics, Vanderbilt University Medical Center, Nashville, TN, USA; Department of Biomedical Informatics, Vanderbilt University Medical Center, Nashville, TN, USA; The Institute for Translational Genomics and Population Sciences, Department of Pediatrics, The Lundquist Institute for Biomedical Innovation at Harbor-UCLA Medical Center, Torrance, CA 90502, USA; Institute of Molecular and Genomic medicine, National Health Research Institutes, Zhunan, Taiwan; Division of Endocrinology and Metabolism, Department of Internal Medicine, Taichung Veterans General Hospital, Taichung, Taiwan; Division of Endocrinology and Metabolism, Department of Internal Medicine, Taipei Veterans General Hospital, Taipei, Taiwan; College of Medicine, National Defense Medical University, Taipei, Taiwan; Division of Endocrinology & Metabolism, Tri-Service General Hospital, National Medical Center, New Taipei City, 237010 Taiwan; University of North Carolina at Chapel Hill, Chapel Hill, NC 27599, USA; University of Alabama at Birmingham, Birmingham, AL 35216, USA; University of Texas Health Science Center at Houston, Houston, TX 77030, USA; Albert Einstein College of Medicine, New York, NY 10461, USA; Henry Ford Health System, Detroit, MI 48202, USA; University of South Carolina, Columbia, SC 29208, USA; Washington University School of Medicine in St. Louis, St. Louis, MO 63110, USA; Washington Center for Bleeding Disorders, Seattle, WA 98101, USA; University of Michigan, Ann Arbor, MI 48109, USA; National Institutes of Health, Rockville, MD 20852, USA; Johns Hopkins University School of Medicine, Baltimore, MD 21205, USA; Mayo Clinic, Rochester, MN 55905, USA; University of Illinois at Chicago, Chicago, IL 60607, USA; Tulane University, New Orleans, LA 70118, USA; University of Texas Rio Grande Valley School of Medicine, Brownsville, TX 78520, USA; Harvard Medical School, Boston, MA 02115, USA; Vitalant Research Institute, San Francisco, CA 94118, USA; University of Arizona, Tucson, AZ 85721, USA; University of Pittsburgh, Pittsburgh, PA 15213, USA; Naseri and Associates Public Health Consultancy Firm and Family Health Clinic, Apia, Samoa; Oceania University of Medicine, Apia, Samoa; University of Maryland School of Medicine, Baltimore, MD 21201, USA; University of Colorado Anschutz Medical Campus, Aurora, CO 80045, USA; Brigham & Women’s Hospital, Boston, MA 02115, USA; Vanderbilt University, Nashville, TN 37235, USA; Wake Forest University School of Medicine, Winston-Salem, NC 27157, USA; Department of Cardiology, Smidt Heart Institute, Cedars–Sinai Medical Center, Los Angeles, CA, USA; Ann and Robert H. Lurie Children’s Hospital of Chicago, Chicago, IL 60611, USA; University of Washington, Seattle WA 98195, USA; University of Vermont, Burlington, VT 05405, USA; Cardiovascular Health Research Unit, Departments of Medicine & Epidemiology, University of Washington, Seattle WA 98195, USA; University of Virginia, Charlottesville, VA 22908, USA; Boston University, Boston, MA 02118, USA; Mass General Brigham, Boston, MA 02114, USA; University of Alabama at Birmingham, Birmingham, AL 35294, USA; Icahn School of Medicine at Mount Sinai, New York, NY 10029, USA; Fred Hutchinson Cancer Research Center, Seattle, WA 98109, USA; Medical College of Wisconsin, Milwaukee, WI, USA; Department of Epidemiology, University of Washington, Seattle, WA 98195, USA; Department of Pathology, Stanford University School of Medicine, Stanford, CA, USA; Institute for Stem Cell Biology and Regenerative Medicine, Stanford University School of Medicine, Stanford, CA, USA; Howard Hughes Medical Institute, Boston, MA, USA; A full list of TOPMed authors appears in the Supplementary Note

## Abstract

With aging, somatic mutations in hematopoietic stem and progenitor cells (HSPC) can give rise to clonal hematopoiesis of indeterminate potential (CHIP), a premalignant state associated with diverse age-related diseases. Here we report the largest multi-ancestry genome-wide analysis of CHIP to date (N = 1,018,305), including individuals of African (N = 85,978), Admixed American (N = 191,371), East Asian (N = 13,532), European (N = 694,015), and South Asian (N = 13,193) ancestry. Multi-ancestry meta-analyses identified 72 genome-wide significant loci, including 44 novel associations implicating genes such as *AFF1*, *ATF7IP*, *ATP8B4*, *BCL2*, *CEBPA*, *CYRIA*, *DNM2*, *ELF1*, *NKX2-3*, *PIK3CB*, *PRDM16*, *RPN1*, *TERC*, and *TRIM4*. Notably, variants at *MECOM* and *PHF20L1* showed opposite allelic effects between *DNMT3A*- and non-*DNMT3A*-driven CHIP, highlighting driver-specific germline influences. Implicated loci converge on pathways regulating telomere maintenance, cell-cycle control, hematopoietic transcription, DNA damage response and immune signaling. These findings support a model in which germline variation both expands the HSPC pool and biases clonal selection in a driver-dependent manner, providing a mechanistic basis for inter-individual heterogeneity in CHIP. Phenome-wide association analyses further linked CHIP to hematologic, neoplastic, and circulatory traits, with enrichment across hematopoietic and non-hematopoietic cell types. Together, this work expands the genomic and phenomic landscape of CHIP and reveals germline–somatic interactions that shape clonal evolution during aging.

## Introduction

Self-renewing cell populations accumulate somatic mutations with aging, although most of these mutations are without functional consequence. Some of these somatic mutations confer a selective advantage leading to clonal expansion^1–3^. In the case of hematopoietic stem cells, driver mutations in genes with diverse functions, including DNA methylation (*DNMT3A*, *TET2*)^4,5^, splicing factors (*SF3B1*, *SRSF2*, *U2AF1, ZRSF2*)^6^, chromatin remodeling (*ASXL1*)^7^, and DNA damage response (DDR; *TP53*, *PPM1D*)^8–10^ can lead to a clonal expansion of hematopoietic stem cells termed clonal hematopoiesis of indeterminate potential (CHIP) when such mutations make up >4% of peripheral blood cells (variant allele fraction, VAF≥2%). CHIP is the pre-cancerous precursor lesion for myeloid hematologic malignancy^11–13^. Numerous studies in human and model systems have linked CHIP to diverse diseases of aging, including coronary artery disease^14,15^, stroke^16^, heart failure^17^, chronic obstructive pulmonary disease^18^, osteoporosis^19^, and chronic liver disease^20^. Intriguingly, CHIP is less frequently observed among patients with Alzheimer’s disease^21^. Characterizing the germline genetic determinants of CHIP offers the opportunity to prioritize unifying features of multiple diseases of aging.

We previously performed a genome-wide association study (GWAS) of CHIP in a study of 3,831 CHIP cases and 61,574 controls from whole-genome sequencing of blood DNA utilizing the National Heart, Lung and Blood Institute (NHLBI) Trans-Omics for Precision Medicine (TOPMed) study^22^. This effort identified three germline genetic risk loci, including one in the *TERT* promoter, the intronic region of *TRIM59*, and a distal enhancer of *TET2* specific to individuals of African ancestry. These findings enabled further work demonstrating that the low-frequency germline variant at *TET2* leads to locally altered methylation and decreased germline *TET2* expression, subsequently promoting the self-renewal and proliferation of hematopoietic stem cells^22^. A subsequent analysis of 200,453 UK Biobank participants expanded the landscape by identifying ten additional risk regions and implicating genes including *PARP1*, *CD164*, *ATM*, *TCL1A*, *SETBP1*, and *CHEK2* in CHIP susceptibility^23^. More recently, three additional studies have refined our understanding of CHIP’s genetic architecture: one large-scale exome sequencing study combining data from 454,803 UK Biobank (UKB) and 173,585 Geisinger participants detected 24 risk loci spanning both common and rare variants^24^; a comparative analysis of the Mexico City Prospective Study (MCPS) with the UKB revealed pronounced ancestry-specific effects—highlighting, for example, novel *TCL1B* risk variants that exert opposing influences on *DNMT3A*- versus *TET2*-driven CHIP^25^; and bidirectional Mendelian randomization analyses have further underscored a dynamic interplay among leukocyte telomere length, *TERT* locus variants, and CHIP across the lifespan^26^. Here we present the largest genome-wide association study of CHIP to date, substantially expanding the genomic and phenomic landscape of clonal hematopoiesis through analysis of over one million ancestrally diverse participants, incorporating newly generated CHIP calls.

Here, we meta-analyzed GWAS from six diverse cohorts for CHIP traits, including 53,274 CHIP cases and 922,528 controls, including 33% individuals of non-European ancestry (**Extended Data Table 1** and **Supplementary Data Table 1**). Using this robust dataset, we perform combined and ancestry-, sex-, and gene-stratified genome-wide discovery analyses, as well as conduct fine-mapping at variant^27^ and gene levels^28^. We also performed multivariate Bayesian analyses^29^ with myeloproliferative neoplasm (MPN)^30^, leukocyte telomere length (LTL)^31^, and expanded mosaic chromosomal alterations (mCAs)^32^-associated alleles to prioritize additional loci for CHIP. Finally, we leverage this large dataset to systematically profile the disease risk of CHIP compared with another form of clonal hematopoiesis—mCAs and further investigate the disease associations by examining cell types^33^ enriched by CHIP (**Extended Data Fig. 1**).

## Results

### Baseline characteristics

We analyzed a total of 1,018,305 individuals across six large-scale cohorts—UKB (N=454,327), All of Us Research Program (AoU; N=245,388), TOPMed (N=74,622), Vanderbilt University Biobank (BioVU; N=54,583), Mass General Brigham Biobank (MGBB; N=52,984), and MCPS (N=136,401)^25^ (see **Methods**). Participants were European (EUR; N=694,015), African (AFR; N=85,978), Admixed American (AMR; N=191,371), East Asian (EAS; N=13,532), and South Asian (SAS; N=13,193) genetic ancestry (**Supplementary Data Table 1**). Across these cohorts, we identified 61,702 CHIP mutations in 55,858 individuals. The overall prevalence of CHIP varied across cohorts, with rates of 6.5% in UKB, 3.9% in AoU, 6.3% in TOPMed, 11.1% in MGBB, 2.9% in BioVU, and 3.1% in MCPS (**Extended Data Table 1**).

CHIP prevalence increased with age (**Fig. 1a; Extended Data Fig. 2a**), and the majority of affected participants (>80%) carried a single CHIP clone (**Fig. 1b; Extended Data Fig. 2b**). The most frequently mutated genes included *DNMT3A*, *TET2*, *ASXL1*, *PPM1D*, *TP53,* and *SF3B1* (**Fig. 1c; Extended Data Fig. 2c**), in line with previous reports^22,23^. CHIP clone size, as measured by variant allele fraction (VAF), was generally larger in TOPMed and AoU than in UKB and MGBB (**Fig. 1d; Extended Data Fig. 2d**), which likely reflected the differences in detection sensitivity between whole-genome sequencing (used in TOPMed and AoU) and whole-exome sequencing (used in UKB and MGBB).

**Fig. 1.**
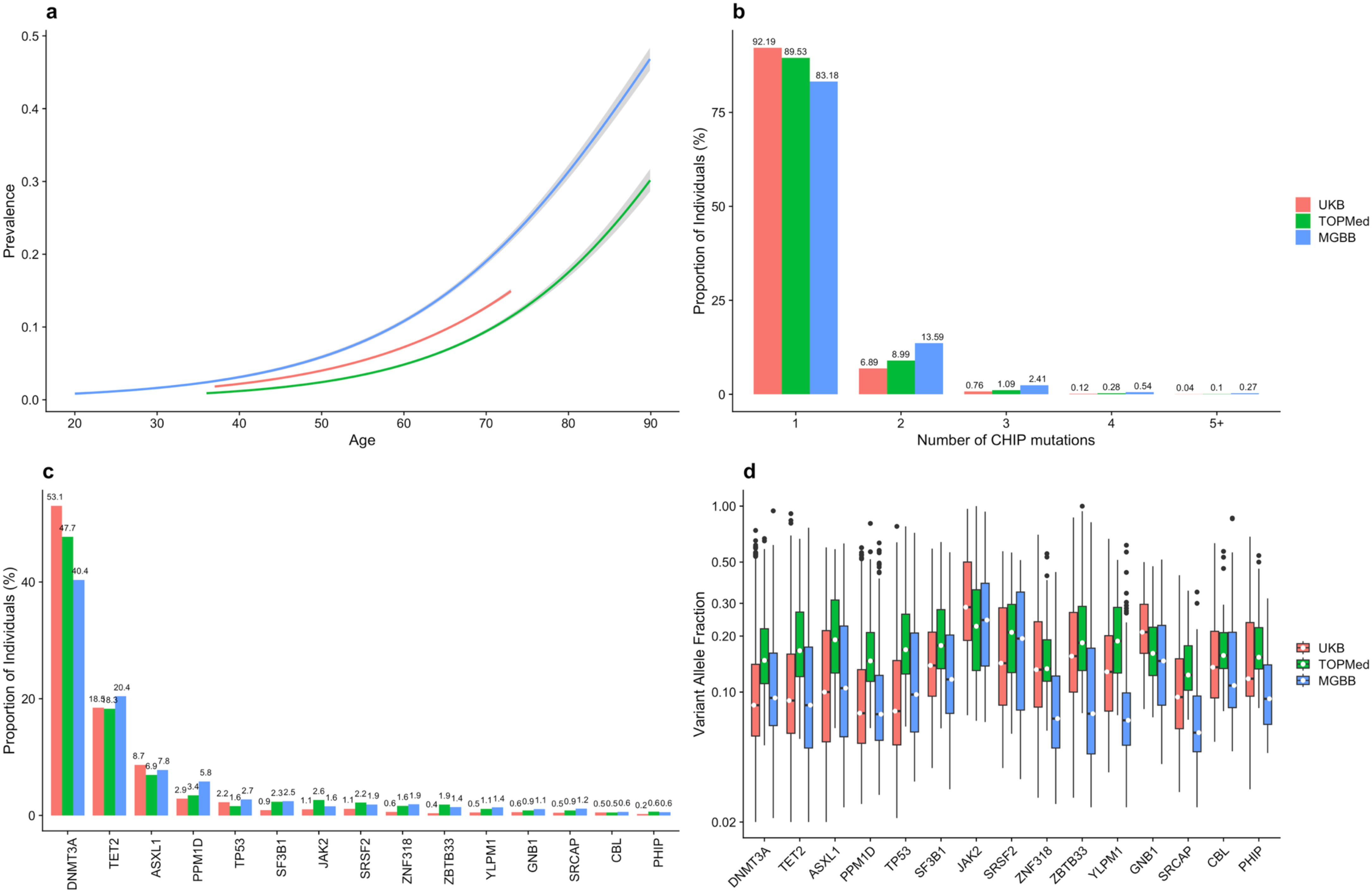
Distribution of CHIP in UKB, TOPMed and MGBB cohort. **a**, CHIP prevalence increased with the donor’s age at the time of blood sampling. The center line represents the general additive model spline, and the shaded region is the 95% confidence interval (N_UKB_=454,327 WES; N_TOPMed_=74,622 WGS; N_MGBB_=52,984 WES). **b**, More than 90% of individuals with CHIP had only one somatic CHIP driver mutation variant identified. **c**, Proportion of individuals carrying a driver mutation in the ten most frequently mutated genes in CHIP. **d**, There was heterogeneity in CHIP clone size as measured by variant allele fraction by CHIP driver gene. Boxplot spanning minimum and maximum values with median variant allele fraction highlighted by white circles. CHIP distribution in the All of Us cohort is presented in **Extended Data** Fig. 2. CHIP: clonal hematopoiesis of indeterminate potential; UKB: UK Biobank; TOPMed: Trans-omics for Precision Medicine; MGBB: Mass General Brigham Biobank; WES: whole-exome samples; WGS: whole-genome samples.

Age at blood draw was strongly associated with a higher prevalence of CHIP across all cohorts (**Fig. 2a; Supplementary Data Table 2**) in multivariable-adjusted logistic regression models (**Methods**). While overall CHIP prevalence did not significantly differ between males and females, males had higher odds of late arising CHIP clones, particularly in *ASXL1*, splicing factors genes, and DDR genes. In contrast, females had a significantly higher prevalence of *DNMT3A* CHIP across all cohorts (**Fig. 2b; Supplementary Data Table 2**). Smoking history (ever smoker or ≥100 cigarettes lifetime) was associated with a higher prevalence of CHIP and CHIP subtypes, with the notable exception of *TET2*, which remained unaffected by smoking status (**Fig. 2c; Supplementary Data Table 2**). CHIP prevalence did not differ significantly between individuals of AFR, AMR, SAS, or EAS ancestry compared to those of EUR ancestry (**Fig. 2d; Supplementary Data Table 2**). These findings comprehensively characterize CHIP across multiple cohorts, highlighting the role of key demographics in CHIP prevalence.

**Fig. 2.**
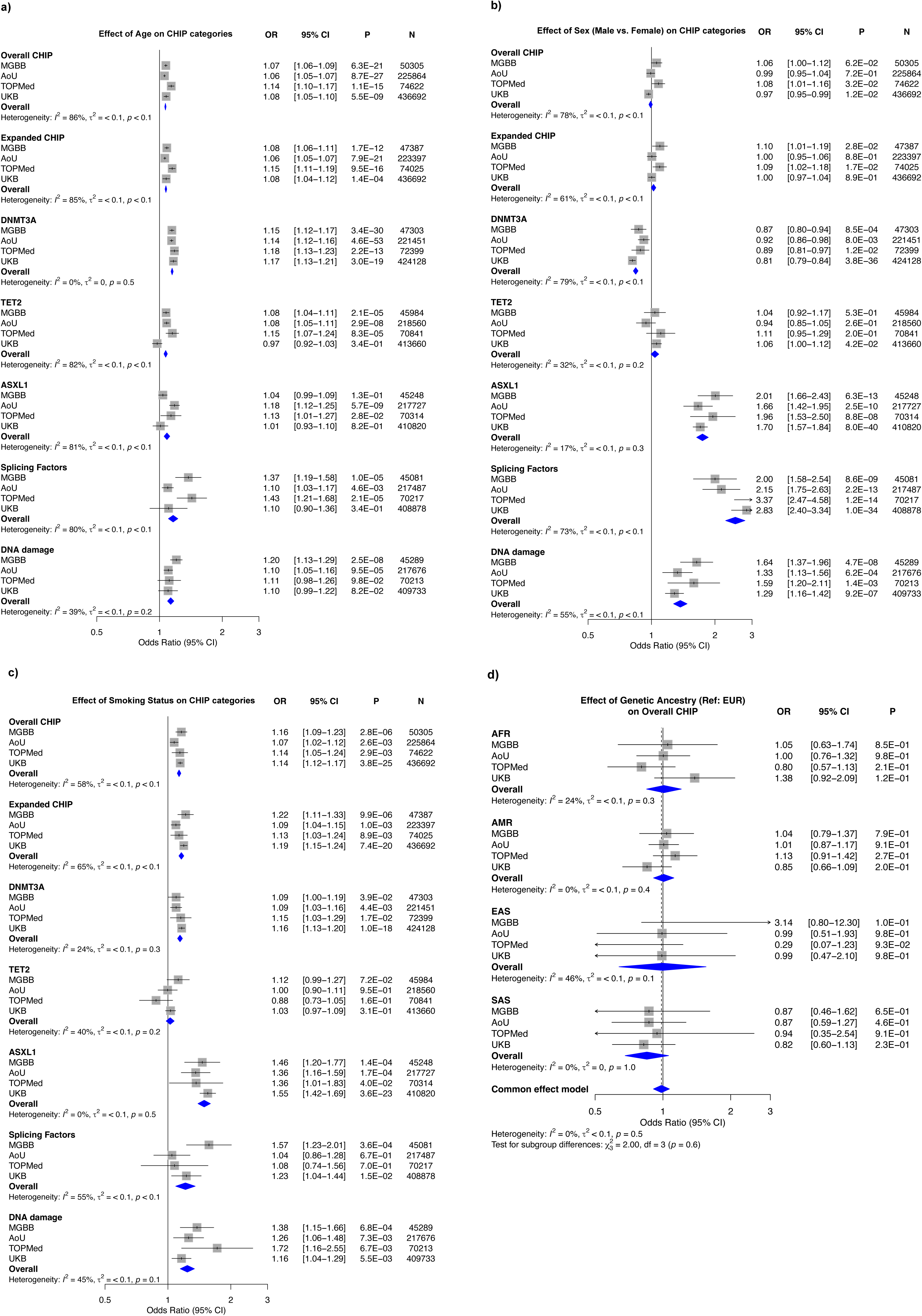
Association of baseline risk factors with CHIP traits. Forest plots showing odds ratio (OR) with a 95% confidence interval (CI) from multivariable logistic regression analyses adjusted for age, age^2^, genetic sex, genetic ancestry, smoking status, first five genetic principal components, and cohort-specific batch effects (Method). a) Age vs. CHIP traits: age is strongly associated with CHIP prevalence. b) Genetic sex vs. CHIP traits: males have lower odds of *DNMT3A* CHIP than females while showing higher odds of *ASXL1*, SF, and DDR CHIP. c) Smoking status vs. CHIP traits: individuals with a smoking history have higher odds of CHIP, and the effect is stronger for *ASXL1*. d) Genetic ancestry vs. overall CHIP prevalence in MGBB (N=45,289), AoU (N=217,676), TOPMed (N=70,213), and UKB (N=409,733): no significant differences between overall CHIP prevalence in AFR, AMR, SAS, and EAS compared to participants with EUR genetic ancestry. DNA damage response (DDR): includes CHIP genes *PPM1D* and/or TP53; Splicing factors [SF]: includes CHIP genes *SF3B1*/*SRSF2*/*U2AF1*/*ZRSR2*; MGBB: Mass General Brigham Biobank; AoU: All of US Research; TOPMed: Trans-omics for Precision Medicine; UKB: UK Biobank.

### Meta-analysis of GWAS identifies novel loci for CHIP traits

We performed multi-ancestry inverse-variance weighted fixed effect meta-analyses of overall CHIP across six cohorts, comprising 53,274 cases and 922,528 controls (N=975,802; **Extended Data Table 1**). We observed no evidence of genomic inflation from population structure (*λ*_GC_ ranging from 1.04 to 1.07; **Extended Data Table 2**). GWAS results are summarized in **Fig. 3a-d**, **Extended Data Figs 3-9**, and detailed significant variants (P<5×10^-8^) in **Supplementary Data Tables 3-17**. We estimated the SNP heritability (*h^2^_SNP_*) of overall and expanded CHIP (i.e., VAF>=10%) to be 9.9% (SD=0.5%) and 11.3% (0.7%) on the liability scale, respectively, using the BLD-LDAK model (see **Methods** and **Extended Data Table 3**). Estimated *h^2^_SNP_* was notably high for gene-specific CHIP subtypes, including *DNMT3A* (13.9%), *TET2* (12.3%), *ASXL1* (12.6%), and DDR genes (21.0%) (**Extended Data Table 3**). Partitioning of *h^2^_SNP_* across functional annotations revealed significant enrichment in promoters and regulatory regions, highlighting a strong role for gene regulation in CHIP predisposition (**Fig. 3e; Extended Data Fig. 10**).

**Fig. 3.**
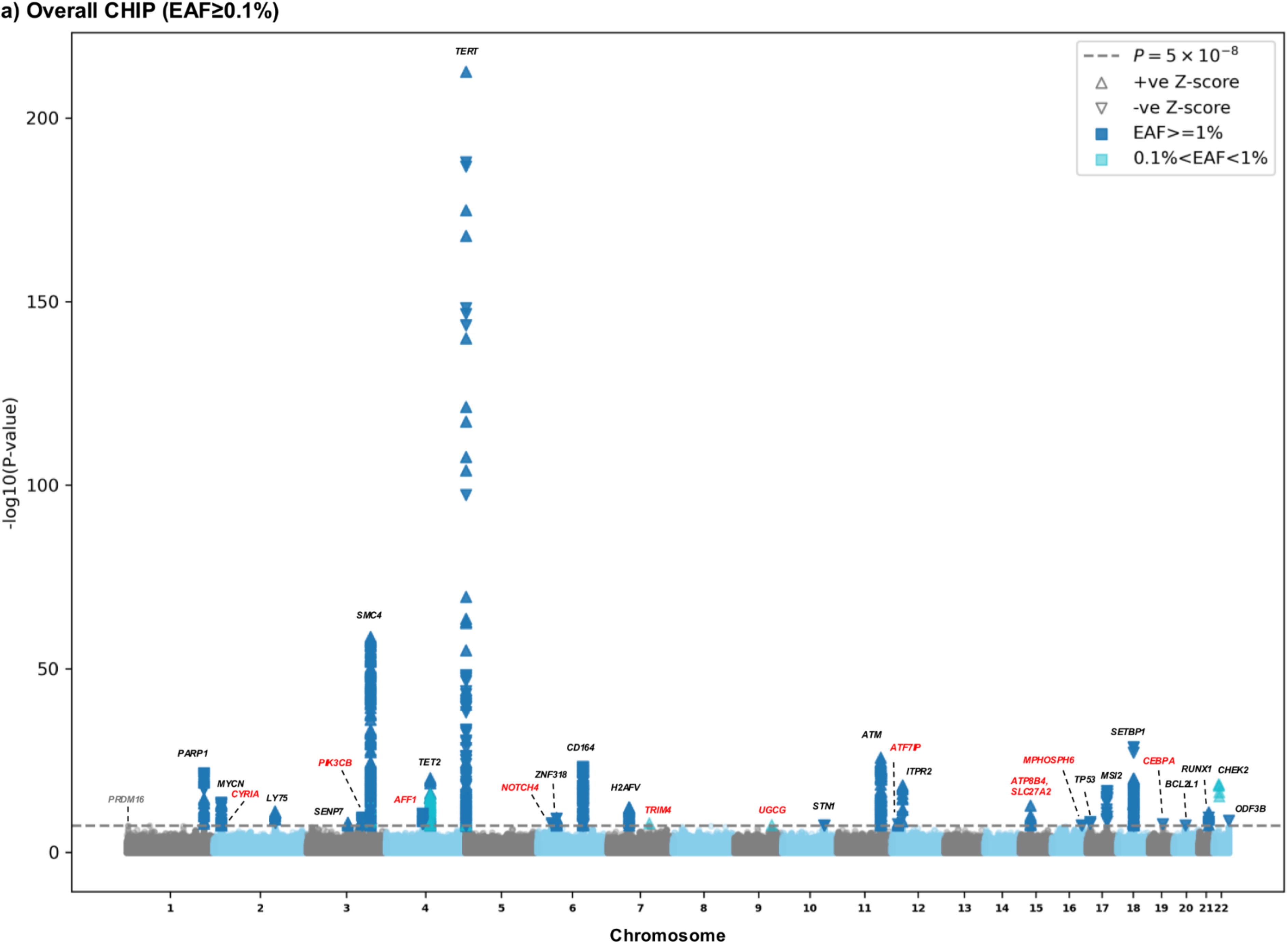
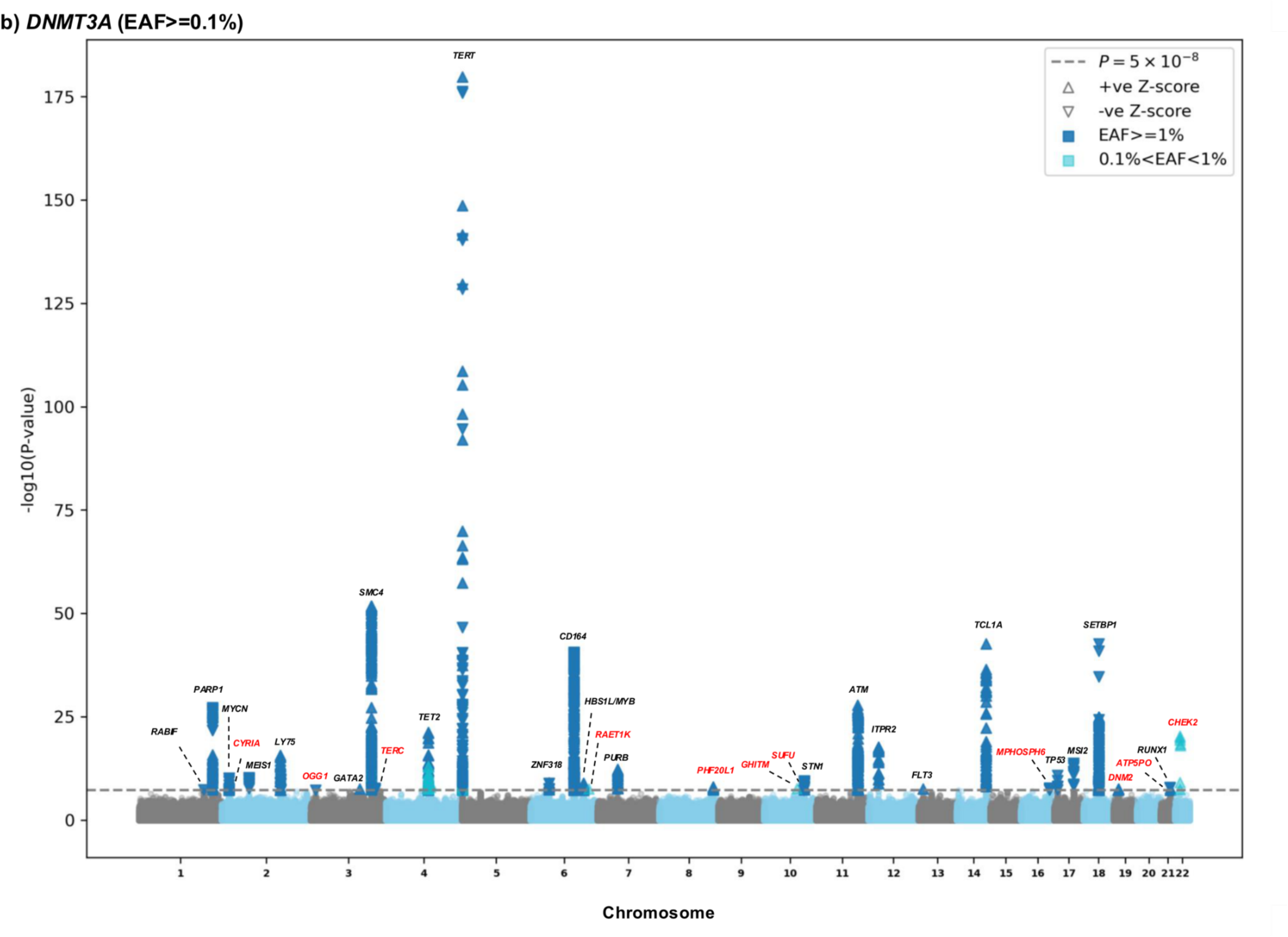
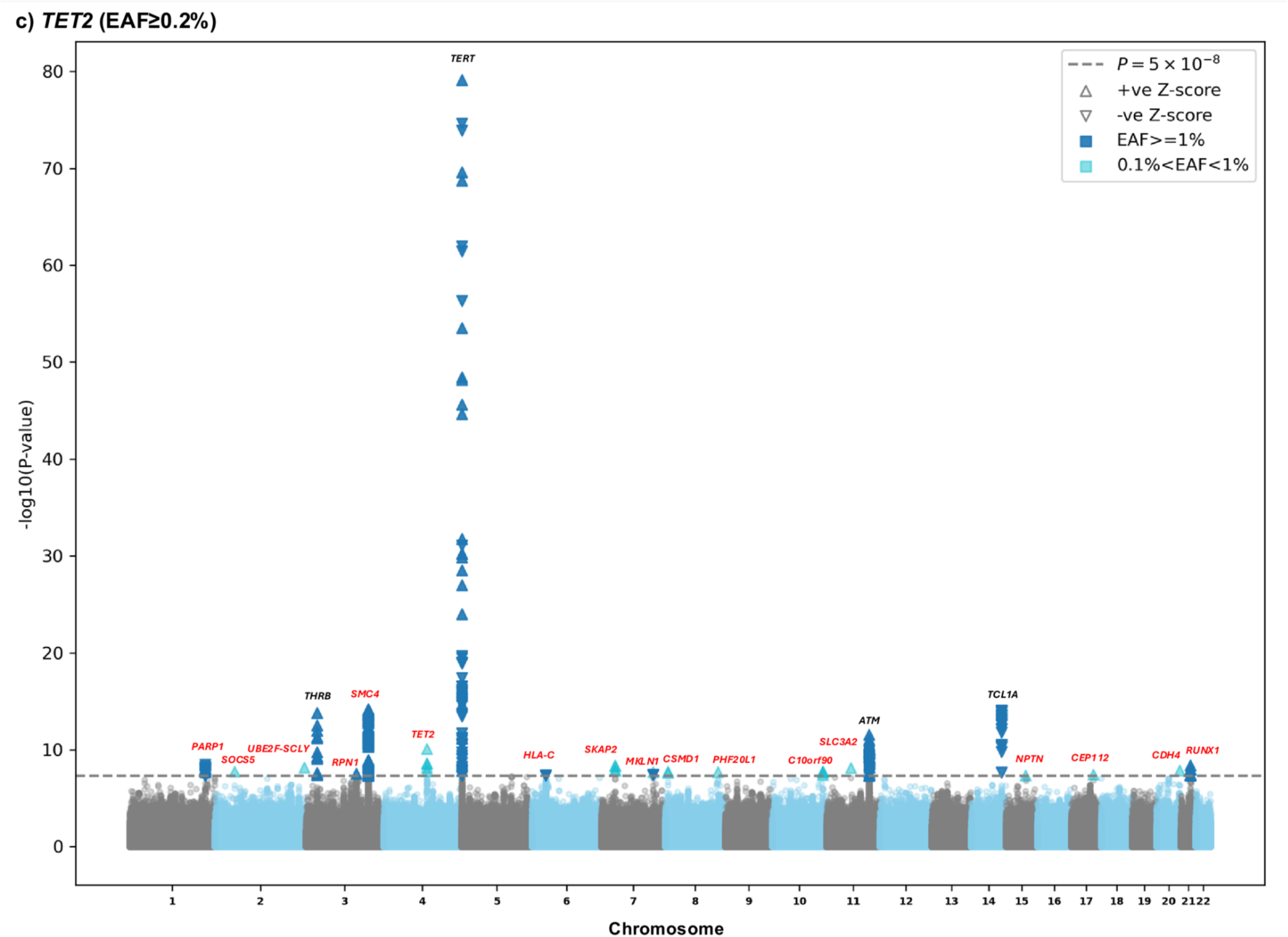
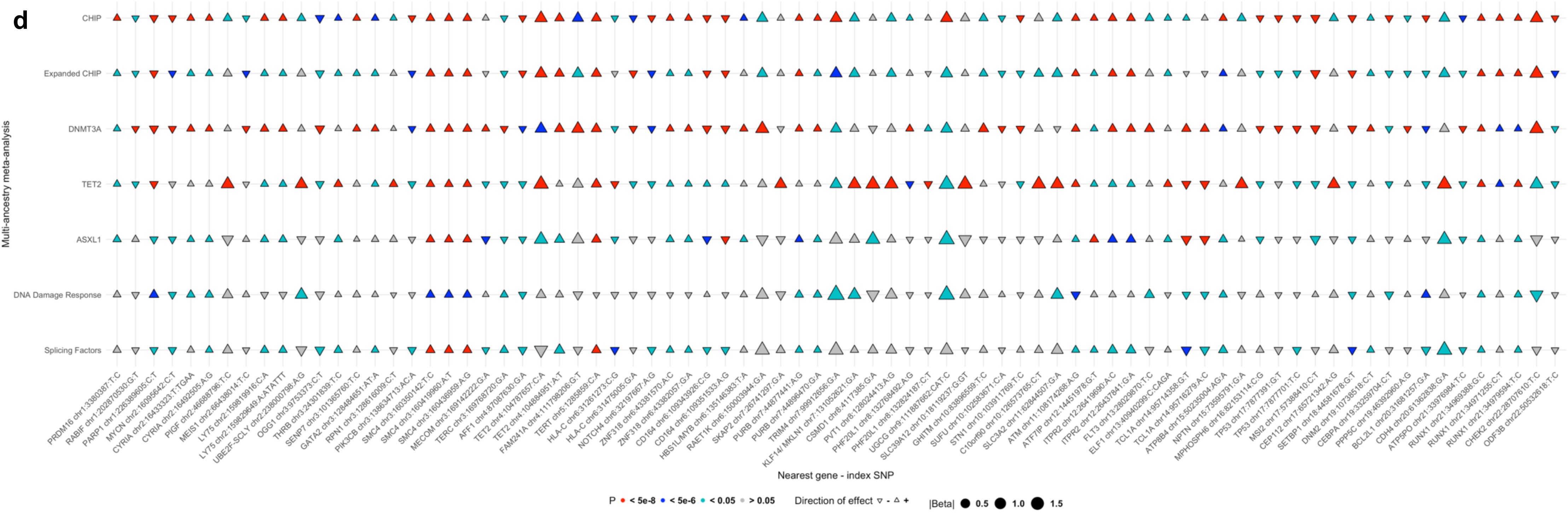
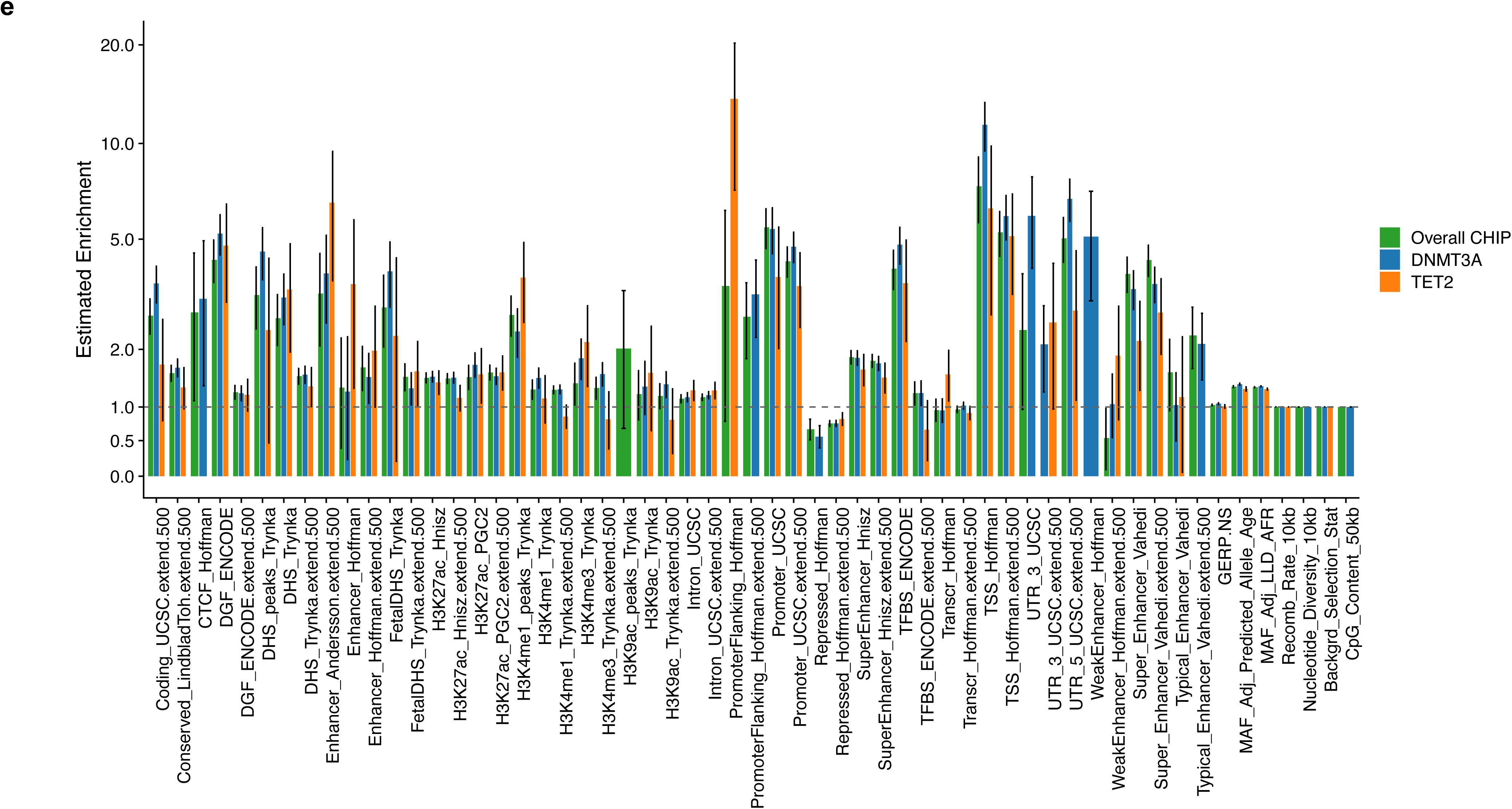
Genetic determinants of CHIP. Gnome-wide meta-analyses of CHIP traits identified (**a**) 31 genome-wide-significant (*P*<5e-8) loci (11 novel) for overall CHIP (effect allele frequency, EAF>= 0.1%; Nmax= 975,182; 53,274 cases), **(b)** 37 loci (13 novel) for *DNMT3A* CHIP (EAF≥0.1%; Nmax= 953,024; 27,968 cases), and (**c**) 24 loci (20 novel) for *TET2* CHIP (EAF≥0.2%; Nmax= 936,857; 10,723 cases). New loci are in red. **(d)** Overlap of overall CHIP lead variants with CHIP subtypes. Significant lead variants from overall CHIP, *DNMT3A*, and *TET2* CHIP GWAS was compared with CHIP categories. (**e)**, Estimated enrichment of SNP heritability (± 1 SD) in genomic functional categories (**Extended Data Table 3**). SNP heritability and enrichment were estimated using summary statistics from multi-ancestry meta-analyses of overall CHIP (N=784,558), *DNMT3A* (N=762,400), *TET2* (N=746,233), CHIP using LDAK ‘SumHer’ function. CHIP: clonal hematopoiesis of indeterminate potential.

The primary meta-analysis of overall CHIP identified 31 genome-wide significant loci (*P* <5×10^-8^), of which 11 are novel (**Fig. 3a; Extended Data Figs 3a, 11; Supplementary Data Tables 3, 10**). The strongest association was at the *TERT* promoter (rs7705526-A, odds ratio OR=1.24, 95 % CI=1.22–1.26, *P*=2.1×10^-213^). Conditional and joint (COJO) analysis resolved these signals into 36 statistically independent variants across 28 loci, revealing marked allelic heterogeneity at established loci like *SMC4*, *TET2*, *TERT,* and *CD164* (**Supplementary Data Table 18-20**). The novel loci, with regional association plots shown in **Extended Data Fig. 11,** implicate genes central to key biological pathways, including PI3K signaling (*PIK3CB*), Notch signaling (*NOTCH4*), transcriptional and chromatin regulation (*PRDM16*, *AFF1*, *ATF7IP*, *CEBPA*), lipid metabolism (*UGCG*, *ATP8B4*), and fundamental cellular processes like cytoskeletal dynamics (*CYRIA*), ubiquitination (*TRIM4*), and cell cycle control (*MPHOSPH6*), thereby broadening the known biological framework of CHIP predisposition^3,34^ (**Table 1**). Fine-mapping prioritized a set of high-confidence causal variants (PIP>60%), with the strongest signals observed at *TERT* (rs7705526, rs2853677, both PIP=100%; and rs35517815, PIP=86.6%). Other highly prioritized variants were rs35119987 at *ATP8B4*, rs201009932 in *SMC4*, and rs144317085 in *TET2*, along with rs10909937 in *PRDM16* and rs78446341 in *LY75* locus (**Supplementary Data Table 21**). Additionally, a parallel analysis of expanded CHIP identified 21 loci (19 novel), including signals at *CPNE9/OGG1*, *MICB*, *PXDNL*, and *TBX6* (**Extended Data Fig. 3b**; **Supplementary Data Table 4**). The majority of expanded CHIP loci overlapped with signals from the primary overall CHIP analysis, and their effect estimates were often larger (**Fig. 3d, Extended Data Fig. 9a**).

**Table 1.**
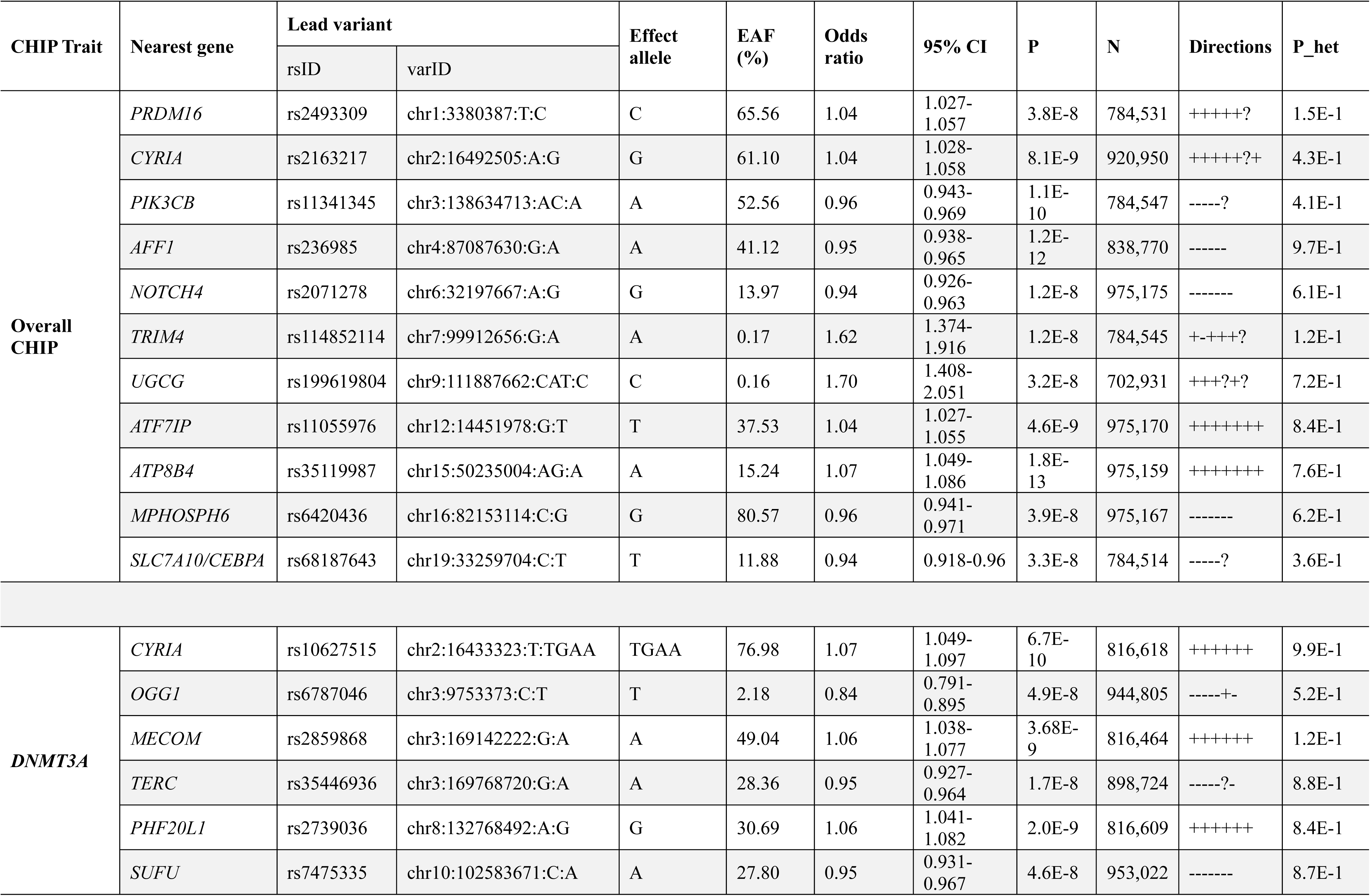
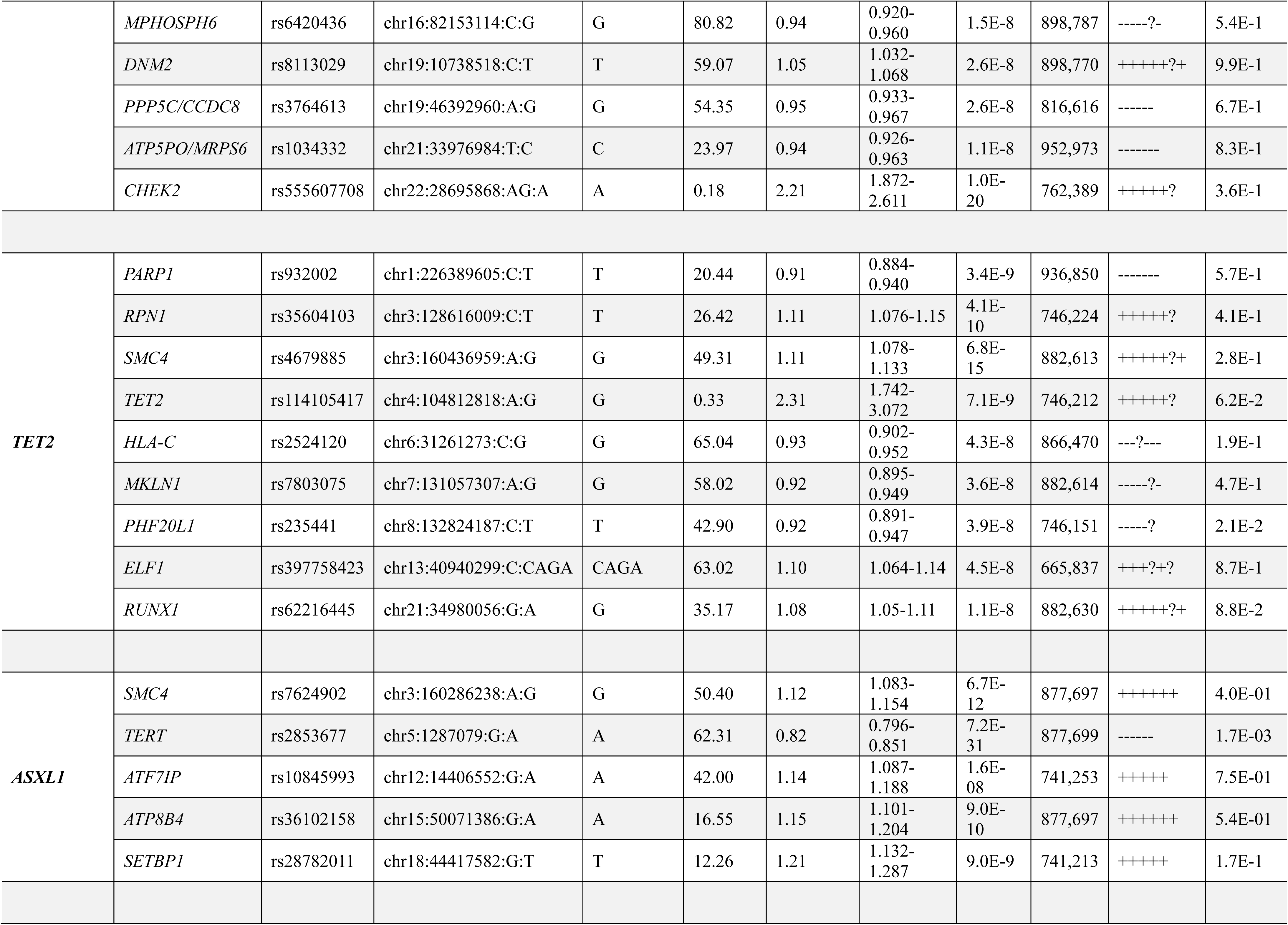
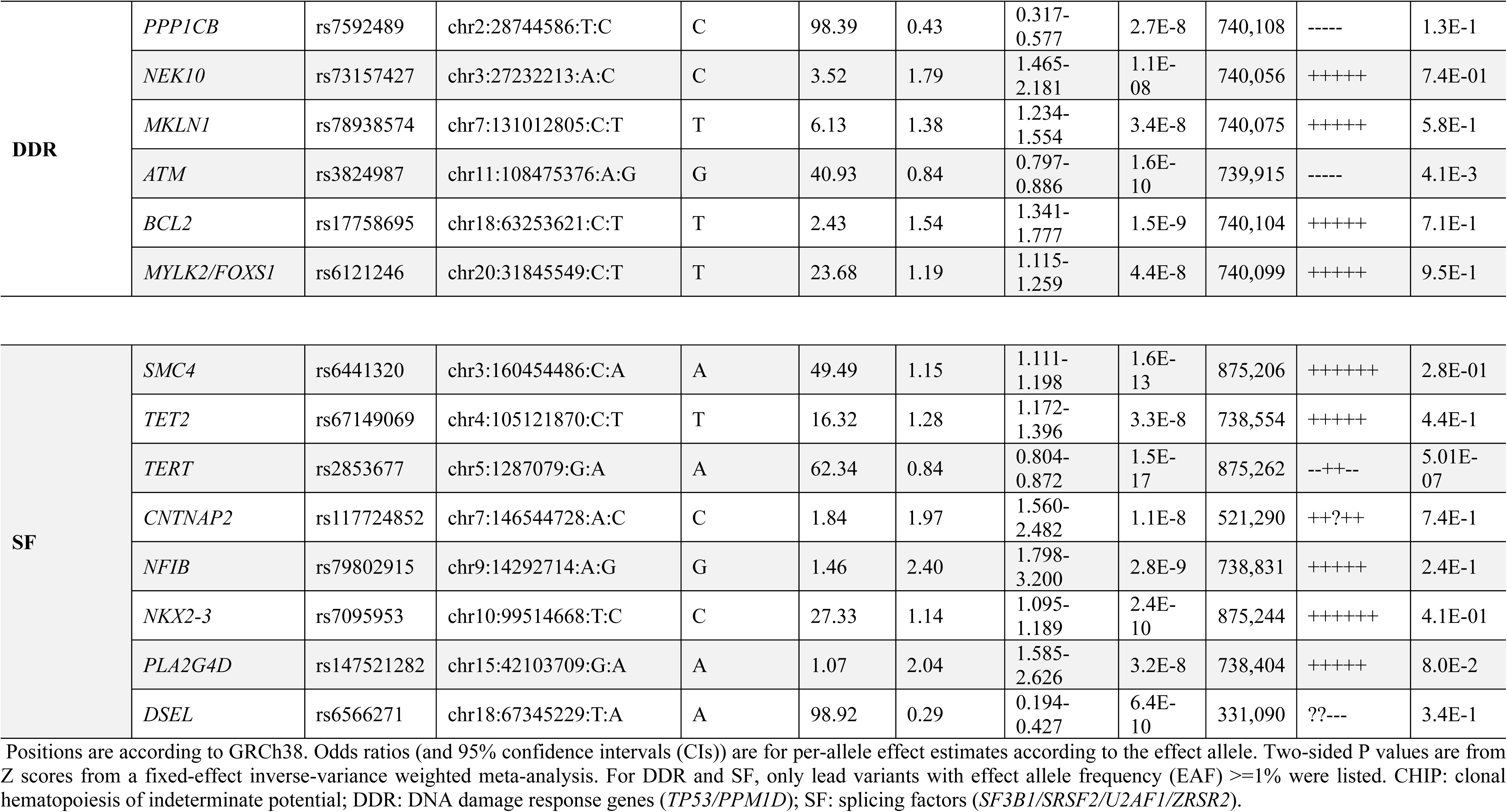
New loci for CHIP traits from meta-analyses.

To further delineate the genetic architecture of CHIP, we performed analyses stratified by ancestry, sex and driver gene. Notably, several loci—including *SMC4*, *TERT* and *SETBP1*—were consistently associated across CHIP traits, and showed concordant effects across ancestry and sex strata (**Fig. 3d; Extended Data Fig. 9**), suggesting shared germline determinants of CHIP traits. In ancestry-stratified analyses, effect estimates for the majority of lead variants identified in the multi-ancestry meta-analysis were highly concordant in EUR samples and showed partial concordance in AFR and AMR populations (**Extended Data Fig. 9b–h**).

These analyses also identified ancestry-specific associations. The EUR-stratified analysis, reflecting the largest sample size, identified several additional loci, including *PRDM16*, *CYRIA*, *PIK3CB*, *MECOM*, *AFF1*, *PHF20L1* and *ATP8B4* (**Extended Data Fig 4, 9b-h and 12; Supplementary Data Table 11**). At the *TET2* locus, distinct ancestry-specific lead variants were observed (EUR: rs144317085-T; AFR: rs114105417-G), consistent with prior reports^22,24^. In AFR-specific analyses, we identified a low-frequency variant at the *ATP8B4* locus (rs369044106-A; effect allele frequency (EAF) = 0.8%; OR = 5.14; P = 6.5×10^-9^) and a total of 20 genome-wide significant loci (at EAF ≥ 2%), including novel signals at *ADGRL4*, *TGFBR3*, *DPPA2*, *SMC4*, *TERT*, *TRIM4*, *PVT1* and *SETBP1* (**Extended Data Fig. 5, 9b-h and 13; Supplementary Data Tables 12-13**). Similarly, in AMR population, we identified eight genome-wide significant loci (EAF>=2%), including signals at *ATP8A1*, *NKX1-1*, *CSMD1*, *LMO7*, and *SETBP1* (**Extended Data Fig. 6, and 9b-h; Supplementary Data Tables 14-15**).

Sex-stratified multi-ancestry analyses were performed to assess the potential differences in genetic effects between males and females. Several loci reached genome-wide significance predominantly in males (including *PURB*, *ITPR2* and *ATP8B4*) or in females (including *MYCN*, *THRB*, *MEIS1*, *PVT1*, *STN1*, *TP53* and *MSI2*) (**Extended Data Figs. 7–8, 9b–h; Supplementary Data Tables 16–17**). However, these loci were also identified in the combined analysis, and Cochran’s *Q*-test did not detect significant heterogeneity in effect sizes between sexes (P_het_ > 0.05), indicating broadly consistent effects. In contrast, we identified a locus associated with DDR CHIP that showed significant sex heterogeneity. A common missense variant in *TADA2A* (rs7211875-C; p.Ser6Pro) was associated with higher odds of DDR CHIP mutations in females (OR=1.35, P=1.4×10^-8^; **Table 2**; **Extended Data Fig. 14**).

**Table 2.** Association of a *TADA2A* missense variant (rs7211875-C; chr17:37411381:T:C; p.Pro6Ser) with DNA damage response (*TP53* and/or *PPM1D*) CHIP in males and females.

| Sex | Study | EAF (%) | Odds ratio | 95% CI | P | N | P<sub>HET</sub> | P<sub>HET</sub> |
| --- | --- | --- | --- | --- | --- | --- | --- | --- |
| Male | UKB | 14.83 | 0.98 | 0.851-1.118 | 7.2E-1 | 187,742 |  | 2.8E-6 |
|  | AoU | 12.80 | 1.03 | 0.815-1.306 | 8.0E-1 | 86,106 |  |  |
|  | TOPMed | 13.40 | 0.83 | 0.827-1.749 | 3.3E-1 | 27,491 |  |  |
|  | MGBB | 14.71 | 0.88 | 0.705-1.108 | 2.8E-1 | 19,916 |  |  |
|  | Meta-analysis | 14.16 | 0.96 | 0.864-1.056 | 3.7E-1 | 321,255 | 6.9E-1 |  |
| Female | UKB | 14.87 | 1.35 | 1.186-1.541 | 1.4E-5 | 220,890 |  |  |
|  | AoU | 12.99 | 1.25 | 0.976-1.589 | 7.8E-2 | 131,546 |  |  |
|  | TOPMed | 13.16 | 1.45 | 1.009-2.087 | 4.5E-2 | 41,025 |  |  |
|  | MGBB | 14.40 | 1.41 | 1.053-1.889 | 2.1E-2 | 25,373 |  |  |
|  | Meta-analysis | 14.09 | 1.35 | 1.215-1.493 | 1.4E-8 | 418,834 | 8.8E-1 |  |

Odds ratios and 95% confidence intervals (CIs) represent per-allele effect estimates for the effect allele rs7211875-C. Two-sided P values were derived from approximate Firth logistic regression in the GWAS analysis or from fixed-effect inverse-variance weighted meta-analysis. EAF: effect allele frequency; CHIP: clonal hematopoiesis of indeterminate potential. P<sub>HET</sub>: Heterogeneity P from Cochran's *Q*-test.

Recognizing that the biology of CHIP is influenced by the initiating driver mutation, we next performed gene-specific meta-analyses for the common drivers (*DNMT3A*, *TET2*, *ASXL1*), splicing factors genes (*SF3B1*, *SRSF2*, *U2AF1, ZRSF2*), and DDR genes (*TP53*, *PPM1D*). These analyses revealed distinct, and in some cases opposing, genetic architectures modulating the fitness of specific mutant clones (**Fig. 3b-d**; **Table 1; Extended Data Figs. 3c-i, 9 and 11; Supplementary Data Tables 5-10, 19 and 20**).

Analysis of *DNMT3A* CHIP yielded 37 loci (13 novel); new loci implicating pathways related to DNA repair (*OGG1*), telomere maintenance (*TERC*) and cell cycle control (*CHEK2*) (**Table 1**; **Fig. 3b**; **Extended Data Fig 3c; Supplementary Data Tables 5, 10 and 19**). A strong association was observed at *MECOM* (rs62116797-T; OR=1.15; P=9.1×10^-12^), a key transcription factor regulating hematopoietic stem cell self-renewal^35,36^, with opposing allelic effect between *DNMT3A* and non-*DNMT3A* CHIP (**Fig. 3d**).

In contrast, analysis of *TET2* CHIP identified 13 loci (9 novel), implicating partially distinct biological pathways, including novel signals at hematopoietic regulators (*RUNX1*), DNA repair genes (*PARP1*) and immune-related loci such as *HLA-C* (**Table 1**; **Fig. 3c; Extended Data Fig 3d**; **Supplementary Data Tables 6, 10 and 20**). Notably, the chromatin reader *PHF20L1* exhibited opposing allelic effects between *DNMT3A* and *TET2*, with increased risk for *DNMT3A* CHIP but decreased risk for *TET2* CHIP (**Fig. 3d**)—echoing the bidirectional associations previously reported at *CD164* and *TCL1A*^23,24,37^. These findings support the existence of distinct germline architectures shaping the fitness landscape of specific mutant clones.

The *ASXL1*-specific analysis identified 12 loci, including 9 novel associations such as *ATF7IP*, *ATP8B4*, and the leukemia predisposition gene^38^ *SETBP1* (**Table 1**; **Fig. 3d; Extended Data Figs. 3e-f, 9a,f and 10; Supplementary Data Tables 7and 10**). Analyses of splicing factors CHIP identified eight novel loci, including *SMC4*, *TET2*, *TERT*, and *NKX2-3*, whereas analyses of DDR CHIP identified six novel loci, including *NEK10*, *ATM* and *BCL2* (**Table 1; Extended Data Figs. 3g-i; Supplementary Data Tables 8-10**). Notably, loci at *PARP1*, *TERT*, *ATM* and *CHEK2* showed opposing allelic effects in DDR CHIP compared to other CHIP subtypes (P<0.05), with *PARP1* and *TERT* risk alleles increasing susceptibility to DDR CHIP, whereas *ATM* and *CHEK2* risk alleles were associated with reduced risk (**Fig. 3d; Extended Data Figs. 9a,g-h**).

Together, these results highlight both shared and subtype-specific determinants of CHIP, implicating convergent pathways—including telomere maintenance, cell-cycle regulation, DNA damage response and regulation of self-renewal—and providing insight into germline-somatic interactions that shape clonal selection in hematopoiesis.

### Advanced analyses refine and expand the genetic architecture of CHIP

To enhance statistical power and leverage the shared genetic architecture between CHIP associated loci and those previously implicated in related hematological traits, including LTL, mCAs, and MPN, we applied a multivariate adaptive shrinkage (MashR) Bayesian model^29^. By jointly analyzing GWAS summary statistics across these correlated traits, Mash prioritized hundreds of additional loci, including 231 loci associated with overall CHIP, 268 with *DNMT3A* CHIP, and 227 with *TET2* CHIP that did not reach genome-wide significance in the primary single-trait analysis (**Supplementary Data Tables 22–24**). Analysis of these newly implicated loci reinforced the central role of genome instability in promoting clonal selection, revealing a striking convergence on pathways involved in DNA damage response, telomere maintenance, and chromatin remodeling.

To systematically identify causal genes within our GWAS loci, we applied the polygenic priority score (PoPS) method, which integrates GWAS data with other multi-omics data to prioritize causal genes based on functional similarity^28^, to summary-level data from the multi-ancestry meta-analysis of overall CHIP, *DNMT3A*, and *TET2* (see **Methods**). This approach identified dozens of high-confidence gene-trait associations (PoPS score>0.50), consistently highlighting key hematopoietic regulators and DNA damage response genes such as *PRDM16*, *PARP1*, *SMC4*, *TERT*, *ATM*, and *RUNX1* across CHIP and its subtypes (**Supplementary Data Tables 25-27**). The consistency between prioritized genes by PoPS and proximity to top GWAS signals suggests high confidence^28^. Further, this prioritization corroborates with the fine-mapped variants at *PARP1, LY75, THRB, SMC4, TERT, ATM, MSI2,* and *SETBP1* loci (**Supplementary Data Table 21**) and previously validated causal variants at *TET2*^22^ *and TCL1A*^37^.

### Phenome-wide analysis links CHIP to diverse clinical outcomes

To define the full clinical landscape of clonal hematopoiesis, we performed a comprehensive phenome-wide association study (PheWAS) of CHIP. We contrasted its effects with those of another age-related clonal phenomenon, mCAs, leveraging datasets totaling 1,116,579 individuals to maximize our discovery power for the mCA comparisons. This large-scale approach revealed strong associations between CHIP and a wide range of both cancerous and non-cancerous diseases.

Among incident hematopoietic and neoplastic conditions (**Fig. 4a; Extended Data Fig. 15; Supplementary Data Table 28**), significant associations were observed with leukemias (CHIP: hazard ratio [HR]=2.72, 95% CI: 2.44-3.03, FDR=2.1×10^-70^; autosomal mCAs: HR=6.79, 95% CI: 6.38-7.25, FDR<1.0×10^-300^), with the largest effect for expanded CHIP and monocytic leukemias (HR=47.2, 95% CI: 28.2-79.2, FDR=3.3×10^-46^) and for expanded autosomal mCAs and chronic lymphoid leukemias (HR=46.0, 95% CI 41.2–51.5, FDR<1.0×10^-300^), aligning with previous reports^2,40^. Besides hematopoietic malignancies, neoplastic associations were also detected with incident respiratory cancer, Kaposi’s Sarcoma, malignancies of the brain or spine, multiple myeloma, neurofibromatosis, melanomas, breast cancer, and cervical cancer for both CHIP and mCAs.

**Fig. 4.**
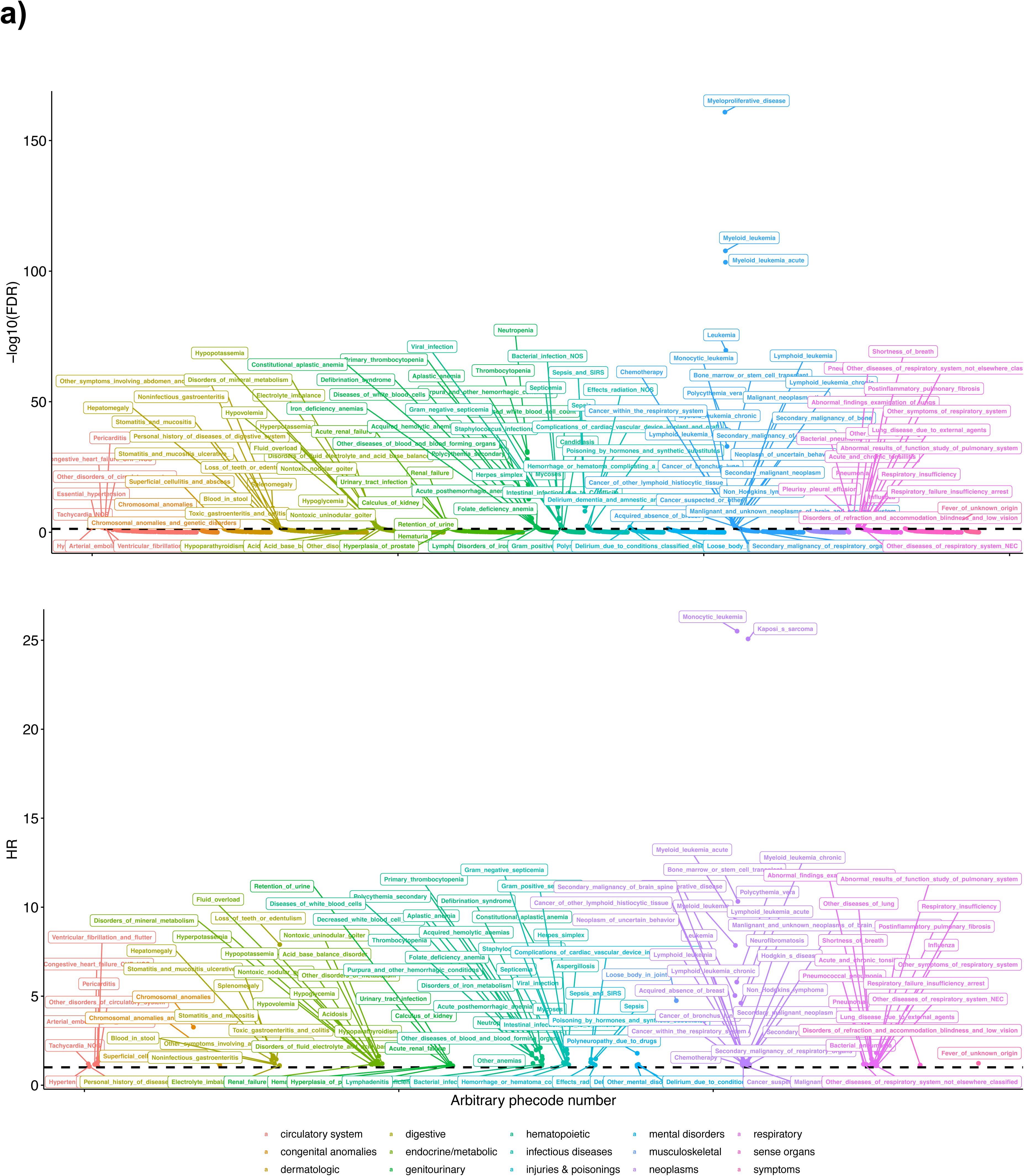
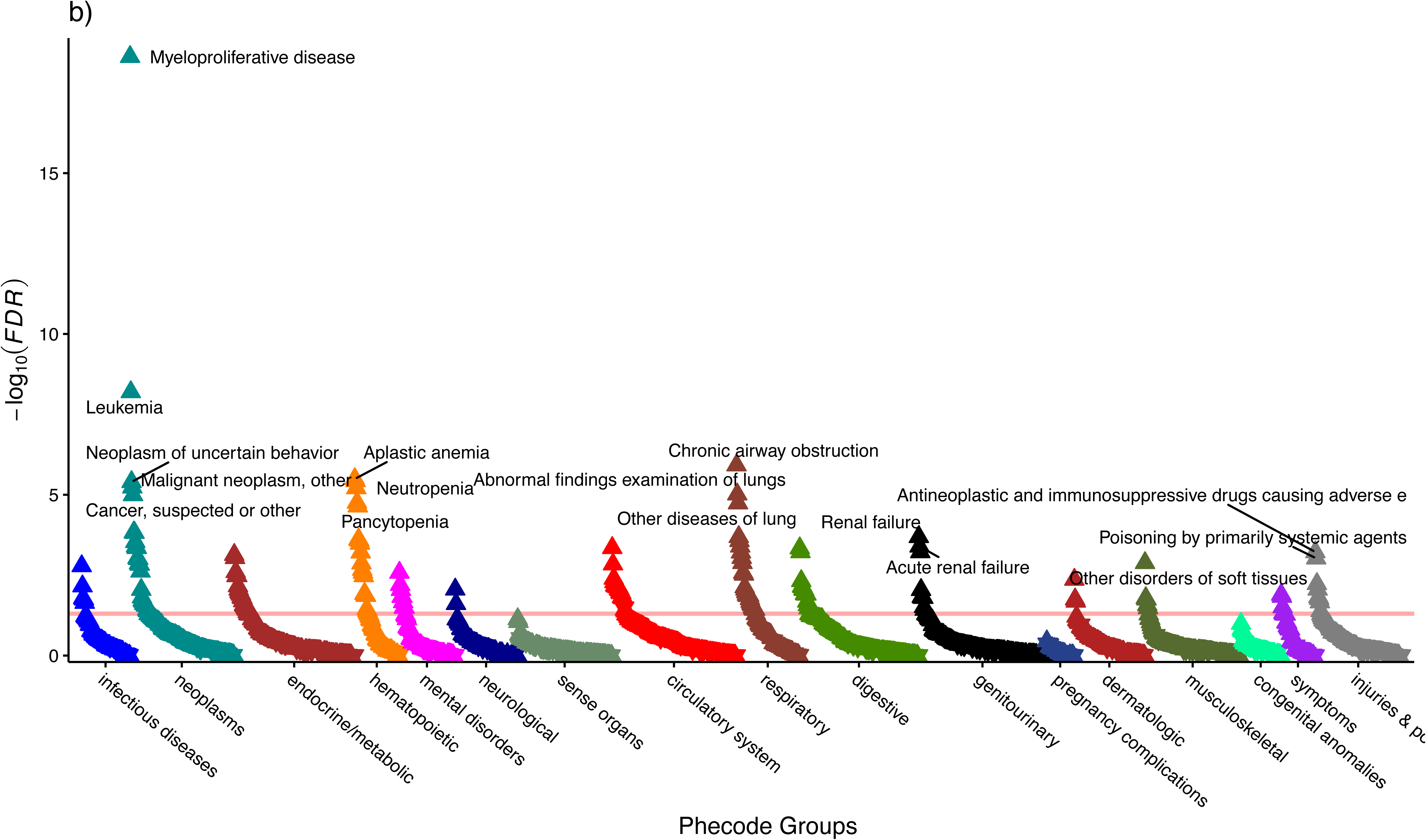
Phenome-wide association analysis of CHIP. a) Meta-analysis of CHIP PheWAS in UKB and MGBB: Significant (FDR<0.05) associations between CHIP and incident phecode phenotypes are presented here. Multivariable adjusted Cox regression included age, age^2^, sex, smoking status (using a 25-factor smoking status adjustment in the UK Biobank and current/prior/never smoker status in MGBB), tobacco use disorder, and first ten genetic principal components. Full summary statistics for CHIP and CHIP subtypes are available in **Supplementary Data Table 28**. b) PheWAS of CHIP in AoU: Associations between CHIP and any phecode phenotypes (i.e. prevalent and/or incident) are presented here. Multivariable adjusted logistic regression in the AoU analysis included age, age^2^, genetic sex, genetic ancestry, first five genetic principal components, status of smoked 100 cigarettes lifetime, batch effects for WGS sequencing center and CHIP calling. Full summary statistics for CHIP and CHIP subtypes are available in **Supplementary Data Table 29.**

For CHIP, across other non-hematopoietic/neoplastic conditions, significant associations were observed with circulatory, gastrointestinal, genitourinary, renal, and infectious conditions, some of which showed consistent associations with mCAs (**Fig. 4a; Extended Data Fig. 16-29; Supplementary Data Table 28**). CHIP was associated with incident myocardial infraction, heart failure, hypertension, tachycardia, ventricular fibrillation, arterial embolism and thrombosis, and pericarditis. In particular, among circulatory conditions, *JAK2* variants were associated with incident valvular disease, non-hypertensive congestive heart failure, cerebrovascular disease, and peripheral vascular disease. Variants of splicing factors were associated with pericarditis, congestive heart failure, subdural hemorrhage, and arteritis. A significant association with incident arterial embolism and thrombosis was detected for expanded mCAs (HR=1.17, 95% CI: 1.08-1.27, FDR=0.002), particularly for expanded ChrX mCAs in females (HR=3.92, 95% CI: 1.92-8.02, FDR=0.03) (**Supplementary Data Table 28)**.

Given the limited follow-up in AoU, we performed separate PheWAS using logistic regression on combined prevalent and incident cases of each phecode^39^ to evaluate potential CHIP associations. Overall CHIP was associated (FDR<0.05) with hematologic and non-hematologic conditions (**Fig. 4b; Extended Data Fig. 30a-j; Supplementary Data Table 29**). In stratified analysis by driver genes, we observed large effect sizes for myeloproliferative disorders, particularly in carriers of *JAK2 V617F,* which increased the odds of myelofibrosis by over 100-fold (OR=112, FDR=2.6E-16), polycythemia vera by almost 47.4-fold (FDR=6.6E-38), and myeloproliferative disease by 37.4-fold (FDR=1.8E-79). *TP53, PPM1D, TET2,* and *ASXL1* CHIP were also associated with higher risk of hematological phenotypes such as myelofibrosis, leukemia, and cytopenia (**Supplementary Data Table 29**). Beyond blood-related conditions, associations extended to other organ systems, for instance, renal impairment, and certain circulatory or respiratory outcomes—though with more modest effect sizes (**Supplementary Data Table 29**). Collectively, these AoU findings reinforce the strong links between specific CHIP driver mutations and both hematologic malignancies and broader systemic phenotypes.

### Polygenic associations of CHIP at a single-cell level corroborated PheWAS findings

We used scDRS^33^ to assess the polygenic enrichment of overall CHIP, *DNMT3A*, and *TET2* GWAS signals across cell populations using single-cell RNA sequencing (scRNA-seq) data from the Tabula Sapiens (TS) human cell atlas^40^. scDRS quantifies the enrichment of GWAS-associated gene expression at single-cell resolution; associations were considered significant at FDR < 0.2 and suggestive at P < 0.01 (see **Methods**). Results are shown in **Fig. 5a–c**. Overall, CHIP, *DNMT3A*, and *TET2* signals were significantly enriched in hematopoietic stem cells from bone marrow. In addition, CHIP and *DNMT3A* showed broader enrichment across multiple cell types, including granulocytes, neutrophils, natural killer (NK) cells, mature enterocytes, hepatocytes, and erythroid progenitors (**Fig. 5a**). These associations were robust to subsampling of cells and genes (**Fig. 5b**). Patterns of enrichment were consistent between cell type-level and tissue-level analysis for CHIP and *DNMT3A*, whereas no significant tissue-level enrichment was observed for *TET2* (**Fig. 5c**).

**Fig. 5.**
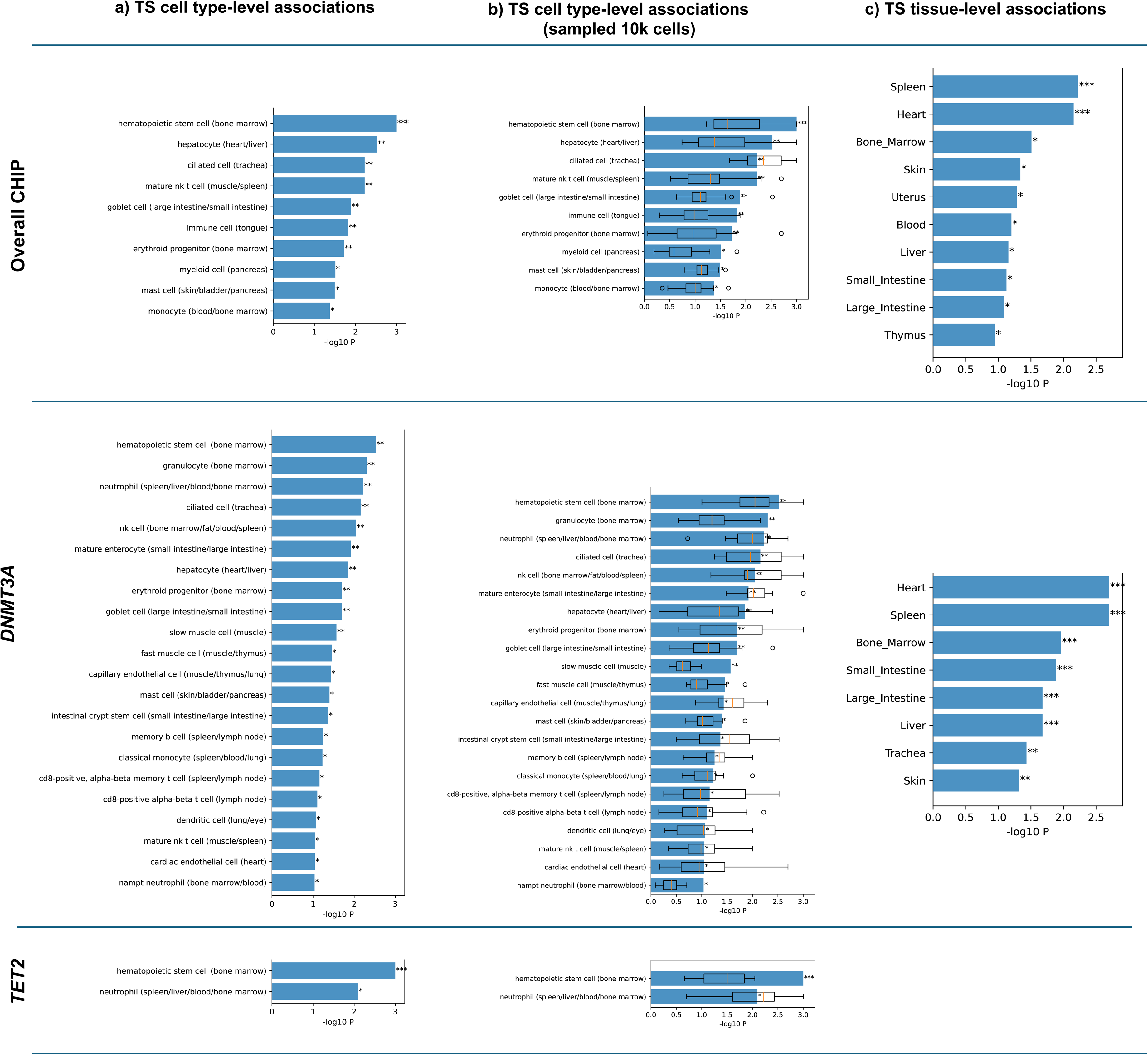
Cell and tissue type-level associations in Tabula Sapiens (TS) human cell atlas. a) Cell type-level associations in TS. b) Sensitivity analysis via subsampling 10K cells from the TS data (25K cells). c) Tissue-level associations in TS. The X-axis represents scDRS –log10 cell / tissue type-level association *P* while the Y-axis represents potentially associated cell / tissue types (*P*<0.01 or *FDR*<0.3) for overall CHIP, *DNMT3A* and *TET2* CHIP, ordered by significance. The boxplots represent the distribution of scDRS –log10 *P* computed from the 20 repetitions of the subsampling experiments. *Denotes *FDR*<0.3, **denotes *FDR*<0.2, and ***denotes *FDR*<0.1 across all cells / tissues for a given trait. CHIP: clonal hematopoiesis of indeterminate potential; scDRS: single-cell disease relevance score.

### Polygenic risk of CHIP predicts hematologic malignancy

To translate our GWAS findings to risk stratification tools, we constructed polygenic risk scores (PRS) for overall CHIP and its subtypes using a discovery set of non-UKB cohorts. When tested in the independent UKB cohort, each PRS was robustly associated with its corresponding CHIP status, with OR ranging from ∼1.07 to ∼1.20 per SD increase in PRS (**Fig. 6a**). For instance, the *DNMT3A*-specific PRS was associated with a 20% increased odds of *DNMT3A*-CHIP per SD increase in the score 1.20 (OR=1.20; 95% CI, 1.18–1.22; P=6.7×10^−112^).

**Fig 6.**
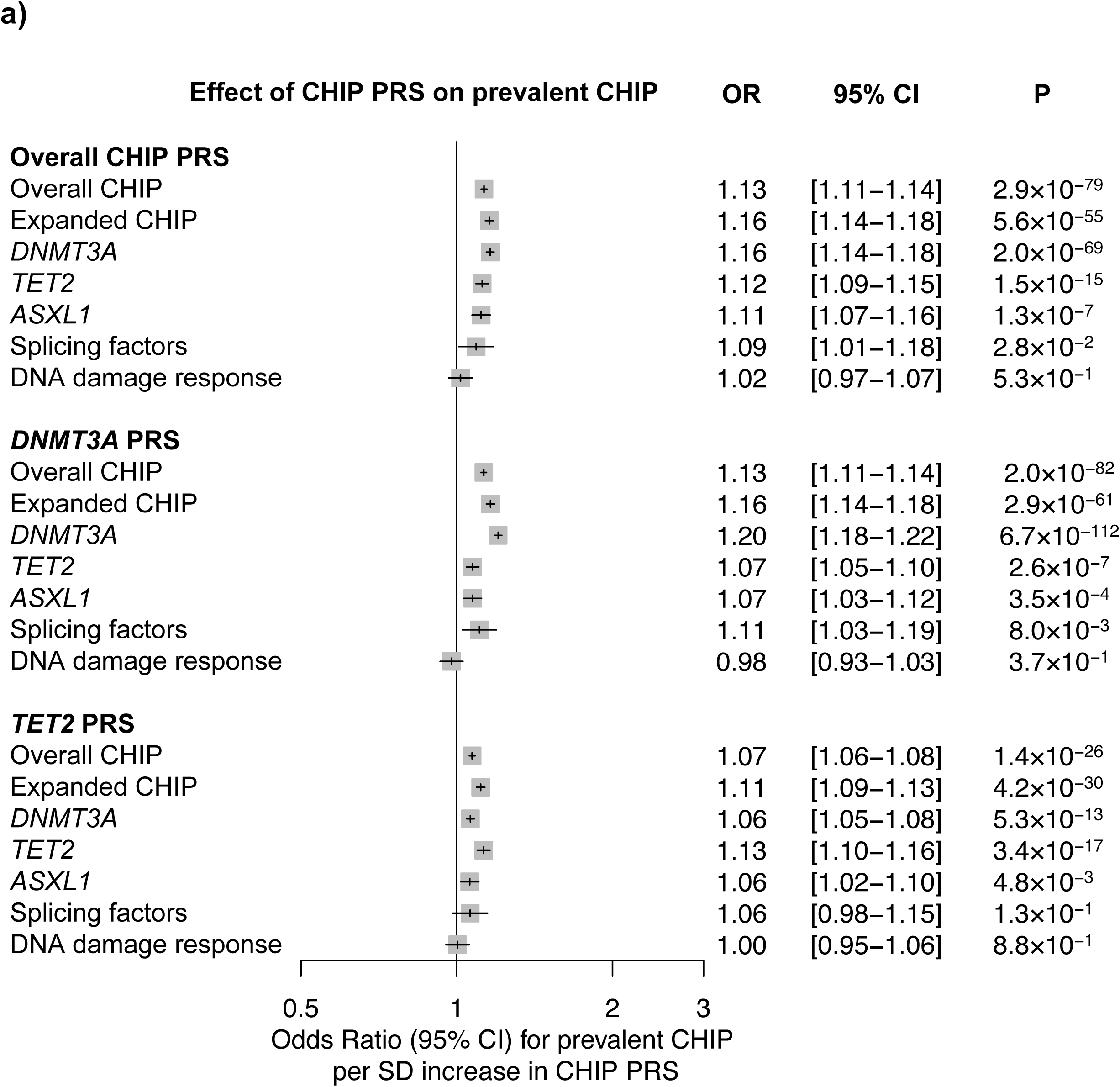
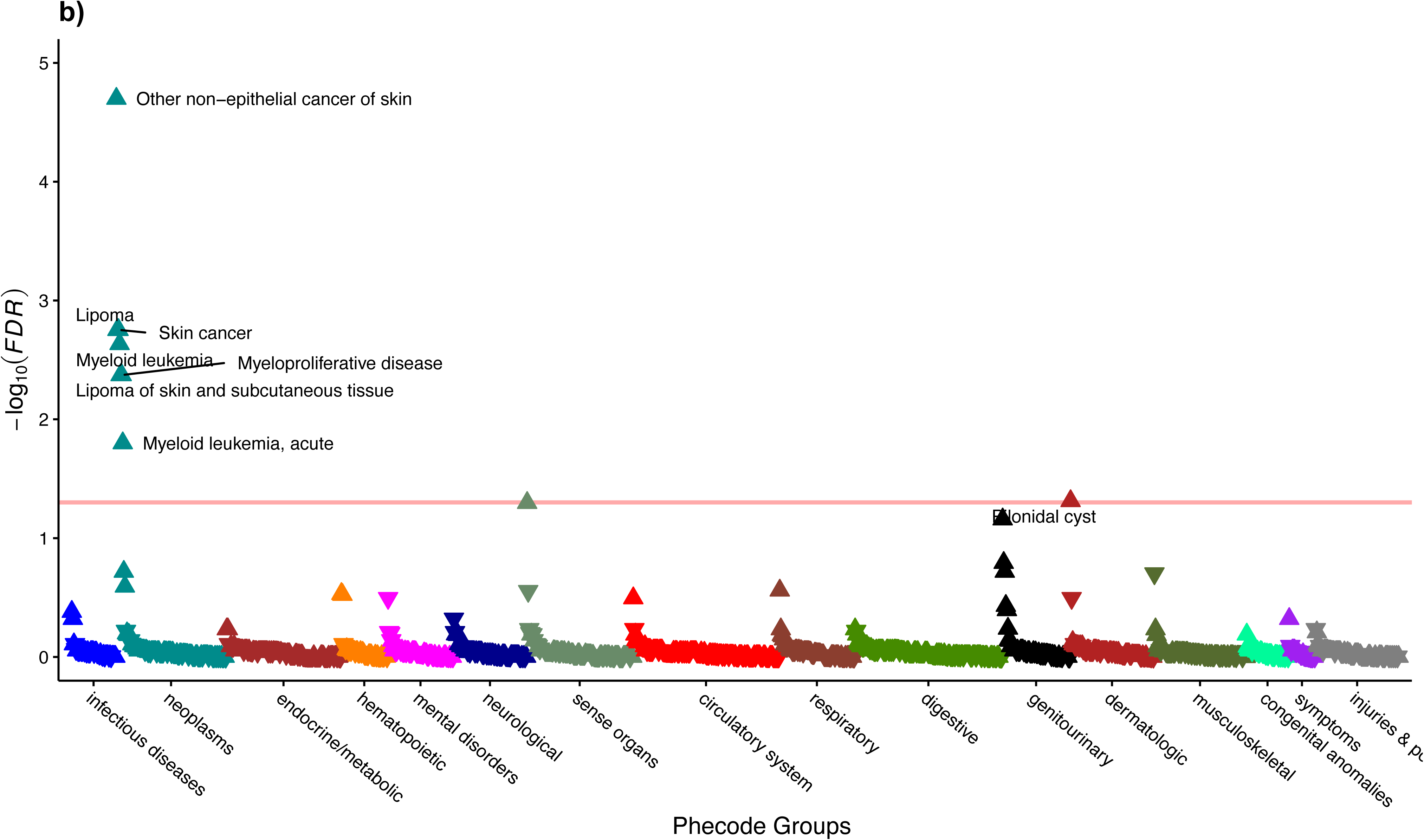
CHIP PRS in UKB. a) Association between CHIP PRS and CHIP prevalence. Multivariable adjusted logistic regression model included age, age^2^, genetic sex, genetic ancestry, smoking status, the first five genetic principal components, and study-specific batch effects to account for potential confounders. **b) PheWAS of CHIP PRS**: association between CHIP PRS and incident disease in UKB was tested using Cox regression including covariates such as age, age^2^, genetic sex, genetic ancestry, smoking status, the first five genetic principal components, and study-specific batch effects to account for potential confounders. Full PheWAS summary statistics for CHIP PRS are available in **Supplementary Data Table 30.**

We next performed a PheWAS of the CHIP PRS in the UKB to explore the broader clinical consequences of a high germline predisposition to CHIP (**Supplementary Data Table 30**). Overall, these PheWAS findings indicate that individuals with higher genetically inferred risk of CHIP are also more likely to develop hematologic neoplasms, such as acute myeloid leukemia (overall CHIP PRS: β=0.20, P=7.4×10^−6^) and myeloproliferative disorders (for example, *TET2* CHIP PRS: β=0.15, P=3.1×10^−7^; expanded CHIP PRS: β=0.14, P=3.0×10^−6^), consistent with known clinical links between CHIP and blood cancers. Elevated CHIP PRS also showed significant associations with certain solid tumors, such as non-epithelial skin cancer and lipomas, as well as hematologic phenotypes (for example, thrombocytopenia, β=0.057, P=2.8×10^−3^ for the overall CHIP PRS). Further, this risk translated into future disease development. Longitudinal analysis confirmed that individuals with a higher CHIP PRS had a significantly increased risk of incident hematologic malignancies (**Fig. 6b** and **Supplementary Data Table 30**), underscoring the clinical potential of these genetic predictors for identifying individuals at elevated cancer risk.

## Discussion

In this comprehensive well-powered multi-ancestry study of over one million individuals, the largest so far, we substantially expanded the genomic and phenomic architecture of clonal hematopoiesis. By moving beyond previous discoveries^22–25^, we revealed a highly polygenic basis for CHIP that is profoundly shaped by the initiating somatic driver mutation. We identified dozens of loci (72 significant regions for overall CHIP, *DNMT3A,* and *TET2* CHIP, including 44 that are novel), systematically mapped the distinct clinical consequence of CHIP and compare it with mCAs to highlight the unique and shared association patterns between the two clonal phenomena, and leveraged genomic methods and tools to explore deeper genomic and phenomic architecture of CHIP. Together, this work provides a foundational atlas that links germline variation to the selection of somatic clones and their subsequent impact on human health and aging.

A central theme emerging from our genetic discoveries is the critical role of germline-encoded genome integrity in shaping CHIP risk. Our primary and driver gene-specific GWAS identified 31 and 37 loci for overall and *DNMT3A* CHIP, respectively, and converged on fundamental pathways of DDR and telomere maintenance. We not only confirmed associations at known DDR genes like *ATM, CHEK2,* and *PARP1* but also uncovered novel loci implicating key regulators such as *MECOM*, a master transcription factor essential for HSC self-renewal^35,36^. The profound influence of this axis is further underscored by our finding that the heritability of DDR CHIP was the highest of any subtype (21.0%). These results not only provide a mechanistic framework for epidemiological observations linking CHIP to exposures such as smoking and cytotoxic therapy^41,42^, but also suggest that inherited variants create a sensitized background upon which environmental mutagens can more readily promote the acquisition or expansion of somatic clones.

Our results also clearly demonstrate that the germline architecture of CHIP is not monolithic but is instead a mosaic of driver-specific landscapes. The genetic associations for *DNMT3A* CHIP are different from those for *TET2* and other CHIP traits, a concept powerfully illustrated by the bidirectional effect at the *MECOM* and *PHF20L1* loci, which confer an increased risk for *DNMT3A* clones but a reduced risk for *TET2* clones, a phenomenon previously reported in *CD164* and *TCL1A*^43^.

These emerging human genetic and functional evidence converges on a core set of germline regulators—*SMC4*, *TERT* and *SETBP1*—that collectively shape the size, durability and competitive fitness of the hematopoietic stem and progenitor cell (HSPC) compartment^26,44–47^. Genome-wide association studies of clonal hematopoiesis consistently nominate these loci as shared risk factors, implicating them as upstream determinants of HSC pool dynamics and mutational opportunity^22–24,48^. Mechanistically, *SMC4*, a core component of the condensin complex, is positioned to regulate higher-order chromatin architecture and mitotic fidelity, thereby sustaining proliferative capacity and genome integrity in actively cycling HSPCs^46,47,49^. In parallel, germline variation at the *TERT* locus links telomere maintenance to clonal hematopoiesis risk, supporting a model in which longer telomere length or stability extends HSC replicative lifespan and permits prolonged clonal persistence^26,44,45,47^. By contrast, *SETBP1* operates at the level of transcriptional and chromatin regulation: experimental studies demonstrate that increased *SETBP1* expression can directly promote self-renewal of hematopoietic progenitors through activation of HOXA-driven programs, effectively conferring stem-like properties outside the canonical HSC compartment^38,50^.

Building on this framework, our results support a hierarchical, data-driven model of germline regulation in clonal hematopoiesis in which inherited variation operates across three distinct but interacting layers. First, a set of shared loci across CHIP subtypes—including *SMC4*, *TERT* and *SETBP1*—define a common “fitness landscape” that expands the effective HSPC pool, prolongs replicative lifespan and increases the probability of acquiring somatic driver mutations, consistent with prior genetic studies of clonal hematopoiesis and telomere biology^44,46,51^. Second, driver-specific germline loci partition this expanded pool into selective niches that favor the outgrowth of particular mutant clones, such that variants at *MYCN*, *CYRIA*, *MEIS1* and *SENP7* preferentially promote *DNMT3A*-CHIP expansion, whereas *THRB*, *RPN1* and *ELF1* bias toward *TET2*-CHIP clones, and *HDAC4*, *ATF7IP* and *ATP8B4* toward *ASXL1*, with additional distinct architectures observed for DNA damage response and splicing factor CHIP. These findings support that germline background encodes a selective landscape that is not uniform but instead tuned to specific epigenetic and transcriptional states imposed by individual driver mutations. Third, we identify a class of loci exhibiting opposing allelic effects across CHIP subtypes, including *MECOM*, *CD164*, *ATM*, *PHF20L1* and *TCL1A*, suggesting that certain germline variants stabilize one clonal trajectory while constraining others, potentially through incompatible chromatin or signaling states. Together, these observations support a unifying model in which germline variation first sets the scale of the HSPC compartment, and then deterministically biases clonal competition in a driver-dependent manner, providing a mechanistic basis for the observed heterogeneity in CHIP architecture across individuals. This discovery transforms previous understanding from a general predisposition to CHIP towards a more granular, driver-aware model of germline-somatic interaction.

By systematically profiling the phenotypic consequences of CHIP, our findings support its role as a systemic, age-related condition with effects extending beyond the hematopoietic system. Phenome-wide analyses not only confirmed the strong link between CHIP and subsequent myeloid malignancies (HR=2.72), particularly for large *JAK2* clones (OR for myelofibrosis > 100), but also demonstrated its significant association with a spectrum of cardiovascular diseases, including coronary artery disease and arterial thrombosis, consistent with previous individual epidemiological studies^1,3,11,14^. Single-cell analyses provide a potential mechanistic link for these pleiotropic effects, revealing polygenic enrichment of CHIP-associated genes in hematopoietic stem cells, as well as in diverse hematopoietic (immune) and non-hematopoietic cell types, including granulocytes, neutrophils, NK cells and hepatocytes—particularly for *DNMT3A*. Finally, the distinct disease associations observed for CHIP (predominantly linked to myeloid malignancies) and mCAs (more strongly associated with lymphoid malignancies) are consistent with prior reports^32,52^ and suggest partially distinct underlying biological mechanisms, despite some shared risk factors.

In addition, in our study, novel statistical methods coupled with well-powered GWAS yielded new insights into CHIP biology. Our study leveraged statistical approaches to access the polygenic enrichment of CHIP, *DNMT3A*, and *TET2* in scRNA-seq data^33^. Corroborating the significant associations with hematologic, immune, and circulatory conditions in pheWAS results. Also, in addition to the large sample size, we further boosted the power for locus discovery through jointly modeling multiple highly correlated traits (e.g. CHIP categories, LTL, MPN, and expanded mCAs) using an empirical Bayes hierarchical approach^29^, which yielded >8 fold increase in locus discovery for CHIP categories; These findings are suggestive and require validation in an independent dataset, however, can be used for hypothesis testing in future studies.

Important limitations of our study include that the cross-sectional nature of our CHIP analysis limits conclusions regarding the temporal evolution between CHIP and these diseases. Also, the technical sensitivity of whole genome and whole exome sequencing precludes the ability to evaluate the clinical significance of CHIP mutations at low VAF, which recent studies suggest can influence disease associations^21,53^.

Overall, our large analyses of the inherited basis of CHIP identified new mechanisms affecting somatic mutation acquisition and clonal fitness with an expanded appreciation for how these somatic mutations shape disease phenotypes.

## Methods

### Study populations

UK Biobank (UKB) is a prospective cohort study of ∼500,000 participants (age range 40-69 at enrollment) from across the United Kingdom with deep genetic (DNA isolated from blood) and phenotypic data^54^. The present study was conducted under UKB application 7089 and 50834. The secondary use of UKB data was approved by the Mass General Brigham Institutional Review Board (protocol 2021P002228).

The All of Us Research Program (AoU) is a nationwide initiative led by the National Institutes of Health (NIH), designed to support precision medicine research by gathering comprehensive health data from a diverse group of participants across the United States^55,56^. This study included 245,388 participants (mean age=52; SD=17; 60.5% Female; 49.8% non-European) with whole-genome sequencing (WGS) data (data release C2022Q4R13; 09/10/2024). The secondary use of AoU data was approved by the Mass General Brigham Institutional Review Board (protocol 2020P001737).

NHLBI Trans-omics for Precision Medicine (TOPMed) is a research program generating genomic data from DNA isolated from blood and other -omics data for more than 80 NHLBI-funded research studies with extensive phenotype data^22,57^. A total of 51 studies with diverse reported ethnicity (40% European or non-Hispanic white, 32% African, African American, or Black, 16% Hispanic/Latino, 10% Asian) were included in the Freeze 8 (https://topmed.nhlbi.nih.gov/topmed-whole-genome-sequencing-methods-freeze-8). Each of the included studies provided informed consent. This study included 74,622 participants from TOPMed that were approved for Study Proposal 11420. The secondary use of dbGaP and TOPMed data was approved by the Mass General Brigham Institutional Review Board (protocol 2016P002395 and 2016P001308).

Mass General Brigham Biobank (MGBB)^58^ is a volunteer biobank of patients receiving care at Mass General Brigham with electronic health records and genetic and phenotypic data on ∼50,000 participants (https://biobank.massgeneralbrigham.org/). The secondary use of Mass General Brigham Biobank data was approved by the Mass General Brigham Institutional Review Board (protocol 2018P001236 and 2020P003064).

The Vanderbilt University Biobank (BioVU) is a de-identified biorepository that includes DNA and plasma from the peripheral blood remaining from routine clinical testing of approximately 250,000 patients^59,60^. The present secondary analyses were approved by the Vanderbilt University Medical Center Institutional Review Board (IRB #201783).

### Sample Exclusion

samples with prevalent hematologic malignancy were excluded from GWAS analysis. The International Classification of Disease (ICD) codes, such as ICD-10: C81-86, C88, C90-96, D45-47; and ICD-9: 200-208, 238.4, 238.71, 238.72-238.75, 238.76 were used to ascertain prevalent hematologic malignancy status in these cohorts.

### CHIP detection

#### UKB

Whole-exome sequencing (WES) of whole blood DNA of 454,927 participants was used to identify somatic mutations. Somatic mutations were called in two batches: batch 1 included 200,628 WES from first release, where 500 random youngest samples were used for panel-of-normal (PON), and batch 2 included the remaining 254,199 WES, where publicly available PON prepared from WGS data from the 1000 genome project (gs://gatk-best-practices/somatic-hg38/1000g_pon.hg38.vcf.gz) was used. We called somatic mutations on 200,128 samples from first batch using Mutect2 software^61^ in the Terra platform (https://portal.firecloud.org/?return=terra#methods/gatk/mutect2-gatk4/20), and 254,199 samples from second batch using Mutect2 software in the UKB Research Analysis Platform (https://ukbiobank.dnanexus.com/landing). PON was used to minimize sequencing artifacts, and the Genome Aggregation Database (gnomAD)^62^ was used to filter likely germline variants from the putative somatic mutations call set. Each Variant Call Format (VCF) file was annotated using ANNOVAR software^63^, and putative CHIP mutations were identified using the pipeline described in Bick et al.^22^ (https://app.terra.bio/#workspaces/terra-outreach/CHIP-Detection-Mutect2; last accessed Feb 7, 2022). For identifying CHIP, pathogenic variants were queried in 74 genes known to drive clonal hematopoiesis and myeloid malignancies (list of variants queried is presented in **Supplementary Data Table 31)**^22^. We kept variants for further curation if (i) total depth of coverage ≥10, (ii) number of reads supporting the alternate allele ≥3, (iii) ≥1 read in both forward and reverse direction supporting the alternate allele, and (iv) VAF≥2%. Finally, CHIP mutations that passed sequence-based filtering were manually curated by a team of hematopathologists. The median depth of coverage for identified CHIP mutations was 77 (mean=80; SD=31.2; range 9-305), and the median number of supporting reads was 7 (mean=10; SD=8.6; range 3-101) in the final call set.

### AoU

In AoU, WGS data from whole blood were available for 245,388 participants (v7 data release). The ancestry categories included 53,942 African/African American (AFR), 40,837 American Admixed/Latino (AMR), 5,381 East Asian (EAS), 123,069 European (EUR), and 2,342 South Asian (SAS) participants (**Supplementary Data Table 1**). CHIP calling from WGS and filtering were conducted in two separate batches. Somatic mutations in known CHIP genes were extracted as previously described by^64^. Variants were excluded if they had total coverage <20, supporting reads <3, or missing support in either forward or reverse strand. Due to poor mapping quality of CHIP variants in *U2AF1*, only variants with ≥5 supporting reads were considered. A total of 10,677 CHIP mutations were identified in 9,755 participants.

#### TOPMed

WGS was performed on 97,691 blood DNA samples from Freeze 8 NHLBI TOPMed dataset using Illumina HiSeq X Ten instruments with a mean depth of at least 30×. All sequences were mapped to the GRCh38 human genome reference following the protocol published previously^65^. Single nucleotide variants (SNV) and short indels were jointly discovered and genotyped across the TOPMed samples using the GotCloud pipeline^66^. The procedure used for CHIP and germline variants calling has been previously described^22,67^.

#### MGBB

WES was performed on whole blood DNA samples from 53,445 MGBB participants with an average coverage of 55X. The CHIP calling procedure described for UKB (above) was applied to WES data from MGBB. PON was created from 100 random WES samples from the youngest participants (age at enrolment ≤21y). Putative CHIP mutations that passed the minimum coverage threshold (≥20 reads) and supporting read criteria (≥3 supporting reads, including at least one read from both forward and reverse strands) were manually curated to generate the final CHIP call set. Hotspot mutations in *U2AF1* (S34F, S34Y, R156H, Q157P, and Q157R) were identified using a custom script available at (https://github.com/MMesbahU/U2AF1_pileup).

#### BioVU

Among 54,583 participants of the BioVU, CHIP was identified from blood DNA using the Illumina Multi-Ethnic Genotyping Array-Expanded (MEGA^EX^) for somatic mutations in four known CHIP driver genes: *DNMT3A*, *JAK2*, *TET2*, and *ASXL1*. We examined nonsense, splice site, and previously reported missense variants genotyped on the MEGA^EX^ array, totaling 29 CHIP mutations. B-allele fractions (BAF) were calculated from the intensity of alternate allele/ (alternate intensity + wild-type intensity), following the method previously developed and validated by^68^. To validate this method of detecting somatic mutations, we evaluated 149 MEGA^EX^-genotyped patients with a putative *JAK2* mutation and performed NGS analysis on the same DNA sample. The two genotyping methods demonstrated high concordance with the NGS detection limit of 5% VAF (*R^2^*=0.9931). Because the MEGA^EX^-sequenced DNA samples were unavailable for *DNMT3A*, *TET2*, or *ASXL1* at the time of the present study, an alternative method was employed for these genes. The mean BAF and its standard deviation were calculated for the population under 40 years of age. Detectable CHIP is extremely rare before this age, allowing for determining the noise in BAF measurements at baseline. The BAFs of the over-40 population were normalized to the under-40 mean and standard deviation. Individuals with a normalized BAF greater than or equal to 6 standard deviations above the mean were considered to have a somatic CHIP mutation. The most common mutations in BioVU samples were *JAK2* V617F and *DNMT3A* R882C/H.

### Genetic ancestry inference

Genetic ancestry for participants in the UKB, TOPMed, and MGBB was inferred by projecting study samples onto reference populations. Variants shared between each target dataset and external reference panels (Human Genome Diversity Project combined with the 1000 Genomes Project, or the 1000 Genomes Project alone) were first identified. The intersecting variants were LD-pruned to remove highly correlated markers, and principal components (PCs) were computed using reference samples only. Study participants were subsequently projected into the reference PC space using per-variant PC loadings. A k-nearest neighbors classifier trained on the first 20 PCs of the reference samples was then used to assign each participant to the most genetically similar reference population. For the All of Us cohort, genetic ancestry assignments were obtained from available cohort data generated using previously described methods (Bick et al. ^69^).

### CHIP prevalence and baseline risk factors

Across cohorts (UKB, AoU, TOPMed, and MGBB), multivariable-adjusted logistic regression was performed to assess the association between baseline risk factors and CHIP, with each model adjusting for age, age², genetic sex, genetic ancestry, and the first five genetic principal components. Smoking status was included, categorized as never, ever, or unknown in UKB, TOPMed, and MGBB, and as having smoked ≥100 cigarettes in a lifetime (no, yes, or unknown) in AoU. Additionally, UKB and TOPMed models adjusted for the genotyping batch, AoU adjusted for WGS sites, and all cohorts adjusted for the CHIP calling batch.

### PheWAS analysis

The PheWAS of CHIP and mCAs with incident phenotypes across all disease organ system categories were performed using Cox proportional hazards models, adjusting for age, age^2^, sex (not used for ChrY and ChrX mCAs analysis), smoking status (using a 25-factor smoking status adjustment in the UKB and current/prior/never smoker status in other cohorts), tobacco use disorder, and principal components 1-10 of genetic ancestry. In the secondary analysis, mCAs associations were meta-analyzed across the multi-ancestry study population of UKB, MGBB, BioVU, and MVP. Time since DNA collection was used as the underlying timescale. The proportional hazards assumption was assessed using Schoenfeld residuals and was not rejected. Individuals with a known history of hematological cancer before DNA collection were excluded. To address multiple testing, an association between CHIP or mCAs and incident health outcomes with FDR<0.05 was considered significant. Analyses of incident events were performed separately in each biobank using the survival package in R (version 3.5, R Foundation). Meta-analyses of the mCAs results were performed using a fixed-effects model from the "meta" R package.

PheWAS in AoU included 1774 phecodes and 11 CHIP categories. Multivariable logistic regression was performed for outcome phecode and exposure CHIP trait adjusted for age, age^2^, genetic sex, genetic ancestry, first five genetic principal components, status of smoked 100 cigarettes lifetime, batch effects for WGS sequencing center and CHIP calling. To comply with AoU policy, the analysis only included cases where ≥40 participants had the phecode and CHIP cases.

### Genome-wide association study (GWAS)

Multi-ancestry GWAS for CHIP traits—including overall CHIP, expanded CHIP (VAF≥10%), *DNMT3A*, *TET2*, *ASXL1*, DNA damage response genes (DDR; *PPM1D*/*TP53*], and splicing factors (SF; *SF3B1/SRSF2/U2AF1/ZRSR2*)—were performed using REGENIE^70^ in UKB, AoU, TOPMed, and MGBB, and SAIGE^71^ in BioVU. CHIP status (VAF≥2% as case "1" and otherwise control "0") was modeled as the outcome in a logistic mixed model, adjusting for age at enrolment, age^2^, genetic sex, genetic ancestry, first ten genetic principal components, and genotyping and CHIP calling batch where applicable. TOPMed analyses were further adjusted for Study, Phase, and sequencing center. Only unrelated individuals (one per pair of first- or second-degree related individuals, or those with ≥ third-degree relatedness) with genotype missingness <10% and complete outcome/covariate data were included. GWAS was conducted in two steps using REGENIE: 1) null model construction via leave-one-out cross-validation (--loocv flag) using ∼500,000 directly genotyped SNV or corresponding variants from WGS (minor allele frequency ≥5%, genotype missingness <10%, Hardy-Weinberg equilibrium *P*>1×10^-15^), and 2) single-variant association testing using imputed/WGS variants with minor allele count ≥40 and INFO score ≥0.30. To account for case-control imbalance, Firth likelihood correction (--firth flag) was applied to variants with *P*<0.01.

We also performed ancestry- (AFR, AMR, EUR) and sex-stratified (male, female) GWAS within in UKB, AoU, TOPMed, and MGBB, adjusting for all covariates except genetic ancestry in ancestry-stratified analyses and genetic sex in sex-stratified analyses.

### Meta-analysis of GWAS

We meta-analyzed multi-ancestry and stratified GWAS results from UKB, AoU, TOPMed, MGBB, and BioVU cohorts using inverse-variance weighted fixed-effect meta-analysis and Cochran’s *Q*-test for heterogeneity in GWAMA software^72^. Additionally, multi-ancestry and AFR and AMR ancestry-stratified meta-analysis was performed including GWAS summary for overall CHIP, *DNMT3A*, *TET2*, and *ASXL1* CHIP from Mexico City Perspective Study (MCPS)^25^. Here, variants at MAF≥0.1% and present in ≥2 studies were included in the meta-analysis. Finally, variants were considered genome-wide significant at *P* <5.0 × 10^-8^, considering 1 million independent variants at a 5% significance level.

### MashR

To nominate potentially associated variants, we leveraged the shared signals across overall CHIP, *DNMT3A* CHIP, *TET2* CHIP, LTL^31^, expanded mCAs^32^, and MPN^30^ associated genome-wide significant loci (*P* < 5 × 10^-8^). We applied mashR (Multivariate Adaptive Shrinkage in R)^29^ to 6.7 million variants across the six traits using the exchangeable *Z* statistic model. Briefly, we fit the empirical Bayes prior by using the maximum SNP across each of the 1,703 linkage disequilibrium (LD) blocks specified in ^73^ and used a random sampling of 40,000 SNP to estimate the relative abundance of each pattern in the overall data set. Armed with this prior information, we then estimated the likelihood and computed posteriors on all variants. Variants with a local false sign rate (LFSR) <0.05 is considered significant. Here, LFSR is the probability that the effect is estimated with the incorrect sign and equivalent to the local false discovery rate^74^.

### Conditional and joint analysis

We performed conditional and joint (COJO)^75^ analyses on summary statistics from the multi-ancestry meta-analyses of overall CHIP, *DNMT3A*, and *TET2* CHIP GWAS to identify conditionally independent signals. Linkage disequilibrium (LD) was estimated using a multi-ancestry reference panel derived from 50k unrelated whole-genome sequenced (WGS) samples in the All of Us (AoU) v7 dataset, ensuring ancestry representation matched the GWAS meta-analysis (20.6% non-European, 79.4% European). The reference panel included individuals from diverse genetic backgrounds: 5,043 African, 3,303 Admixed American, 926 East Asian, 39,704 European, 52 Middle Eastern, and 975 South Asian.

We included variants with minor allele frequency (MAF) >0.1% and excluded those with allele frequency discrepancies >0.2 between GWAS summary statistics and reference samples. COJO analyses were performed by conditioning on the lead variant (*P*=5.0×10^-8^) from each chromosome, iteratively identifying independent variants (*P*=5.0×10^-8^) with LD threshold (*r^2^*<0.8) within a 10 Mb window. Finally, in cases where multiple independent variants were detected within the same chromosome, these were fitted in a final joint analysis to identify conditionally independent genome-wide significant variants (*P*=5.0×10^-8^).

### SNP-based heritability (*h^2^_SNP_*)

We estimated *h^2^_SNP_* using the summary statistics from meta-analysis of multi-ancestry GWAS of CHIP traits GWAS in UKB, AoU, TOPMed, and MGBB. We used the SumHer function of LDAK software^76,77^ with precomputed LD tagging prepared from HapMap3 UKB AFR sample (BLD-LDAK model: https://genetics.ghpc.au.dk/doug/TaggingFiles/bld.ldak.hapmap.afr.tagging.gz). The BLD-LDAK AFR LD tagging was prepared using 1.0-1.2M non-ambiguous HapMap3 SNP from 2,577 African individuals in UKB. Using ∼1.0M overlapping SNP from the summary statistics, *h^2^_SNP_* was estimated on a liability scale using average sample prevalence in GWAS cohorts (**Extended Data Table 3**).

### CHIP polygenic risk score (PRS)

To derive PRS, we used MegaPRS^78^ with precomputed HapMap3 European linkage disequilibrium (LD) tagging (https://genetics.ghpc.au.dk/doug/gbr.hapmap.tar.gz). CHIP PRS was calculated in UKB based on a multi-ancestry meta-analysis of CHIP GWAS conducted in AoU, TOPMed, and MGBB. The association between CHIP PRS (standardized to a mean of 0 and standard deviation of 1) and CHIP prevalence was evaluated using multivariable-adjusted logistic regression. Covariates included age, age^2^, genetic sex, genetic ancestry, smoking status, the first five genetic principal components, and genotyping and CHIP calling batch.

PheWAS of CHIP PRS in UKB: multivariable adjusted Cox regression was performed including covariates age, age^2^, genetic sex, genetic ancestry, smoking status, the first five genetic principal components, and genotyping and CHIP calling batch.

### Statistical fine-mapping

To infer putative causal variants in the associated loci, we conducted LD-informed statistical fine-mapping. We derived the LD-matrix of tested variants in each locus using TOPMed imputed UKB dosage of European ancestries by LDstore2 software (version 2.0)^79^. Using derived LD matrices, we applied statistical fine-mapping using the summary statistics obtained from the European meta-analysis (only included GWAS of EUR samples from UKB, TOPMed, AoU, MGBB, BioVU) by FINEMAP software (version 1.4)^27^. We only kept variants tested in at least 400,000 individuals to obtain consistent statistical power for each variant. FINEMAP’s shotgun stochastic search algorithm was used with a configuration of a maximum of 10 causal variants. A credible set with maximum posterior probability and variants with posterior inclusion probability (PIP)≥0.1 was reported for each locus.

### Polygenic prioritization of causal genes

We implemented PoPS to leverage the full genome-wide signal for nominating causal genes. Details of these methods have been described elsewhere^28^. Briefly, PoPS is a novel similarity-based gene prioritization approach that assesses the polygenic enrichments of gene features, including cell-type-specific gene expression, protein-protein interaction networks, and biological pathways, through training linear models to predict gene-level association scores from those features and converting the gene p-values from the linear models to *Z*-scores that reflect the confidence on its causal role to the given locus. We used the summary statistics of multi-ancestry meta-analysis of CHIP, *DNMT3A*, and *TET2* GWAS and LD reference panel of European ancestry individuals from the 1000 Genomes Project (phase 3). In total, 57,543 features were considered for analysis, and those who passed marginal feature selection were carried forward to the linear models for computation. We computed a PoPS score for all protein-coding genes within a defined 500kb window around each of the significant genomic regions. We prioritized the gene with the PoPS score >0.50 in each locus.

### CHIP associations at the cell-type level

scDRS group analysis was used to assess the polygenic associations of CHIP, *DNMT3A*, and *TET2* at cell type and tissue level. Details of the methods have been described elsewhere^33^. Briefly, scDRS is a novel statistical method that associates individual cells in a scRNA-seq data to a trait GWAS based on the aggregate expression across a set of putative GWAS trait genes, assessing statistical significance using appropriately matched control genes; furthermore, the scDRS group analysis computes a p-value for a cell group (e.g., cell type or tissue) based on the association of cells within the given cell group. Following the scDRS guideline, the putative trait gene sets were constructed as the top 1,000 MAGMA genes, where MAGMA^80^, an existing gene-scoring method, was applied to GWASs from UKB participants of European ancestry using a set of 489 unrelated individuals of European ancestry from phase 3 1000 Genomes Project as an LD reference panel^81^ . scRNA-seq data TS human cell atlas^40^ (FACS data with 26,813 cells, 24 tissues, and 68 cell types with more than 50 cells) was used for this analysis. Multiple testing correlation (FDR) was applied to each trait separately across all cell types (68 for TS) or across all tissues (24 for TS). To increase power, associations were considered significant at FDR<0.2, and all suggestive associations with *P*<0.01 were reported. Sensitivity analyses were conducted by repeating the analysis using down-sampled scRNA-seq data (to 50K cells) and down-sampled putative trait gene sets (to 500 genes) separately, each with 20 repetitions.

## Supporting information

Supplemental Author List

## Data availability

The meta-GWAS summary statistics will be publicly available through the Common Metabolic Diseases Knowledge Portal (https://hugeamp.org/). TOPMed individual-level genotype and phenotype data are available through restricted access via the dbGaP. The genotypes and phenotypes of UKB and AoU participants are available by application to the UKB (https://www.ukbiobank.ac.uk/register-apply/) and AoU (https://allofus.nih.gov/), respectively. Publicly funded genotyping and exome sequencing data for 13,500 participants from the MGBB are available in dbGAP (https://www.ncbi.nlm.nih.gov/projects/gap/cgi-bin/study.cgi?study_id=phs002018.v1.p1). For access to the full MGBB data and potential collaboration, the researchers can contact MGBB team online (https://www.massgeneralbrigham.org/en/research-andinnovation/participate-in-research/biobank/for-researchers).

## Code availability

The codes used to generate data in main and supplemental tables and figures are publicly available via GitHub at https://github.com/MMesbahU/meta-gwas-of-clonal-hematopoiesis. CHIP calling pipeline is available at https://app.terra.bio/#workspaces/terra-outreach/CHIP-Detection-Mutect, and the custom script for mutations in *U2AF1* is available at https://github.com/MMesbahU/U2AF1_pileup.

## Acknowledgments

We thank the investigators and their studies for contributing samples and/or data to the current work and the participants in those studies who made this research possible. This research has been conducted using the UKB Resource under Application Number 7089 and 50834. This research is based on data from the BioVU, which is supported by NIH Grant UL1 RR024975-01and UL1 TR000445-06. This research is also based on data from the Million Veteran Program, Office of Research and Development, Veterans Health Administration, and was supported by Million Veteran Program-MVP000 (dbGaP accession: phs001672). This publication does not represent the views of the Department of Veteran Affairs or the United States Government.

## TOPMed

We gratefully acknowledge the studies and participants who provided biological samples and data for TOPMed. The WHI program is funded by the National Heart, Lung, and Blood Institute, National Institutes of Health, U.S. Department of Health and Human Services through contracts HHSN268201600018C, HHSN268201600001C, HHSN268201600002C, HHSN268201600003C, and HHSN268201600004C. The COPDGene project described was supported by Award Number U01 HL089897 and Award Number U01 HL089856 from the National Heart, Lung, and Blood Institute. The content is solely the responsibility of the authors and does not necessarily represent the official views of the National Heart, Lung, and Blood Institute or the National Institutes of Health. Whole genome sequencing (WGS) for the Trans-Omics in Precision Medicine (TOPMed) program was supported by the National Heart, Lung and Blood Institute (NHLBI). WGS for “NHLBI TOPMed: Multi-Ethnic Study of Atherosclerosis (MESA)” (phs001416.v1.p1) was performed at the Broad Institute of MIT and Harvard (3U54HG003067-13S1). Centralized read mapping and genotype calling, along with variant quality metrics and filtering were provided by the TOPMed Informatics Research Center (3R01HL-117626-02S1). Phenotype harmonization, data management, sample-identity QC, and general study coordination, were provided by the TOPMed Data Coordinating Center (3R01HL-120393-02S1). The MESA projects are conducted and supported by the National Heart, Lung, and Blood Institute (NHLBI) in collaboration with MESA investigators. Support for MESA is provided by contracts 75N92025D00022, 75N92020D00001, HHSN268201500003I, N01-HC-95159, 75N92025D00026, 75N92020D00005, N01-HC-95160, 75N92020D00002, N01-HC-95161, 75N92025D00024, 75N92020D00003, N01-HC-95162, 75N92025D00027, 75N92020D00006, N01-HC-95163, 75N92025D00025, 75N92020D00004, N01-HC-95164, 75N92025D00028, 75N92020D00007, N01-HC-95165, N01-HC-95166, N01-HC-95167, N01-HC-95168, N01-HC-95169, UL1-TR-000040, UL1-TR-001079, UL1-TR-001420, UL1TR001881, and R01HL105756. The authors thank the MESA participants and the MESA investigators and staff for their valuable contributions. A full list of participating MESA investigators and institutions can be found at http://www.mesa-nhlbi.org. Cardiovascular Health Study (CHS) is supported by contracts 75N92021D00006, HHSN268201200036C, HHSN268200800007C, HHSN268201800001C, N01-HC85079, N01-HC-85080, N01-HC-85081, N01-HC-85082, N01-HC-85083, N01-HC-85084, N01-HC-85085, N01-HC-85086, N01-HC-35129, N01-HC-15103, N01-HC-55222, N01-HC-75150, N01-HC-45133, and N01-HC-85239; grant numbers R01HL172803, U01 HL080295, U01 HL130114 and R01 HL059367 from the National Heart, Lung, and Blood Institute, and R01 AG023629 from the National Institute on Aging, with additional contribution from the National Institute of Neurological Disorders and Stroke. GeneSTAR was supported by the National Institutes of Health/National Heart, Lung, and Blood Institute (U01 HL72518, HL087698, HL112064) and by a grant from the National Institutes of Health/National Center for Research Resources (M01-RR000052) to the Johns Hopkins General Clinical Research Center. The COPDGene project is also supported by the COPD Foundation through contributions made to an Industry Advisory Board comprised of AstraZeneca, Boehringer Ingelheim, GlaxoSmithKline, Novartis, Pfizer, Siemens and Sunovion. The My Life, Our Future (MLOF) samples and data are made possible through the partnership of Bloodworks Northwest, the American Thrombosis and Hemostasis Network, the National Hemophilia Foundation, and Bioverativ. Framingham Heart Study (FHS) acknowledges the support of contracts NO1-HC-25195, HHSN268201500001I, 75N92025D00012 and 75N92019D00031 from the National Heart, Lung and Blood Institute and grant supplement R01 HL092577-06S1 for this research. We also acknowledge the dedication of the FHS study participants without whom this research would not be possible. The Mount Sinai BioMe Biobank has been supported by The Andrea and Charles Bronfman Philanthropies and in part by Federal funds from the NHLBI and NHGRI (U01HG00638001; U01HG007417; X01HL134588). We thank all participants in the Mount Sinai Biobank. We also thank all our recruiters who have assisted and continue to assist in data collection and management and are grateful for the computational resources and staff expertise provided by Scientific Computing at the Icahn School of Medicine at Mount Sinai. The Atherosclerosis Risk in Communities study (ARIC) has been funded in whole or in part with Federal funds from the National Heart, Lung, and Blood Institute, National Institutes of Health, Department of Health and Human Services (contract numbers HHSN268201700001I, HHSN268201700002I, HHSN268201700003I, HHSN268201700004I and HHSN268201700005I). The Jackson Heart Study (JHS) is supported and conducted in collaboration with Jackson State University (HHSN268201300049C and HHSN268201300050C), Tougaloo College (HHSN268201300048C), and the University of Mississippi Medical Center (HHSN268201300046C and HHSN268201300047C) contracts from the National Heart, Lung, and Blood Institute (NHLBI) and the National Institute for Minority Health and Health Disparities (NIMHD). The Coronary Artery Risk Development in Young Adults Study (CARDIA) is conducted and supported by the National Heart, Lung, and Blood Institute (NHLBI) in collaboration with the University of Alabama at Birmingham (HHSN268201300025C & HHSN268201300026C), Northwestern University (HHSN268201300027C), University of Minnesota (HHSN268201300028C), Kaiser Foundation Research Institute (HHSN268201300029C), and Johns Hopkins University School of Medicine (HHSN268200900041C). CARDIA is also partially supported by the Intramural Research Program of the National Institute on Aging (NIA) and an intra-agency agreement between NIA and NHLBI (AG0005). The SAPPHIRE study is supported by the Fund for Henry Ford Hospital, the American Asthma Foundation, the National Heart Lung and Blood Institute (R01HL118267, X01HL134589), the National Institute of Allergy and Infectious Diseases (R01AI079139), and the National Institute of Diabetes and Digestive and Kidney Diseases (R01DK113003). The Rare Variants for Hypertension in Taiwan Chinese (THRV) is supported by the National Heart, Lung, and Blood Institute (NHLBI) grant (R01HL111249) and its participation in TOPMed is supported by an NHLBI supplement (R01HL111249-04S1). SAPPHIRe was supported by NHLBI grants (U01HL54527, U01HL54498) and Taiwan funds, and the other cohorts were supported by Taiwan funds.

The Genetic Epidemiology Network of Salt-Sensitivity (GenSalt) was supported by research grants (U01HL072507, R01HL087263, and R01HL090682) from the NHLBI. Collection of the San Antonio Family Study (SAFS) data was supported in part by National Institutes of Health (NIH) grants R01 HL045522, MH078143, MH078111 and MH083824; and whole genome sequencing of SAFS subjects was supported by U01 DK085524 and R01 HL113323. The HyperGEN Study is part of the National Heart, Lung, and Blood Institute (NHLBI) Family Blood Pressure Program; collection of the data represented here was supported by grants U01 HL054472 (MN Lab), U01 HL054473 (DCC), U01 HL054495 (AL FC), and U01 HL054509 (NC FC). The HyperGEN: Genetics of Left Ventricular Hypertrophy Study was supported by NHLBI grant R01 HL055673 with whole-genome sequencing made possible by supplement -18S1. The Hispanic Community Health Study/Study of Latinos (HCHS_SOL) is a collaborative study supported by contracts from the National Heart, Lung, and Blood Institute (NHLBI) to the University of North Carolina (HHSN268201300001I / N01-HC-65233), University of Miami (HHSN268201300004I / N01-HC-65234), Albert Einstein College of Medicine (HHSN268201300002I / N01-HC-65235), University of Illinois at Chicago – HHSN268201300003I / N01-HC-65236 Northwestern Univ), and San Diego State University (HHSN268201300005I / N01-HC-65237). The following Institutes/Centers/Offices have contributed to the HCHS/SOL through a transfer of funds to the NHLBI: National Institute on Minority Health and Health Disparities, National Institute on Deafness and Other Communication Disorders, National Institute of Dental and Craniofacial Research, National Institute of Diabetes and Digestive and Kidney Diseases, National Institute of Neurological Disorders and Stroke, NIH Institution-Office of Dietary Supplements. The ECLIPSE study (NCT00292552) was sponsored by GlaxoSmithKline. Mayo Clinic Venous Thromboembolism Study (Mayo_VTE)) is funded by NHLBI grants HL66216 and HL83141 and NHGRI grants HG04735, HG06379, and research support provided by Mayo Foundation. Samoan Adiposity Study date collection was funded by NIH grant R01-HL093093. Support for GENOA was provided by the National Heart, Lung and Blood Institute (HL054457, HL054464, HL054481, and HL087660) of the National Institutes of Health. The TOPMed component of the Amish Research Program was supported by NIH grants R01 HL121007, U01 HL072515, and R01 AG18728. Funding for BAGS was provided by National Institutes of Health (NIH) R01HL104608, R01HL087699, and HL104608 S1. Cleveland Family Study (CFS) is supported by grant numbers-HL 046389; HL113338;1R35HL135818. GOLDN is supported by U01 HL072524. Whole-genome sequencing in GOLDN was funded by NHLBI grant R01 HL104135 and supplement R01 HL104135-04S1. The Heart and Vascular Health (HVH) Study was supported by grants HL068986, HL085251, HL095080, and HL073410 from the National Heart, Lung, and Blood Institute. The Boston Early-Onset COPD (EOCOPD) Study was supported by R01 HL113264 and U01 HL089856 from the National Heart, Lung, and Blood Institute. The Diabetes Heart Study (DHS) was supported by R01 HL92301, R01 HL67348, R01 NS058700, R01 AR48797, R01 DK071891, R01 AG058921, the General Clinical Research Center of the Wake Forest University School of Medicine (M01 RR07122, F32 HL085989), the American Diabetes Association, and a pilot grant from the Claude Pepper Older Americans Independence Center of Wake Forest University Health Sciences (P60 AG10484). The Women’s Genome Health Study (WGHS) is supported by the National Heart, Lung, and Blood Institute (HL043851 and HL080467) and the National Cancer Institute (CA047988 and UM1CA182913). The most recent cardiovascular endpoints were supported by ARRA funding HL099355.

## Author Contributions

A.G.B. and P.N. designed and conceived the study. A.G.B. and P.N. supervised the study. M.M.U., Z.Y., J.S.W., A.N., C.V., S.K., T.N., S.M.U., S.M.Z., K.P., B.T., A.J.S., T.M.M., M.Y.W., S.M.H., R.B., S.D.J., M.A.R., M.C.H., W.E.H., M.J.Z., J.D., J.B.H., V.G.S., G.K.G., C.J.G., H.A.A, K.C., Y.V.S., S.P., P.W.F.W., G.G, Y.X., M.R.S., Y-D.I.C., W.H-H.S., Y-J.H., L.M.R., B.A.H., M.F., R.C.K., K.L.W., D.K.A., M.P., B.A.K., S.K., P.A.P., J.A.S., R.A.M., L.R.Y., L.C.B., M.d.A., D.D., J.H., J.B., S.T.W., B.C., D.A.M., R.L.M., T.N., S.V., B.D.M., K.C.B., S.R., D.S., B.M.S., N.D.P., B.I.F., D.W.B., C.M.A., R.K., N.S., J.F., R.T., J.C.B., B.M.P., K.D.T., S.S.R., J.I.R, E.B., V.S.R., S.L., P.T.E., Z.W., R.J.F.L., J.H.Y., M.H.C., E.K.S., G.A., C.K., P.L.A., A.P.R., S.J., B.L.E., A.B., and P.N. acquired, analyzed or interpreted the data. M.M.U., Z.Y., T.N., S.M.U., S.K., S.M.Z, A.B., and P.N. drafted the manuscript. All authors critically reviewed the manuscript.

