## Extended Data Figures 1-30 for "Genomic and phenomic landscape of clonal hematopoiesis in over a million ancestrally diverse participants"

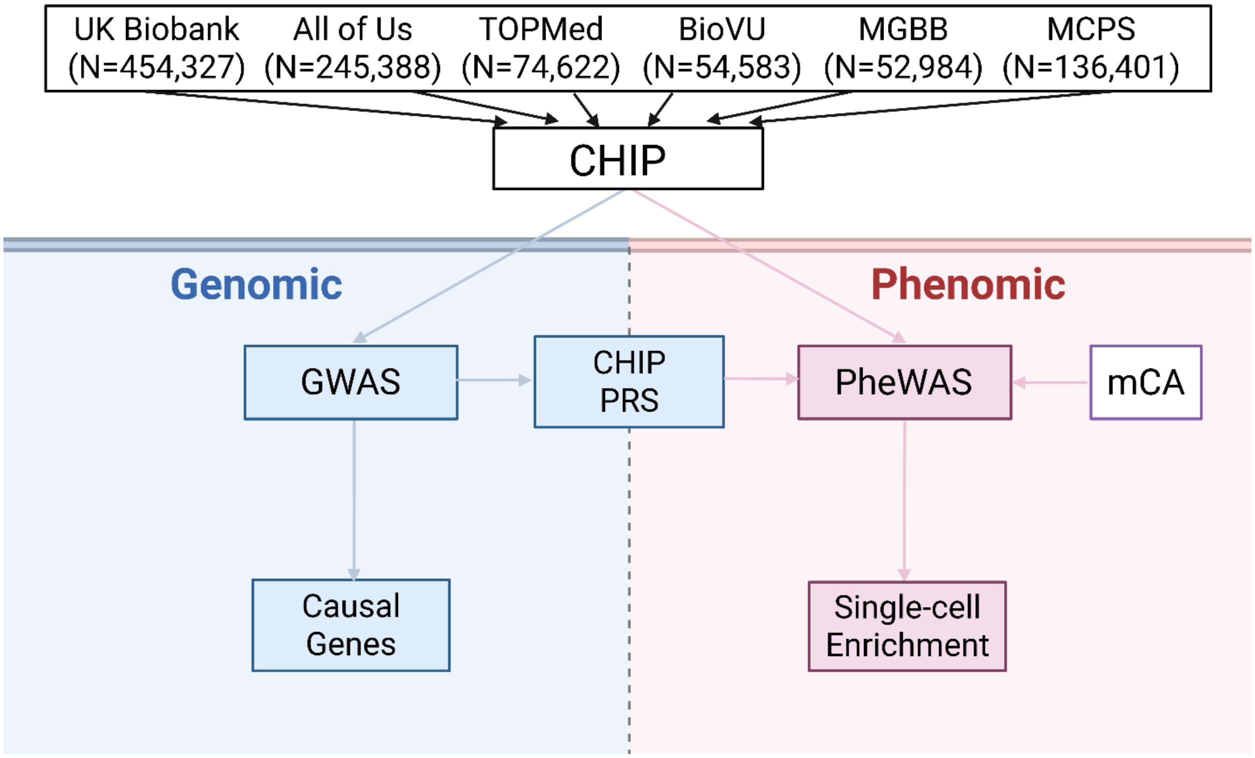

**Extended Data Figure 1 | Flowchart of the study.** CHIP: clonal hematopoiesis of indeterminate potential; mCA: mosaic chromosomal alterations; MGBB: Mass General Brigham Biobank; TOPMed: Trans-Omics for Precision Medicine; PRS: polygenic risk score; PheWAS: phenome-wide association study; BioVU: Vanderbilt BioVU; MCPS: Mexico City Prospective Study.

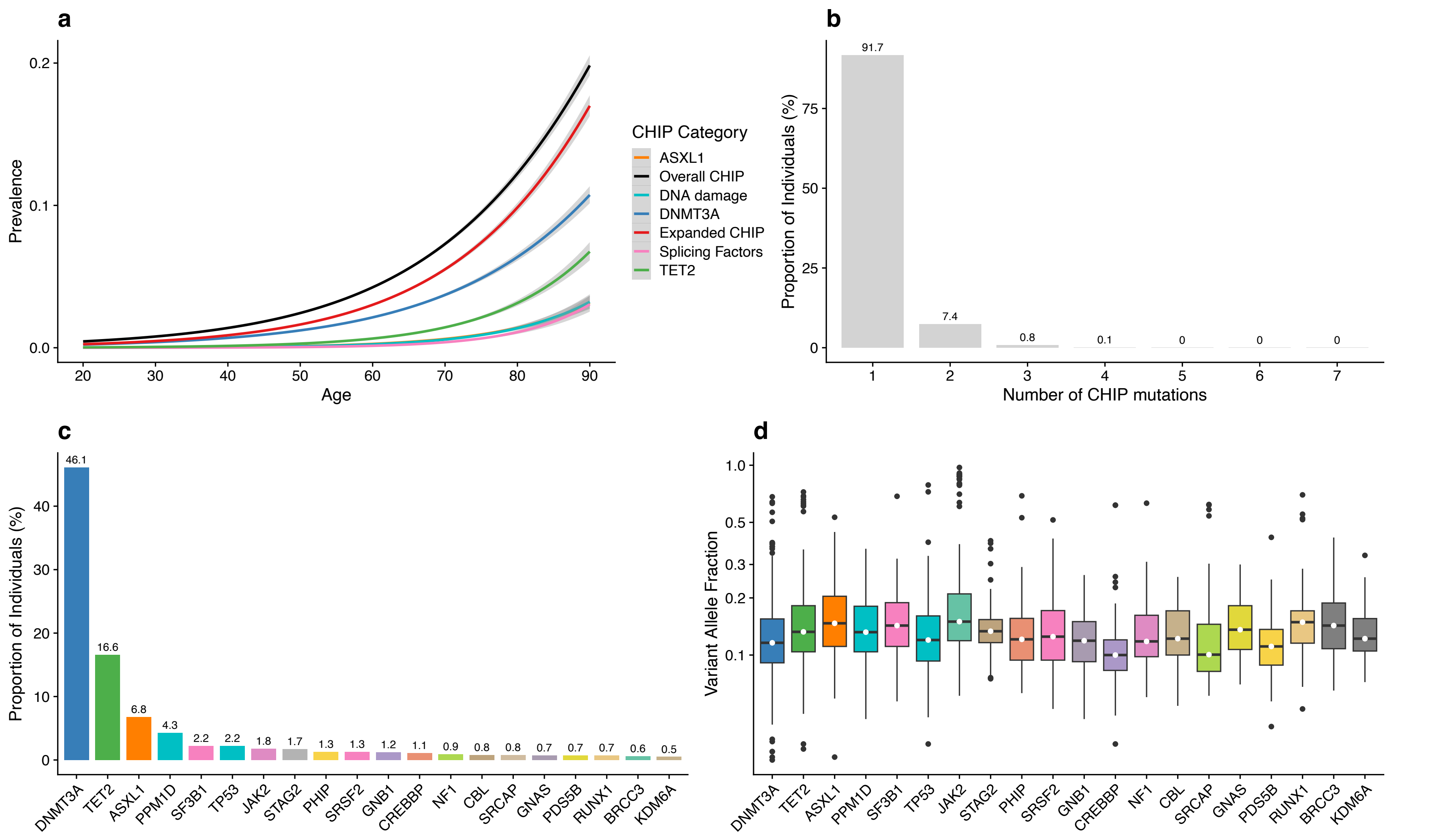

**Extended Data Figure 2 | Distribution of CHIP in the All of Us Research cohort (AoU; N=245,388).** a) Prevalence of CHIP in AoU. The center line represents the general additive model spline, and the shaded region is the 95% confidence interval. Here, 9737 CHIP, 7043 expanded CHIP (VAF>=10%), 4751 *DNMT3A*, 1682 *TET2*, 721 *ASXL1*, 460 *SF*, and 670 *DDR* cases vs 235651 controls. b) More than 90% of individuals with CHIP had only one somatic CHIP driver mutation. c) Proportion of individuals carrying a driver mutation in frequently mutated CHIP genes. d) Distribution of CHIP clones (variant allele fraction) by drivers. Boxplot spanning minimum and maximum values with median variant allele fraction highlighted by white circles. CHIP: clonal hematopoiesis of indeterminate potential; DDR: DNA damage response genes (TP53/PPM1D); SF: splicing factors (SF3B1/SRSF2/U2AF1/ZRSR2).

a) Overall CHIP (EAF≥0.1%)

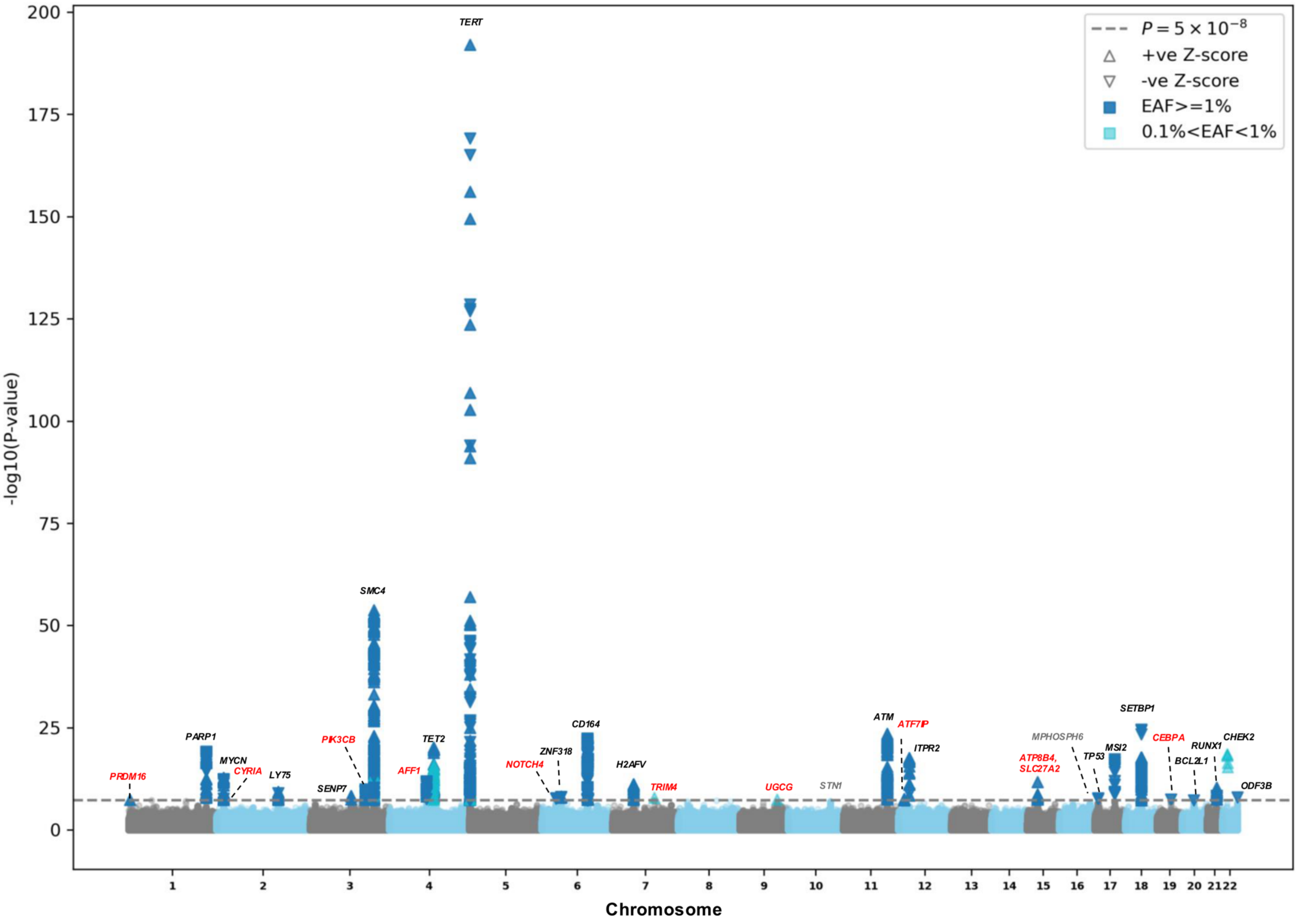

b) Expanded CHIP (VAF≥10%)

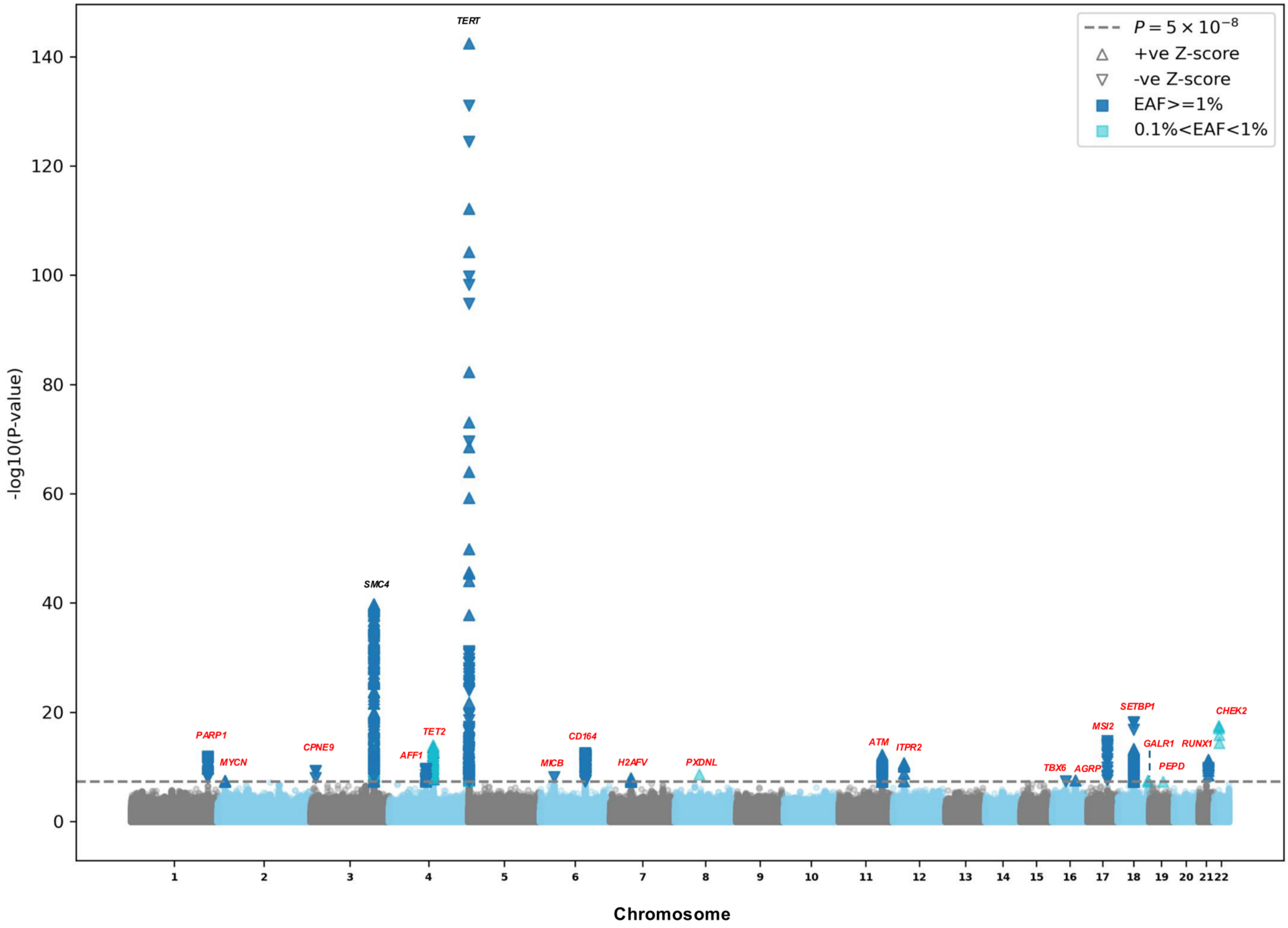

c) *DNMT3A* (EAF≥0.1%)

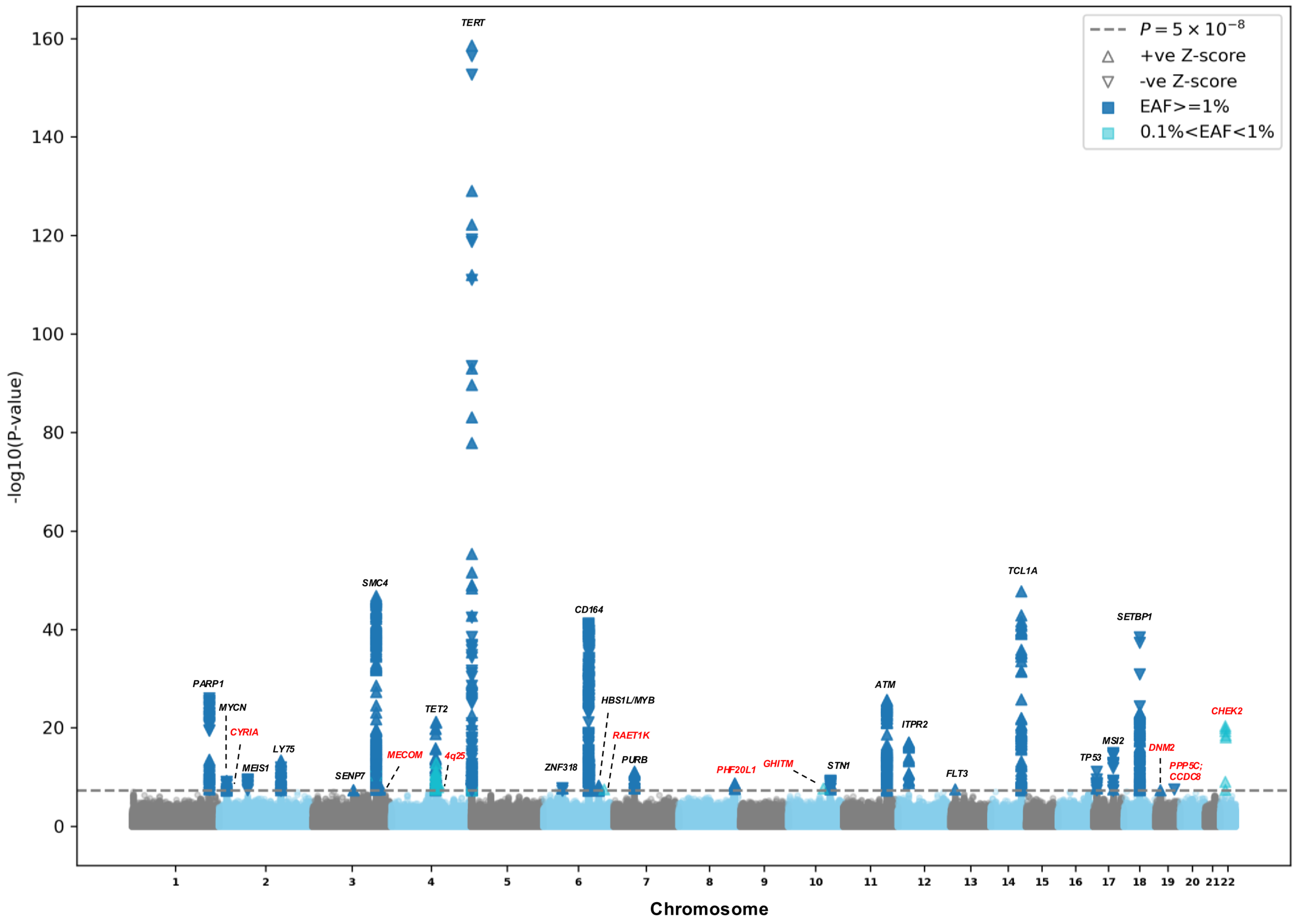

d) *TET2* (EAF≥0.2%)

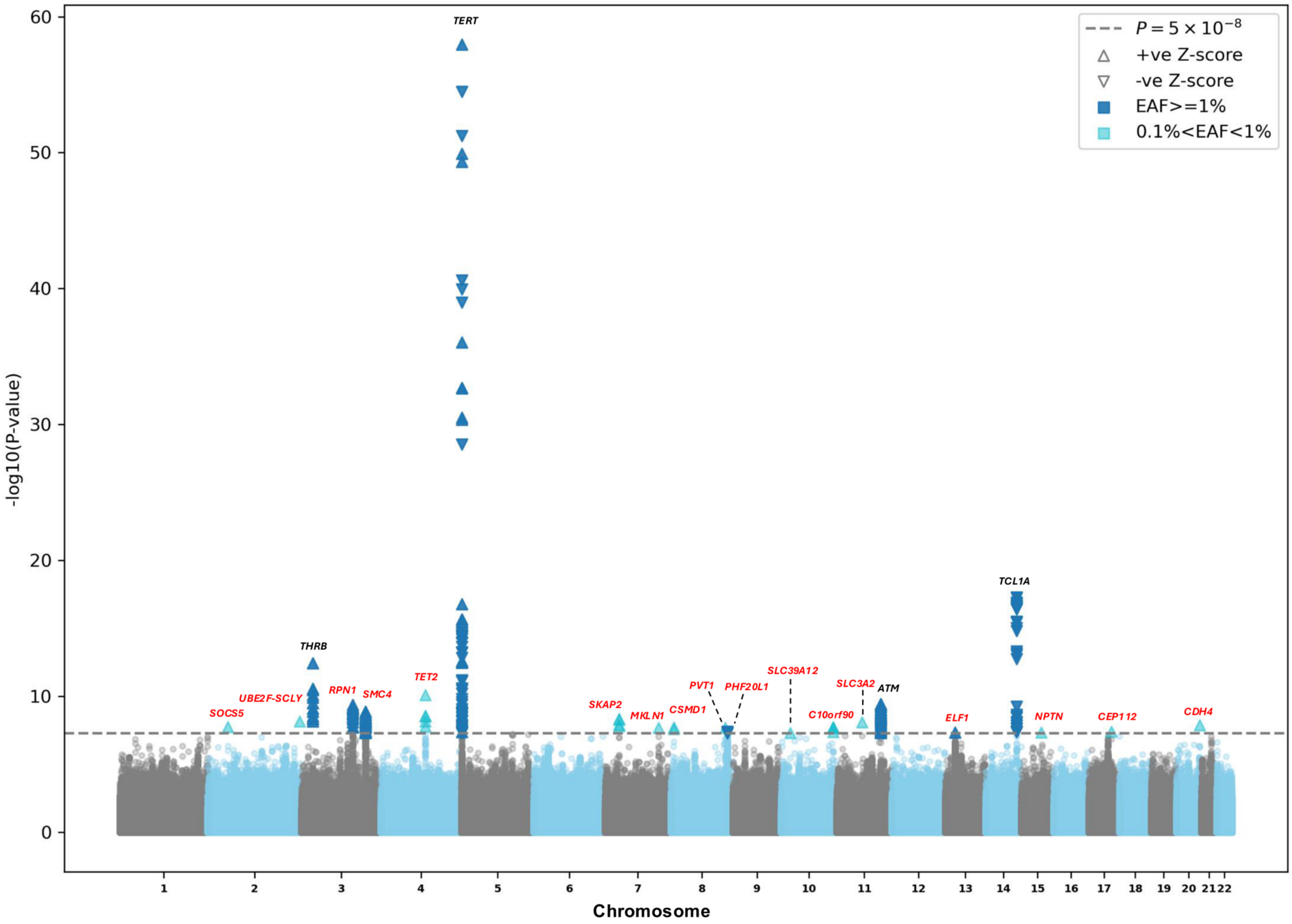

TERT
THRB
TCL1A
TET2
SOCS5
UBE2F-SCLY
RPN1
SMC4
SKAP2
MKLN1
CSMD1
PVT1
PHF20L1
SLC39A12
C10orf90
SLC3A2
ATM
ELF1
NPTN
CEP112
CDH4
P = 5 × 10^-8
+ve Z-score
-ve Z-score
EAF>=1%
0.1%<EAF<1%
-log10(P-value)
Chromosome
0
10
20
30
40
50
60

e) *ASXL1* (EAF≥0.5%)

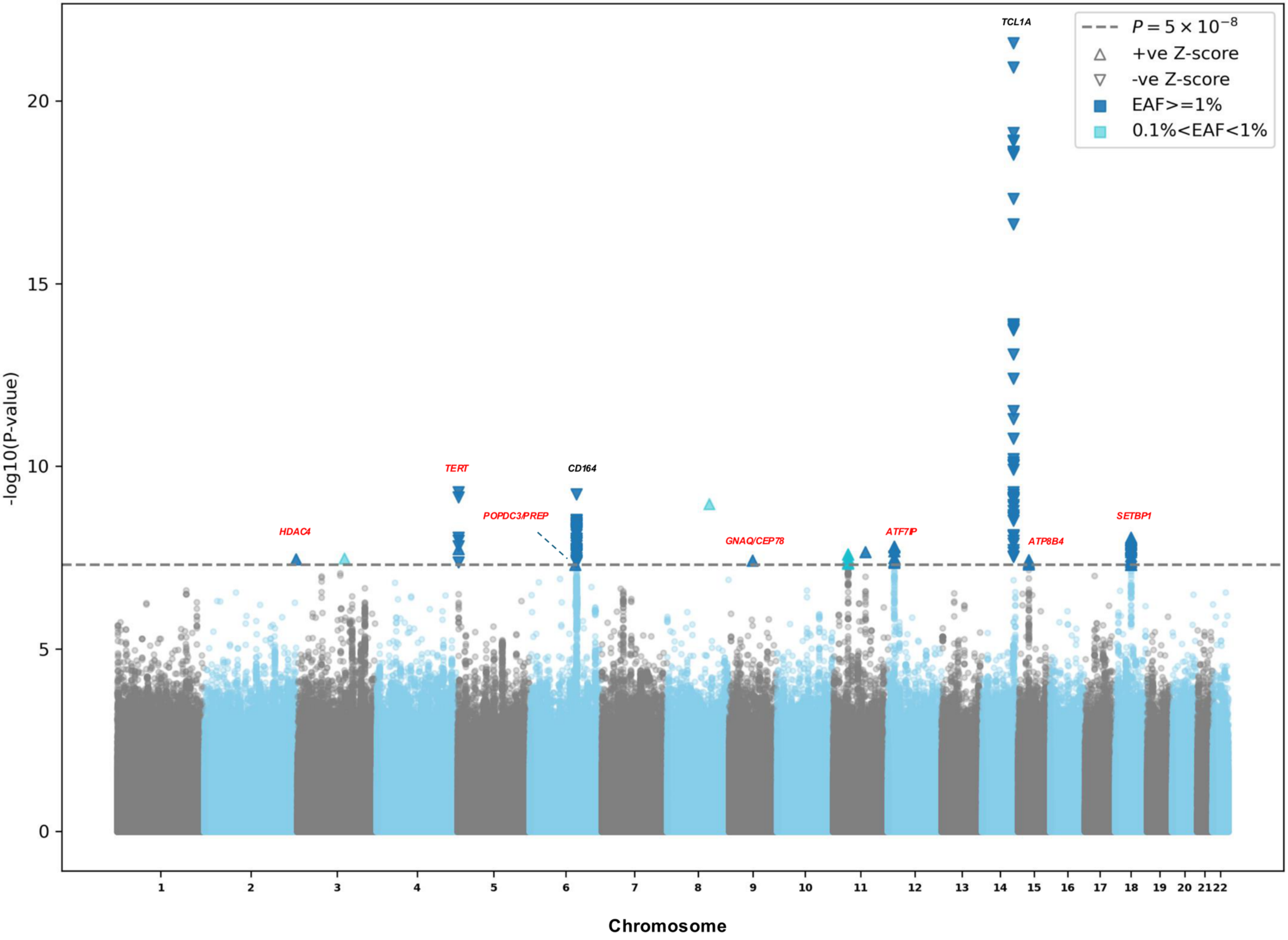

**f) *ASXL1 (with MCPS)***

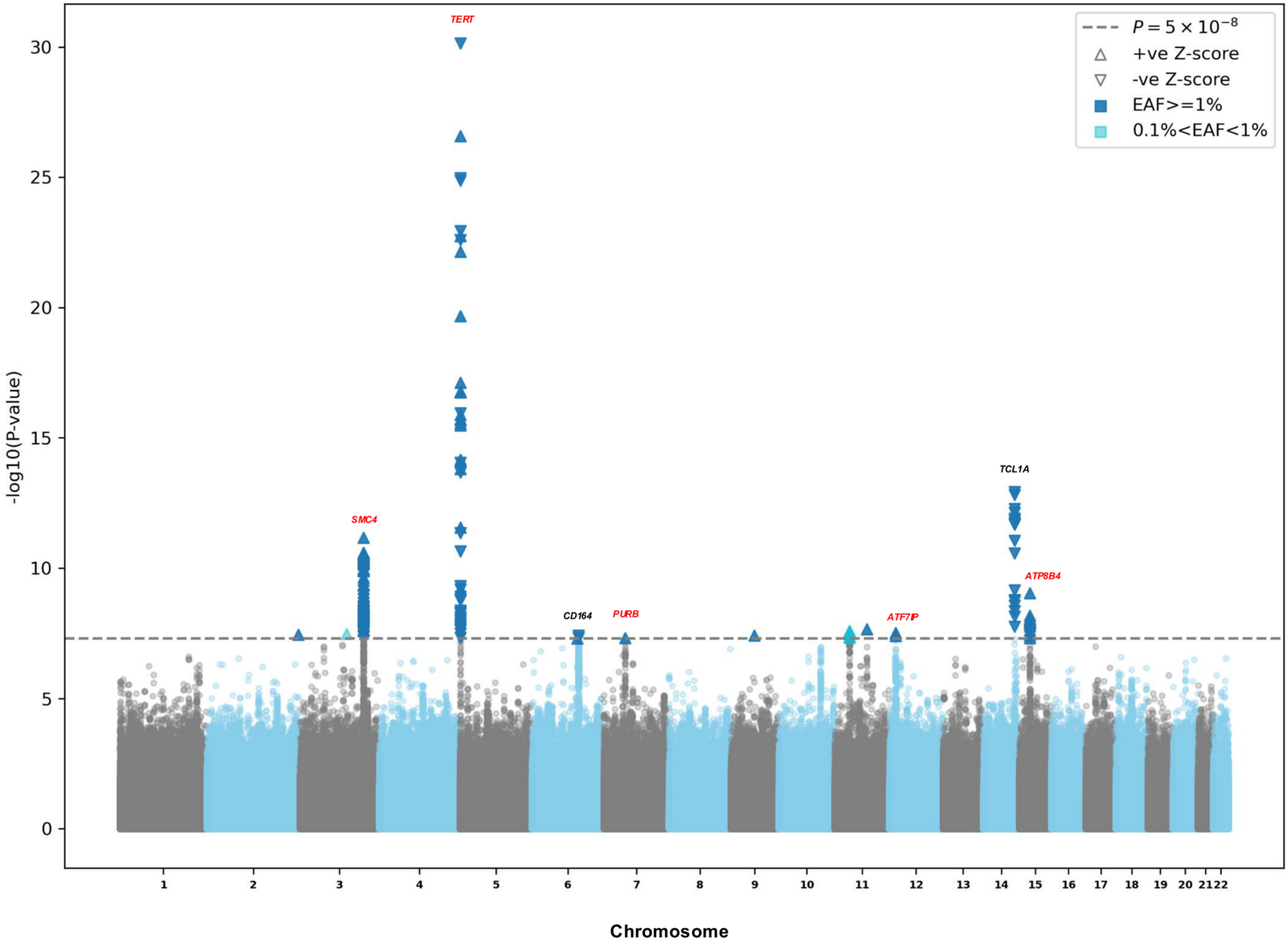

g) DDR

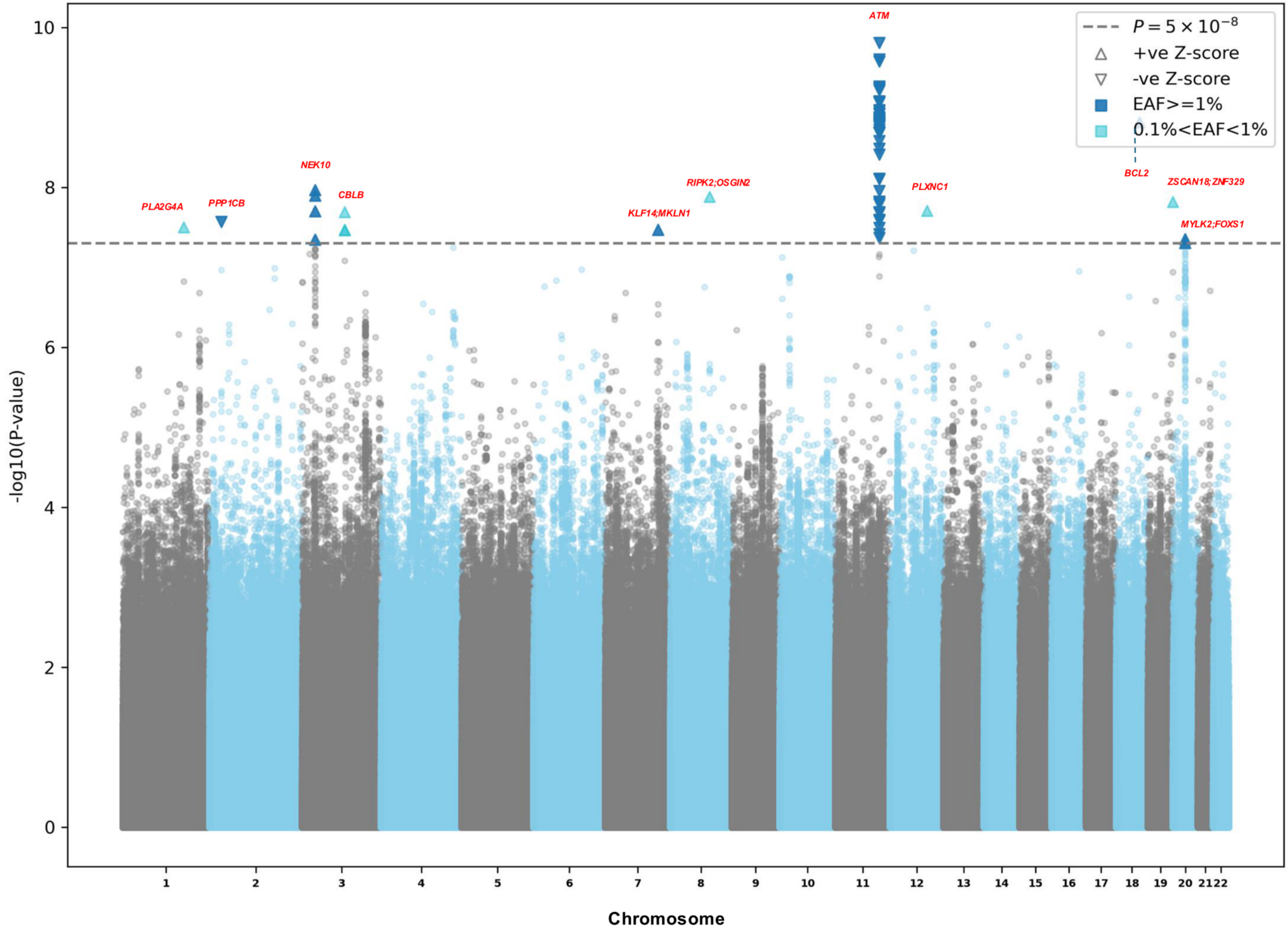

ATM
NEK10
PLA2G4A
PPP1CB
CBLB
KLF14;MKLN1
RIPK2;OSGIN2
PLXNC1
BCL2
ZSCAN18;ZNF329
MYLK2;FOXS1
P = 5 × 10^-8
+ve Z-score
-ve Z-score
EAF>=1%
0.1%<EAF<1%
-log10(P-value)
0
2
4
6
8
10
Chromosome

h) SF (EAF>0.5%)

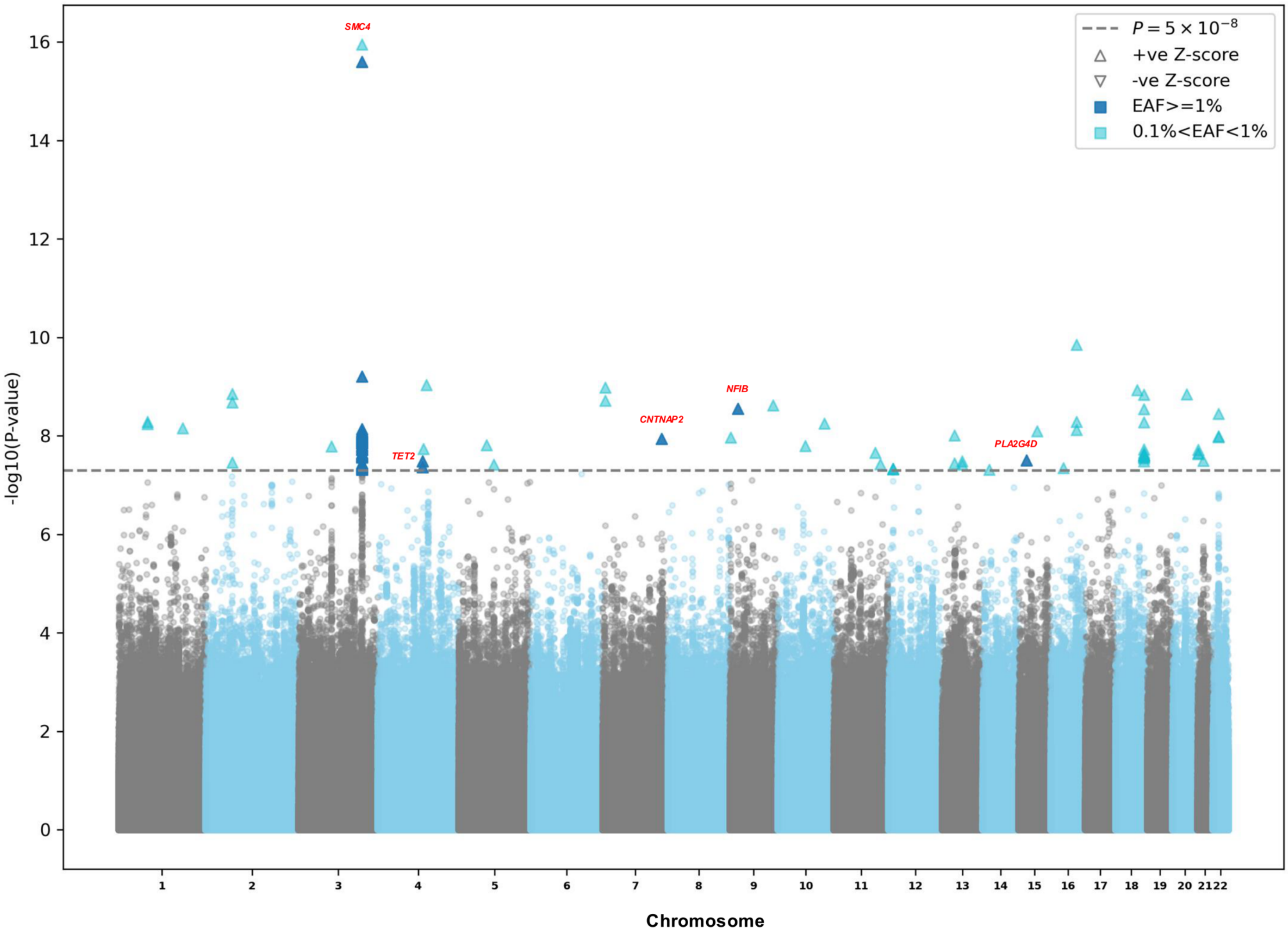

SMC4
TET2
CNTNAP2
NFIB
PLA2G4D
$P = 5 \times 10^{-8}$
+ve Z-score
-ve Z-score
EAF>=1%
0.1%<EAF<1%
-log10(P-value)
16
14
12
10
8
6
4
2
Chromosome

i) SF (*with MCPS*)

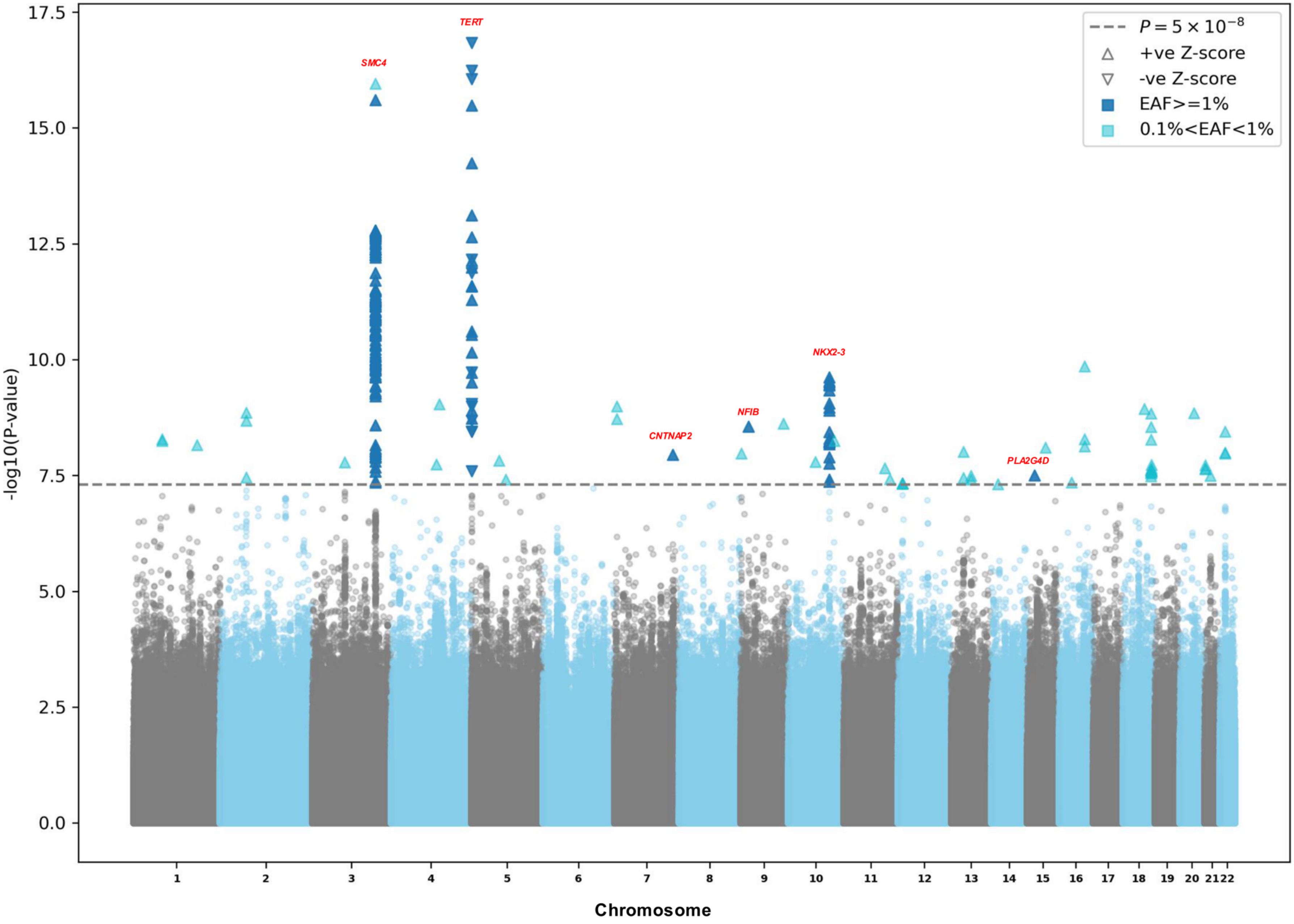

**Extended Data Figure 3 | Manhattan plots for multi-ancestry CHIP GWAS meta-analyses.** a) Manhattan plot for overall CHIP GWAS in UKB, AoU, TOPMed, MGBB and BioVU (N=838,781). b) Manhattan plot for expanded CHIP (VAF≥10%) GWAS in UKB, AoU, TOPMed, and MGBB (N= 779,016). c) Manhattan plot for *DNMT3A* GWAS in UKB, AoU, TOPMed, MGBB and BioVU (N= 816,623). d) Manhattan plot for *TET2* GWAS in UKB, AoU, TOPMed, MGBB and BioVU (N=800,453). e) Manhattan plot for *ASXL1* GWAS in UKB, AoU, TOPMed, and MGBB (N=741,305). f) Manhattan plot for *ASXL1* GWAS in UKB, AoU, TOPMed, MGBB, and MCPS (N=877,706). g) Manhattan plot for DDR GWAS in UKB, AoU, TOPMed, and MGBB (N=740,114). h) Manhattan plot for SF GWAS in UKB, AoU, TOPMed, and MGBB (N=738,868). i) Manhattan plot for SF GWAS in UKB, AoU, TOPMed, MGBB and MCPS (N=875,269). EAF: effect allele frequency; UKB: UK biobank; AoU: All of Us Research; TOPMed: Trans-omics for Precision medicine; MGBB: Mass General and Brigham biobank; BioVU: Vanderbilt BioVU; MCPS: Mexico City Prospective Study; CHIP: clonal hematopoiesis of indeterminate potential; DDR: DNA damage response genes (*TP53/PPM1D*); SF: splicing factors (*SF3B1/SRSF2/U2AF1/ZRSR2*).

a)

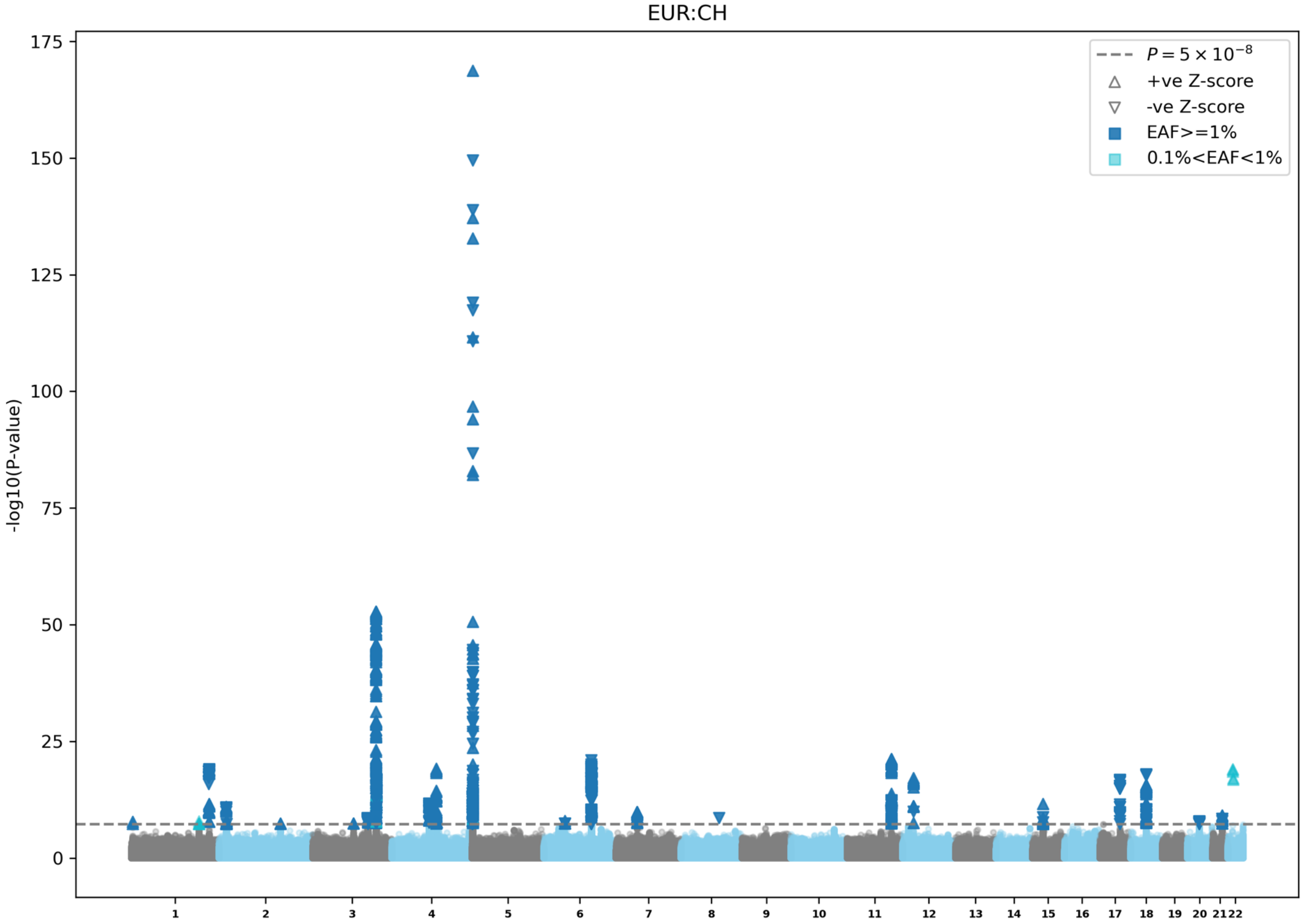
EUR:CH
-log10(P-value)
0
25
50
75
100
125
150
175
$P = 5 \times 10^{-8}$
+ve Z-score
-ve Z-score
EAF>=1%
0.1%<EAF<1%

b)

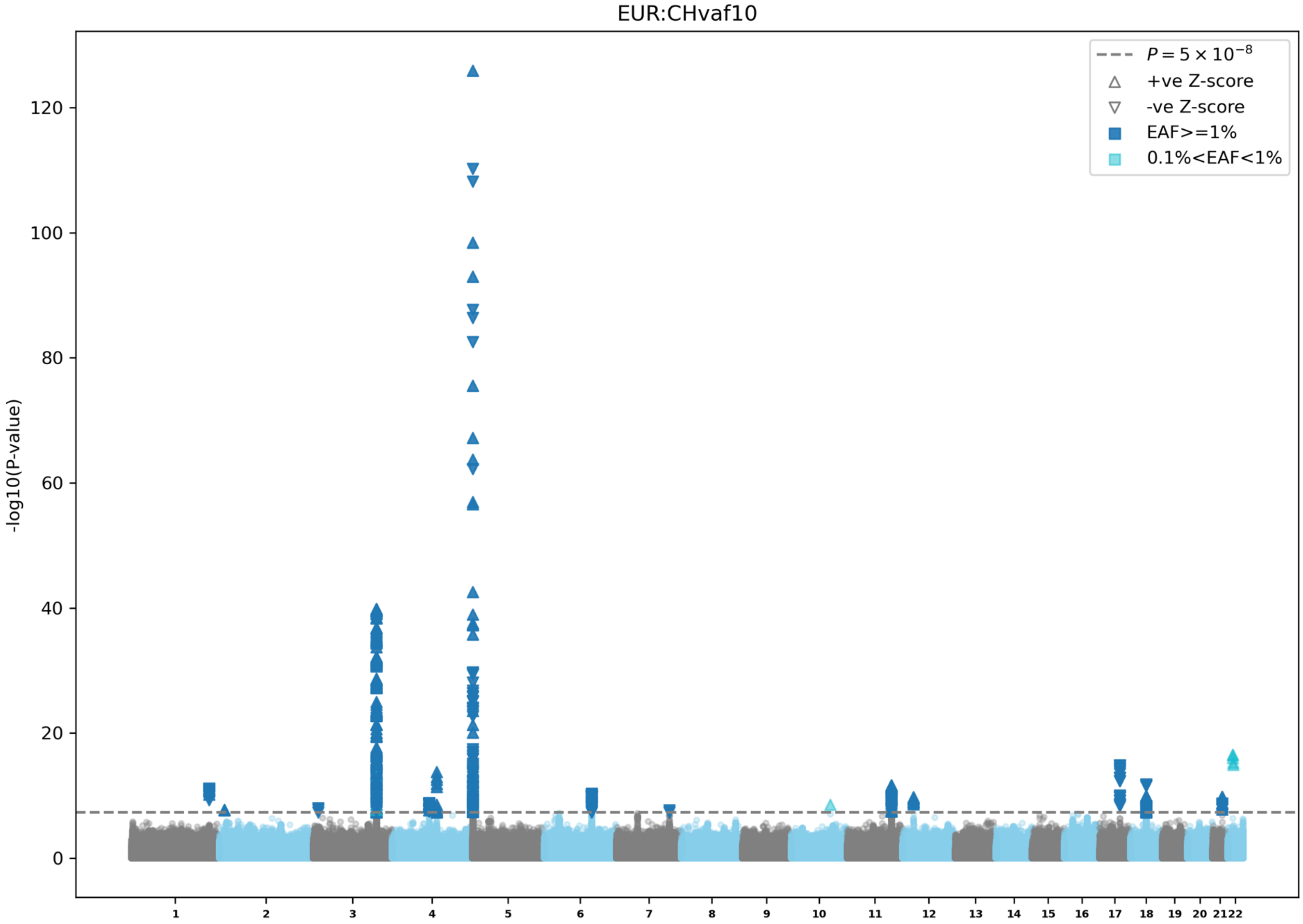

EUR:CHvaf10
$P = 5 \times 10^{-8}$
+ve Z-score
-ve Z-score
EAF>=1%
0.1%<EAF<1%
-log10(P-value)
0
20
40
60
80
100
120

c)

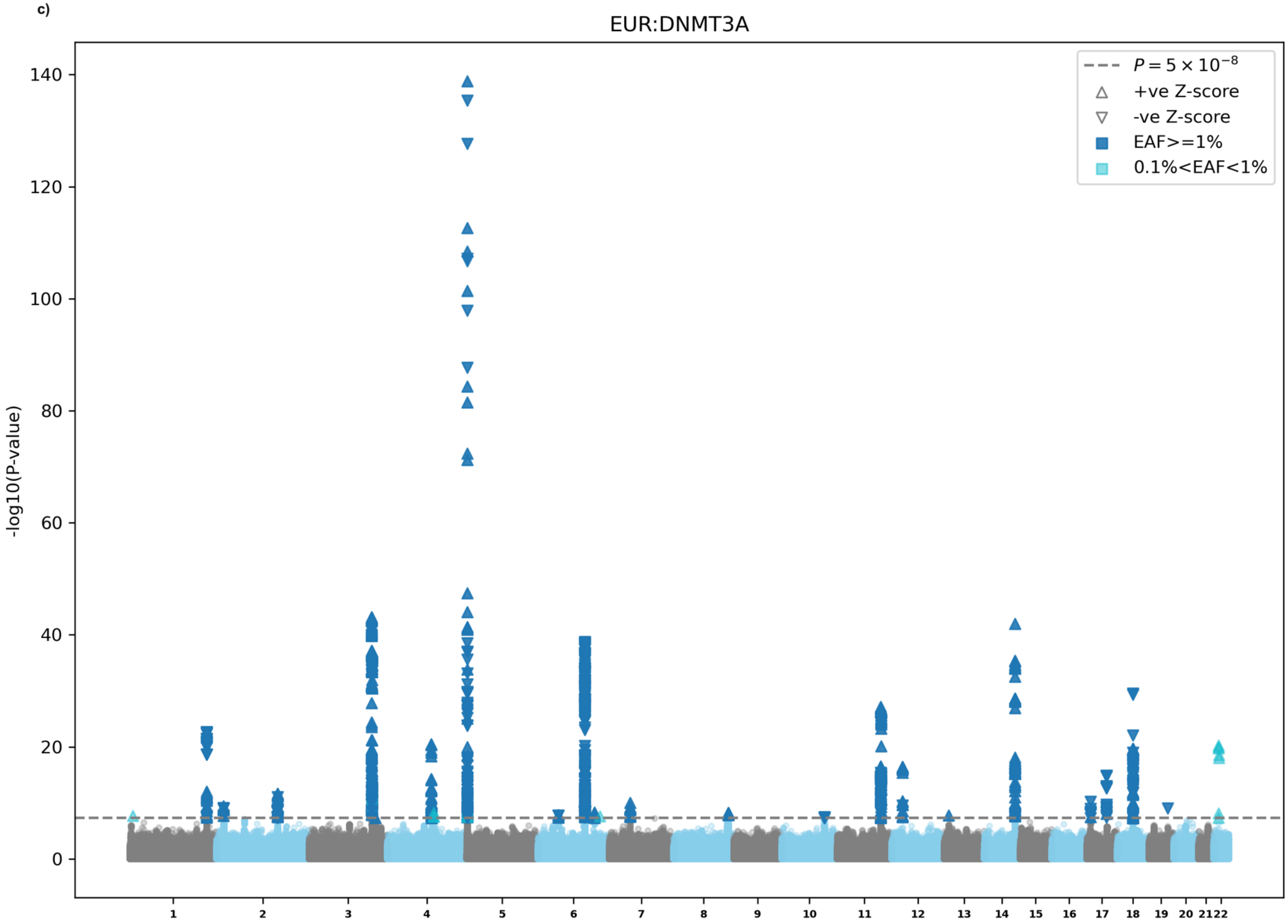

EUR:DNMT3A
$P = 5 \times 10^{-8}$
+ve Z-score
-ve Z-score
EAF>=1%
0.1%<EAF<1%
-log10(P-value)
0
20
40
60
80
100
120
140
1 2 3 4 5 6 7 8 9 10 11 12 13 14 15 16 17 18 19 20 21 22

d)

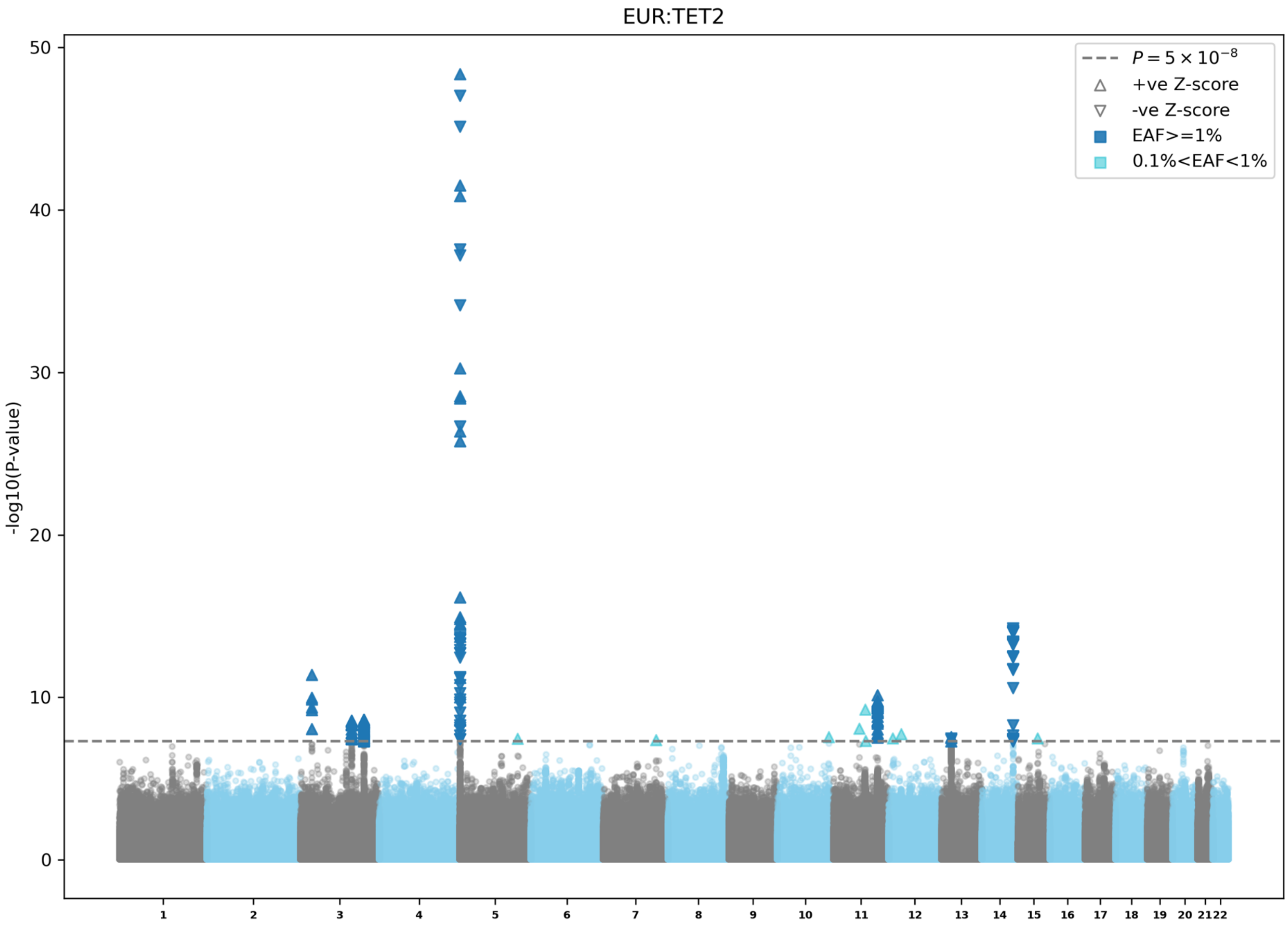

EUR:TET2
-log10(P-value)
50
40
30
20
10
0
$P = 5 \times 10^{-8}$
+ve Z-score
-ve Z-score
EAF>=1%
0.1%<EAF<1%

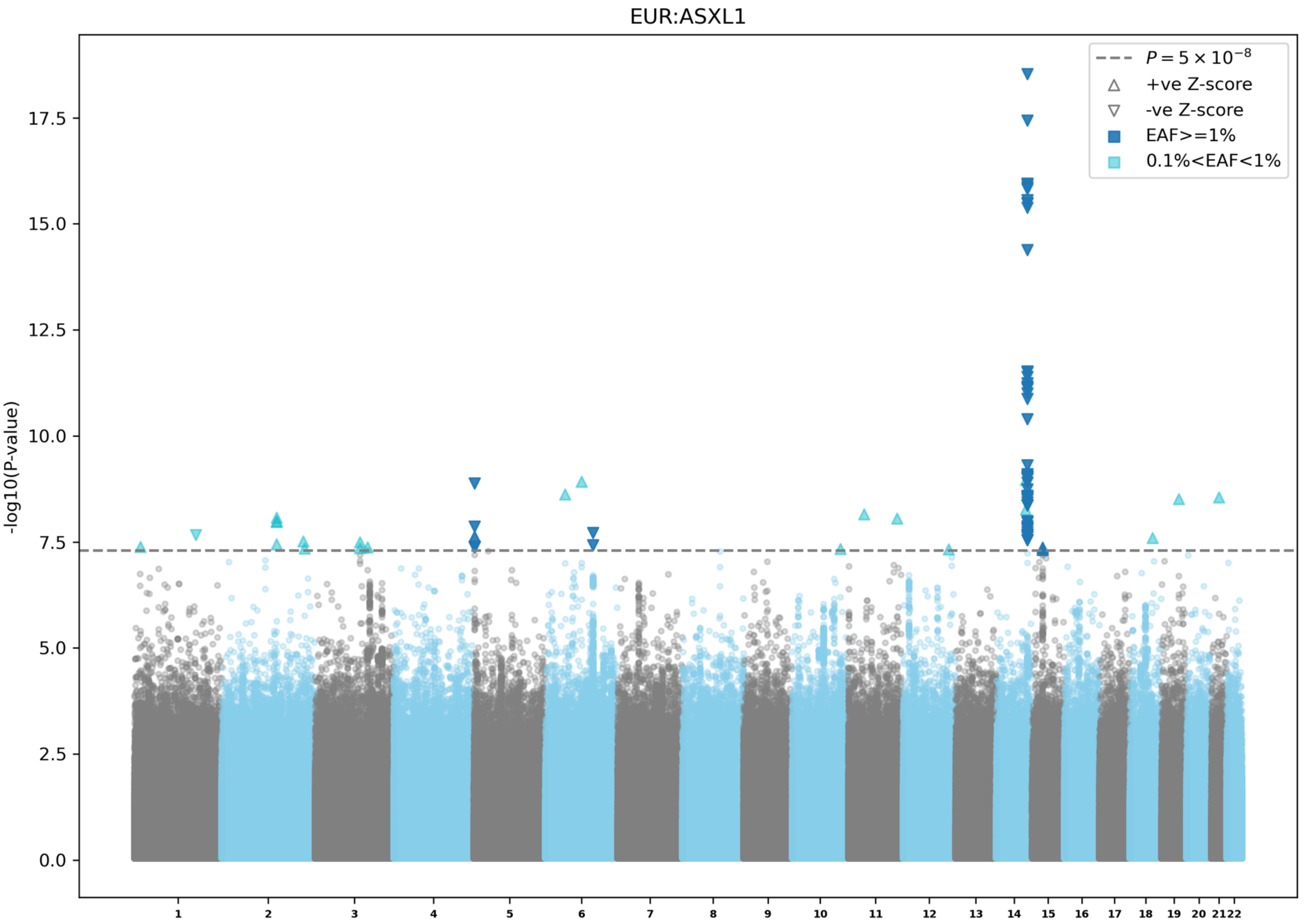

EUR:ASXL1
$P = 5 \times 10^{-8}$
+ve Z-score
-ve Z-score
EAF>=1%
0.1%<EAF<1%
-log10(P-value)
0.0
2.5
5.0
7.5
10.0
12.5
15.0
17.5

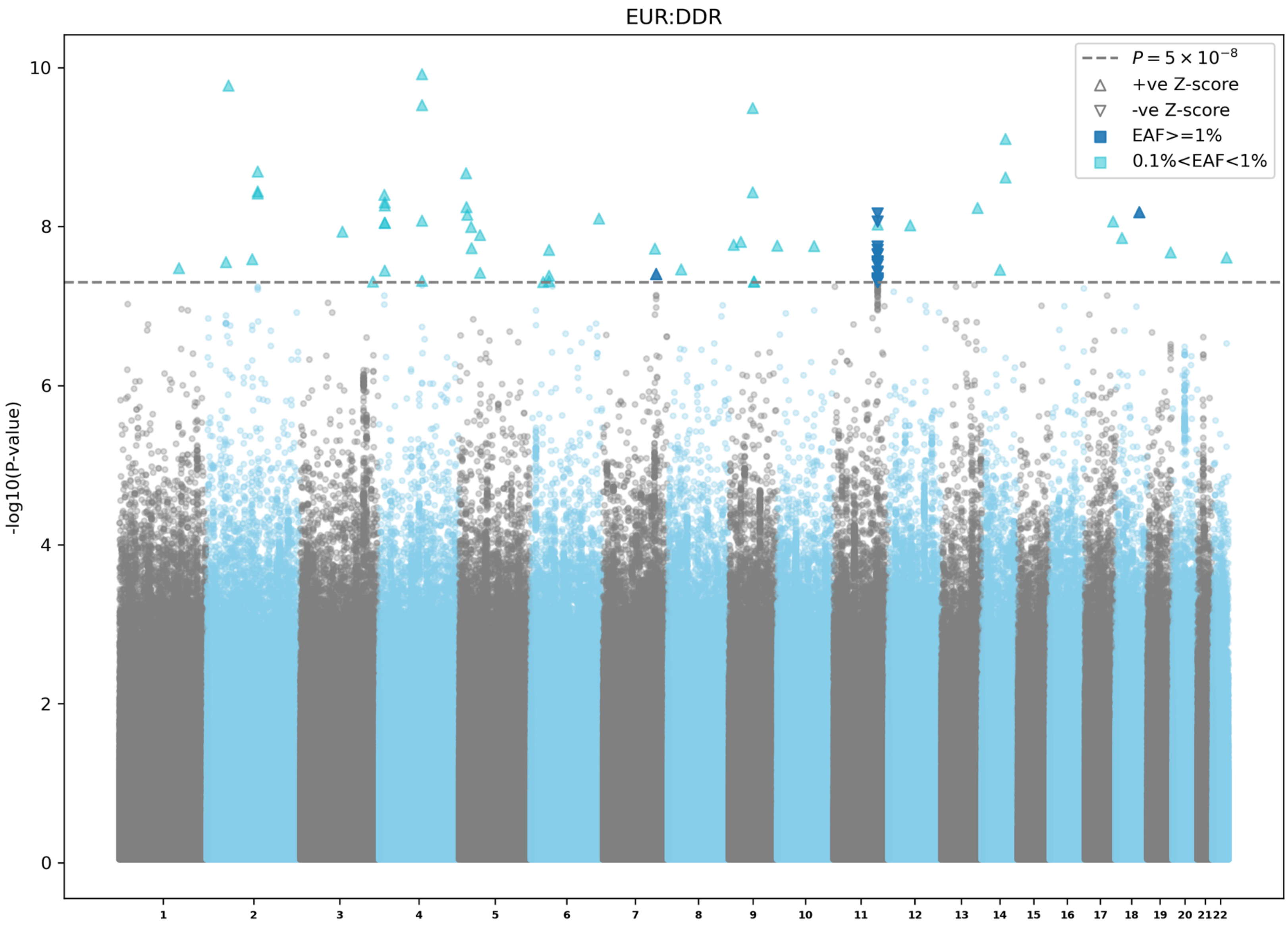

EUR:DDR
-log10(P-value)
0
2
4
6
8
10
$P = 5 \times 10^{-8}$
+ve Z-score
-ve Z-score
EAF>=1%
0.1%<EAF<1%

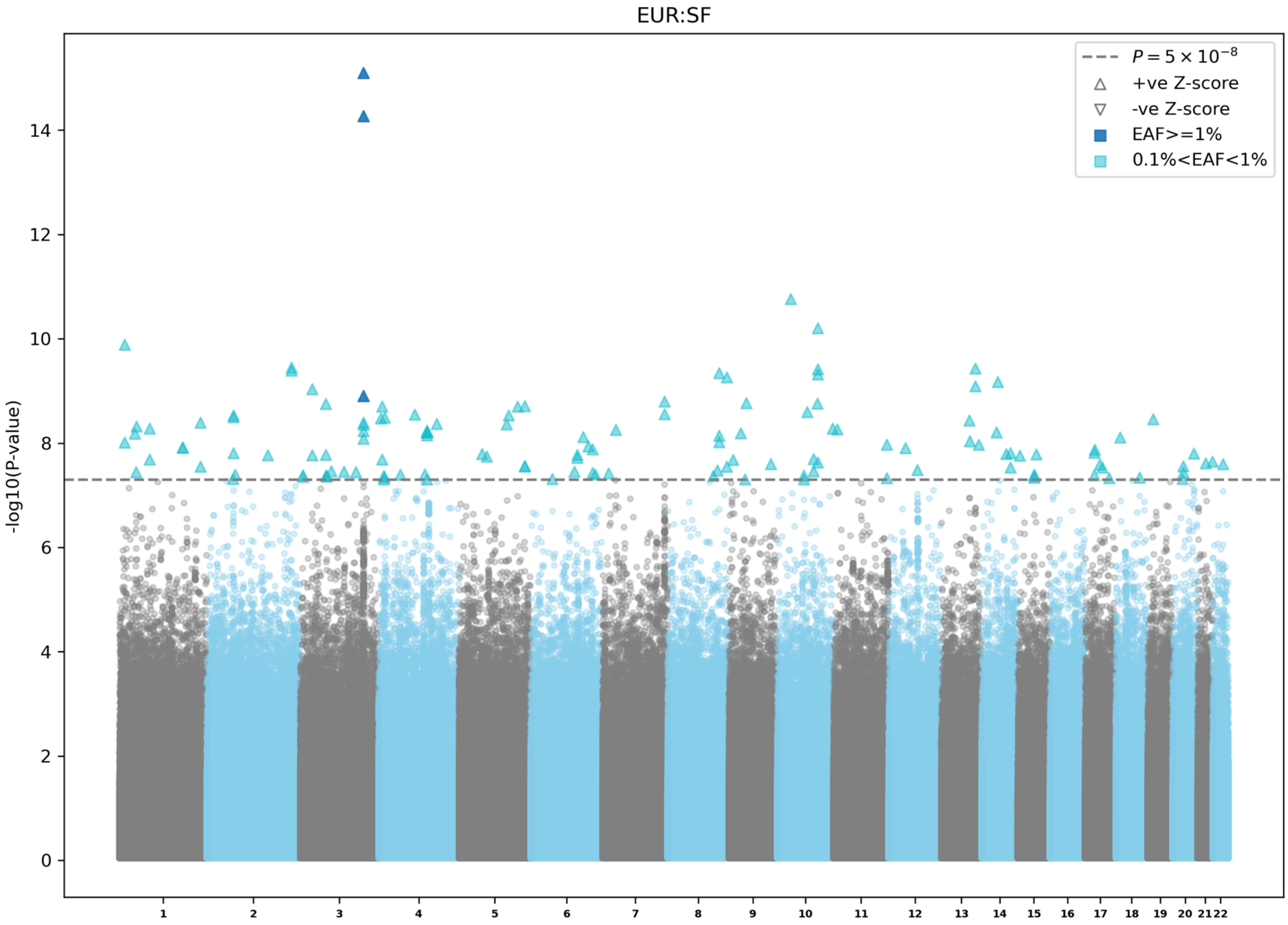
EUR:SF
-log10(P-value)
$P = 5 \times 10^{-8}$
+ve Z-score
-ve Z-score
EAF>=1%
0.1%<EAF<1%
0
2
4
6
8
10
12
14
1 2 3 4 5 6 7 8 9 10 11 12 13 14 15 16 17 18 19 20 21 22

**Extended Data Figure 4 | Manhattan plots for European ancestry specific CHIP GWAS meta-analyses.** a) Overall CHIP GWAS in UKB, AoU, TOPMed, MGBB and BioVU. b) Expanded CHIP (VAF≥10%) GWAS in UKB, AoU, TOPMed, and MGBB. c) *DNMT3A* CHIP GWAS in UKB, AoU, TOPMed, MGBB and BioVU. d) *TET2* CHIP GWAS in UKB, AoU, TOPMed, MGBB and BioVU. e) *ASXL1* CHIP GWAS in UKB, AoU, TOPMed, and, MGBB. f) DDR CHIP GWAS in UKB, AoU, TOPMed, and MGBB. g) SF CHIP GWAS in UKB, AoU, TOPMed, and MGBB. EAF: effect allele frequency; UKB: UK biobank; AoU: All of Us Research; TOPMed: Trans-omics for Precision medicine; MGBB: Mass General and Brigham biobank; BioVU: Vanderbilt BioVU; CHIP: clonal hematopoiesis of indeterminate potential; DDR: DNA damage response genes (*TP53/PPM1D*); SF: splicing factors (*SF3B1/SRSF2/U2AF1/ZRSR2*).

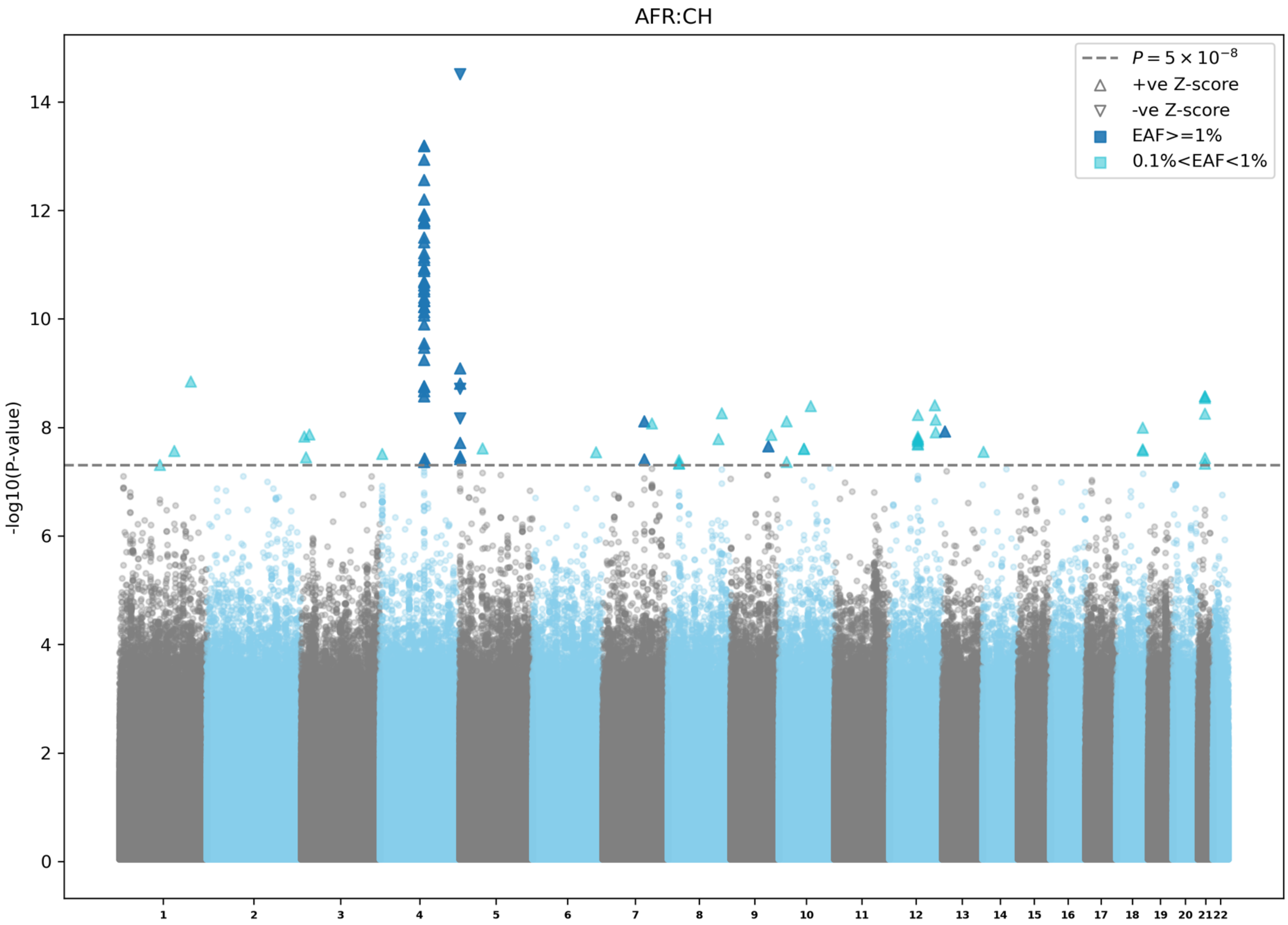

AFR:CH
$P = 5 \times 10^{-8}$
+ve Z-score
-ve Z-score
EAF>=1%
0.1%<EAF<1%
-log10(P-value)
0
2
4
6
8
10
12
14
1 2 3 4 5 6 7 8 9 10 11 12 13 14 15 16 17 18 19 20 21 22

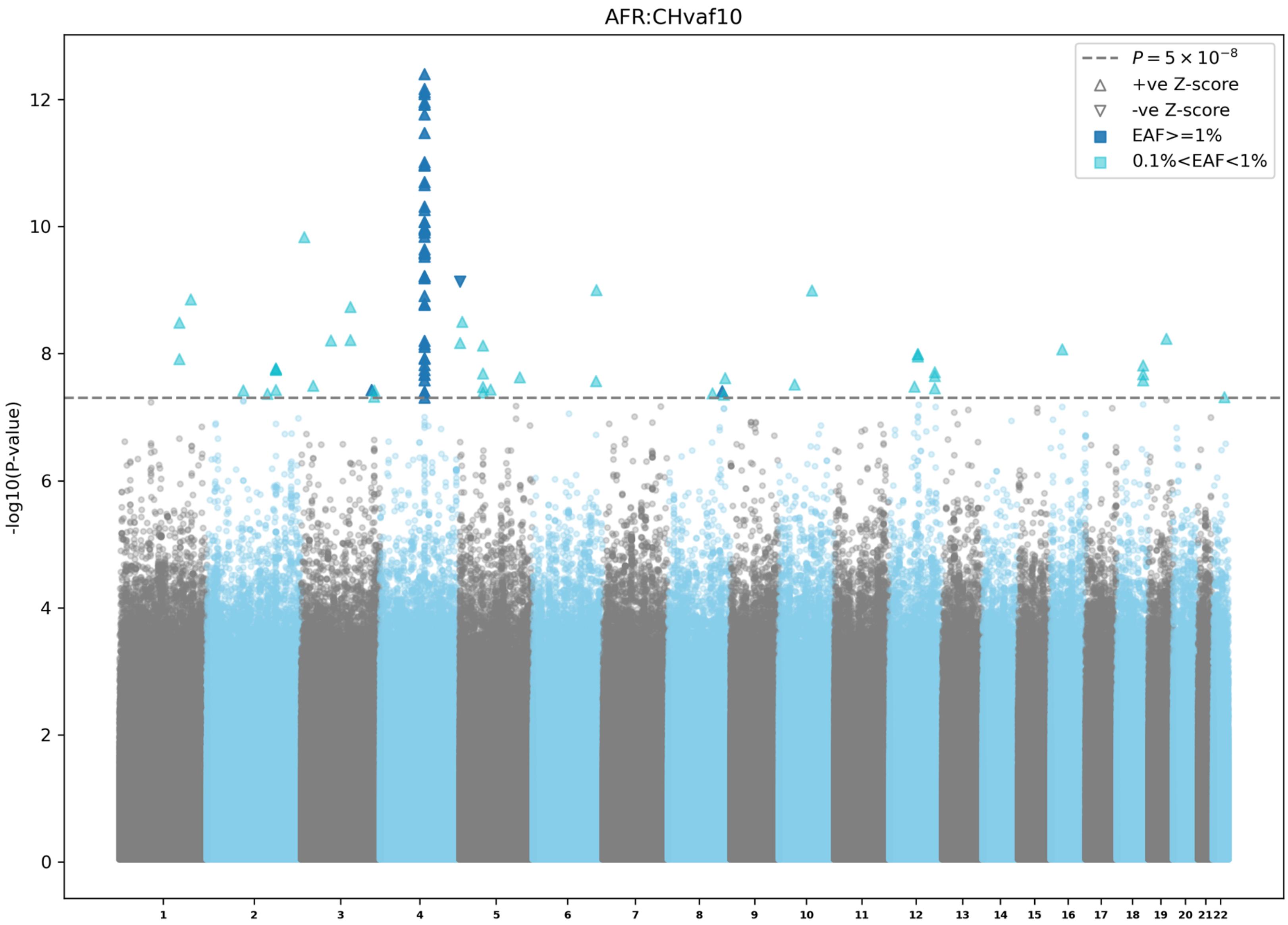

AFR:CHvaf10
$P = 5 \times 10^{-8}$
+ve Z-score
-ve Z-score
EAF>=1%
0.1%<EAF<1%
-log10(P-value)
0
2
4
6
8
10
12

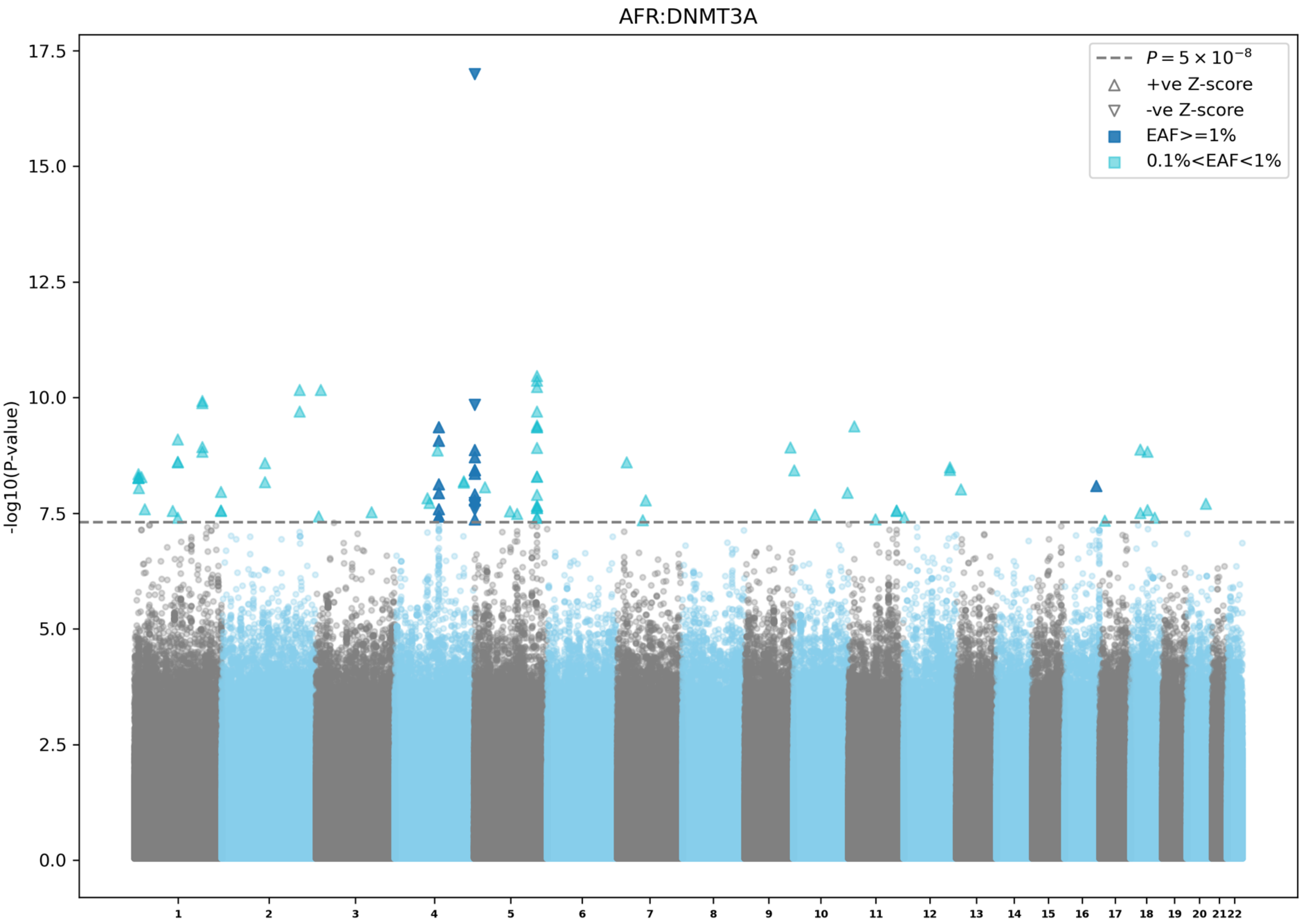

AFR:DNMT3A
-log10(P-value)
17.5
15.0
12.5
10.0
7.5
5.0
2.5
0.0
$P = 5 \times 10^{-8}$
+ve Z-score
-ve Z-score
EAF>=1%
0.1%<EAF<1%

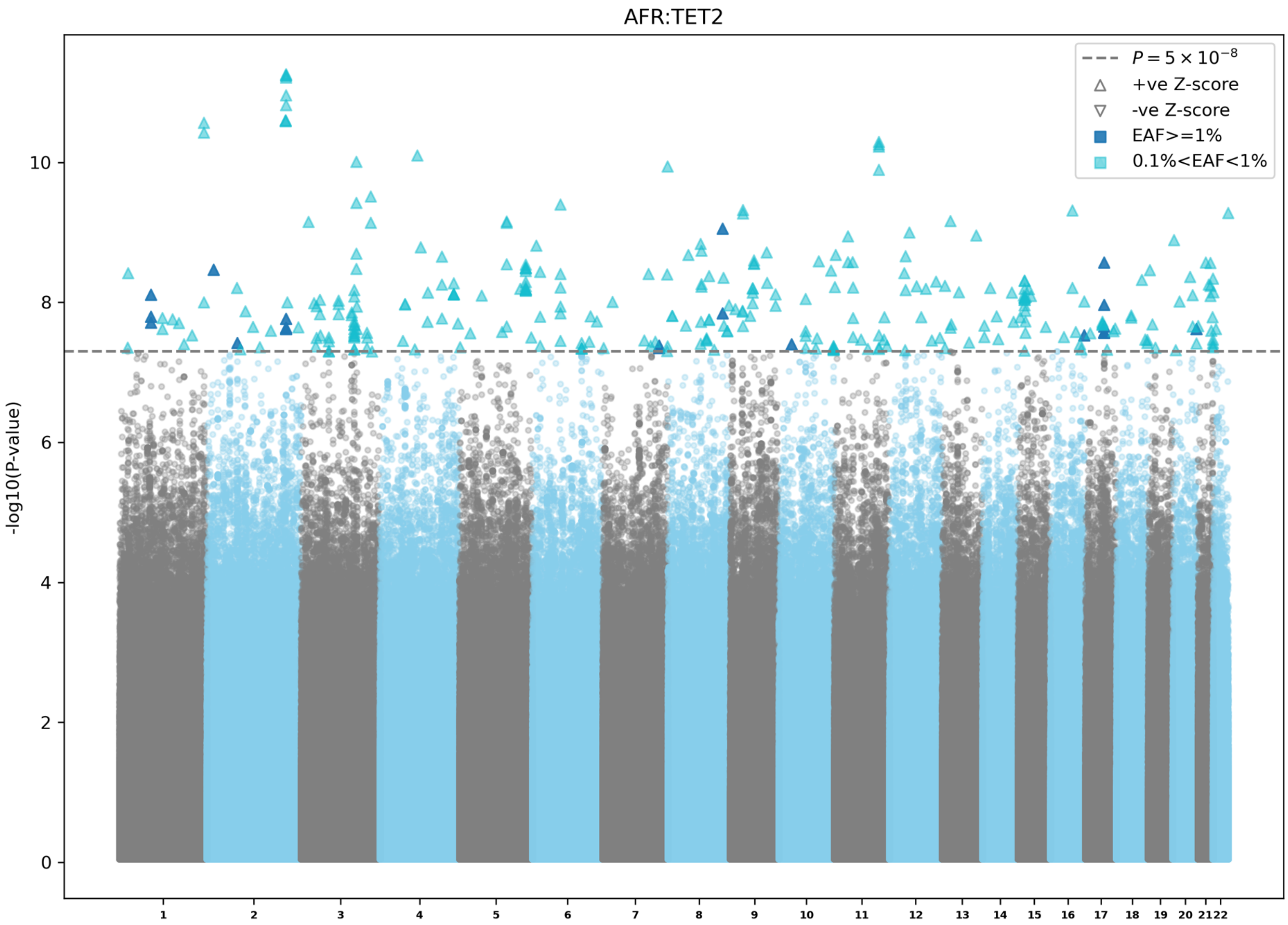

AFR:TET2
$P = 5 \times 10^{-8}$
+ve Z-score
-ve Z-score
EAF>=1%
0.1%<EAF<1%
-log10(P-value)
0
2
4
6
8
10

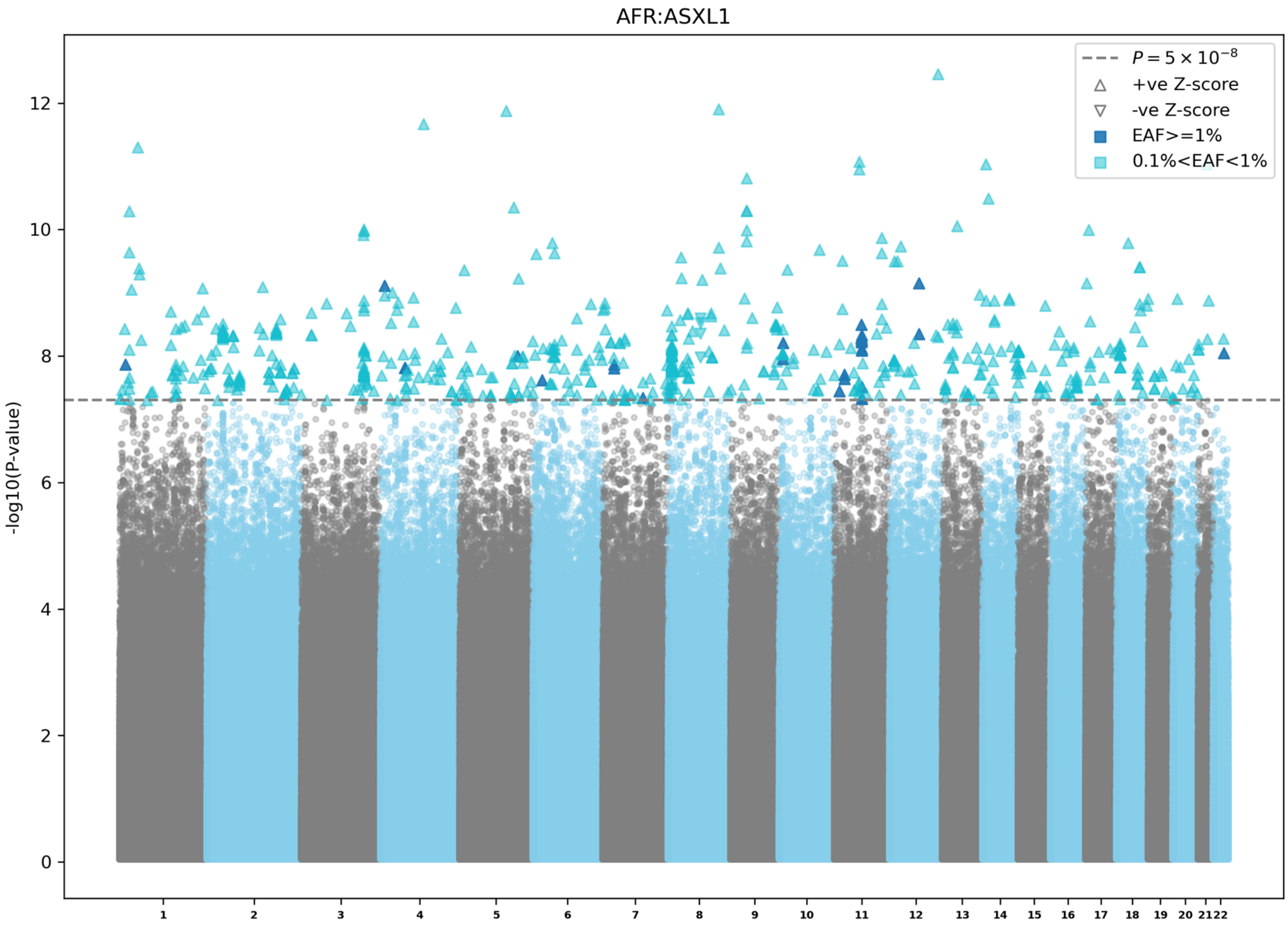

AFR:ASXL1
$P = 5 \times 10^{-8}$
+ve Z-score
-ve Z-score
EAF>=1%
0.1%<EAF<1%
-log10(P-value)
0
2
4
6
8
10
12

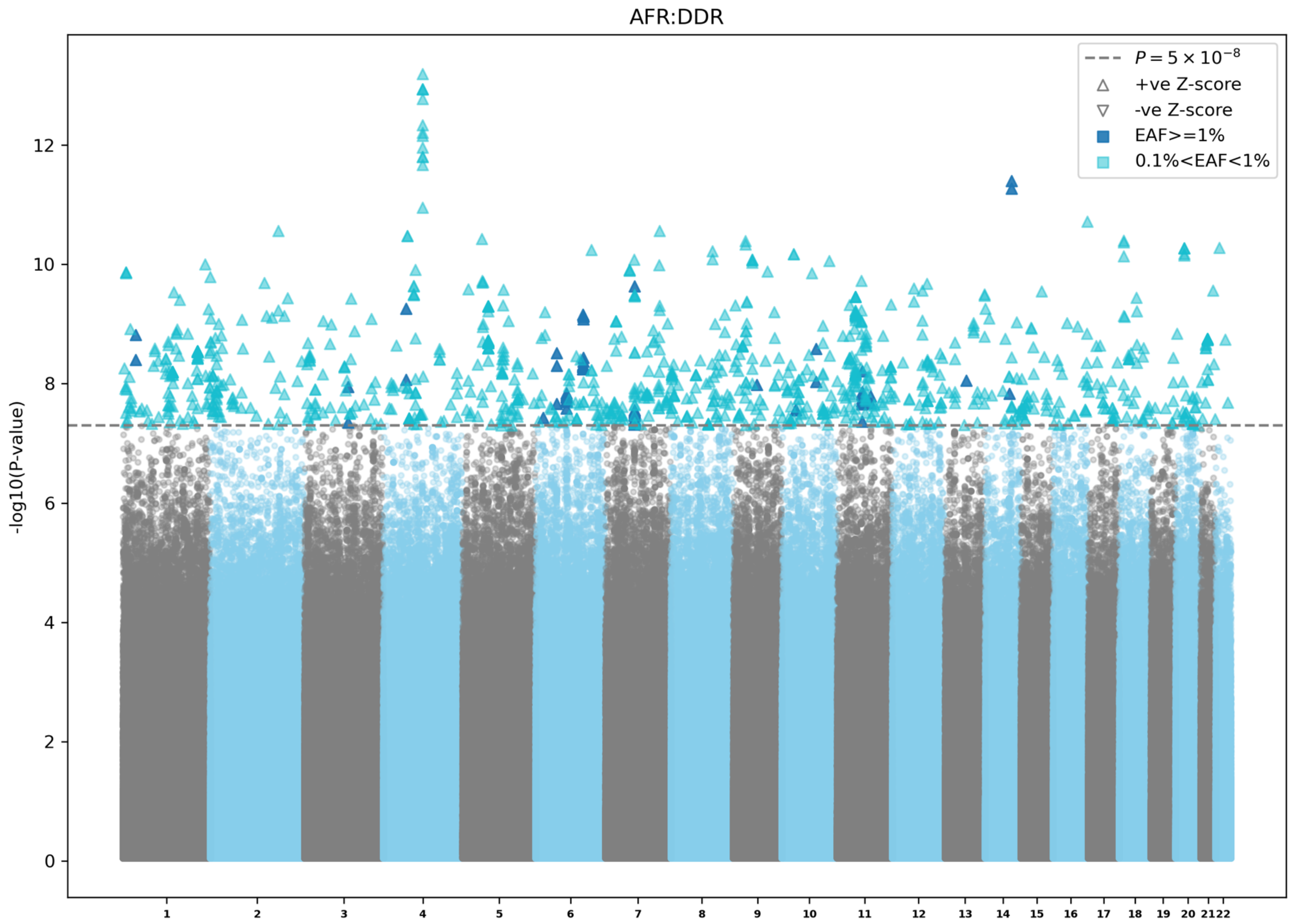

AFR:DDR
$P = 5 \times 10^{-8}$
+ve Z-score
-ve Z-score
EAF>=1%
0.1%<EAF<1%
-log10(P-value)
0
2
4
6
8
10
12

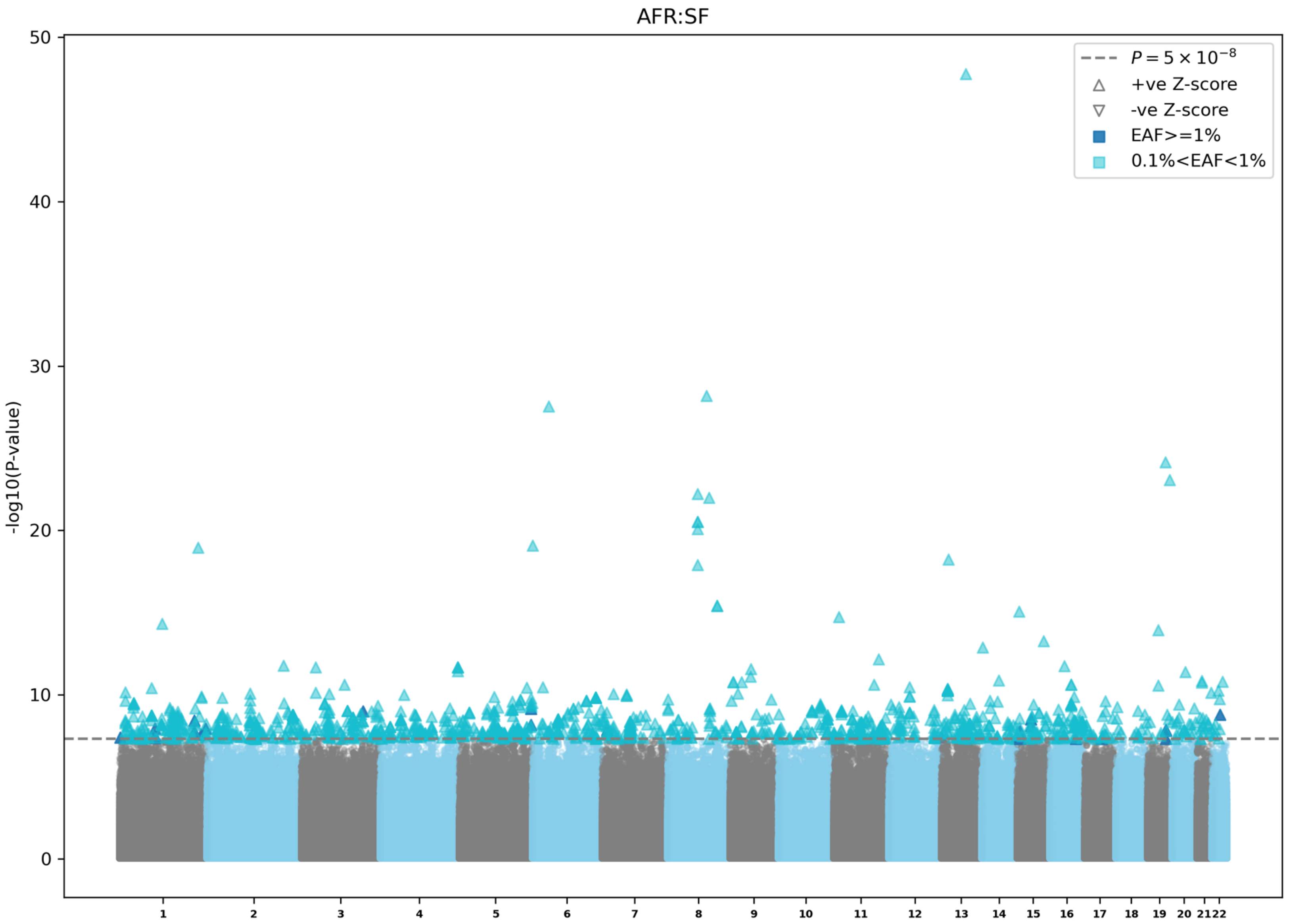

AFR:SF
-log10(P-value)
0
10
20
30
40
50
$P = 5 \times 10^{-8}$
+ve Z-score
-ve Z-score
EAF>=1%
0.1%<EAF<1%

**Extended Data Figure 5 | Manhattan plots for African ancestry specific CHIP GWAS meta-analyses.** a) Overall CHIP GWAS in UKB, AoU, TOPMed, and MGBB. b) Expanded CHIP (VAF≥10%) GWAS in UKB, AoU, TOPMed, and MGBB. c) *DNMT3A* CHIP GWAS in UKB, AoU, TOPMed, and MGBB. d) *TET2* CHIP GWAS in UKB, AoU, TOPMed, and MGBB. e) *ASXL1* CHIP GWAS in UKB, AoU, TOPMed, and MGBB. f) DDR CHIP GWAS in UKB, AoU, TOPMed, and MGBB. g) SF CHIP GWAS in UKB, AoU, TOPMed, and MGBB. EAF: effect allele frequency; UKB: UK biobank; AoU: All of Us Research; TOPMed: Trans-omics for Precision medicine; MGBB: Mass General and Brigham biobank; CHIP: clonal hematopoiesis of indeterminate potential; DDR: DNA damage response genes (*TP53/PPM1D*); SF: splicing factors (*SF3B1/SRSF2/U2AF1/ZRSR2*).

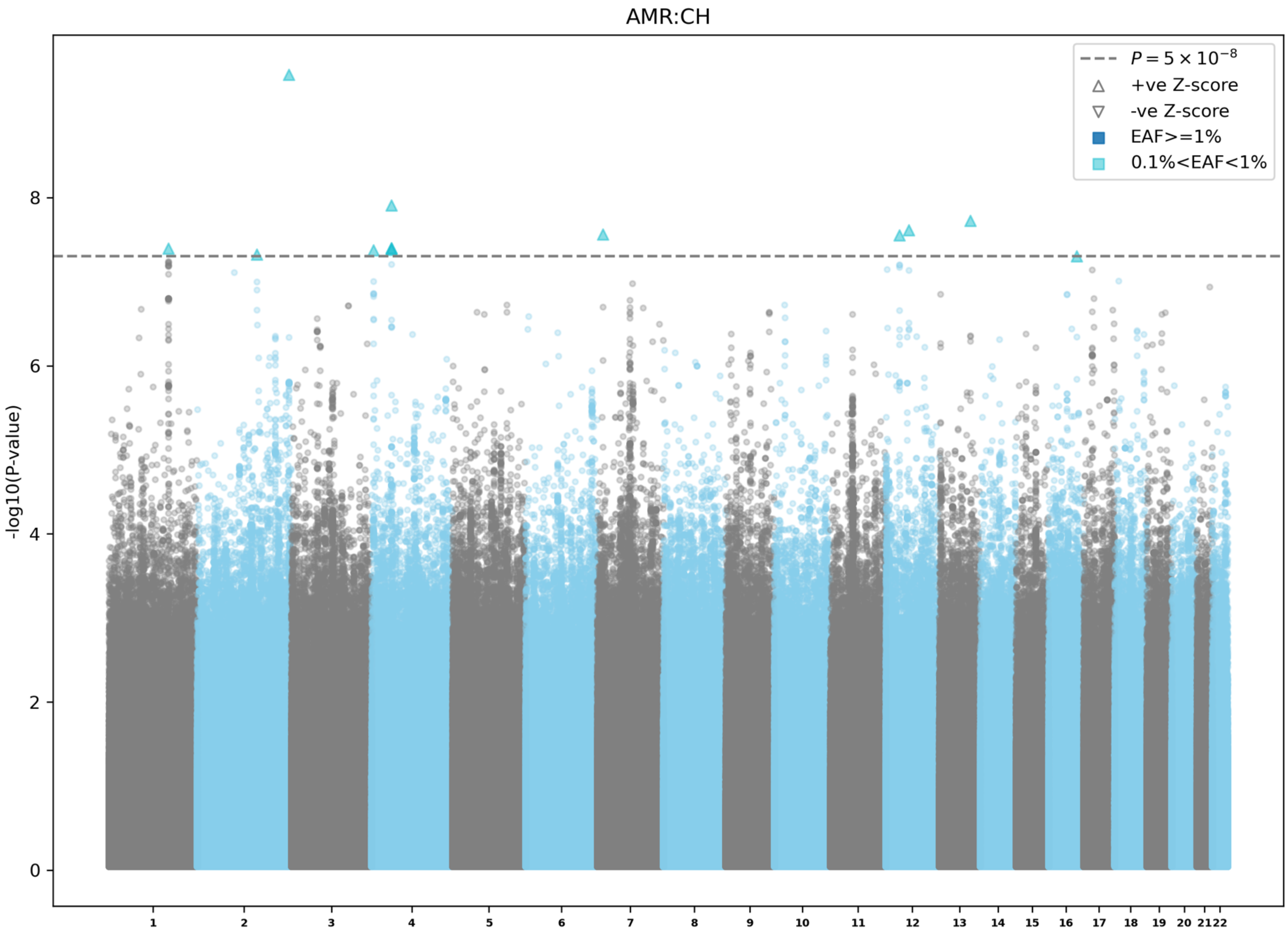
AMR:CH
$P = 5 \times 10^{-8}$
+ve Z-score
-ve Z-score
EAF>=1%
0.1%<EAF<1%
-log10(P-value)
0
2
4
6
8
1 2 3 4 5 6 7 8 9 10 11 12 13 14 15 16 17 18 19 20 21 22

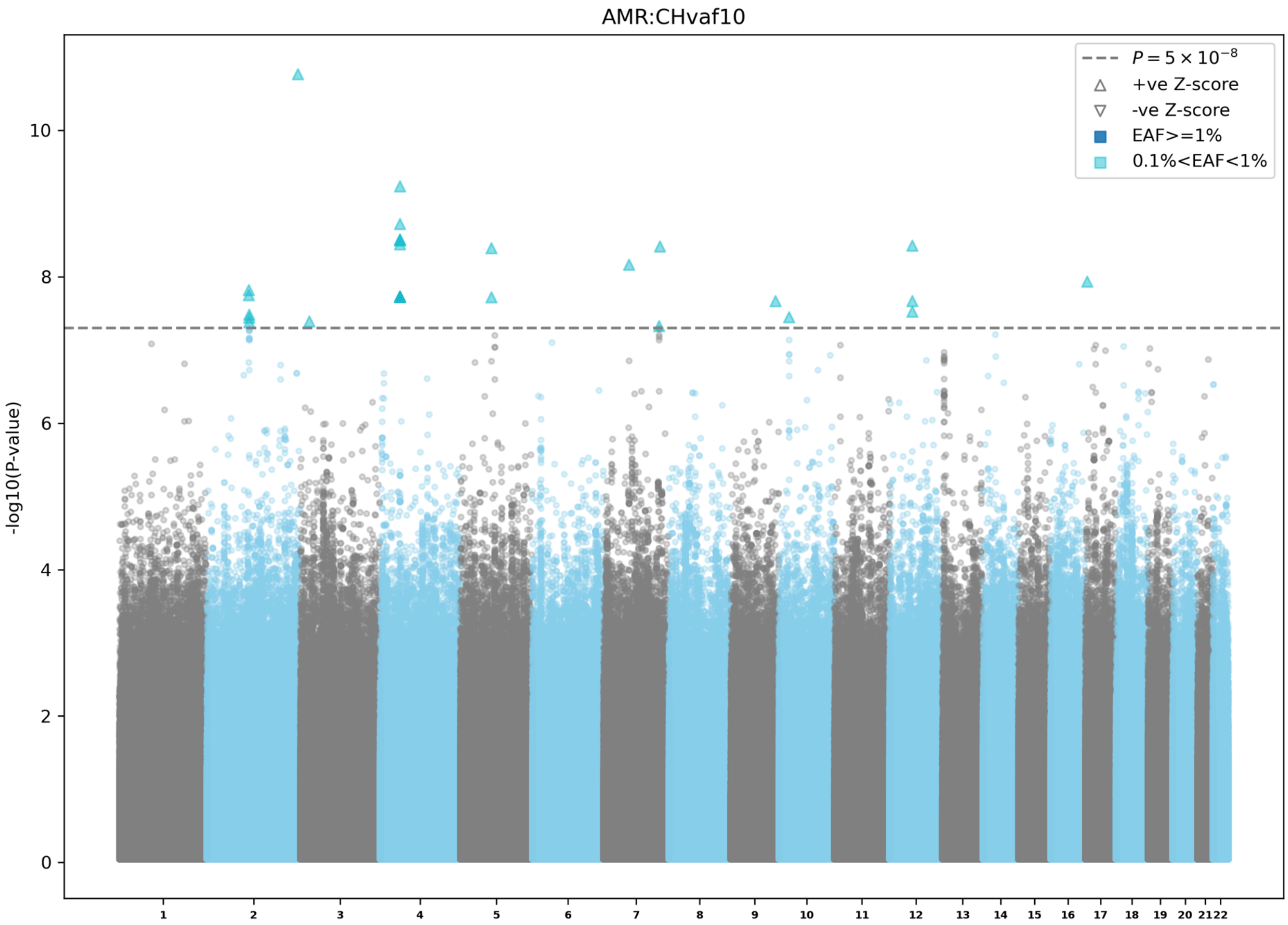
AMR:CHvaf10
$P = 5 \times 10^{-8}$
+ve Z-score
-ve Z-score
EAF>=1%
0.1%<EAF<1%
-log10(P-value)
0
2
4
6
8
10
1 2 3 4 5 6 7 8 9 10 11 12 13 14 15 16 17 18 19 20 21 22

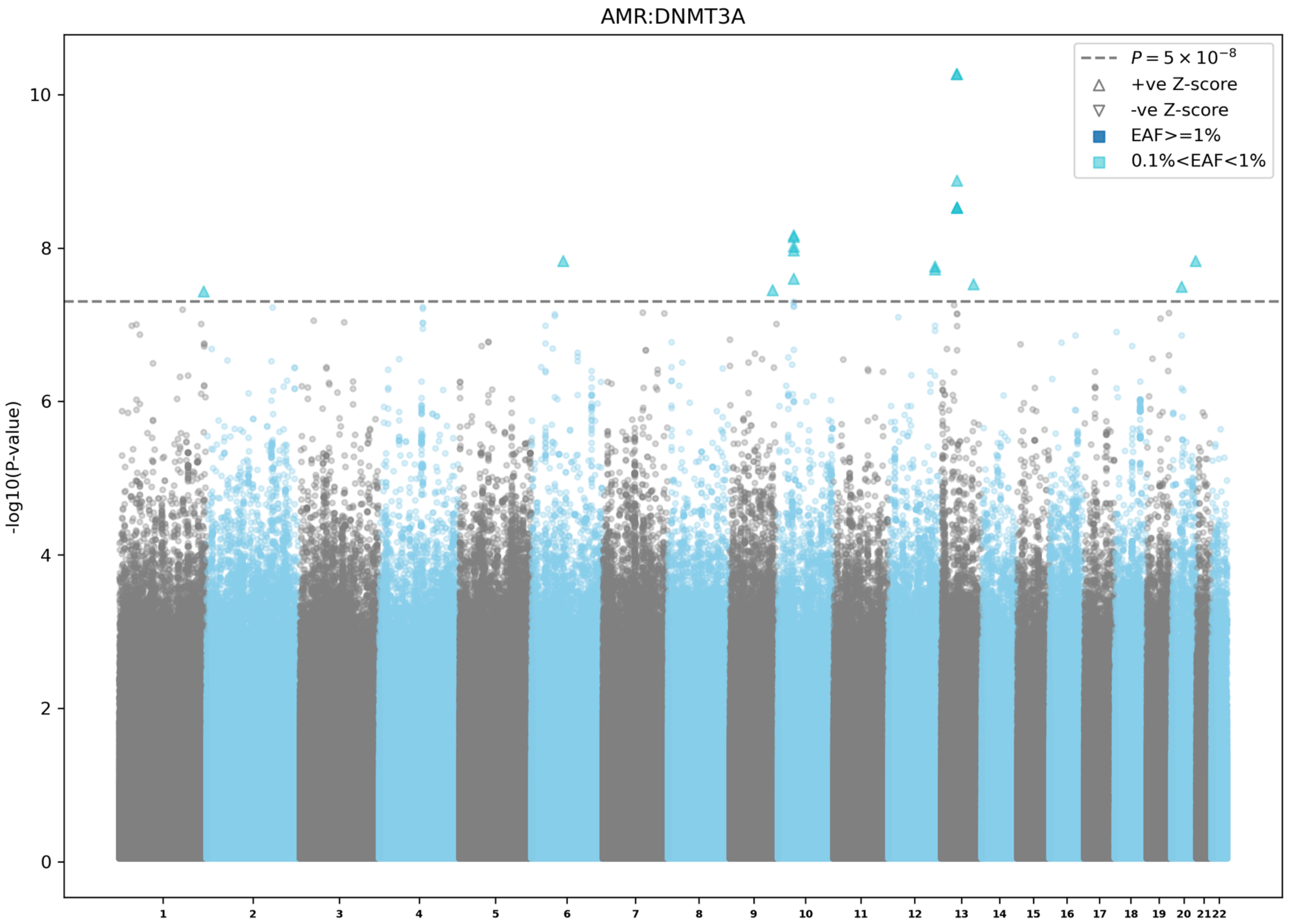

AMR:DNMT3A
$P = 5 \times 10^{-8}$
+ve Z-score
-ve Z-score
EAF>=1%
0.1%<EAF<1%
-log10(P-value)
0
2
4
6
8
10

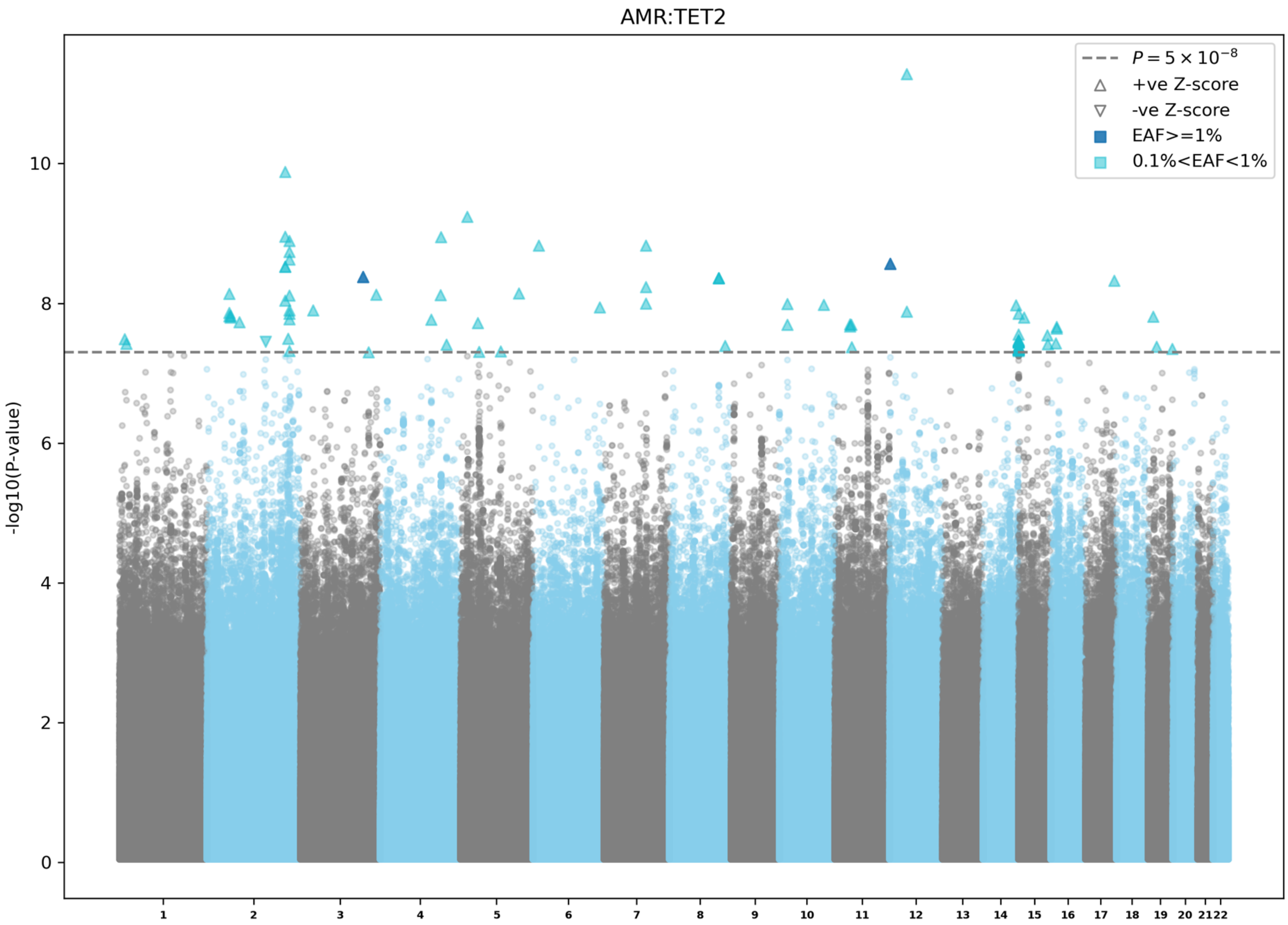
AMR:TET2
$P = 5 \times 10^{-8}$
+ve Z-score
-ve Z-score
EAF>=1%
0.1%<EAF<1%
-log10(P-value)
0
2
4
6
8
10
1 2 3 4 5 6 7 8 9 10 11 12 13 14 15 16 17 18 19 20 21 22

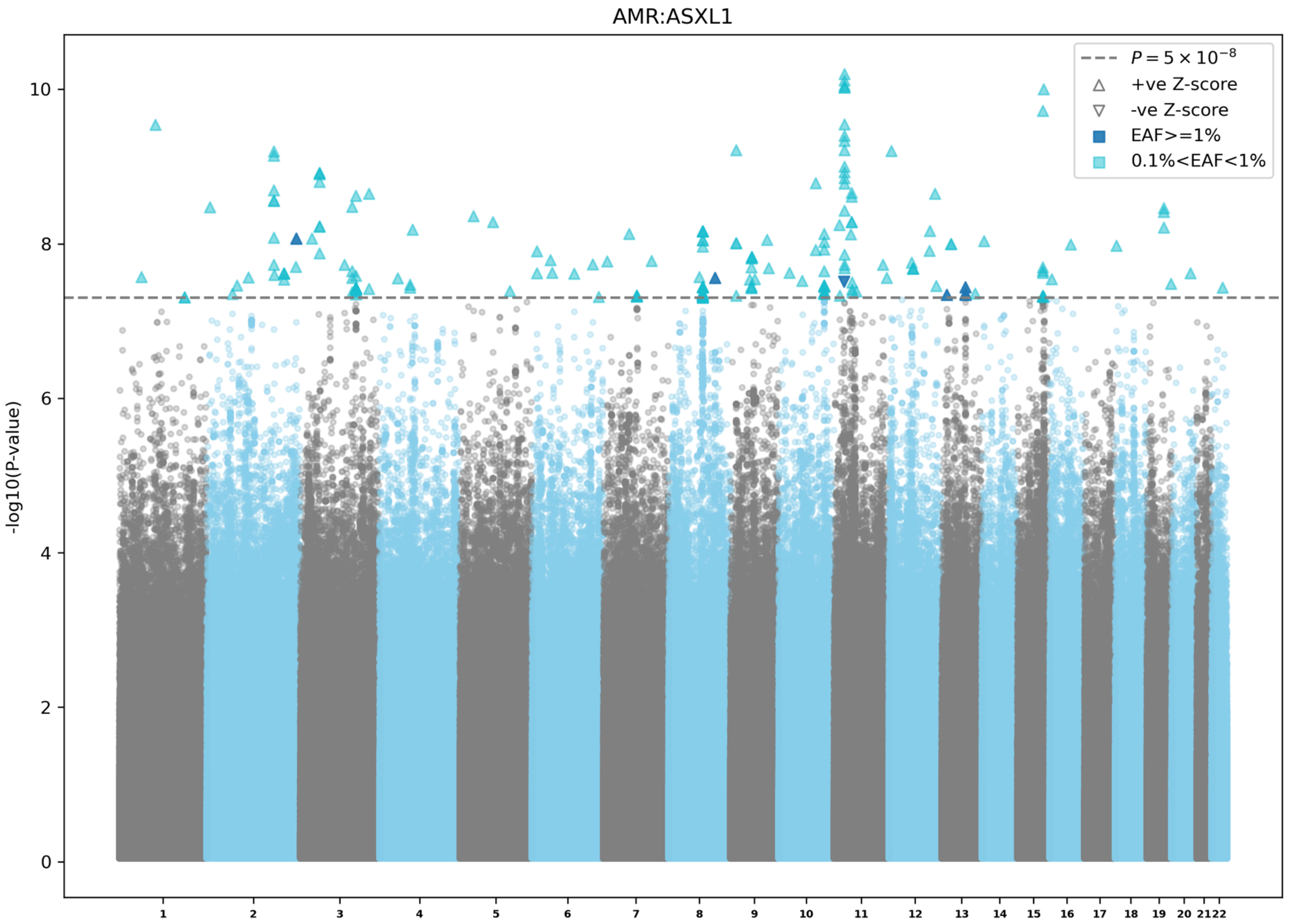

AMR:ASXL1
$P = 5 \times 10^{-8}$
+ve Z-score
-ve Z-score
EAF>=1%
0.1%<EAF<1%
-log10(P-value)
0
2
4
6
8
10

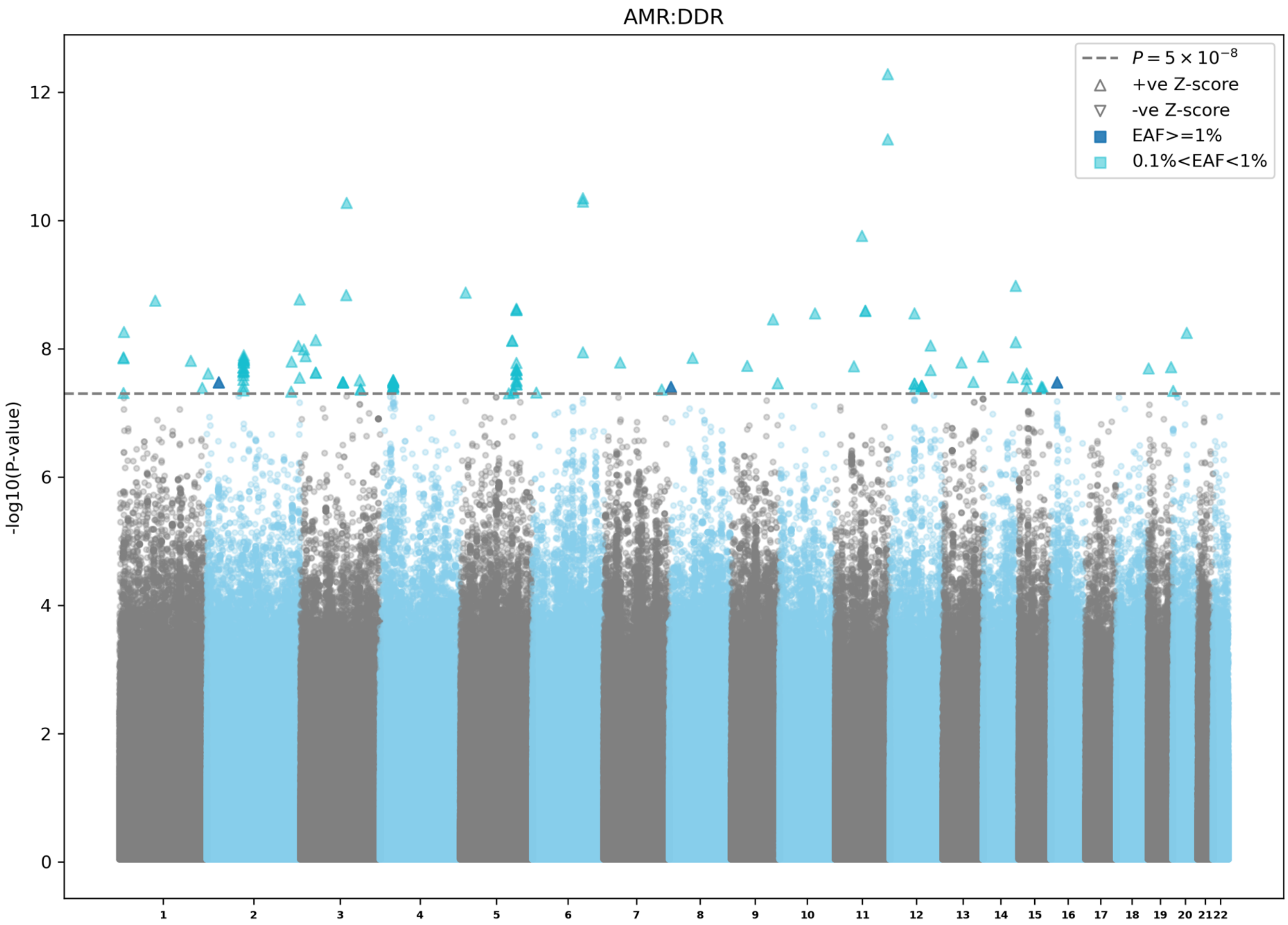

AMR:DDR
$P = 5 \times 10^{-8}$
+ve Z-score
-ve Z-score
EAF>=1%
0.1%<EAF<1%
-log10(P-value)
0
2
4
6
8
10
12
1 2 3 4 5 6 7 8 9 10 11 12 13 14 15 16 17 18 19 20 21 22

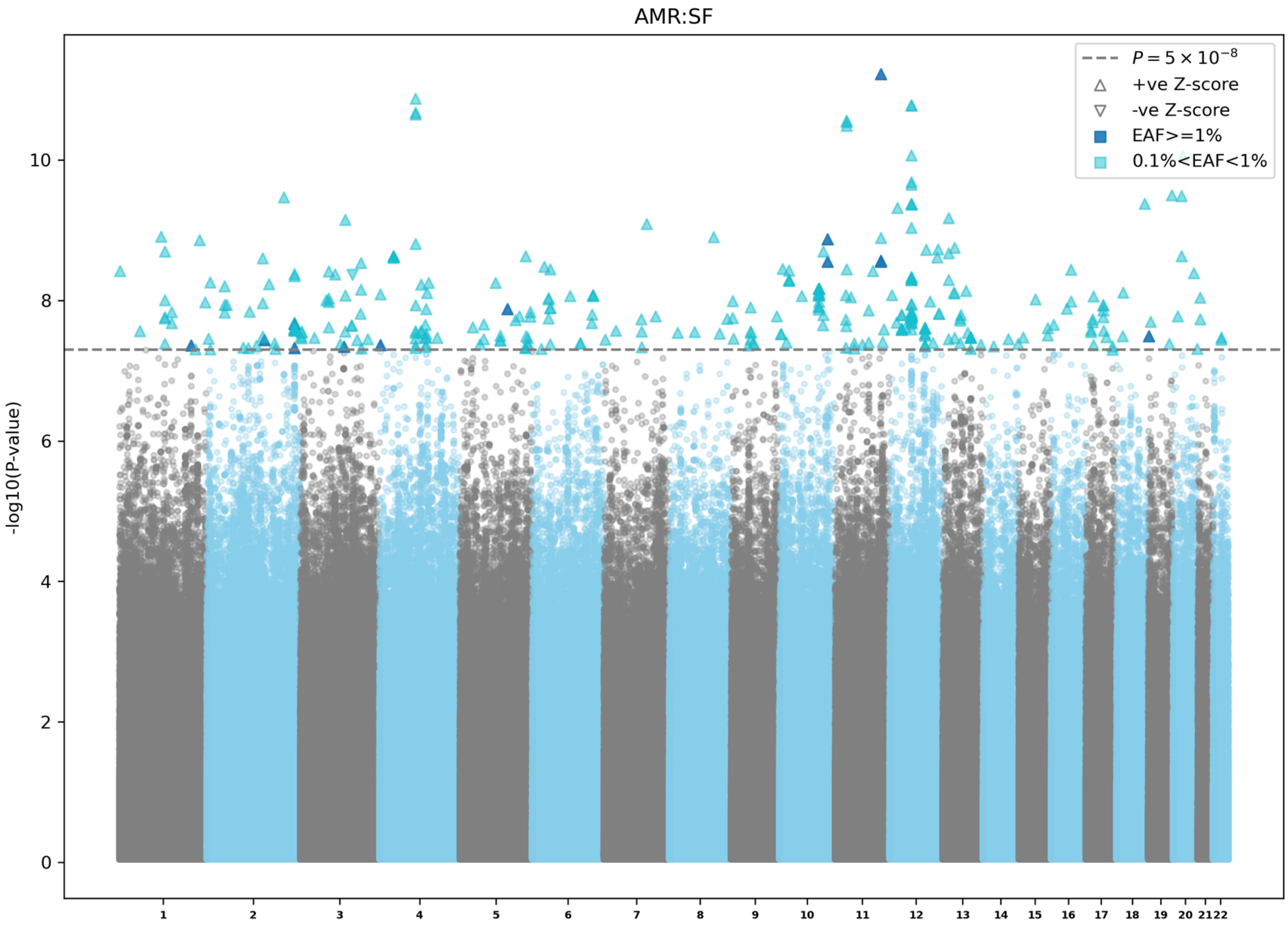
AMR:SF
$P = 5 \times 10^{-8}$
+ve Z-score
-ve Z-score
EAF>=1%
0.1%<EAF<1%
-log10(P-value)
0
2
4
6
8
10

**Extended Data Figure 6 | Manhattan plots for Admixed American ancestry specific CHIP GWAS meta-analyses.** a) Overall CHIP GWAS in UKB, AoU, TOPMed, and MGBB. b) Expanded CHIP (VAF≥10%) GWAS in UKB, AoU, TOPMed, and MGBB. c) *DNMT3A* CHIP GWAS in UKB, AoU, TOPMed, and MGBB. d) *TET2* CHIP GWAS in UKB, AoU, TOPMed, and MGBB. e) *ASXL1* CHIP GWAS in UKB, AoU, TOPMed, and MGBB. f) DDR CHIP GWAS in UKB, AoU, TOPMed, and MGBB. g) SF CHIP GWAS in UKB, AoU, TOPMed, and MGBB. EAF: effect allele frequency; UKB: UK biobank; AoU: All of Us Research; TOPMed: Trans-omics for Precision medicine; MGBB: Mass General and Brigham biobank; CHIP: clonal hematopoiesis of indeterminate potential; DDR: DNA damage response genes (*TP53/PPM1D*); SF: splicing factors (*SF3B1/SRSF2/U2AF1/ZRSR2*).

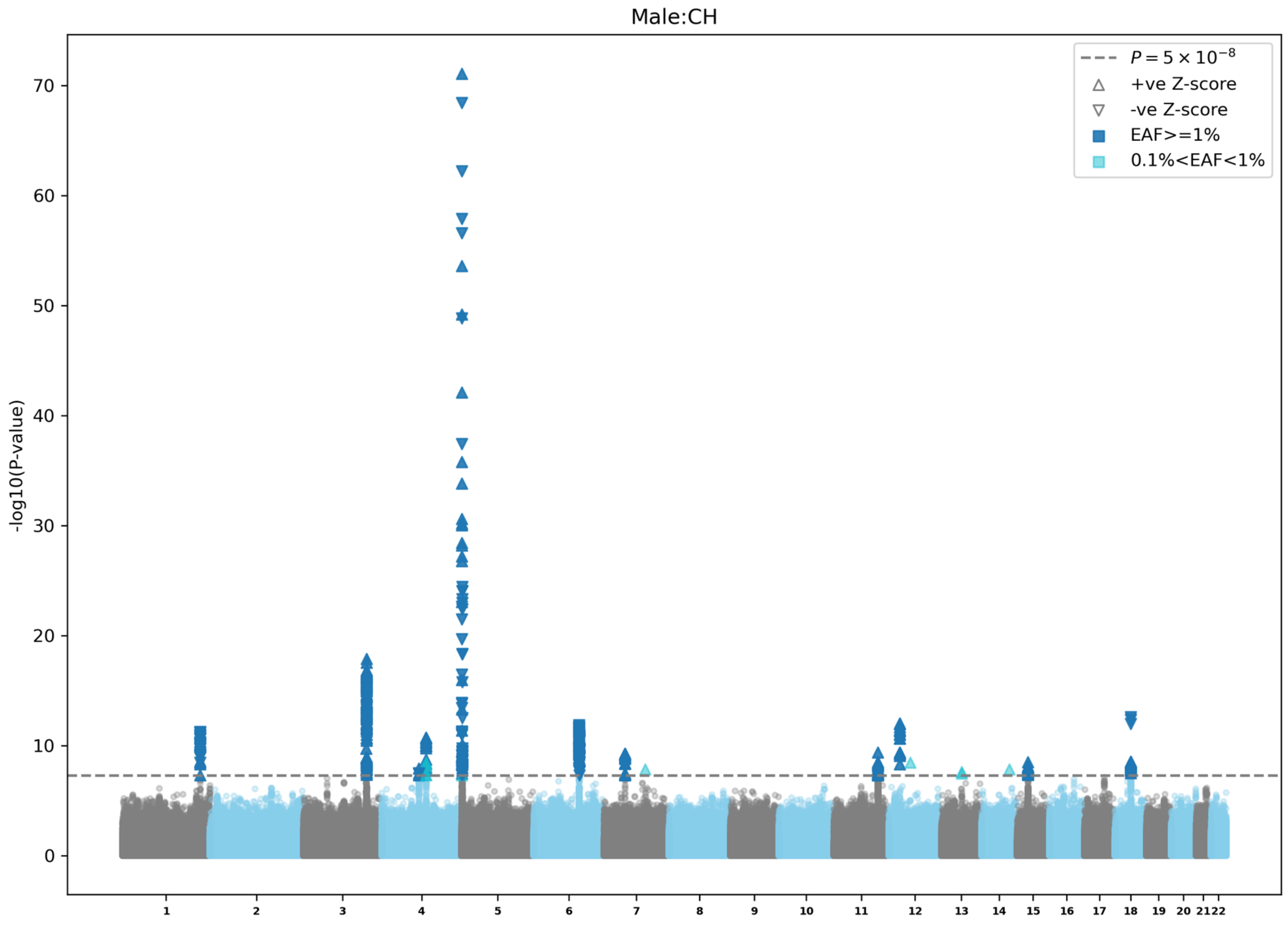

Male:CH
-log10(P-value)
0
10
20
30
40
50
60
70
$P = 5 \times 10^{-8}$
+ve Z-score
-ve Z-score
EAF>=1%
0.1%<EAF<1%

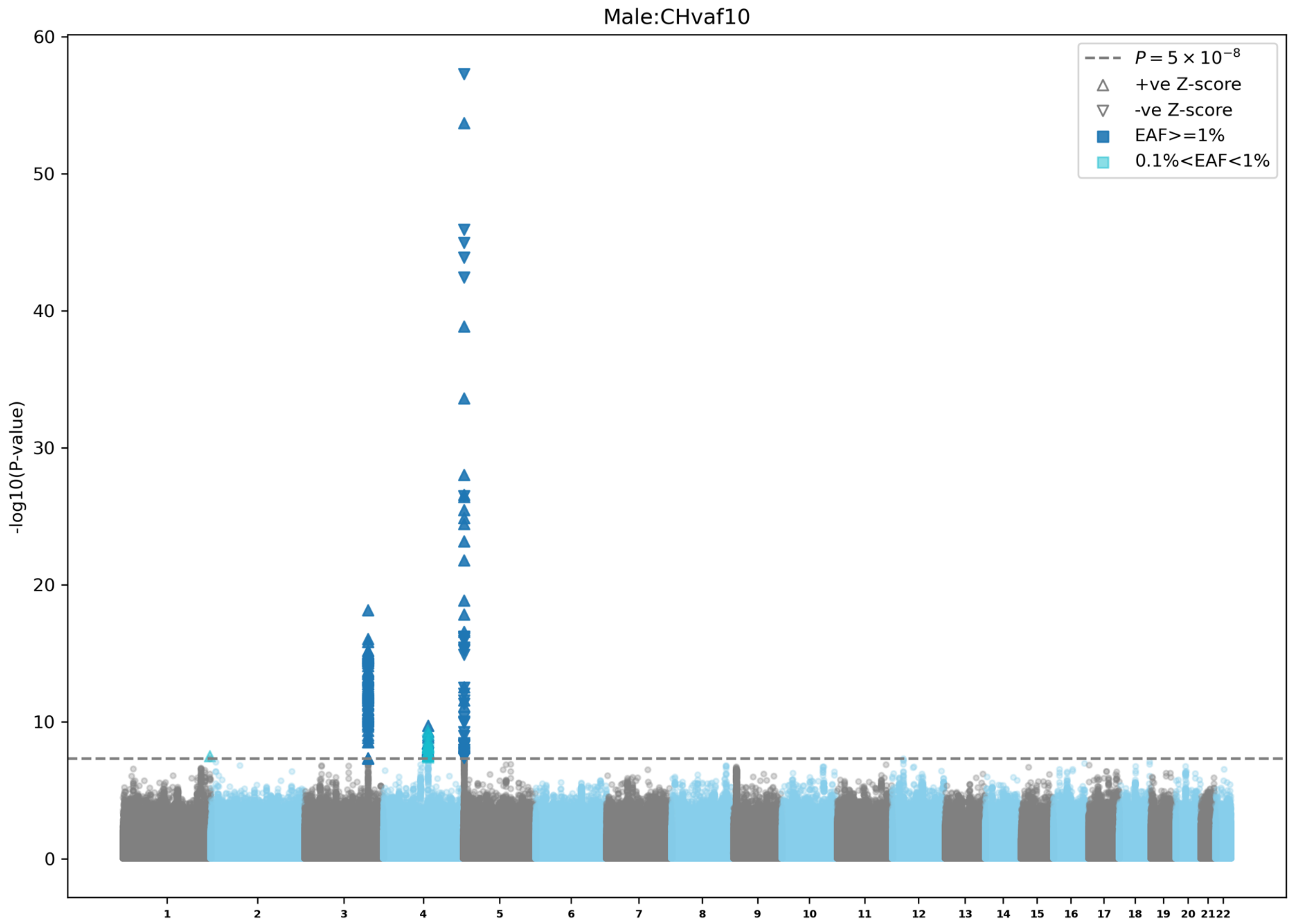

Male:CHvaf10
$P = 5 \times 10^{-8}$
+ve Z-score
-ve Z-score
EAF>=1%
0.1%<EAF<1%
-log10(P-value)
0
10
20
30
40
50
60

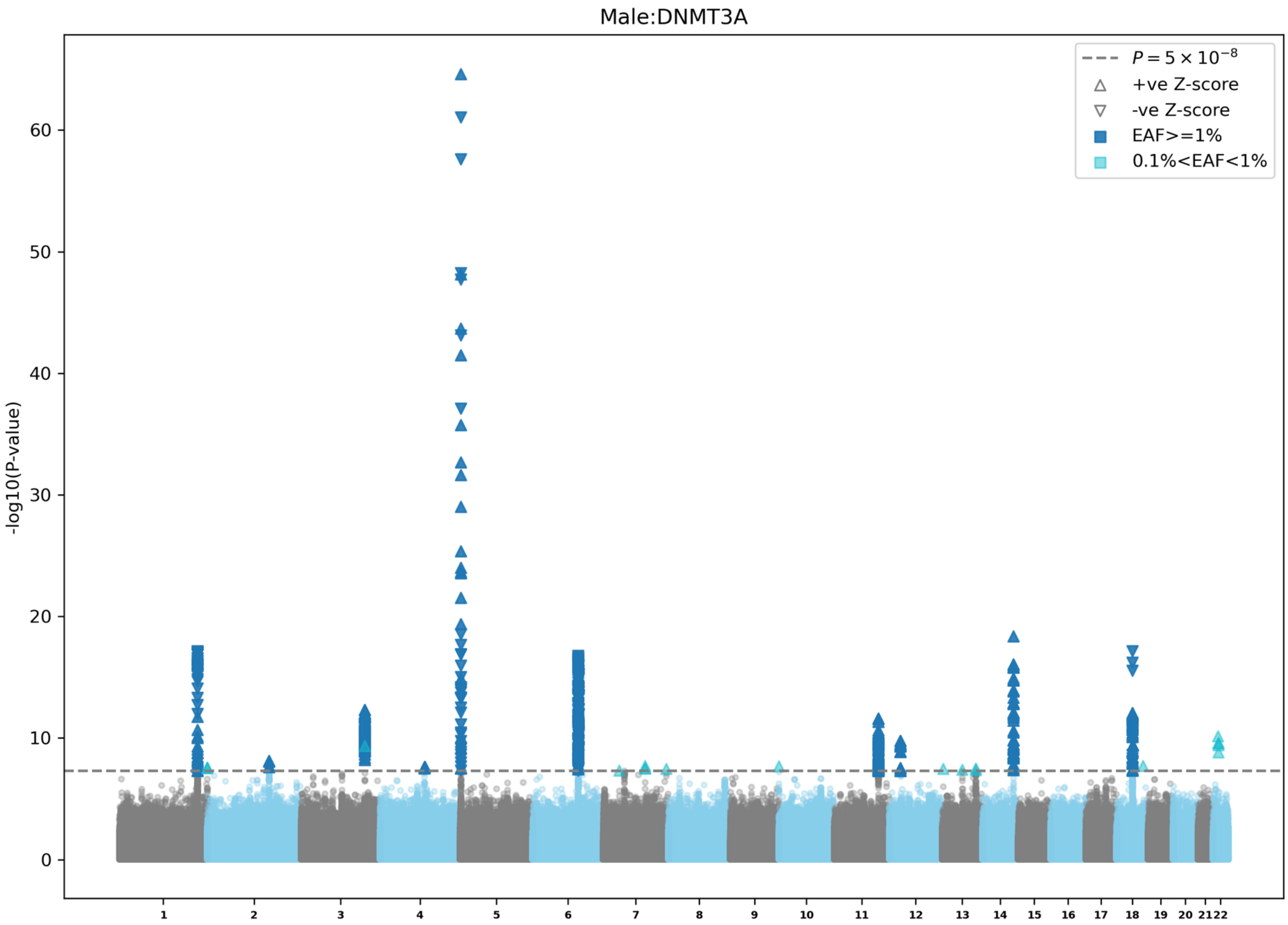
Male:DNMT3A
$P = 5 \times 10^{-8}$
+ve Z-score
-ve Z-score
EAF>=1%
0.1%<EAF<1%
-log10(P-value)
0
10
20
30
40
50
60

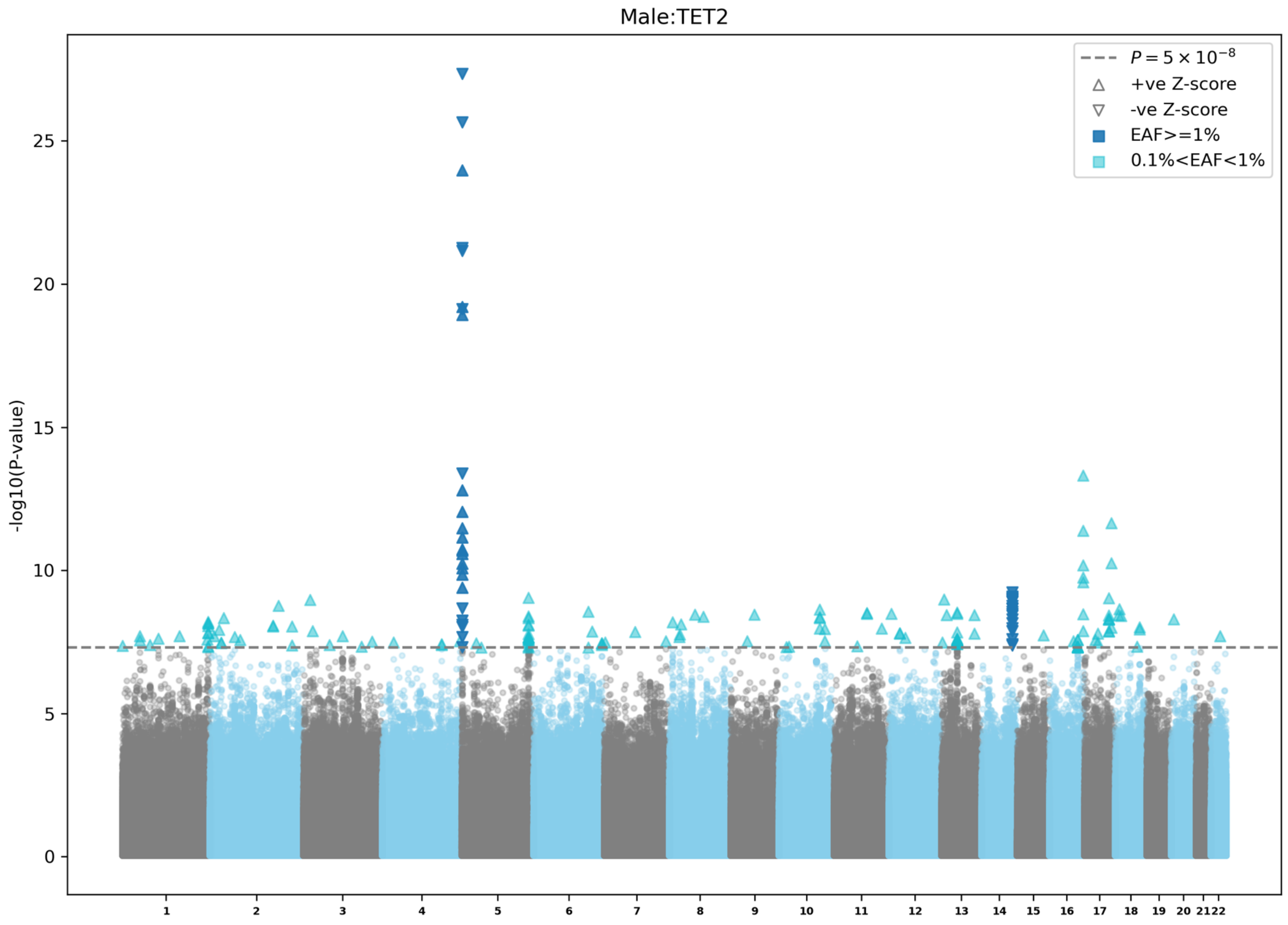
Male:TET2
-log10(P-value)
0
5
10
15
20
25
$P = 5 \times 10^{-8}$
+ve Z-score
-ve Z-score
EAF>=1%
0.1%<EAF<1%
1 2 3 4 5 6 7 8 9 10 11 12 13 14 15 16 17 18 19 20 21 22

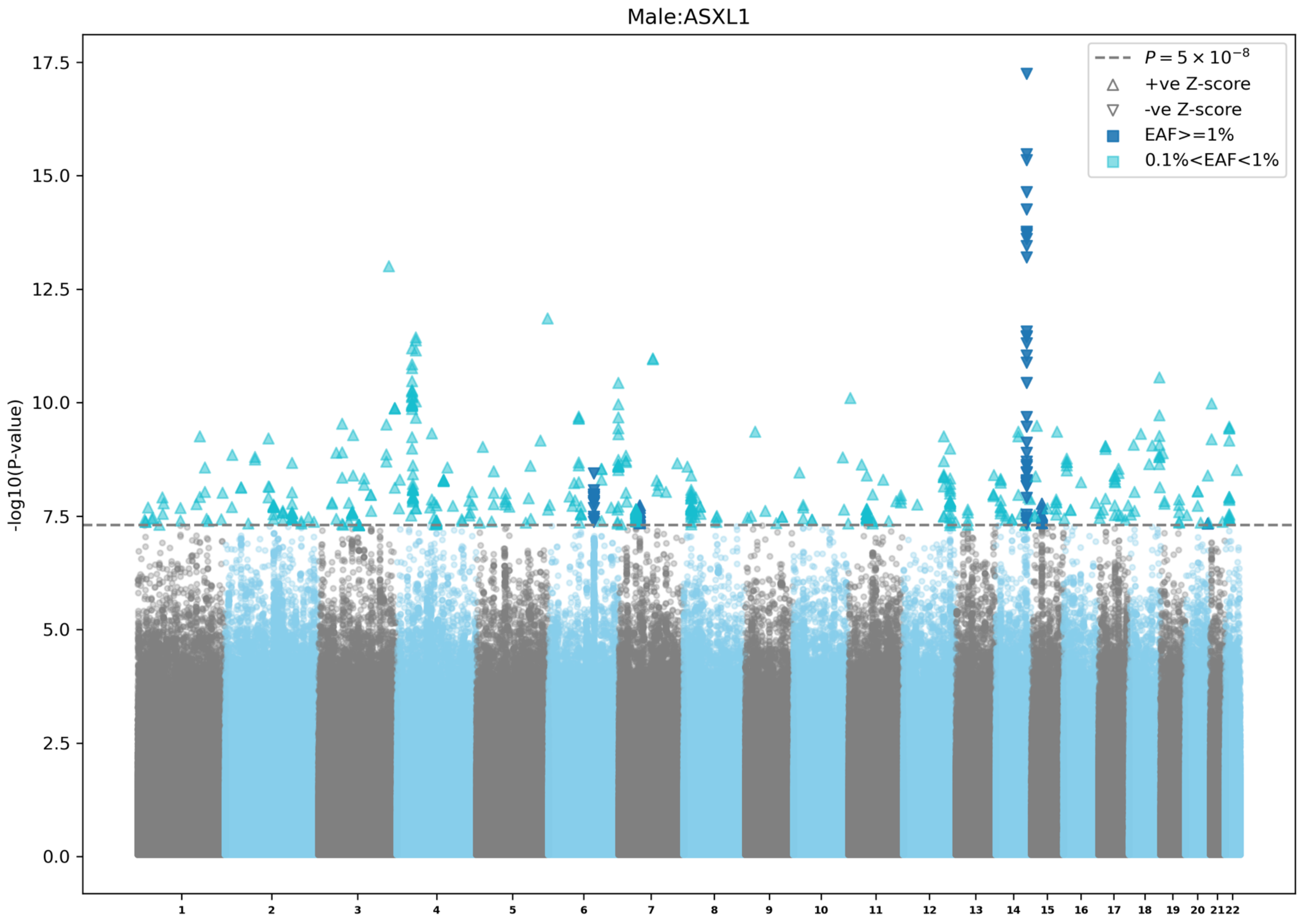

Male:ASXL1
-log10(P-value)
17.5
15.0
12.5
10.0
7.5
5.0
2.5
0.0
$P = 5 \times 10^{-8}$
+ve Z-score
-ve Z-score
EAF>=1%
0.1%<EAF<1%

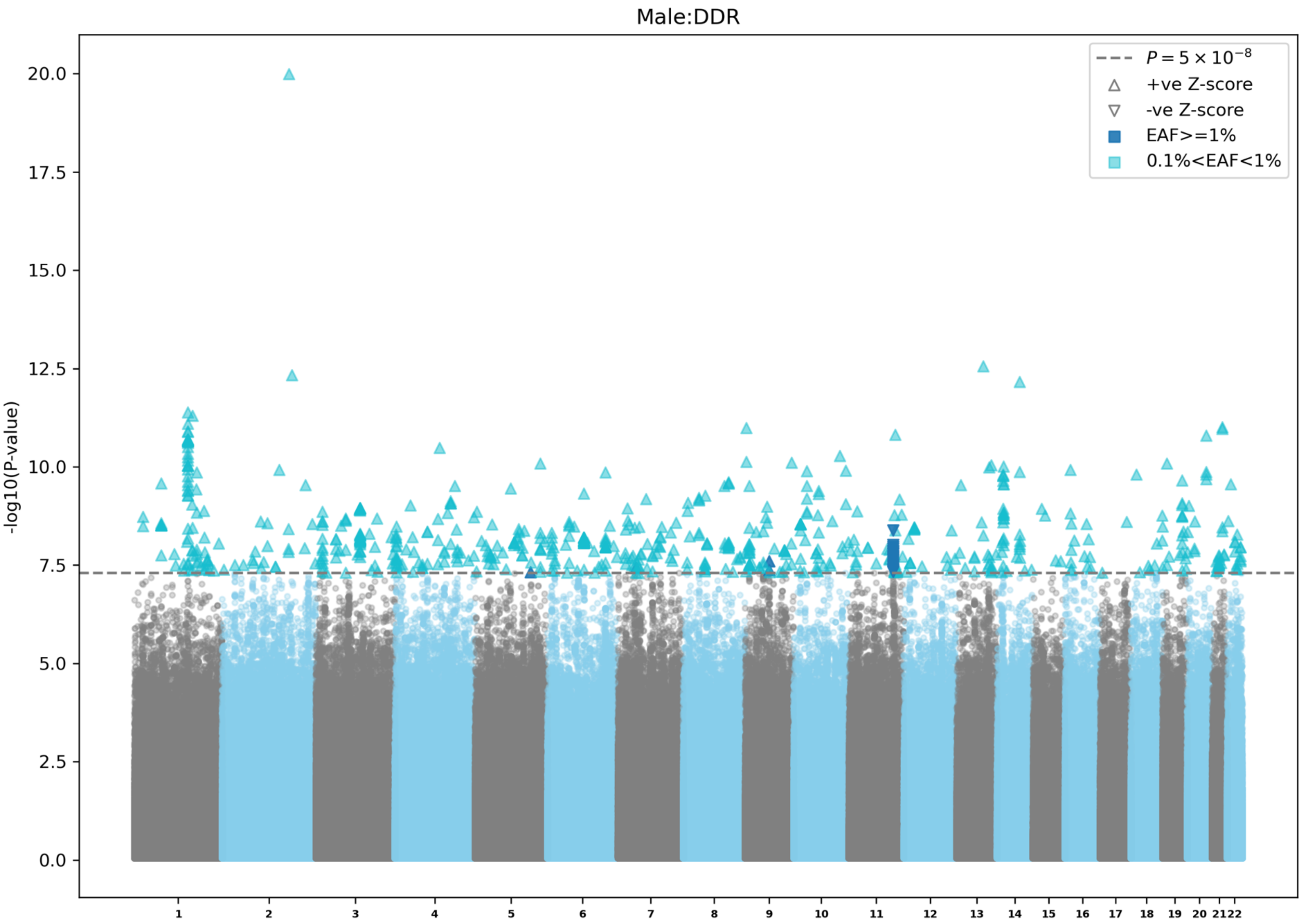

Male:DDR
-log10(P-value)
20.0
17.5
15.0
12.5
10.0
7.5
5.0
2.5
0.0
$P = 5 \times 10^{-8}$
+ve Z-score
-ve Z-score
EAF>=1%
0.1%<EAF<1%

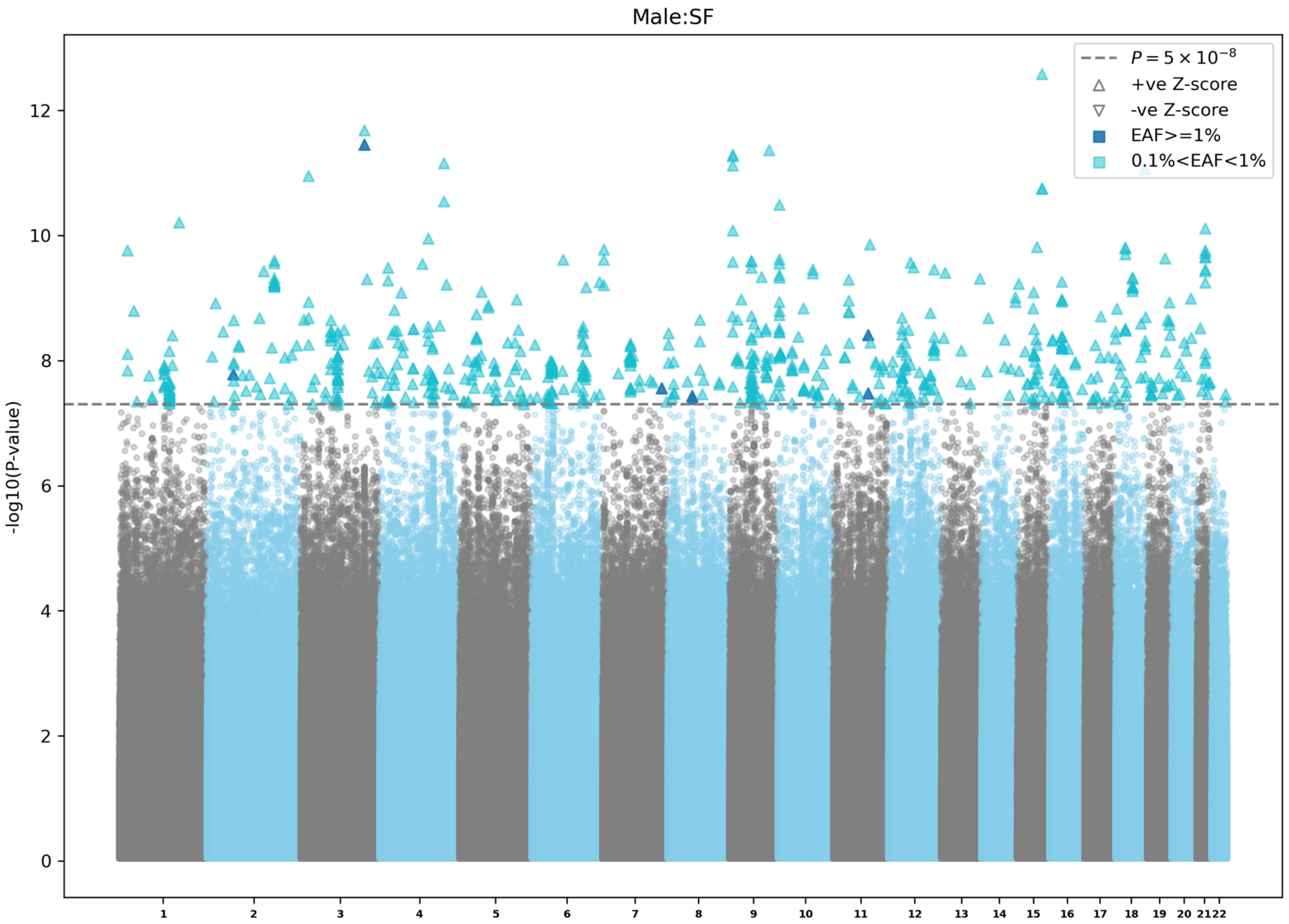

Male:SF
$P = 5 \times 10^{-8}$
+ve Z-score
-ve Z-score
EAF>=1%
0.1%<EAF<1%
-log10(P-value)
0
2
4
6
8
10
12
1 2 3 4 5 6 7 8 9 10 11 12 13 14 15 16 17 18 19 20 21 22

**Extended Data Figure 7 | Manhattan plots for male specific CHIP GWAS meta-analyses.** a) Overall CHIP GWAS in UKB, AoU, TOPMed, and MGBB. b) Expanded CHIP (VAF≥10%) GWAS in UKB, AoU, TOPMed, and MGBB. c) *DNMT3A* CHIP GWAS in UKB, AoU, TOPMed, and MGBB. d) *TET2* CHIP GWAS in UKB, AoU, TOPMed, and MGBB. e) *ASXL1* CHIP GWAS in UKB, AoU, TOPMed, and MGBB. f) DDR CHIP GWAS in UKB, AoU, TOPMed, and MGBB. g) SF CHIP GWAS in UKB, AoU, TOPMed, and MGBB. EAF: effect allele frequency; UKB: UK biobank; AoU: All of Us Research; TOPMed: Trans-omics for Precision medicine; MGBB: Mass General and Brigham biobank; CHIP: clonal hematopoiesis of indeterminate potential; DDR: DNA damage response genes (*TP53/PPM1D*); SF: splicing factors (*SF3B1/SRSF2/U2AF1/ZRSR2*).

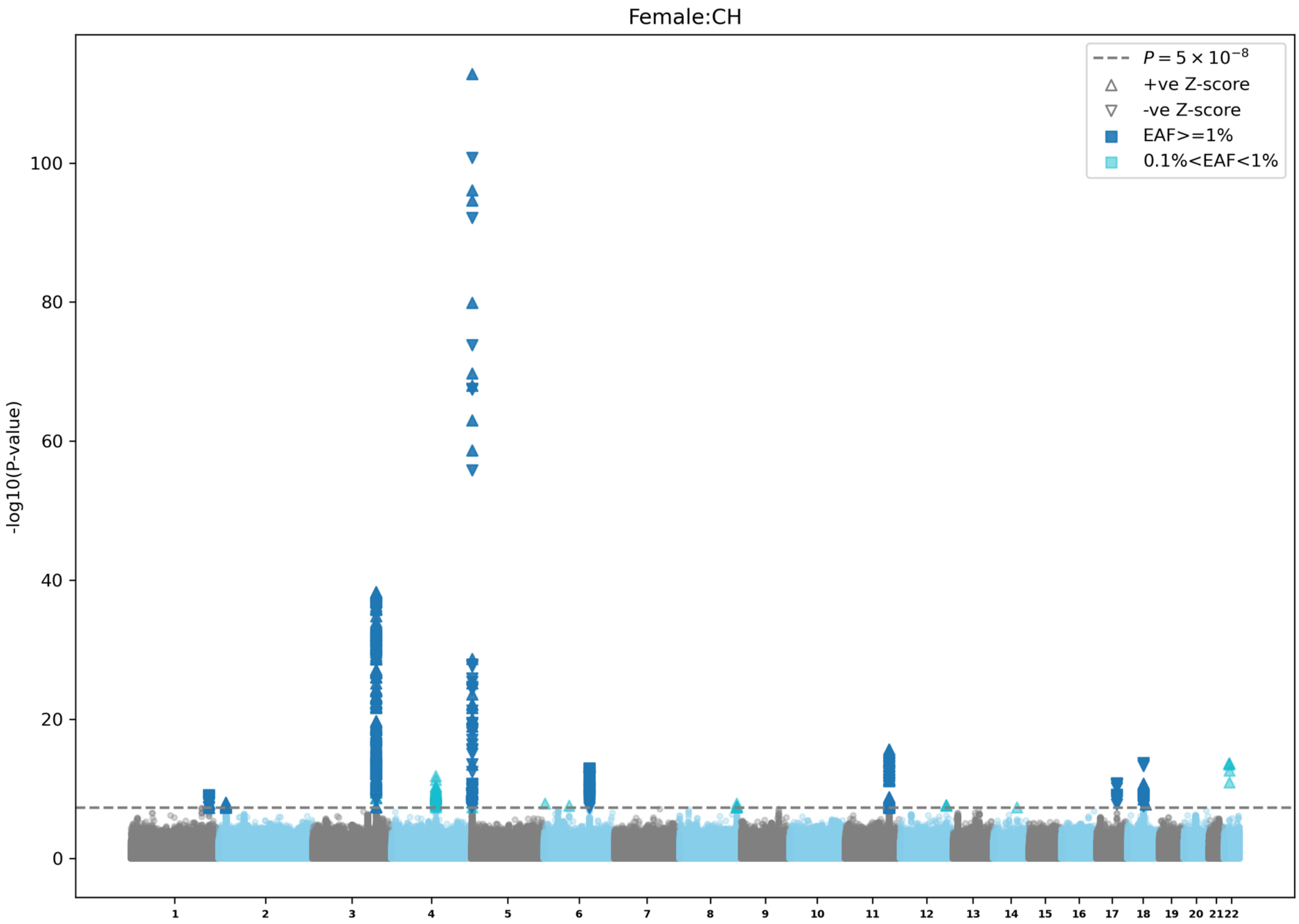
Female:CH
$P = 5 \times 10^{-8}$
+ve Z-score
-ve Z-score
EAF>=1%
0.1%<EAF<1%
-log10(P-value)
0
20
40
60
80
100

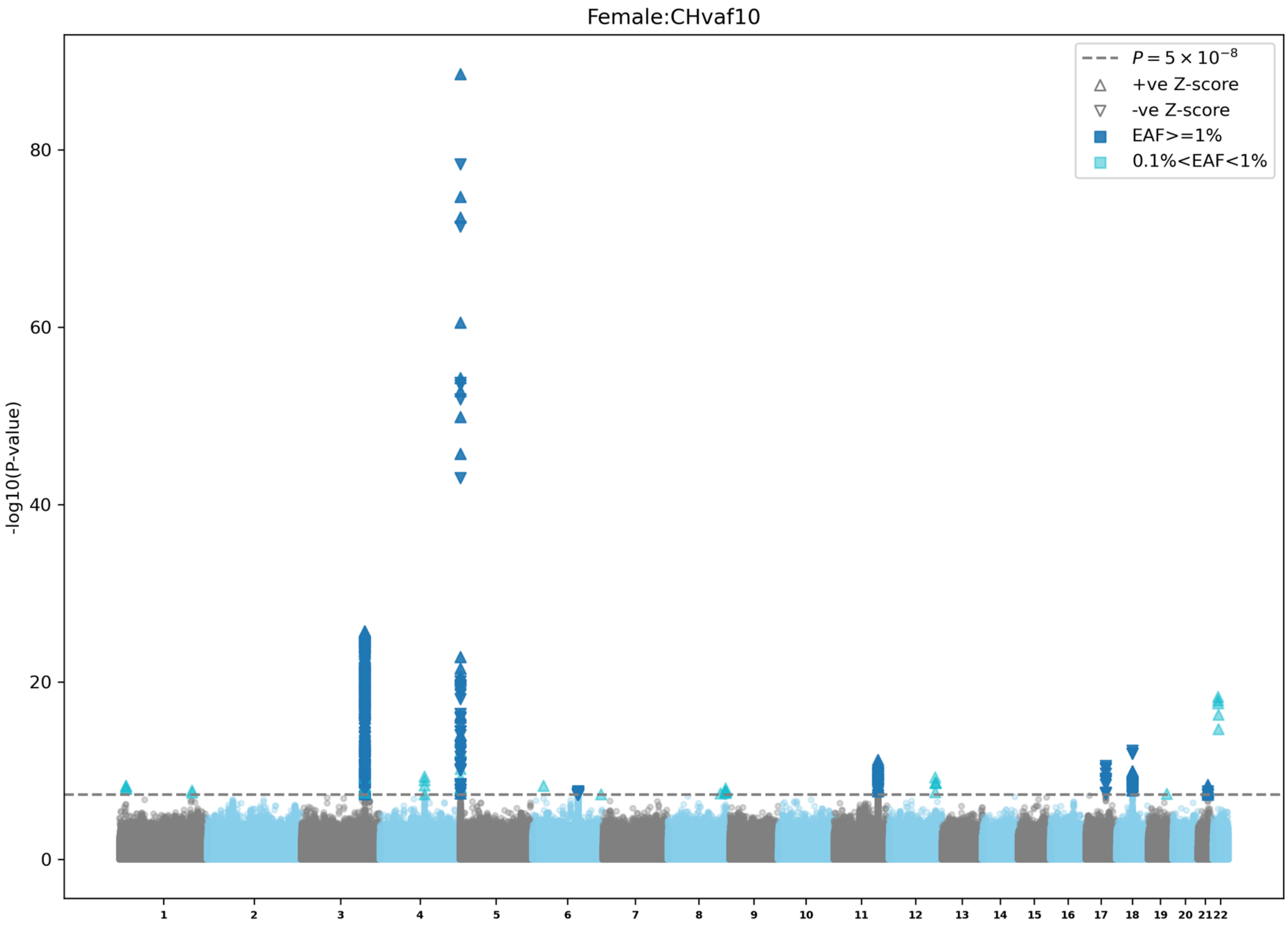

Female:CHvaf10
$P = 5 \times 10^{-8}$
+ve Z-score
-ve Z-score
EAF>=1%
0.1%<EAF<1%
-log10(P-value)
0
20
40
60
80

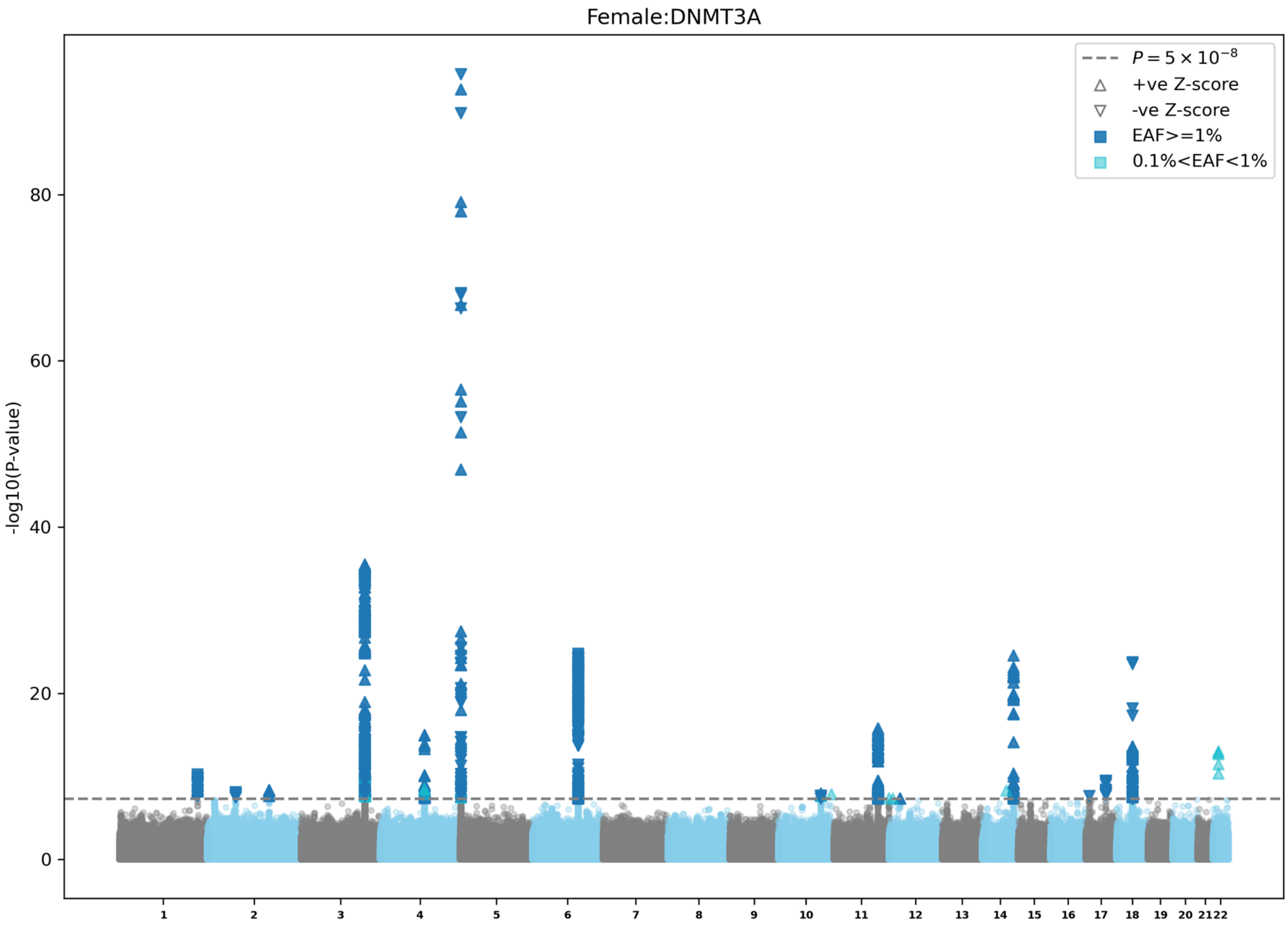

Female:DNMT3A
-log10(P-value)
0
20
40
60
80
$P = 5 \times 10^{-8}$
+ve Z-score
-ve Z-score
EAF>=1%
0.1%<EAF<1%

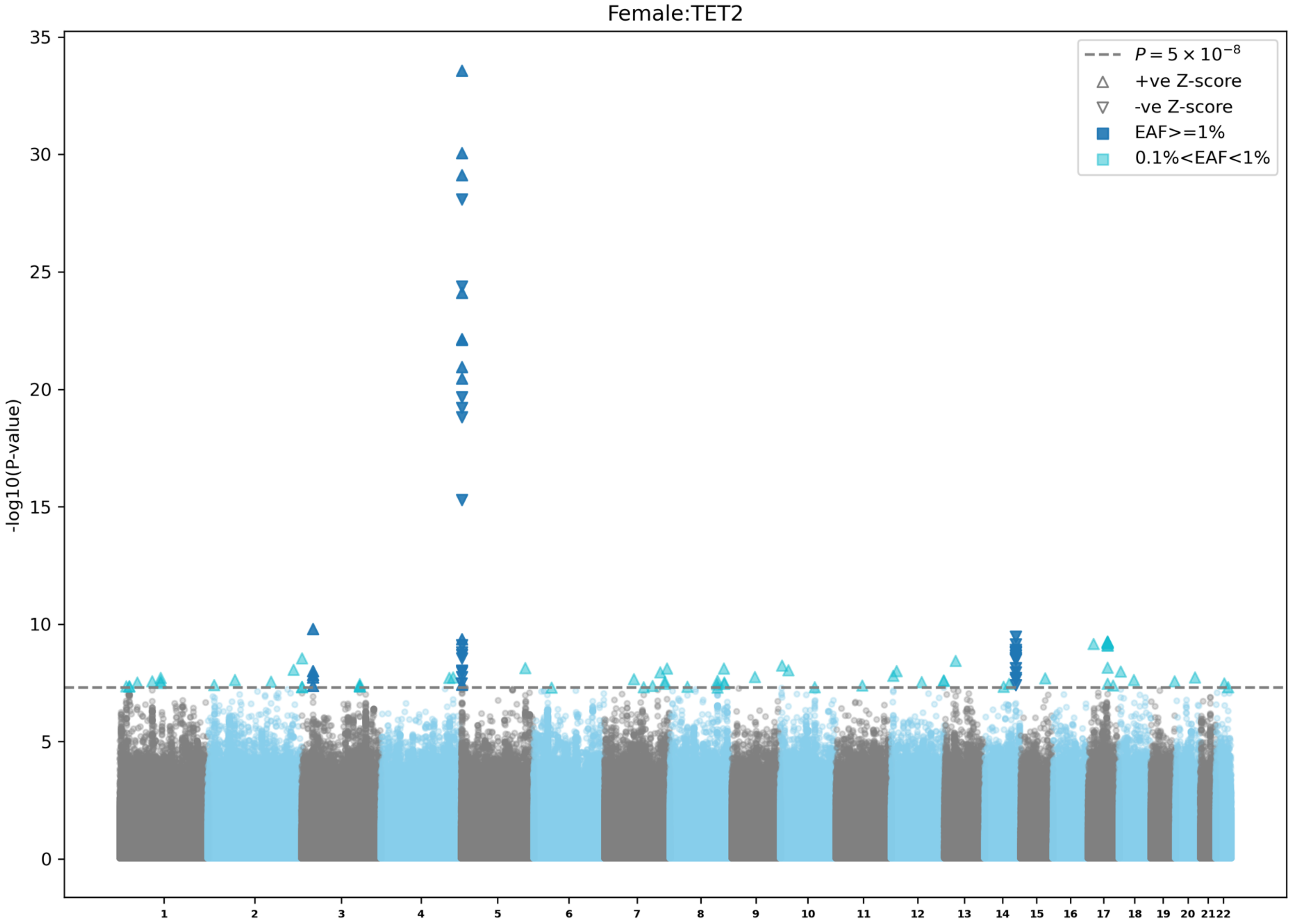

Female:TET2
-log10(P-value)
35
30
25
20
15
10
5
0
$P = 5 \times 10^{-8}$
+ve Z-score
-ve Z-score
EAF>=1%
0.1%<EAF<1%

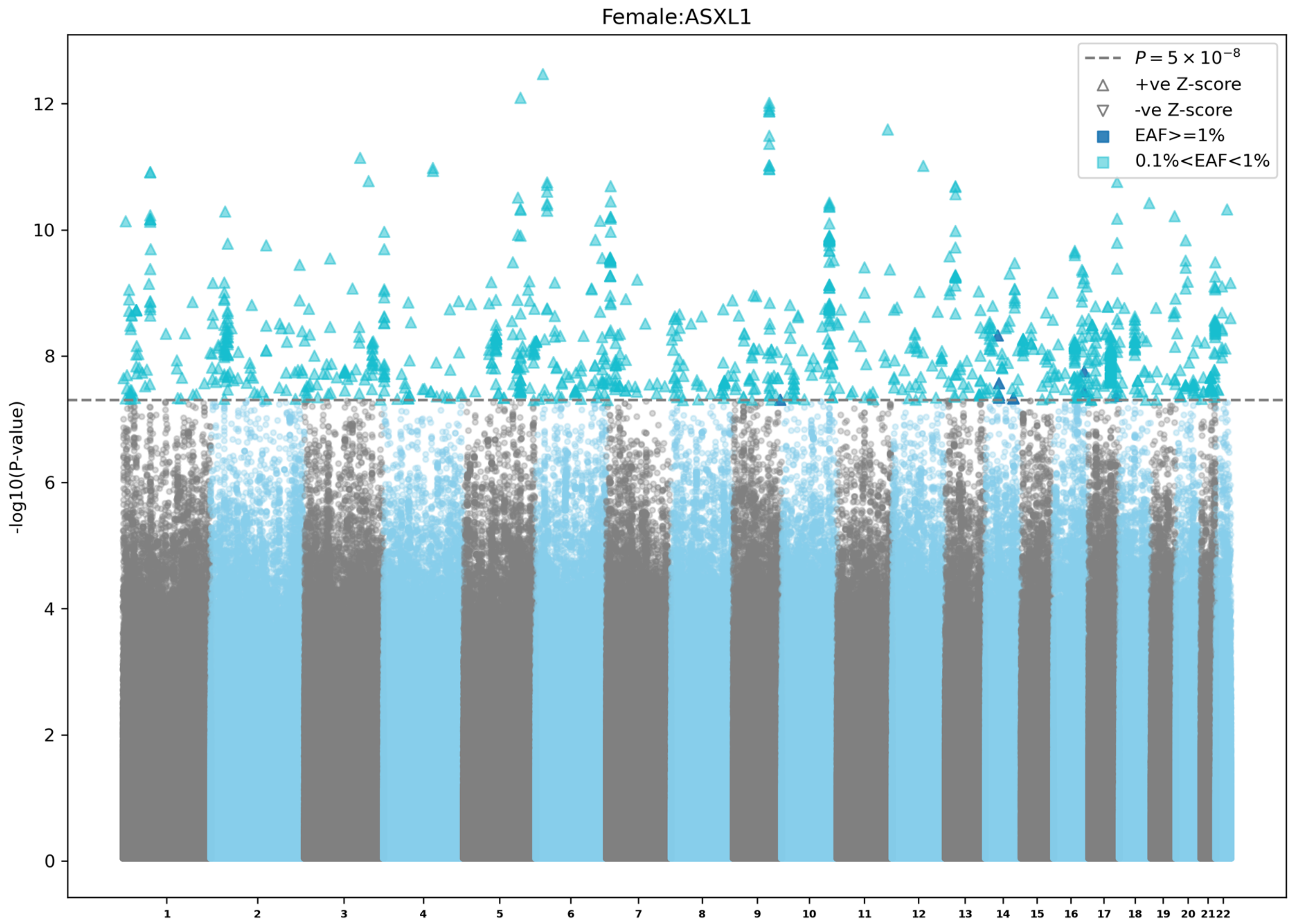

Female:ASXL1
$P = 5 \times 10^{-8}$
+ve Z-score
-ve Z-score
EAF>=1%
0.1%<EAF<1%
-log10(P-value)
0
2
4
6
8
10
12

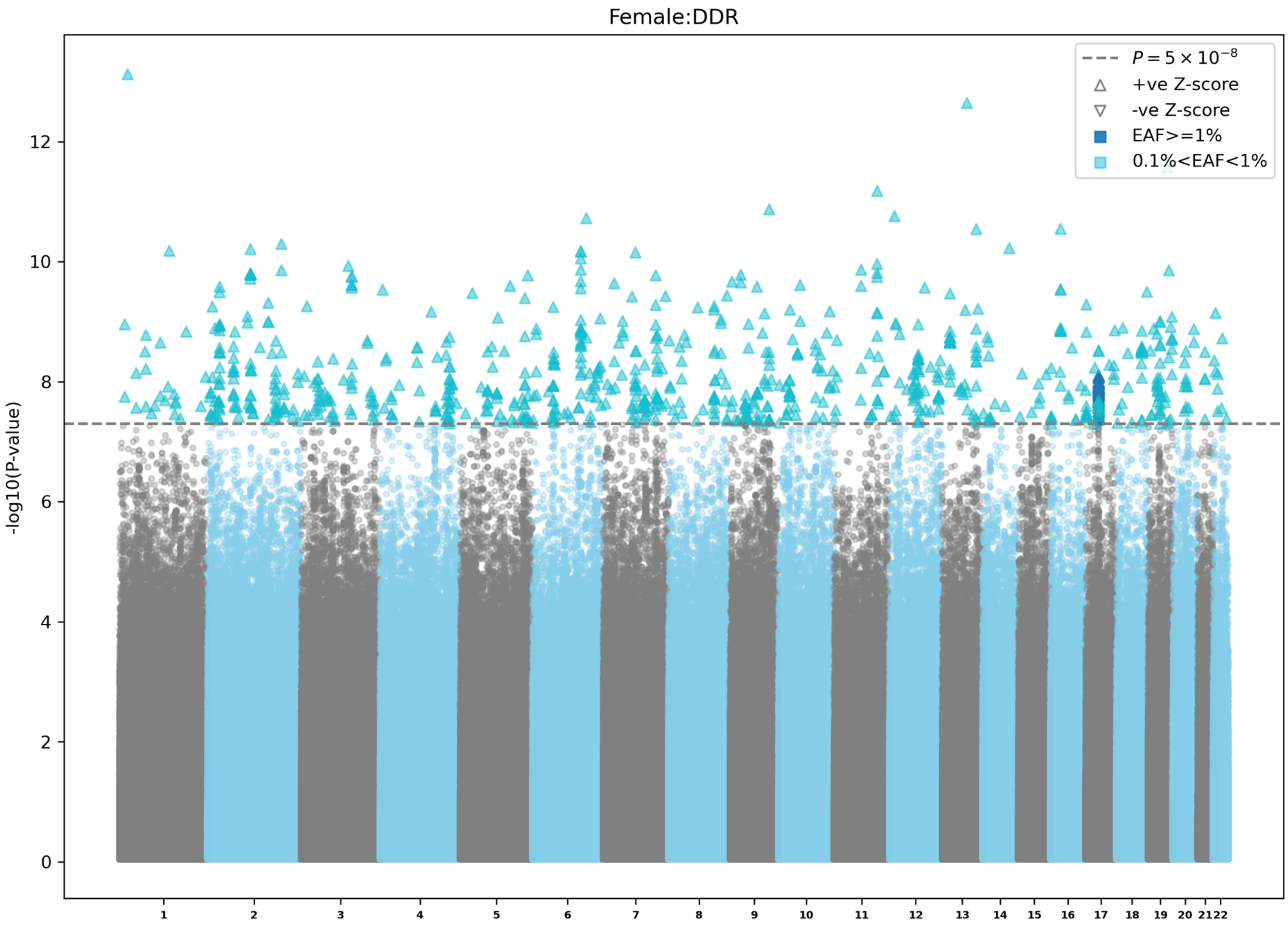
Female:DDR
-log10(P-value)
$P = 5 \times 10^{-8}$
+ve Z-score
-ve Z-score
EAF>=1%
0.1%<EAF<1%
0
2
4
6
8
10
12

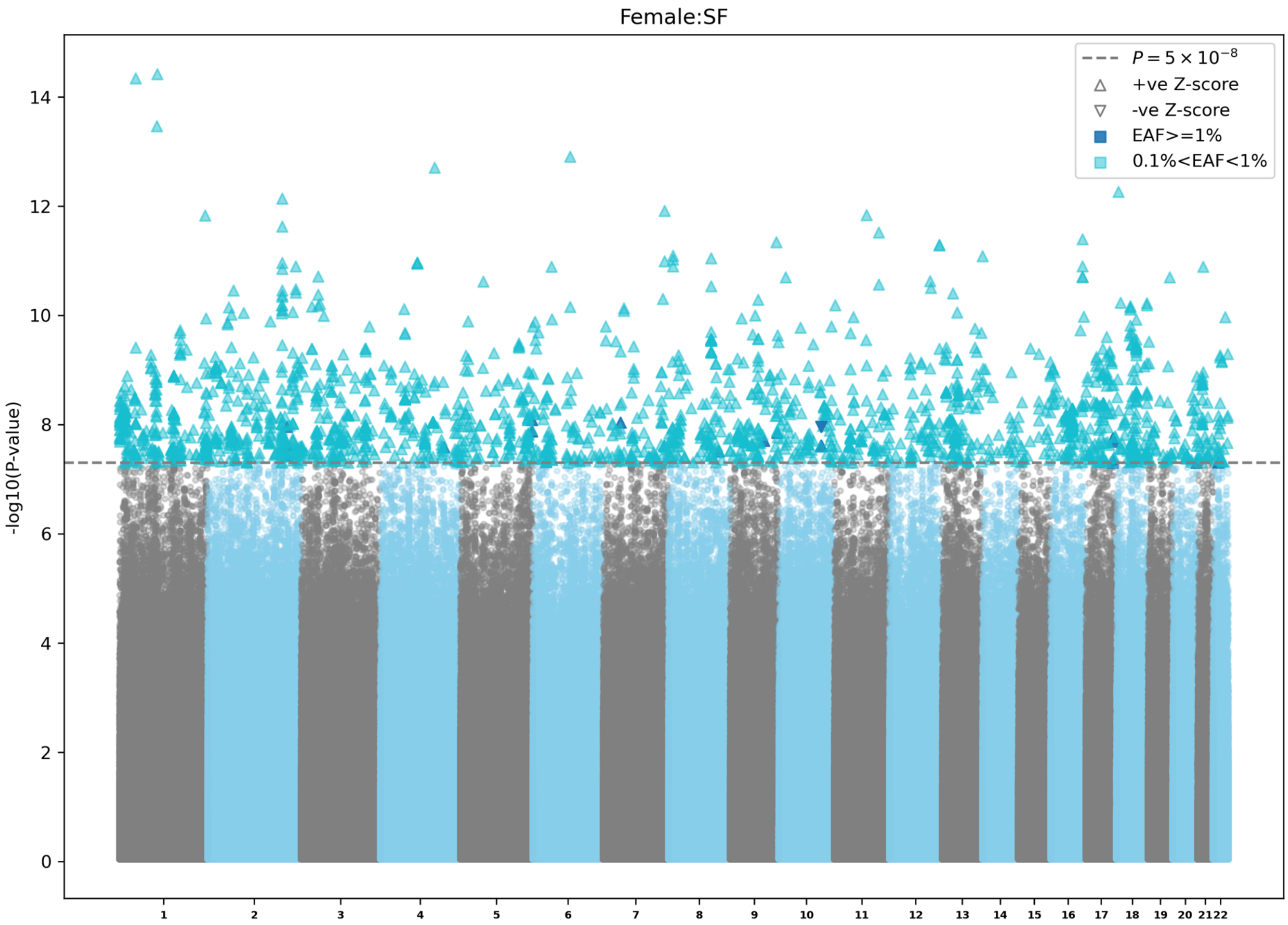

Female:SF
$P = 5 \times 10^{-8}$
+ve Z-score
-ve Z-score
EAF>=1%
0.1%<EAF<1%
-log10(P-value)
0
2
4
6
8
10
12
14
1 2 3 4 5 6 7 8 9 10 11 12 13 14 15 16 17 18 19 20 21 22

**Extended Data Figure 8 | Manhattan plots for female specific CHIP GWAS meta-analyses.** a) Overall CHIP GWAS in UKB, AoU, TOPMed, and MGBB. b) Expanded CHIP (VAF≥10%) GWAS in UKB, AoU, TOPMed, and MGBB. c) *DNMT3A* CHIP GWAS in UKB, AoU, TOPMed, and MGBB. d) *TET2* CHIP GWAS in UKB, AoU, TOPMed, and MGBB. e) *ASXL1* CHIP GWAS in UKB, AoU, TOPMed, and MGBB. f) DDR CHIP GWAS in UKB, AoU, TOPMed, and MGBB. g) SF CHIP GWAS in UKB, AoU, TOPMed, and MGBB. EAF: effect allele frequency; UKB: UK biobank; AoU: All of Us Research; TOPMed: Trans-omics for Precision medicine; MGBB: Mass General and Brigham biobank; CHIP: clonal hematopoiesis of indeterminate potential; DDR: DNA damage response genes (*TP53/PPM1D*); SF: splicing factors (*SF3B1/SRSF2/U2AF1/ZRSR2*).

#### a) Multi-ancestry meta-GWAS of CHIP traits

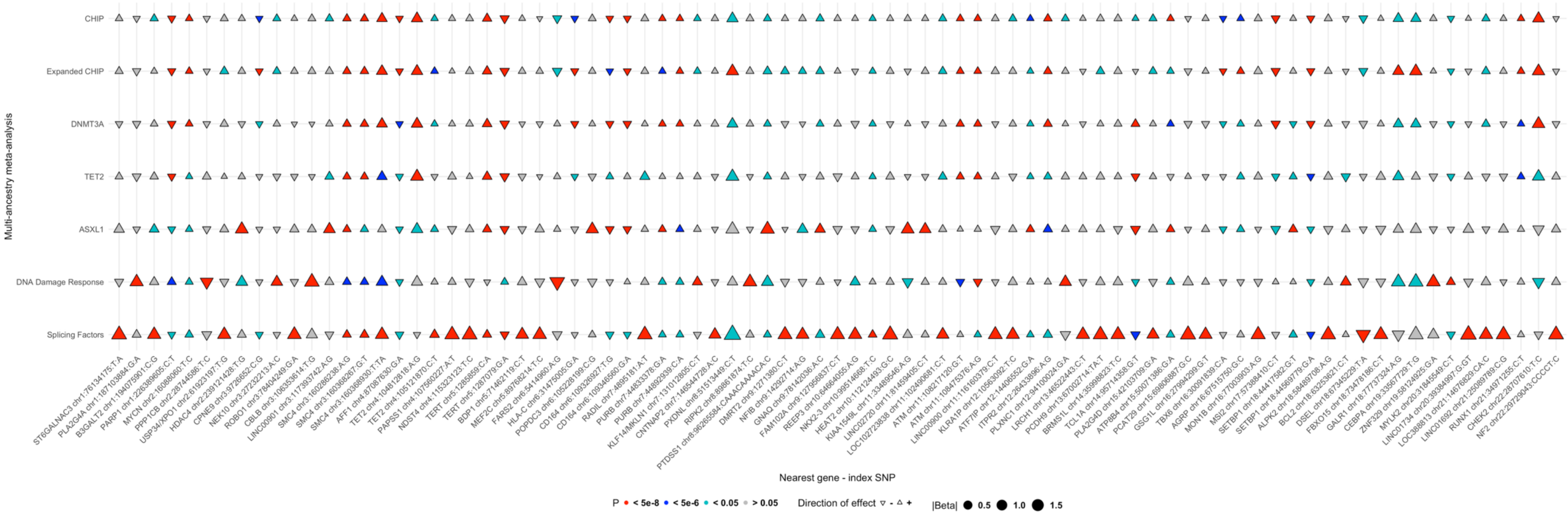

**b) Stratified meta-GWAS of overall CHIP**

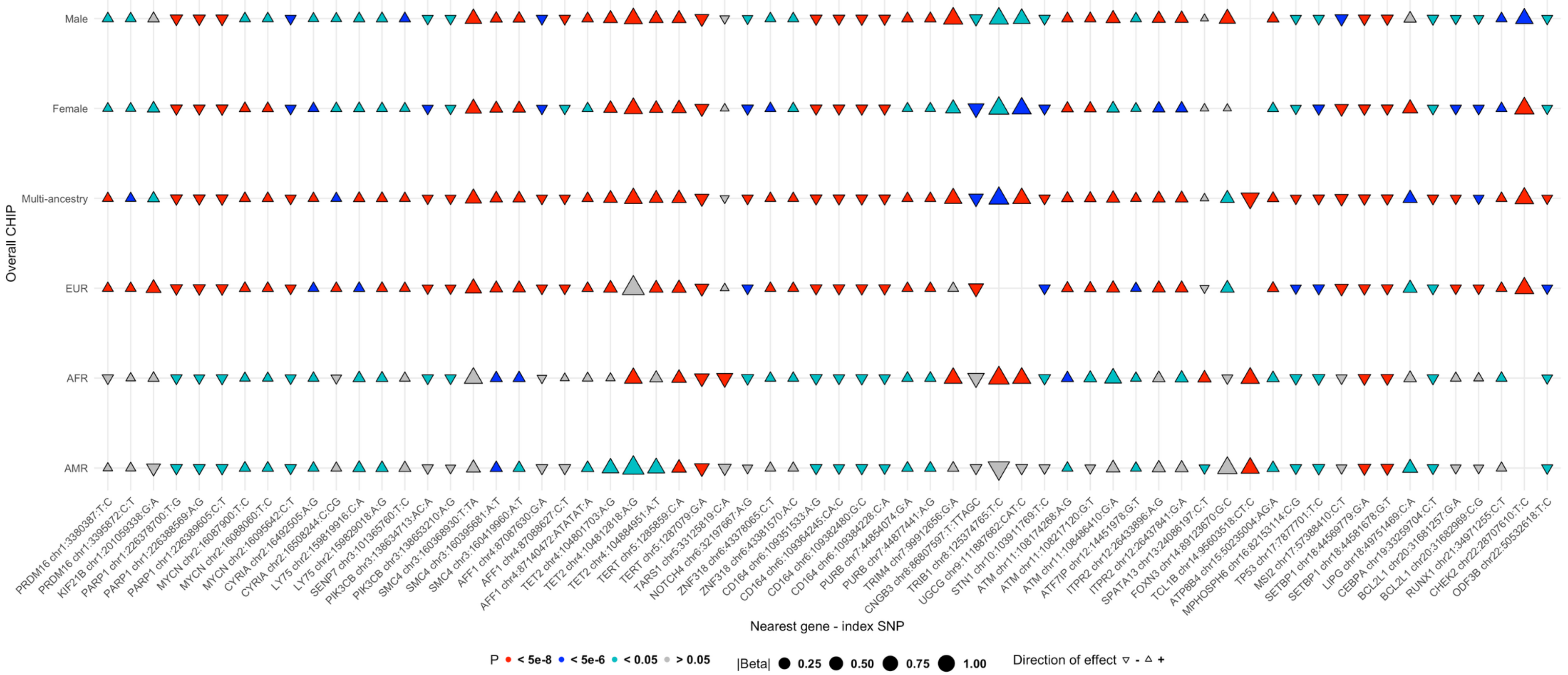

#### c) Stratified meta-GWAS of expanded CHIP

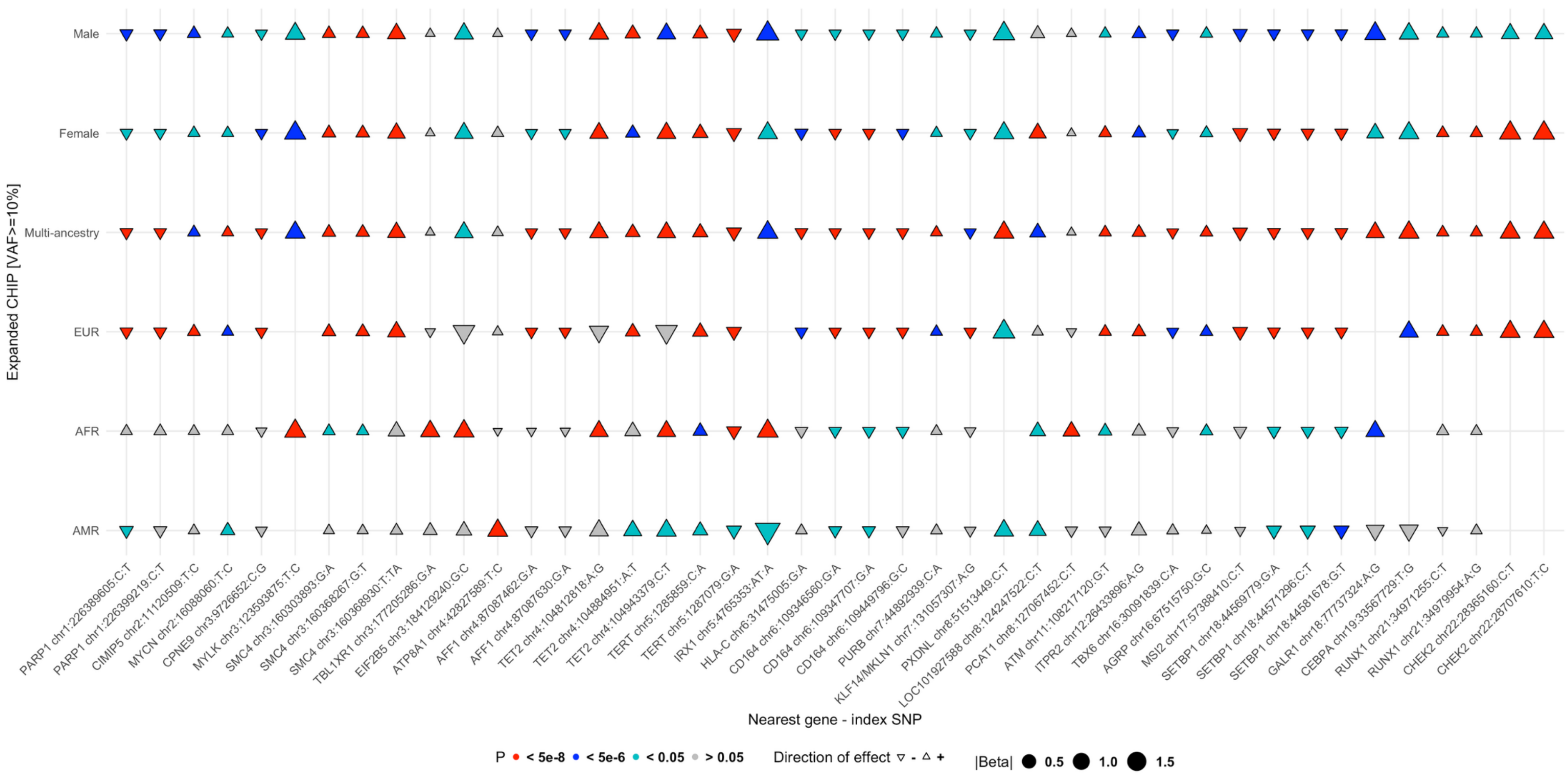

**d) Stratified meta-GWAS of *DNMT3A* CHIP**

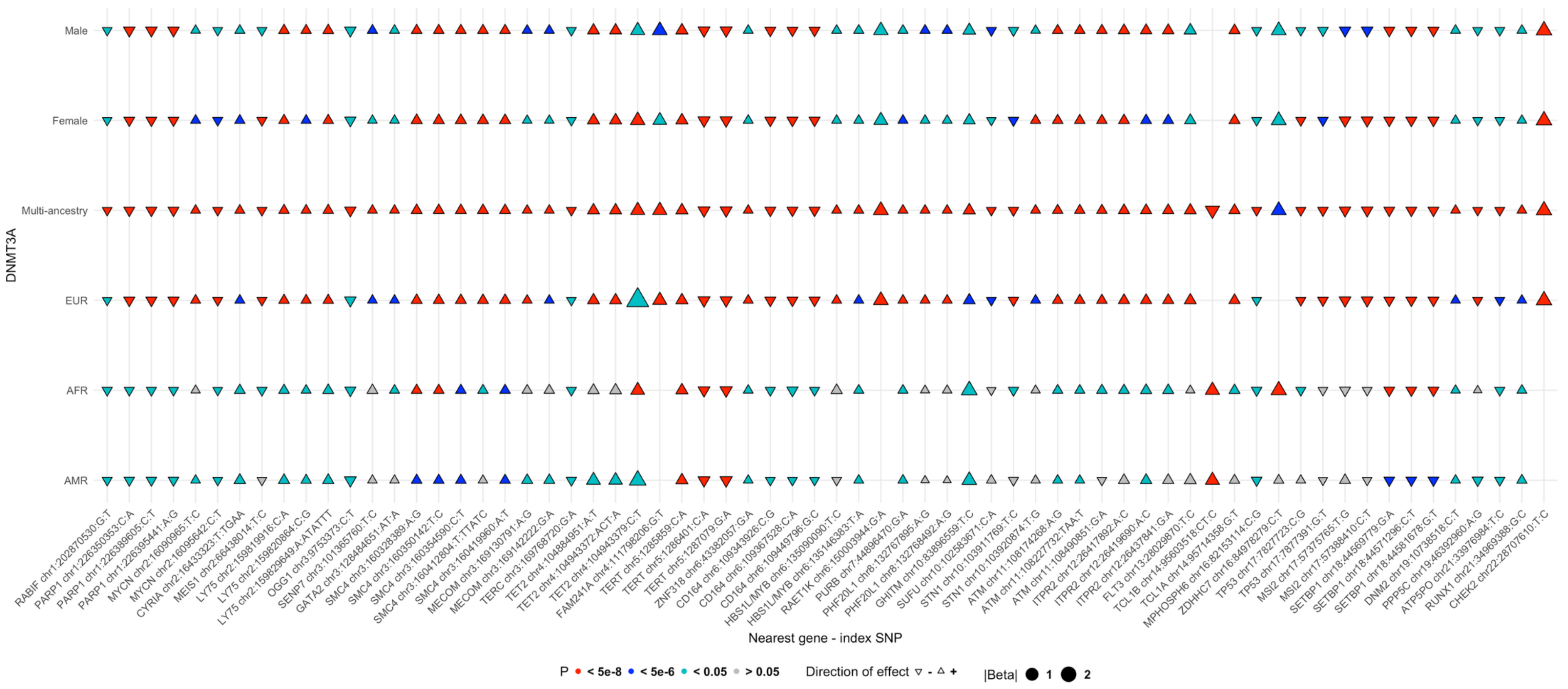

**e) Stratified meta-GWAS of *TET2* CHIP**

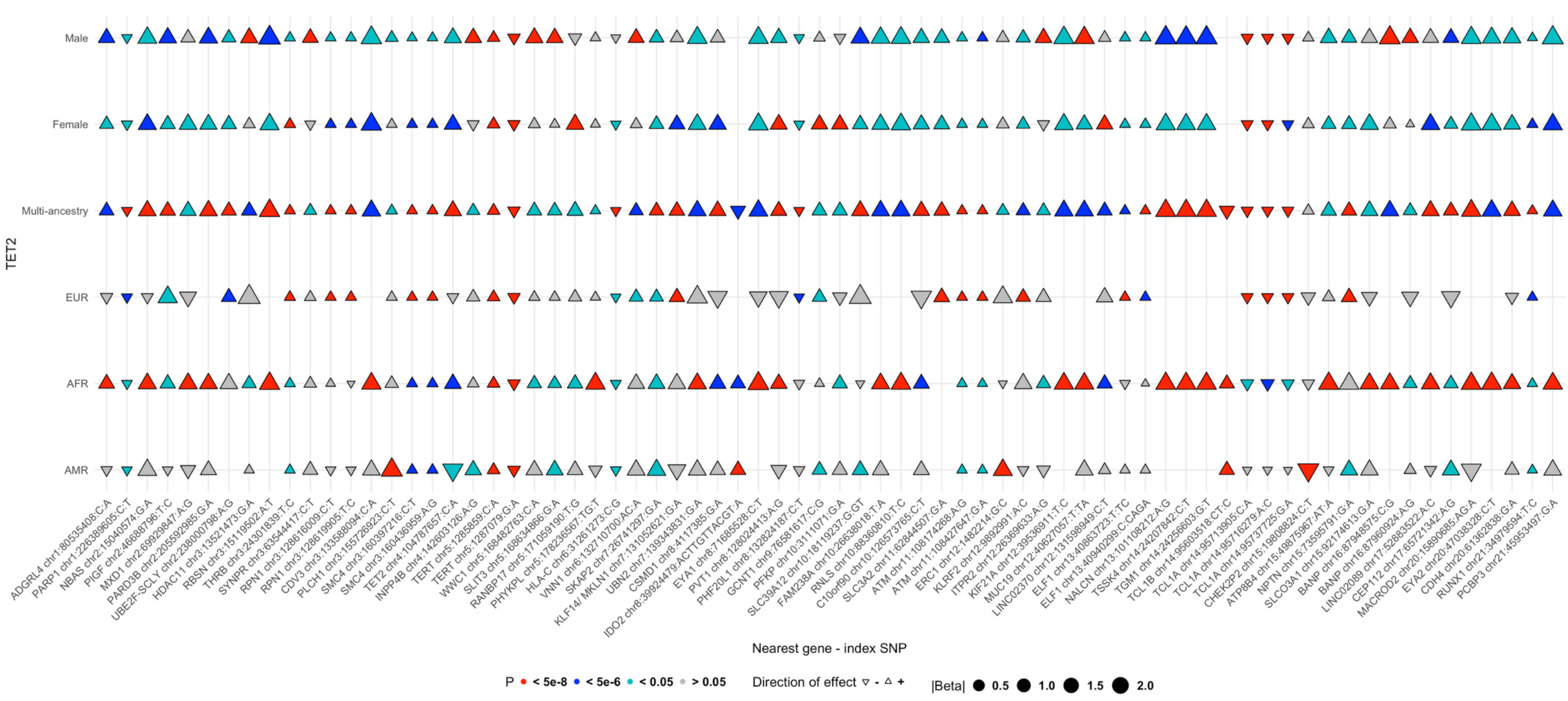

#### f) Stratified meta-GWAS of *ASXL1* CHIP

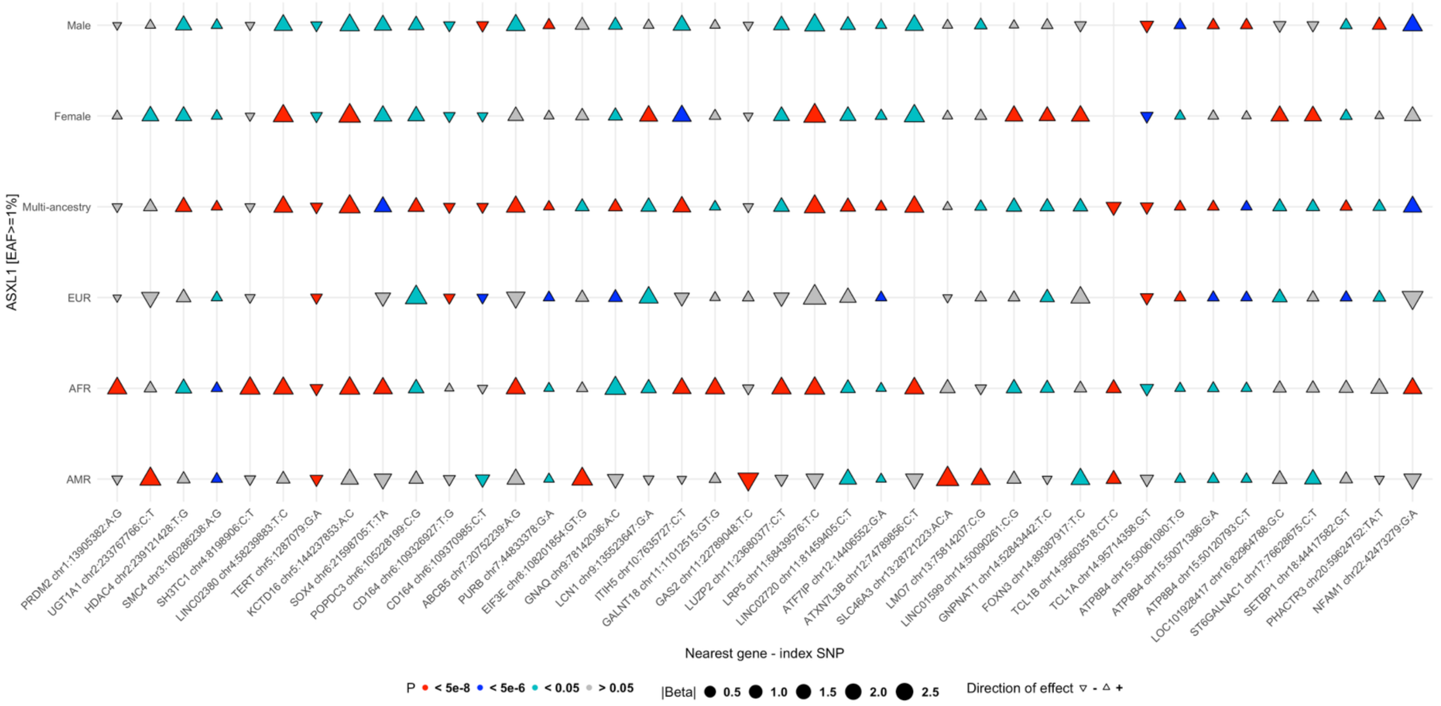

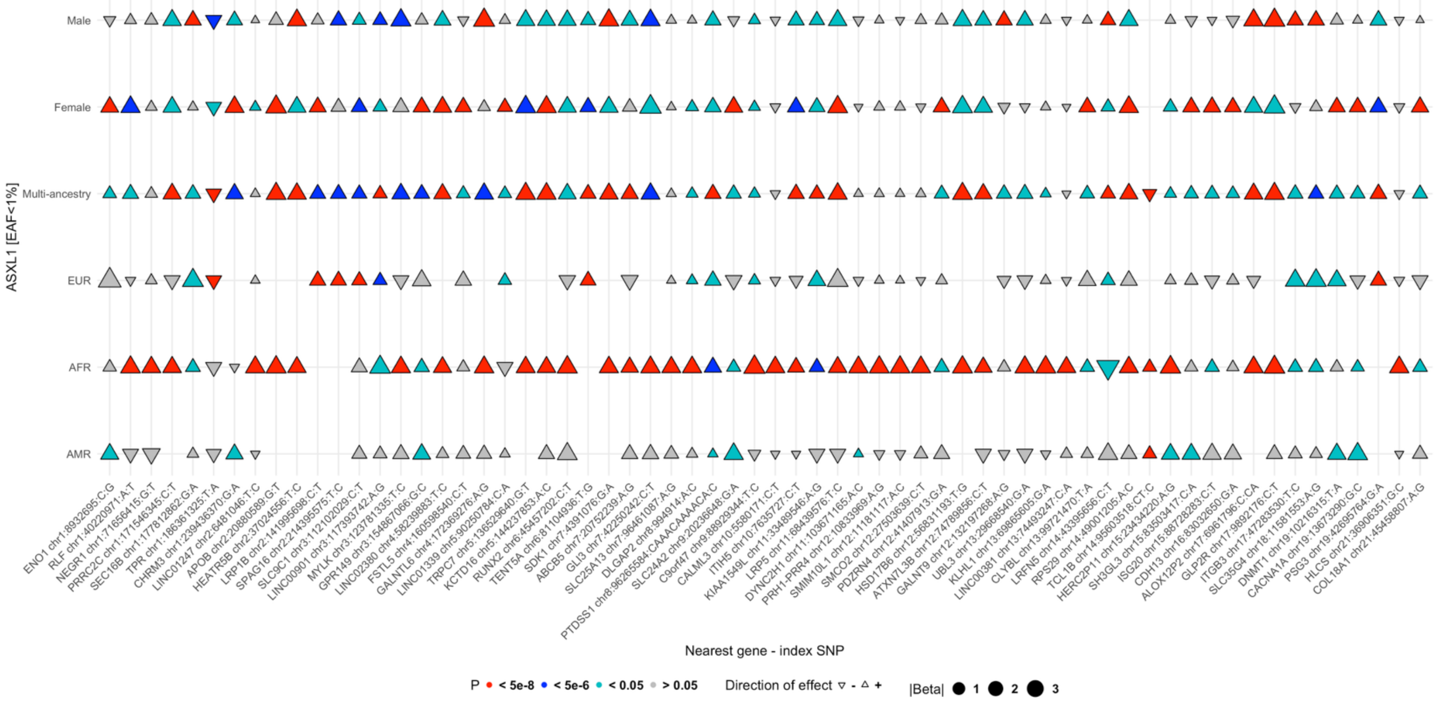

#### g) Stratified meta-GWAS of DNA damage response CHIP

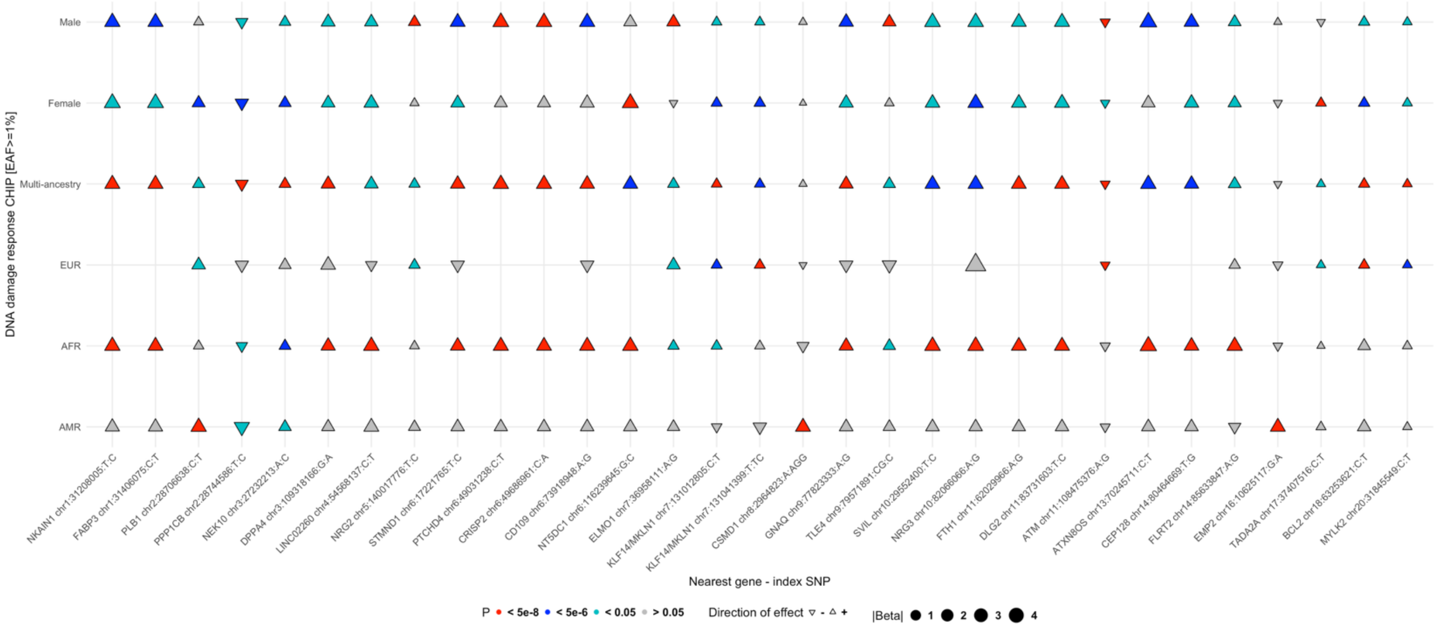

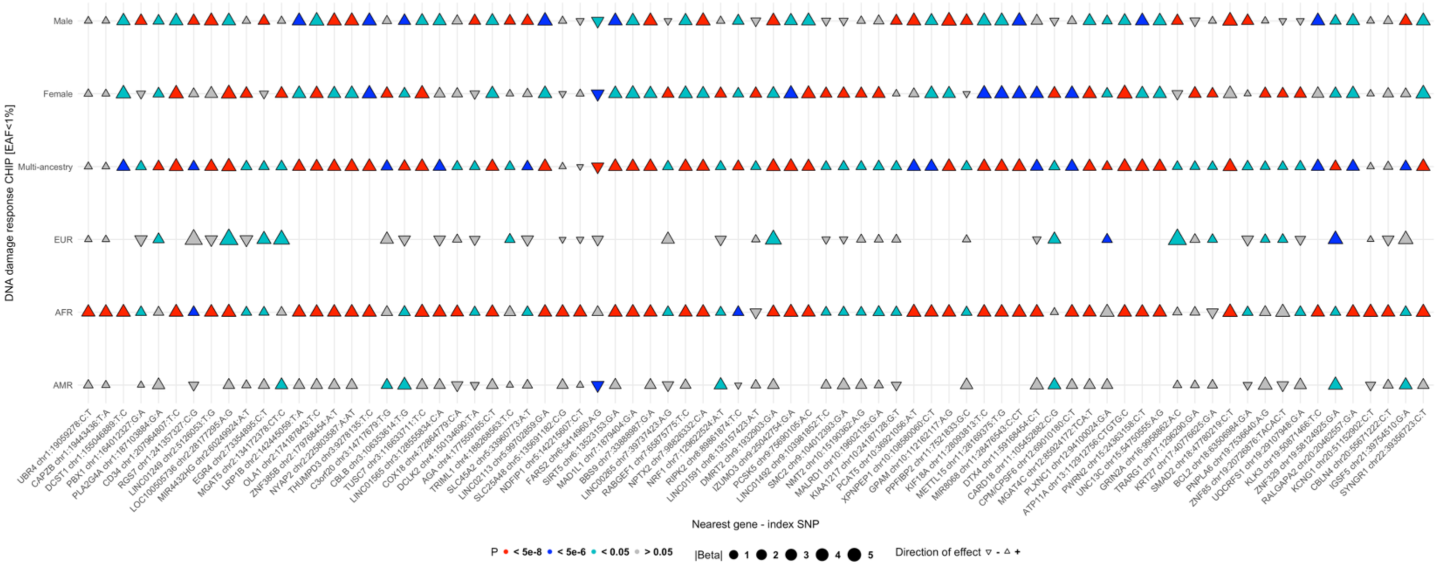

#### h) Stratified meta-GWAS of splicing factors CHIP

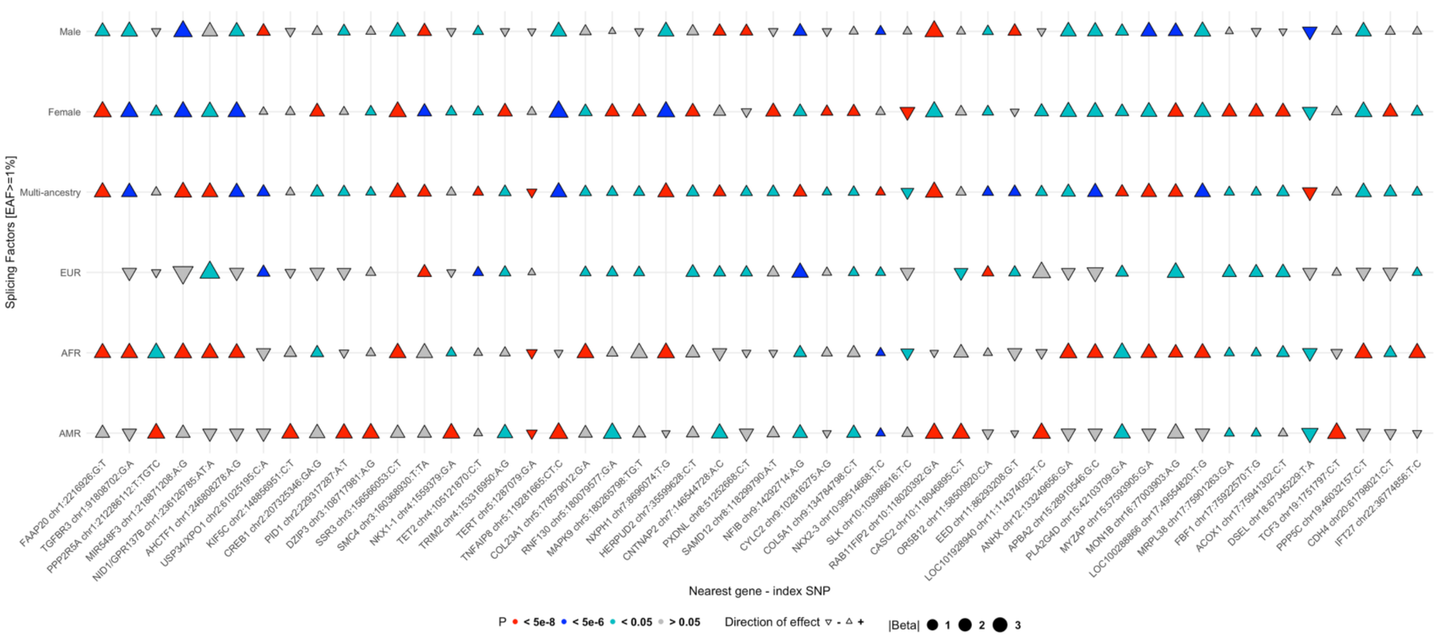

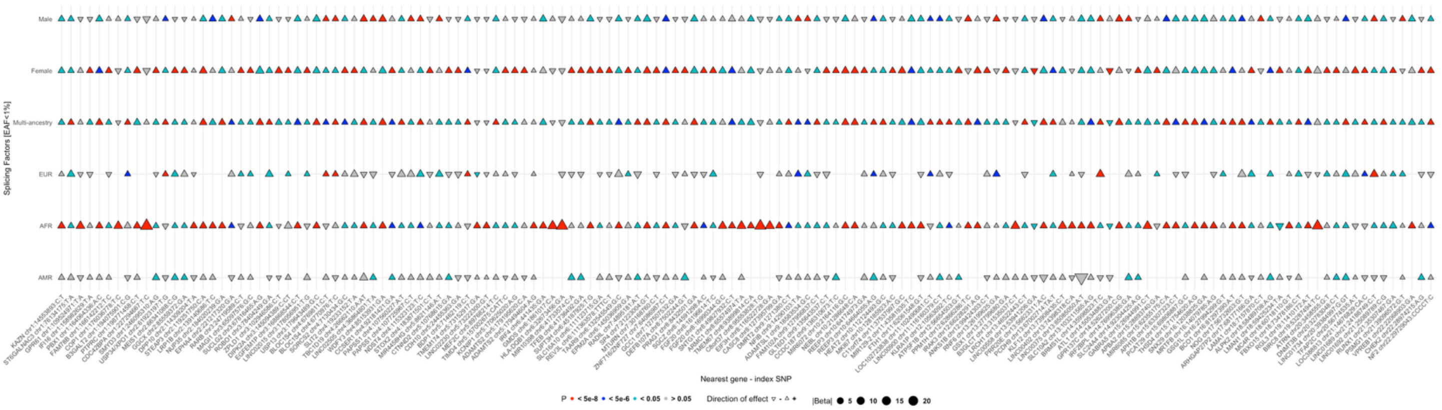

**Extended Data Figure 9 | Overlap of lead variants across CHIP categories.** Lead variants from overall CHIP, expanded CHIP (variant allele fraction VAF>=10%), *DNMT3A*, *TET2*, *ASXL1*, DNA damage response (*TP53/PPM1D)* and splicing factors (*SF3B1/SRSF2/U2AF1/ZRSR2*) CHIP were compared with multi-ancestry and ancestry, sex and driver-gene stratified meta-analyses. a) Multi-ancestry meta-GWAS of CHIP traits: significant lead variants from expanded CHIP, *ASXL1*, DDR and SF CHIP GWAS was compared with CHIP categories. b) Stratified meta-GWAS of overall CHIP. c) Stratified meta-GWAS of expanded CHIP. d) Stratified meta-GWAS of *DNMT3A* CHIP. e) Stratified meta-GWAS of *TET2* CHIP. f) Stratified meta-GWAS of *ASXL1* CHIP. g) Stratified meta-GWAS of DNA damage response CHIP. h) Stratified meta-GWAS of splicing factors CHIP. CHIP: clonal hematopoiesis of indeterminate potential.

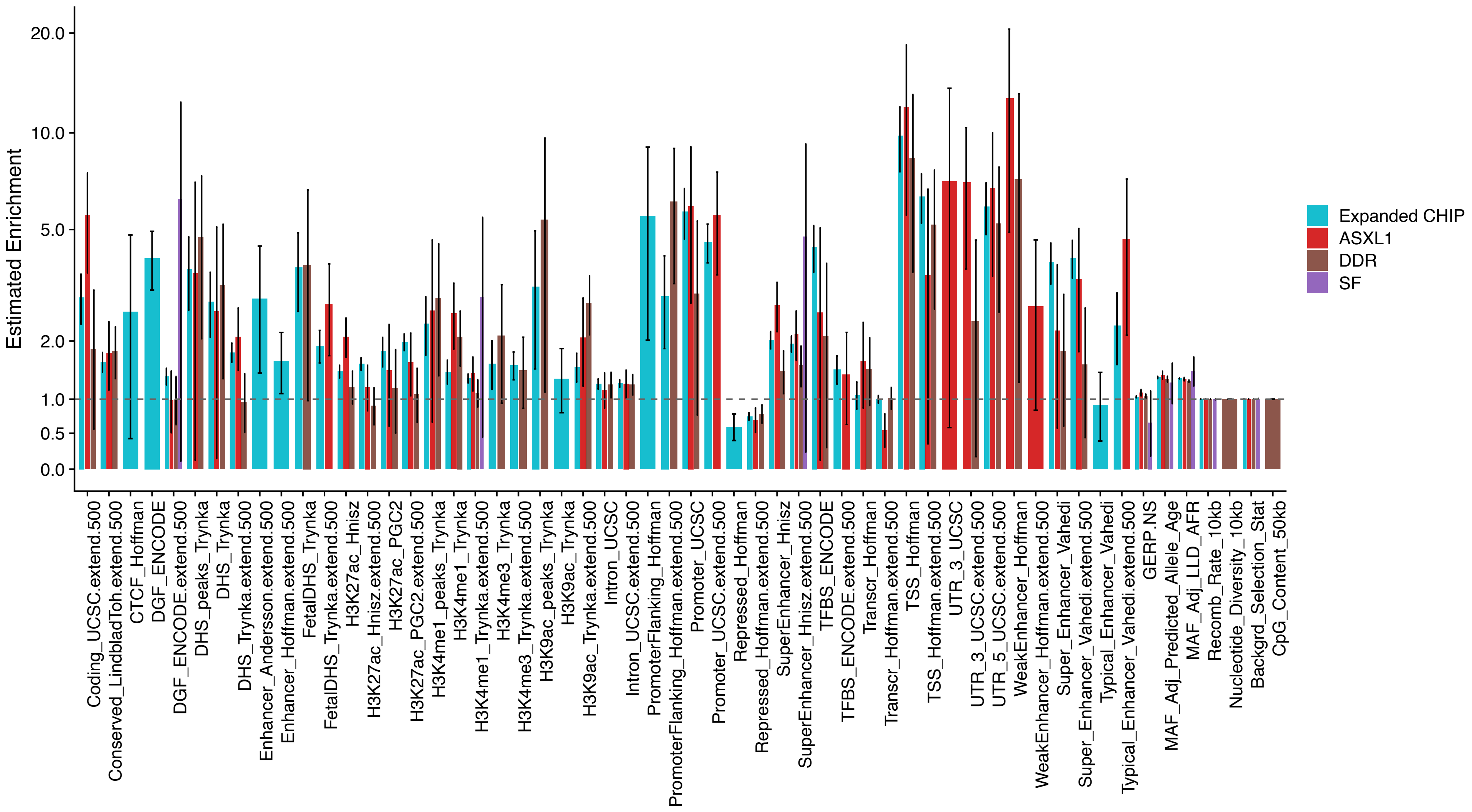

**Extended Data Figure 10 | Estimated enrichment of SNP heritability (± 1 SD) in genomic functional categories.** SNP heritability and enrichment were estimated using summary statistics from multi-ancestry meta-analyses of expanded CHIP (N=779,016), *ASXL1* (N=741,305), DDR (N=740,114), and SF (N=738,868) CHIP using LDAK 'SumHer' function. CHIP: clonal hematopoiesis of indeterminate potential; DDR: DNA damage response genes (*TP53/PPM1D*); SF: splicing factors (*SF3B1/SRSF2/U2AF1/ZRSR2*).

### Overall CHIP

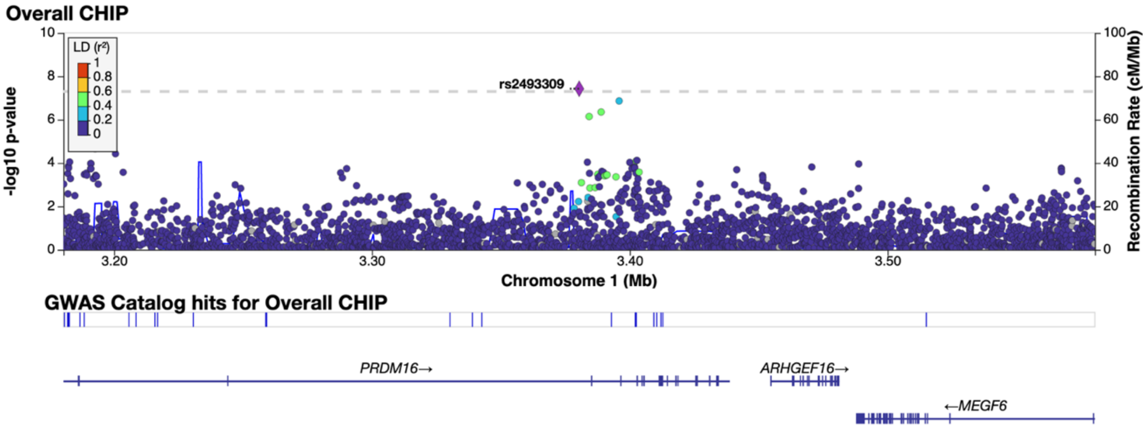

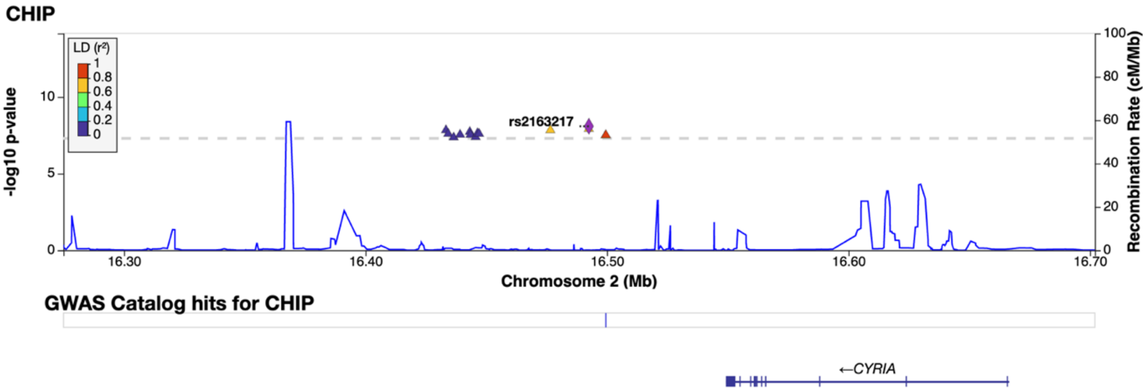

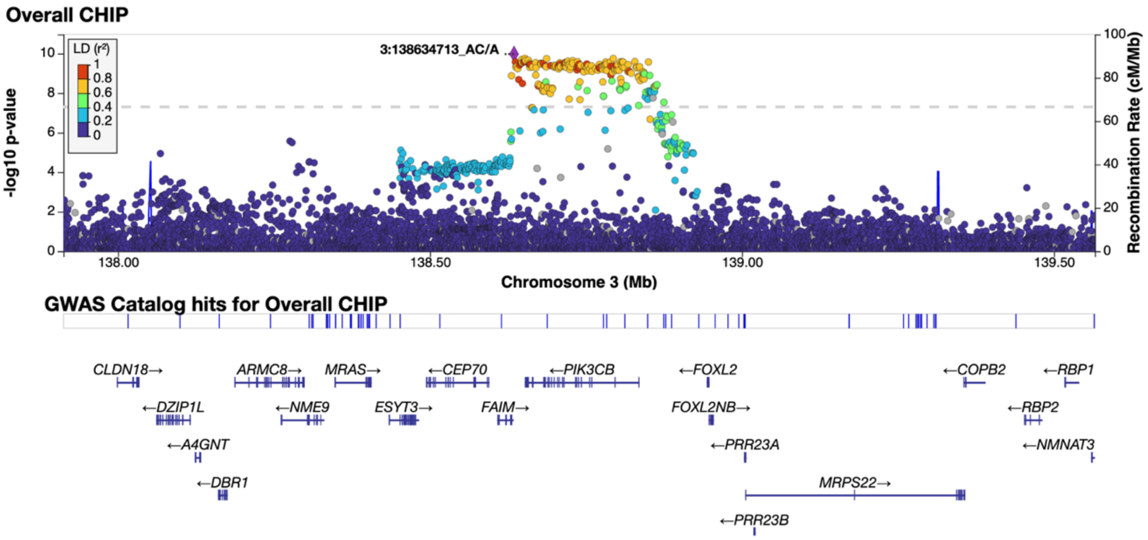

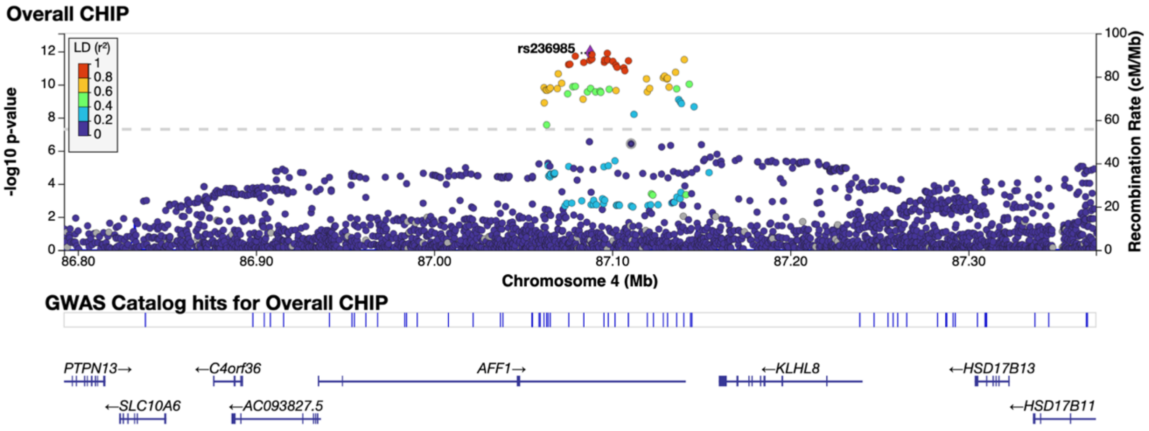

Overall CHIP
LD (r²)
1
0.8
0.6
0.4
0.2
0
rs236985
-log10 p-value
Recombination Rate (cM/Mb)
Chromosome 4 (Mb)
86.80
86.90
87.00
87.10
87.20
87.30
GWAS Catalog hits for Overall CHIP
PTPN13→
←C4orf36
AFF1→
←KLHL8
←HSD17B13
←SLC10A6
←AC093827.5
←HSD17B11

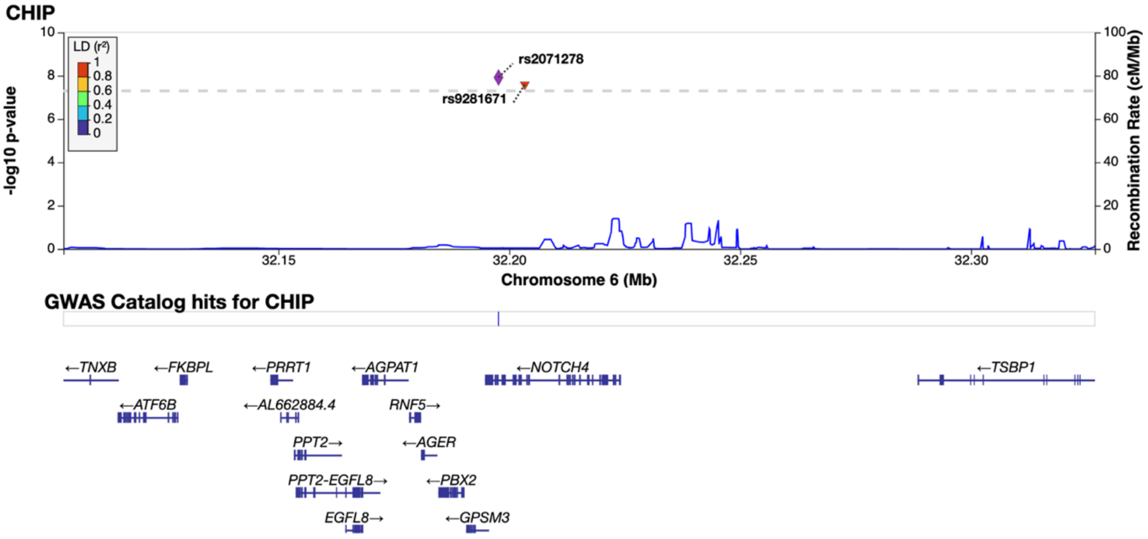

CHIP
LD (r²)
1
0.8
0.6
0.4
0.2
0
rs2071278
rs9281671
-log10 p-value
Recombination Rate (cM/Mb)
Chromosome 6 (Mb)
32.15
32.20
32.25
32.30
GWAS Catalog hits for CHIP
←TNXB
←FKBPL
←PRRT1
←AGPAT1
←NOTCH4
←TSBP1
←ATF6B
←AL662884.4
RNF5→
PPT2→
←AGER
PPT2-EGFL8→
←PBX2
EGFL8→
←GPSM3

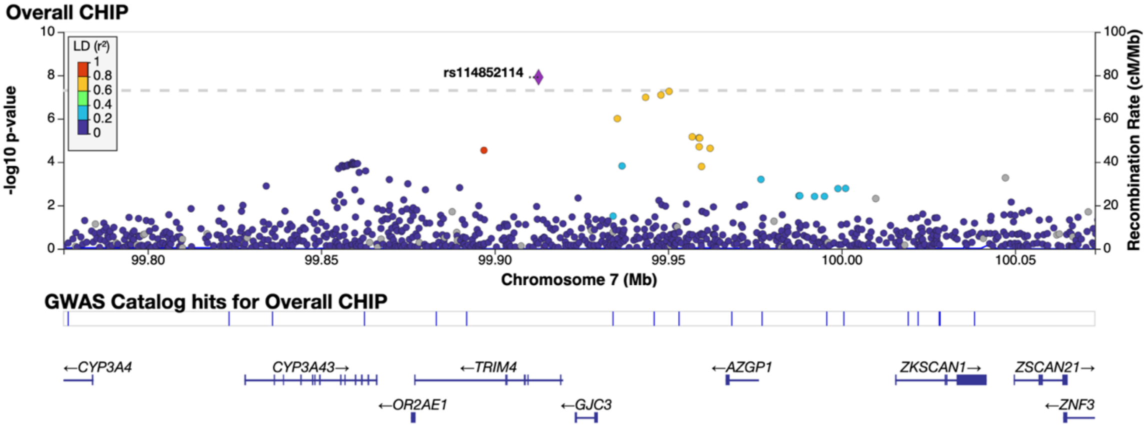

Overall CHIP
LD (r²)
1
0.8
0.6
0.4
0.2
0
rs114852114
-log10 p-value
Recombination Rate (cM/Mb)
Chromosome 7 (Mb)
99.80
99.85
99.90
99.95
100.00
100.05
GWAS Catalog hits for Overall CHIP
←CYP3A4
CYP3A43→
←TRIM4
←AZGP1
ZKSCAN1→
ZSCAN21→
←OR2AE1
←GJC3
←ZNF3

#### Overall CHIP

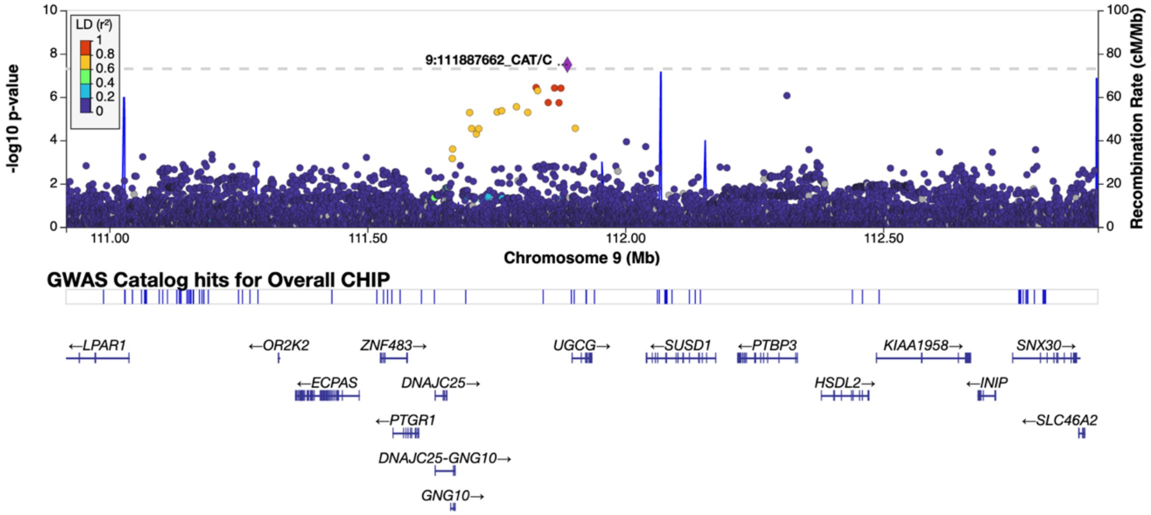

#### CHIP

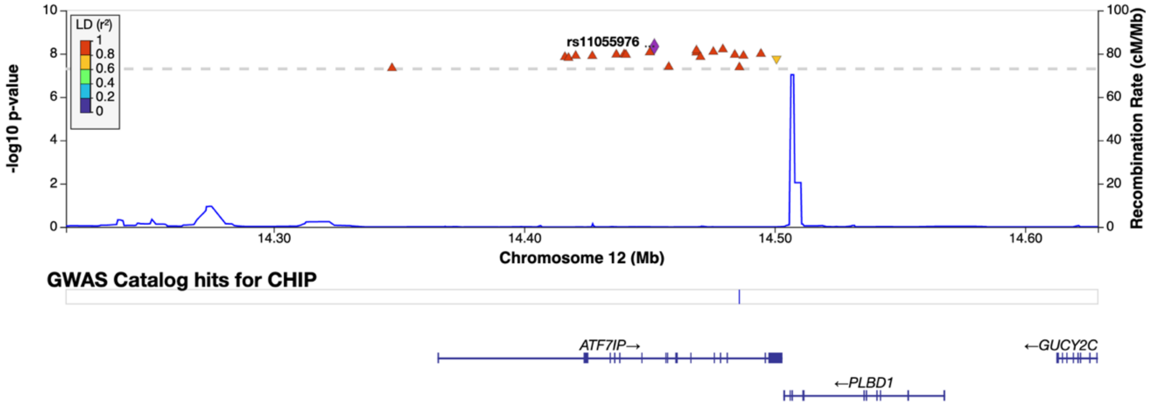

#### CHIP

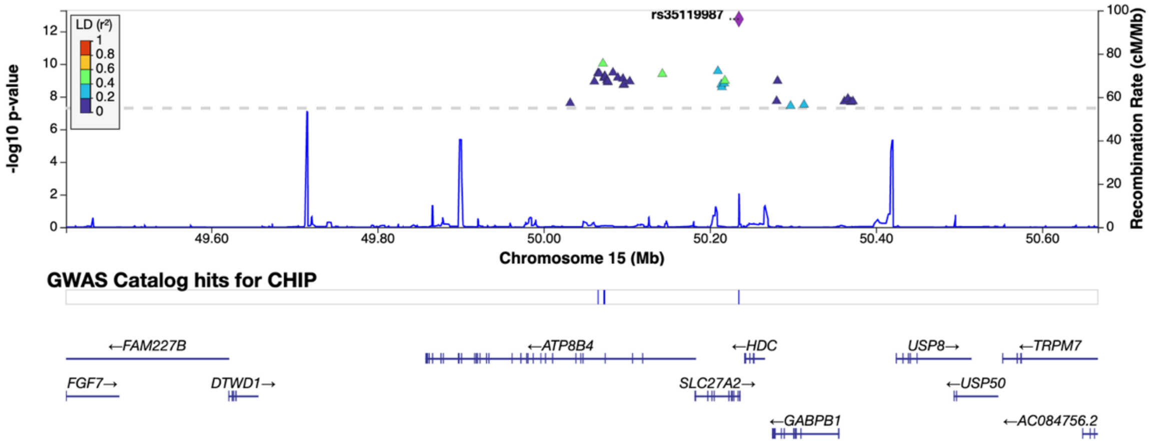

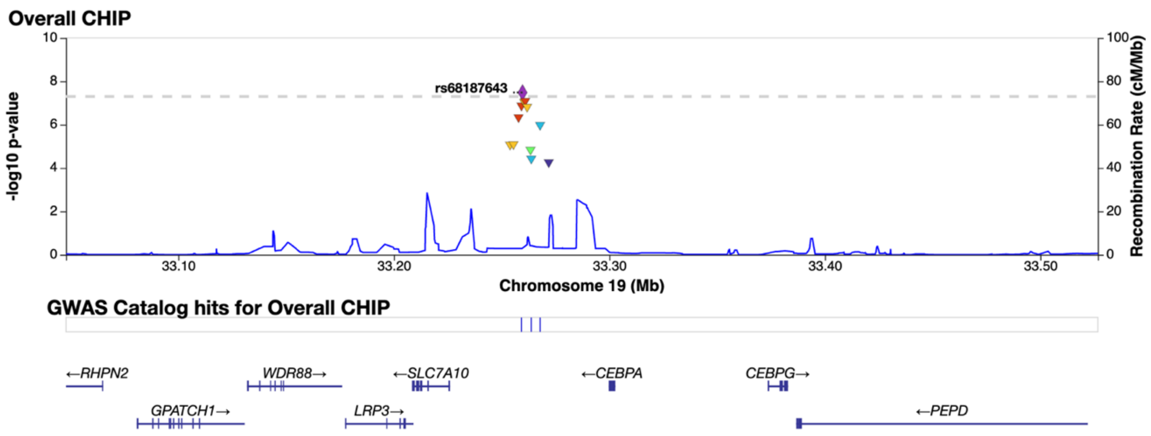
Overall CHIP
-log10 p-value
Recombination Rate (cM/Mb)
rs68187643
Chromosome 19 (Mb)
33.10
33.20
33.30
33.40
33.50
GWAS Catalog hits for Overall CHIP
←RHPN2
WDR88→
←SLC7A10
←CEBPA
CEBPG→
GPATCH1→
LRP3→
←PEPD

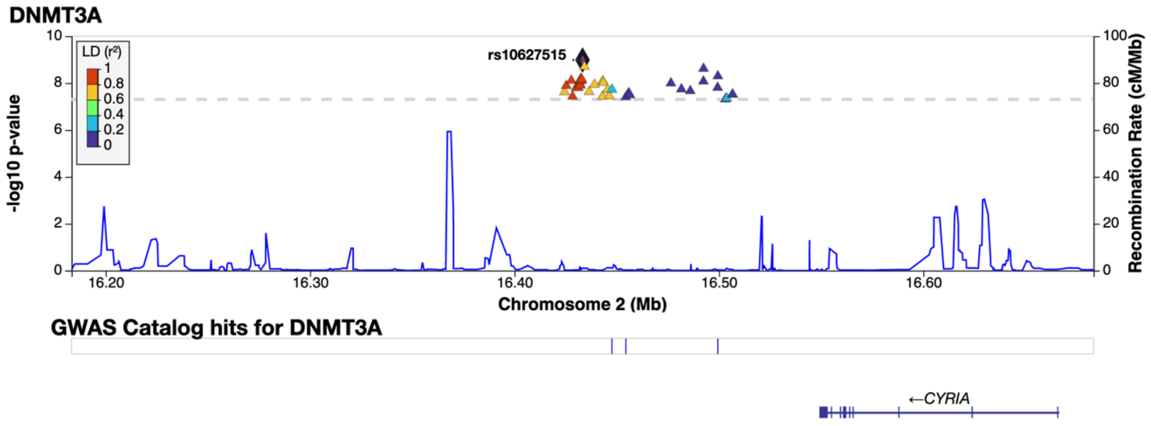

DNMT3A
LD (r²)
rs10627515
-log10 p-value
Recombination Rate (cM/Mb)
16.20
16.30
16.40
16.50
16.60
Chromosome 2 (Mb)
GWAS Catalog hits for DNMT3A
←CYRIA

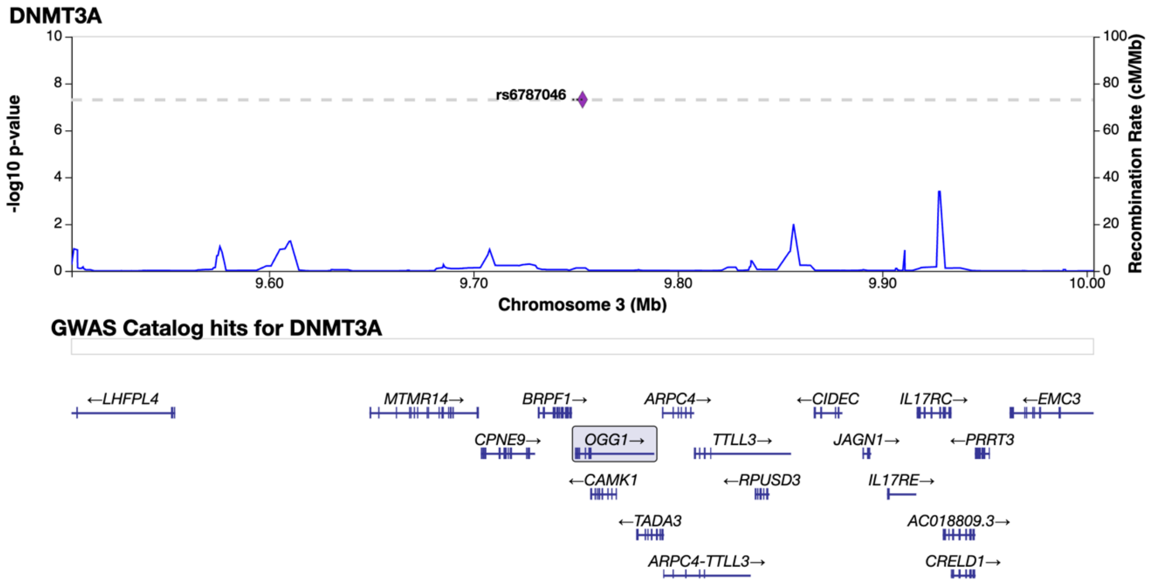

DNMT3A
rs6787046
-log10 p-value
Recombination Rate (cM/Mb)
9.60
9.70
9.80
9.90
10.00
Chromosome 3 (Mb)
GWAS Catalog hits for DNMT3A
←LHFPL4
MTMR14→
BRPF1→
ARPC4→
←CIDEC
IL17RC→
←EMC3
CPNE9→
OGG1→
TTLL3→
JAGN1→
←PRRT3
←CAMK1
←RPUSD3
IL17RE→
←TADA3
AC018809.3→
ARPC4-TTLL3→
CRELD1→

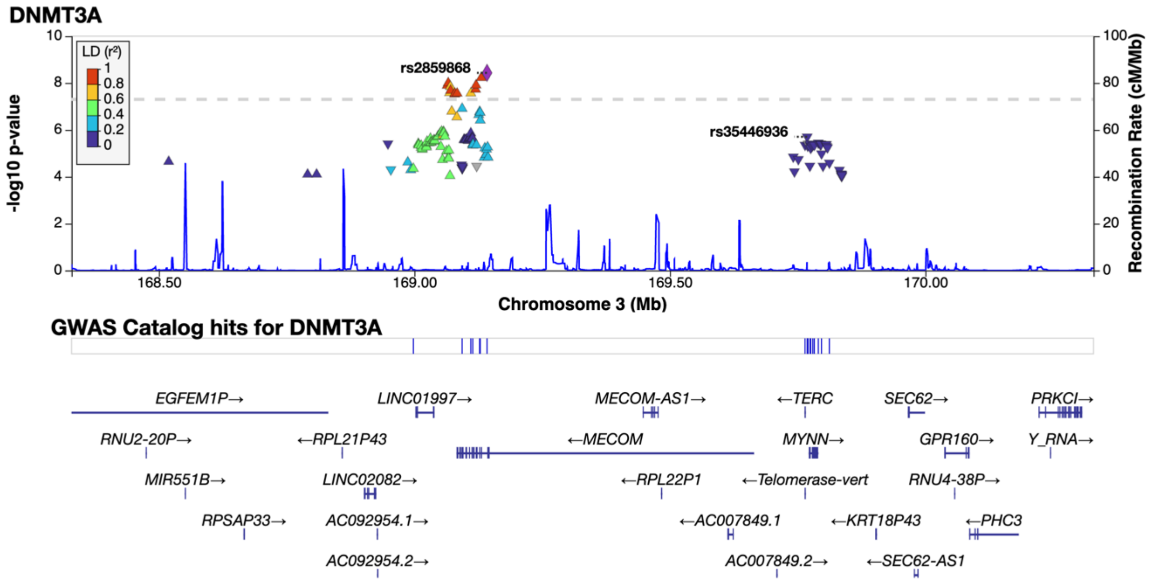

DNMT3A
LD (r²)
rs2859868
rs35446936
-log10 p-value
Recombination Rate (cM/Mb)
168.50
169.00
169.50
170.00
Chromosome 3 (Mb)
GWAS Catalog hits for DNMT3A
EGFEM1P→
LINC01997→
MECOM-AS1→
←TERC
SEC62→
PRKCI→
RNU2-20P→
←RPL21P43
←MECOM
MYNN→
GPR160→
Y_RNA→
MIR551B→
LINC02082→
←RPL22P1
←Telomerase-vert
RNU4-38P→
RPSAP33→
AC092954.1→
←AC007849.1
←KRT18P43
←PHC3
AC092954.2→
AC007849.2→
←SEC62-AS1

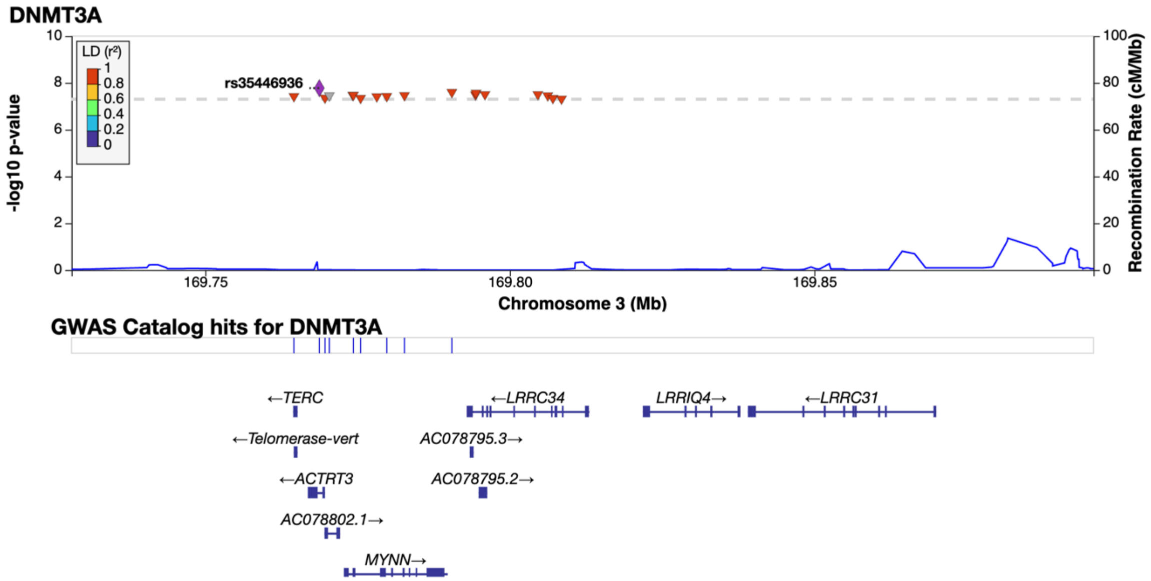
DNMT3A
rs35446936
LD (r²)
-log10 p-value
Recombination Rate (cM/Mb)
Chromosome 3 (Mb)
GWAS Catalog hits for DNMT3A
←TERC
←Telomerase-vert
←ACTRT3
AC078802.1→
MYNN→
←LRRC34
AC078795.3→
AC078795.2→
LRRIQ4→
←LRRC31

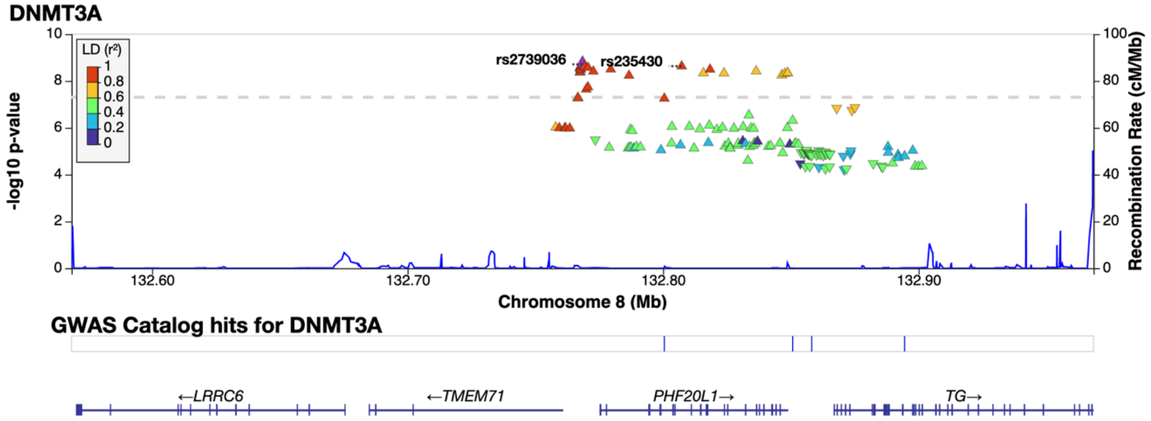
DNMT3A
rs2739036
rs235430
LD (r²)
-log10 p-value
Recombination Rate (cM/Mb)
Chromosome 8 (Mb)
GWAS Catalog hits for DNMT3A
←LRRC6
←TMEM71
PHF20L1→
TG→

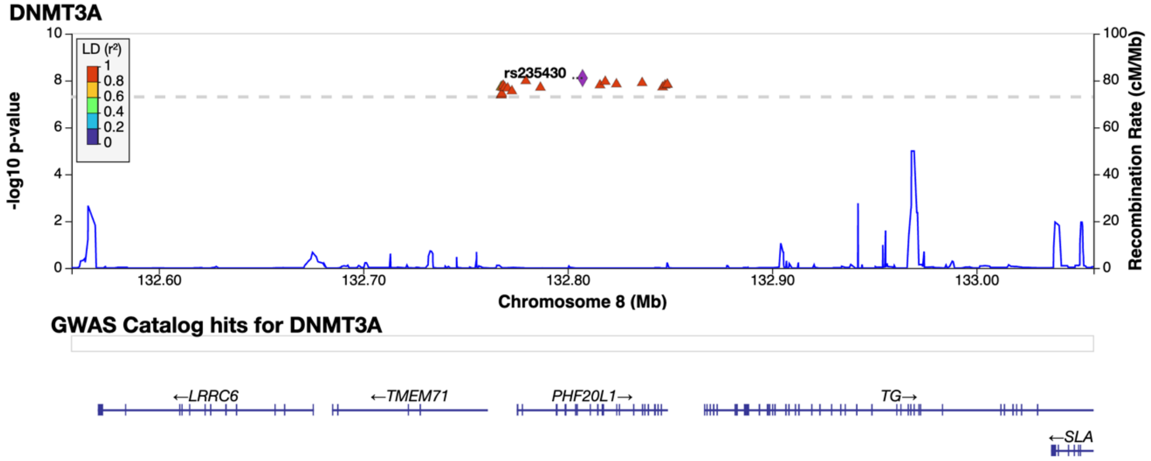
DNMT3A
rs235430
LD (r²)
-log10 p-value
Recombination Rate (cM/Mb)
Chromosome 8 (Mb)
GWAS Catalog hits for DNMT3A
←LRRC6
←TMEM71
PHF20L1→
TG→
←SLA

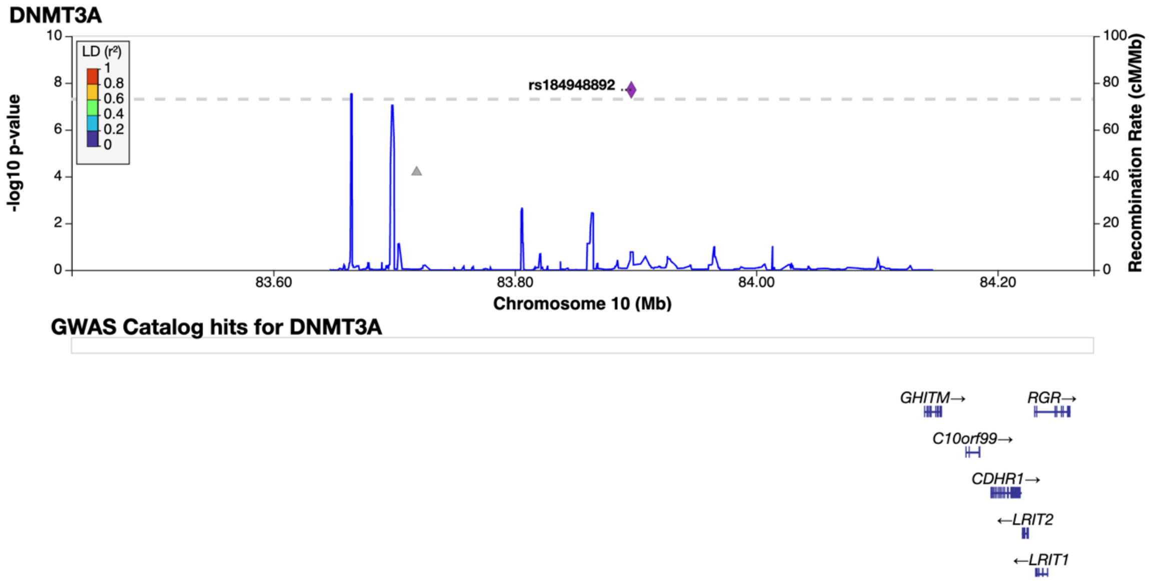
DNMT3A
LD (r²)
rs184948892
-log10 p-value
Recombination Rate (cM/Mb)
Chromosome 10 (Mb)
GWAS Catalog hits for DNMT3A
GHITM→
RGR→
C10orf99→
CDHR1→
←LRIT2
←LRIT1

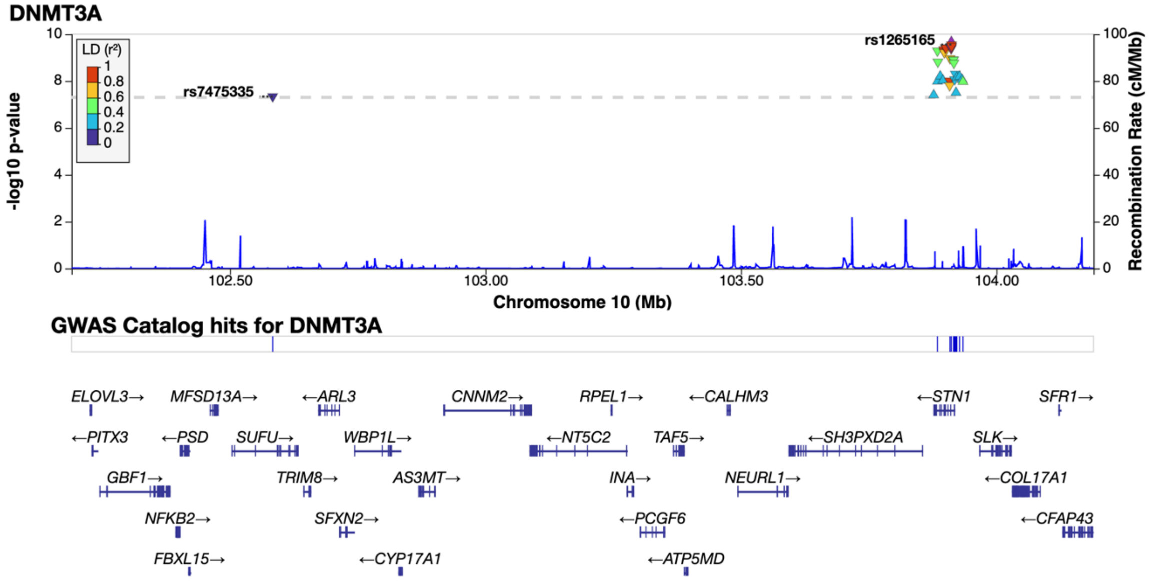
DNMT3A
LD (r²)
rs1265165
rs7475335
-log10 p-value
Recombination Rate (cM/Mb)
Chromosome 10 (Mb)
GWAS Catalog hits for DNMT3A
ELOVL3→ MFSD13A→ ←ARL3 CNNM2→ RPEL1→ ←CALHM3 ←STN1 SFR1→
←PITX3 ←PSD SUFU→ WBP1L→ ←NT5C2 TAF5→ ←SH3PXD2A SLK→
GBF1→ TRIM8→ AS3MT→ INA→ NEURL1→ ←COL17A1
NFKB2→ SFXN2→ ←PCGF6 ←CFAP43
FBXL15→ ←CYP17A1 ←ATP5MD

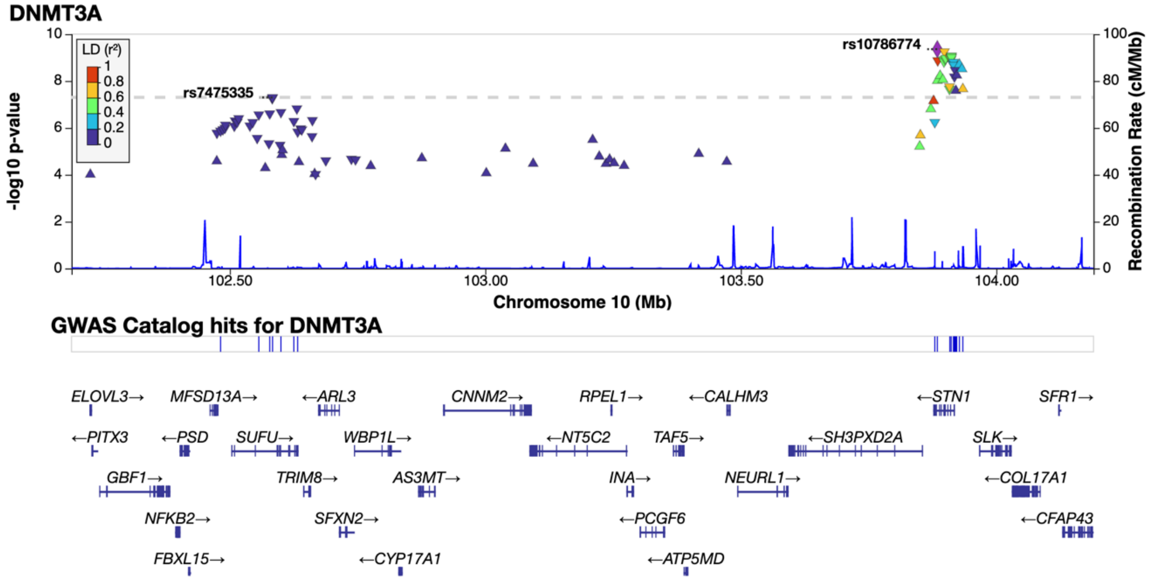
DNMT3A
LD (r²)
rs10786774
rs7475335
-log10 p-value
Recombination Rate (cM/Mb)
Chromosome 10 (Mb)
GWAS Catalog hits for DNMT3A
ELOVL3→ MFSD13A→ ←ARL3 CNNM2→ RPEL1→ ←CALHM3 ←STN1 SFR1→
←PITX3 ←PSD SUFU→ WBP1L→ ←NT5C2 TAF5→ ←SH3PXD2A SLK→
GBF1→ TRIM8→ AS3MT→ INA→ NEURL1→ ←COL17A1
NFKB2→ SFXN2→ ←PCGF6 ←CFAP43
FBXL15→ ←CYP17A1 ←ATP5MD

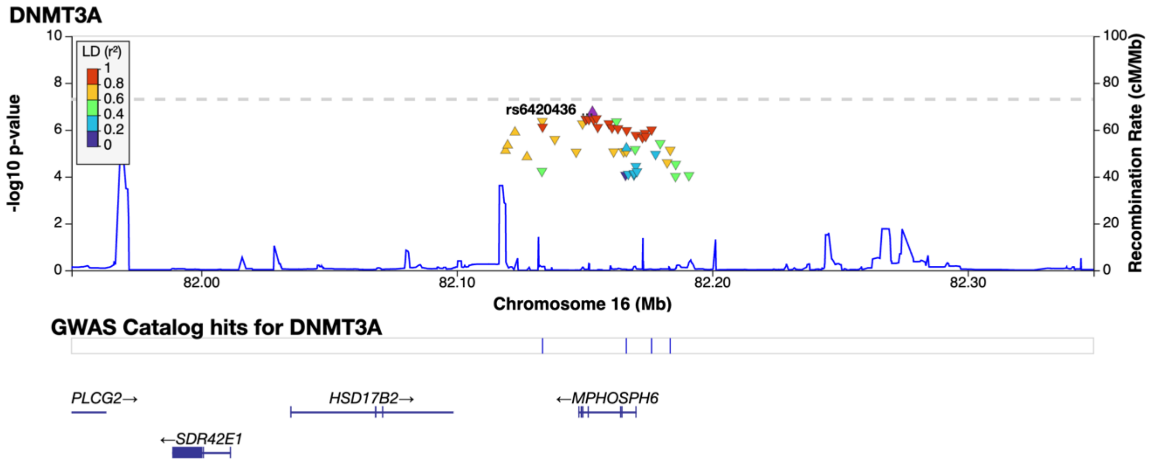
DNMT3A
rs6420436
-log10 p-value
Recombination Rate (cM/Mb)
Chromosome 16 (Mb)
GWAS Catalog hits for DNMT3A
PLCG2→
HSD17B2→
←MPHOSPH6
←SDR42E1

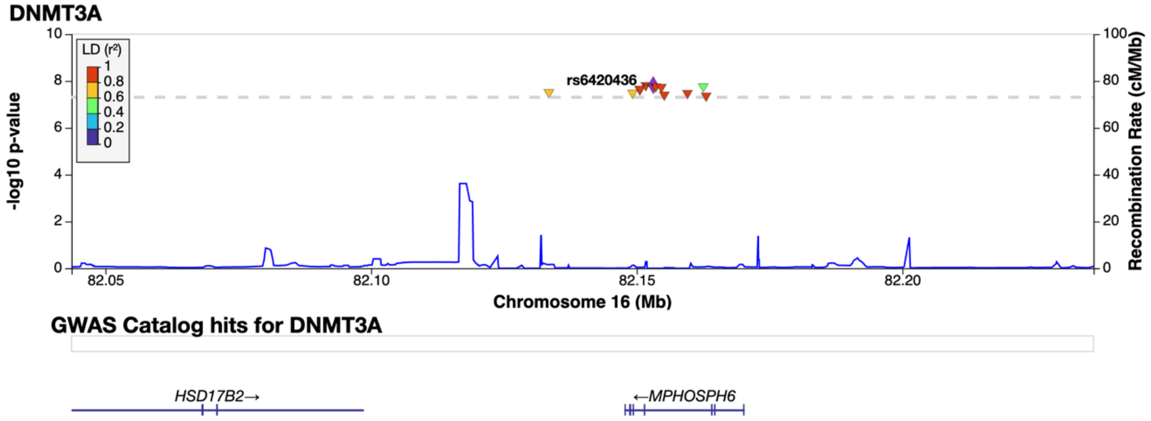
DNMT3A
rs6420436
-log10 p-value
Recombination Rate (cM/Mb)
Chromosome 16 (Mb)
GWAS Catalog hits for DNMT3A
HSD17B2→
←MPHOSPH6

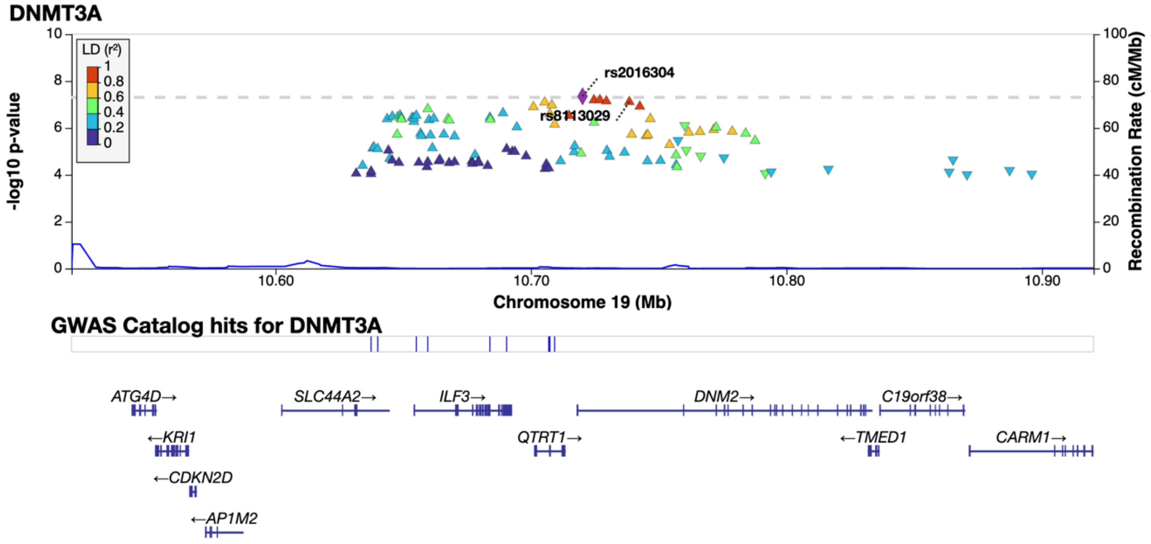
DNMT3A
rs2016304
rs8113029
-log10 p-value
Recombination Rate (cM/Mb)
Chromosome 19 (Mb)
GWAS Catalog hits for DNMT3A
ATG4D→
SLC44A2→
ILF3→
DNM2→
C19orf38→
←KRI1
QTRT1→
←TMED1
CARM1→
←CDKN2D
←AP1M2

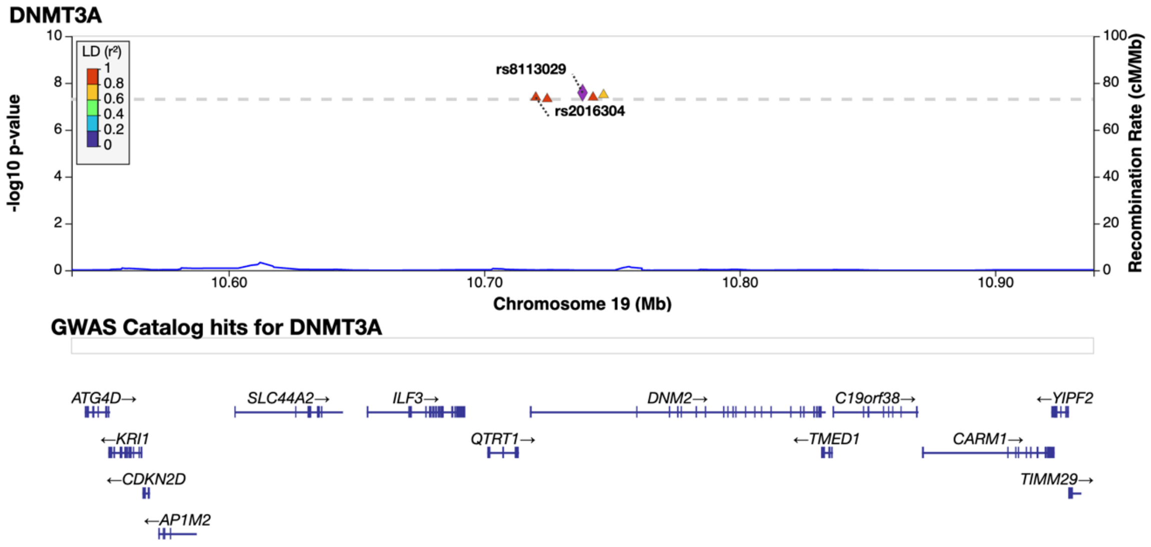
DNMT3A
LD (r²)
1
0.8
0.6
0.4
0.2
0
rs8113029
rs2016304
-log10 p-value
Recombination Rate (cM/Mb)
10.60
10.70
10.80
10.90
Chromosome 19 (Mb)
GWAS Catalog hits for DNMT3A
ATG4D→
SLC44A2→
ILF3→
DNM2→
C19orf38→
←YIPF2
←KRI1
QTRT1→
←TMED1
CARM1→
←CDKN2D
TIMM29→
←AP1M2

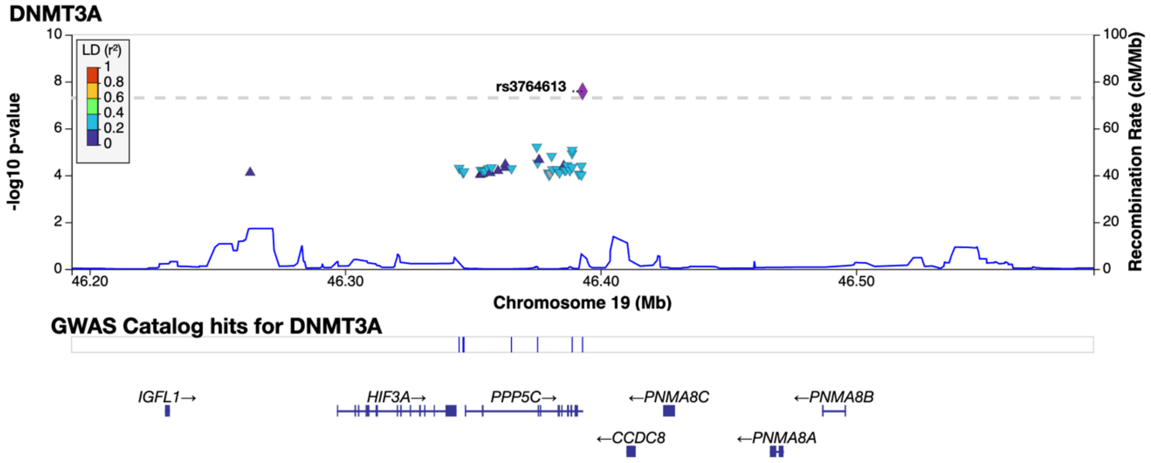
DNMT3A
LD (r²)
1
0.8
0.6
0.4
0.2
0
rs3764613
-log10 p-value
Recombination Rate (cM/Mb)
46.20
46.30
46.40
46.50
Chromosome 19 (Mb)
GWAS Catalog hits for DNMT3A
IGFL1→
HIF3A→
PPP5C→
←PNMA8C
←PNMA8B
←CCDC8
←PNMA8A

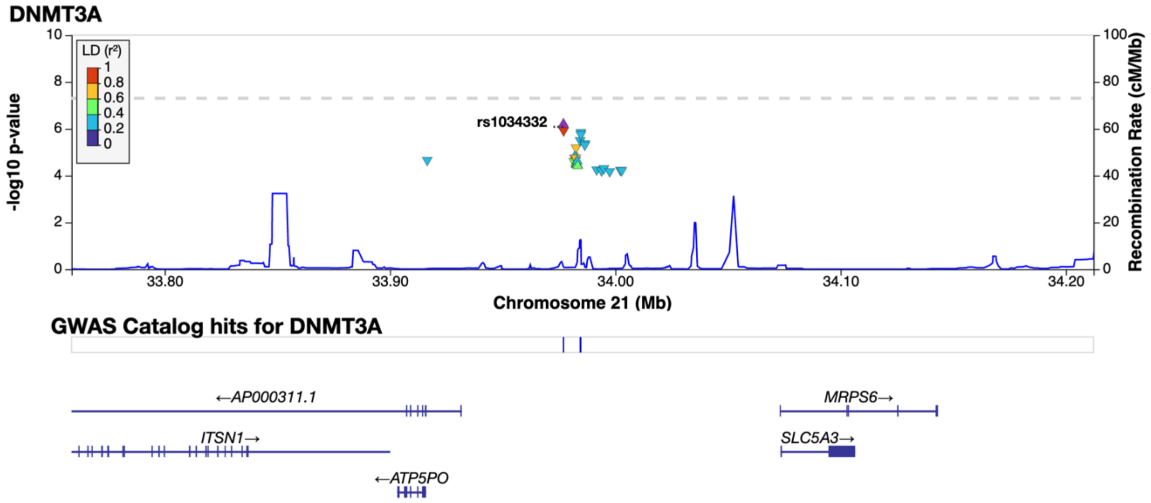
DNMT3A
LD (r²)
1
0.8
0.6
0.4
0.2
0
rs1034332
-log10 p-value
Recombination Rate (cM/Mb)
33.80
33.90
34.00
34.10
34.20
Chromosome 21 (Mb)
GWAS Catalog hits for DNMT3A
←AP000311.1
MRPS6→
ITSN1→
SLC5A3→
←ATP5PO

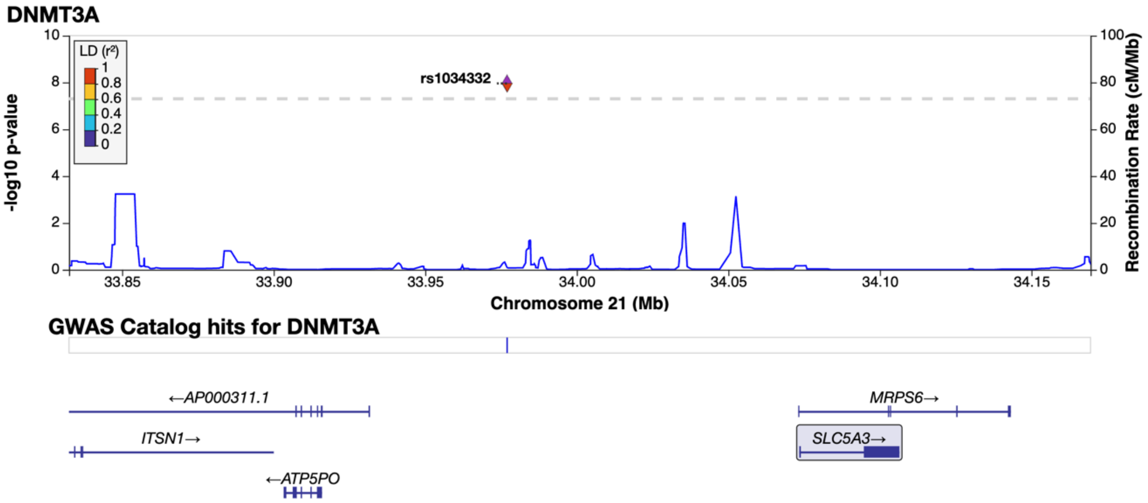
DNMT3A
LD (r²)
1
0.8
0.6
0.4
0.2
0
rs1034332
-log10 p-value
Recombination Rate (cM/Mb)
Chromosome 21 (Mb)
33.85
33.90
33.95
34.00
34.05
34.10
34.15
GWAS Catalog hits for DNMT3A
←AP000311.1
ITSN1→
←ATP5PO
MRPS6→
SLC5A3→

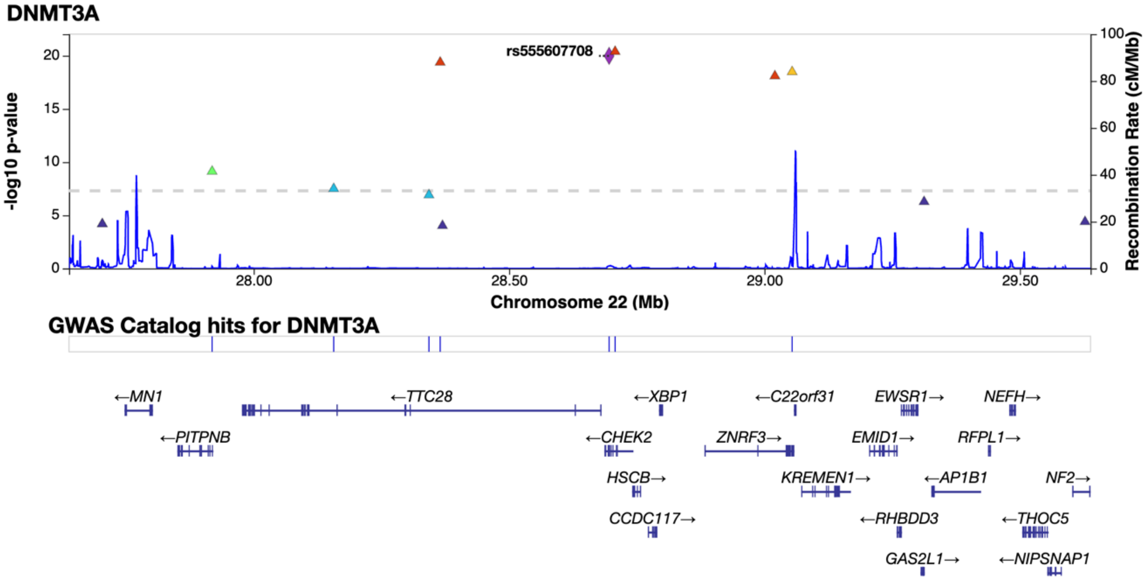
DNMT3A
rs555607708
-log10 p-value
Recombination Rate (cM/Mb)
Chromosome 22 (Mb)
28.00
28.50
29.00
29.50
GWAS Catalog hits for DNMT3A
←MN1
←PITPNB
←TTC28
←XBP1
←CHEK2
HSCB→
CCDC117→
←C22orf31
ZNRF3→
KREMEN1→
EWSR1→
EMID1→
←RHBDD3
GAS2L1→
NEFH→
RFPL1→
←AP1B1
NF2→
←THOC5
←NIPSNAP1

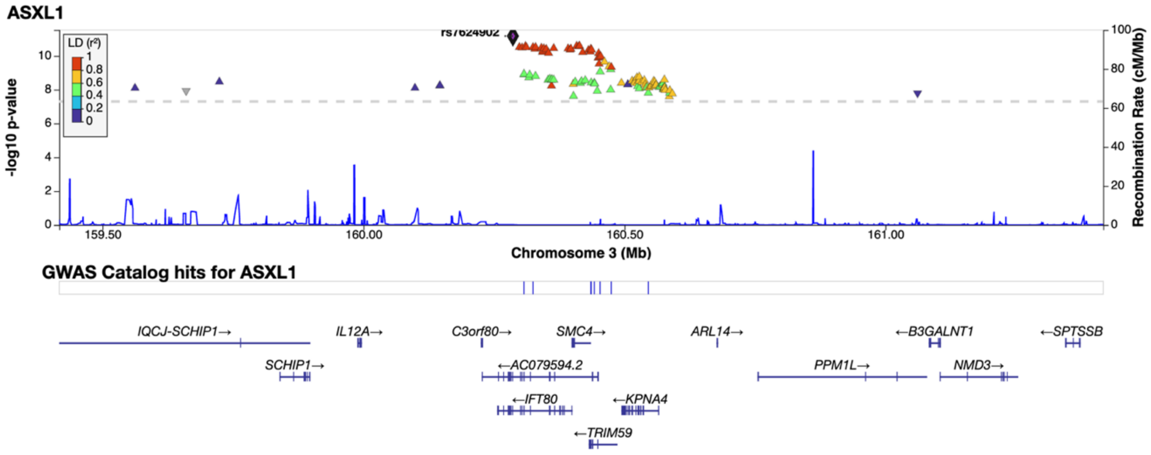
ASXL1
rs7624902
LD (r²)
-log10 p-value
Recombination Rate (cM/Mb)
Chromosome 3 (Mb)
GWAS Catalog hits for ASXL1
IQCJ-SCHIP1→
IL12A→
C3orf80→
SMC4→
ARL14→
←B3GALNT1
←SPTSSB
SCHIP1→
←AC079594.2
PPM1L→
NMD3→
←IFT80
←KPNA4
←TRIM59

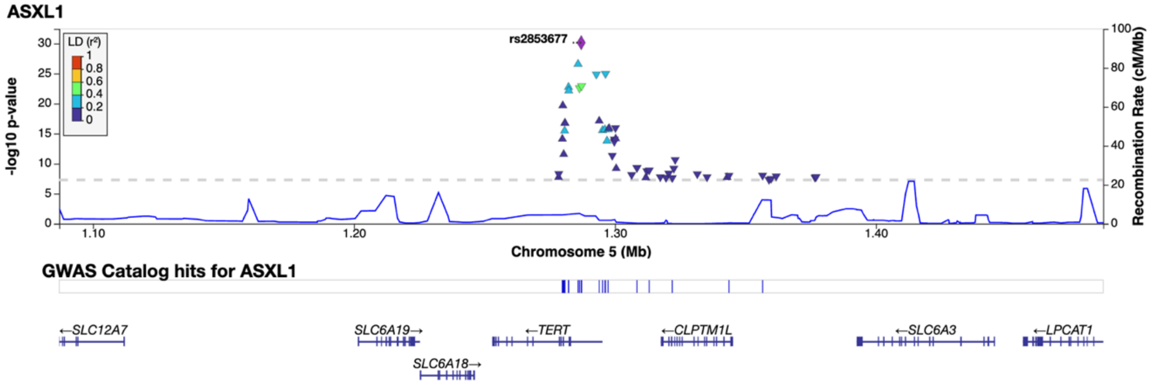
ASXL1
rs2853677
LD (r²)
-log10 p-value
Recombination Rate (cM/Mb)
Chromosome 5 (Mb)
GWAS Catalog hits for ASXL1
←SLC12A7
SLC6A19→
←TERT
←CLPTM1L
←SLC6A3
←LPCAT1
SLC6A18→

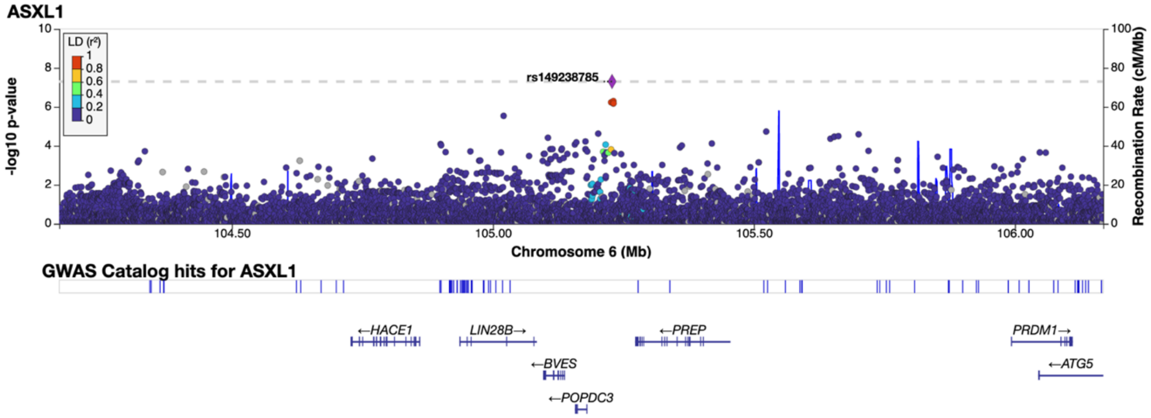
ASXL1
rs149238785
LD (r²)
-log10 p-value
Recombination Rate (cM/Mb)
Chromosome 6 (Mb)
GWAS Catalog hits for ASXL1
←HACE1
LIN28B→
←PREP
PRDM1→
←BVES
←ATG5
←POPDC3

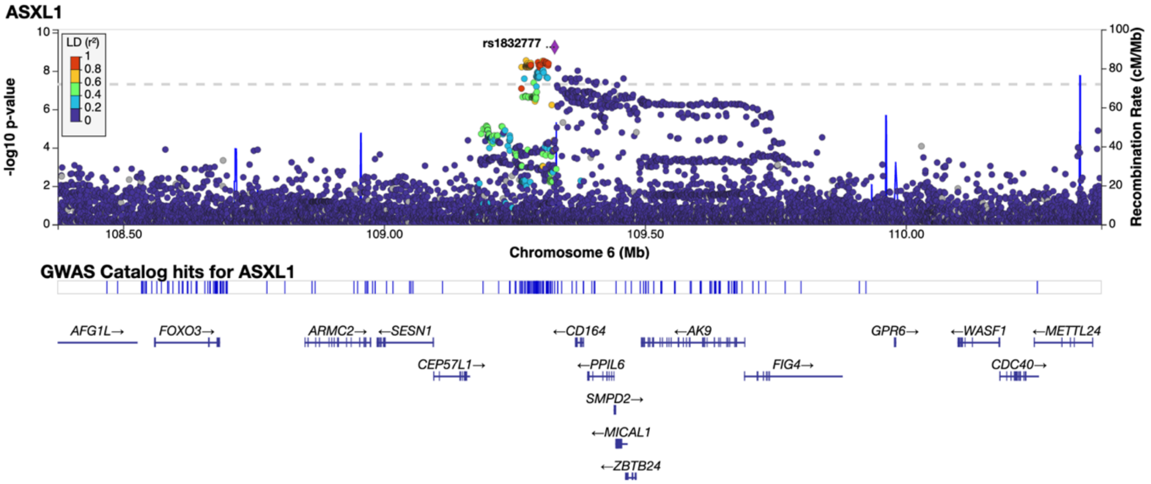
ASXL1
-log10 p-value
LD (r²)
1
0.8
0.6
0.4
0.2
0
rs1832777
Recombination Rate (cM/Mb)
108.50
109.00
109.50
110.00
Chromosome 6 (Mb)
GWAS Catalog hits for ASXL1
AFG1L→
FOXO3→
ARMC2→
←SESN1
CEP57L1→
←CD164
←PPIL6
SMPD2→
←MICAL1
←ZBTB24
←AK9
FIG4→
GPR6→
←WASF1
CDC40→
←METTL24

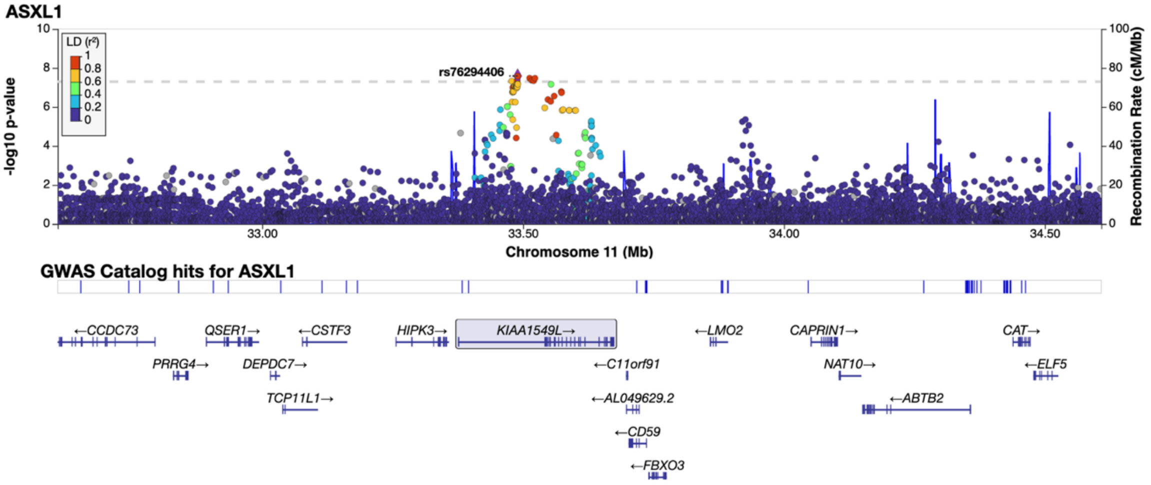
ASXL1
-log10 p-value
LD (r²)
1
0.8
0.6
0.4
0.2
0
rs76294406
Recombination Rate (cM/Mb)
33.00
33.50
34.00
34.50
Chromosome 11 (Mb)
GWAS Catalog hits for ASXL1
←CCDC73
PRRG4→
QSER1→
DEPDC7→
TCP11L1→
←CSTF3
HIPK3→
KIAA1549L→
←C11orf91
←AL049629.2
←CD59
←FBXO3
←LMO2
CAPRIN1→
NAT10→
←ABTB2
CAT→
←ELF5

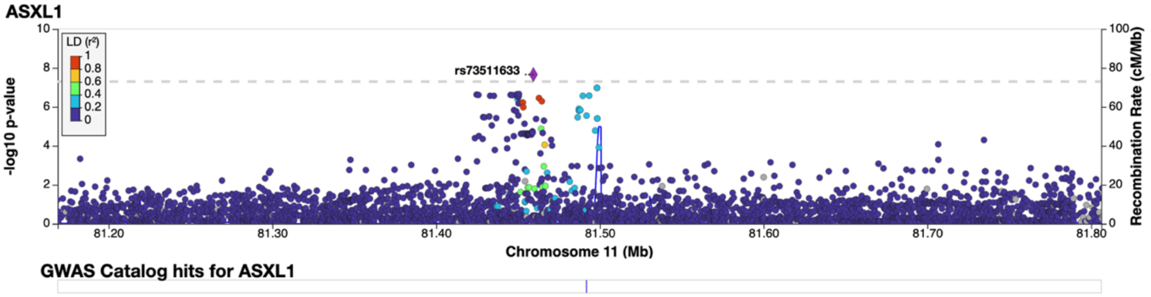
ASXL1
-log10 p-value
LD (r²)
1
0.8
0.6
0.4
0.2
0
rs73511633
Recombination Rate (cM/Mb)
81.20
81.30
81.40
81.50
81.60
81.70
81.80
Chromosome 11 (Mb)
GWAS Catalog hits for ASXL1

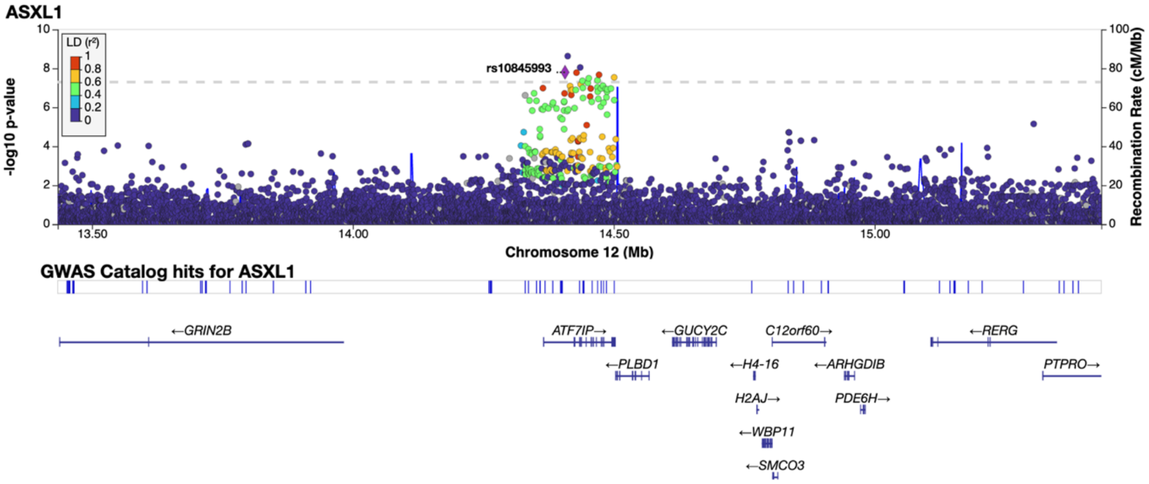
ASXL1
LD (r²)
1
0.8
0.6
0.4
0.2
0
rs10845993
-log10 p-value
Recombination Rate (cM/Mb)
Chromosome 12 (Mb)
13.50
14.00
14.50
15.00
GWAS Catalog hits for ASXL1
←GRIN2B
ATF7IP→
←GUCY2C
C12orf60→
←RERG
←PLBD1
←H4-16
←ARHGDIB
PTPRO→
H2AJ→
PDE6H→
←WBP11
←SMCO3

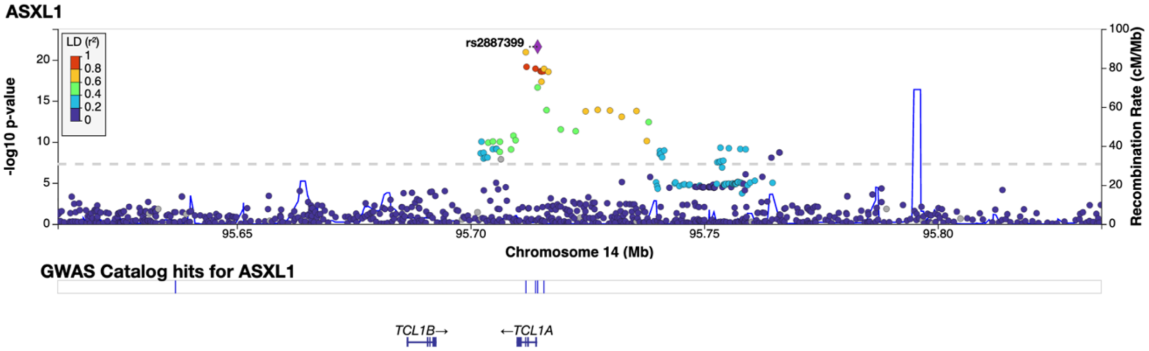
ASXL1
LD (r²)
1
0.8
0.6
0.4
0.2
0
rs2887399
-log10 p-value
Recombination Rate (cM/Mb)
Chromosome 14 (Mb)
95.65
95.70
95.75
95.80
GWAS Catalog hits for ASXL1
TCL1B→
←TCL1A

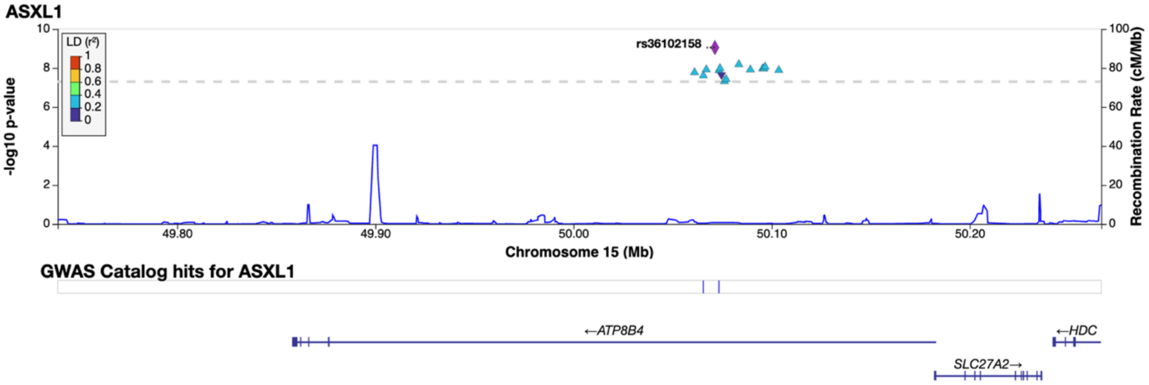
ASXL1
LD (r²)
1
0.8
0.6
0.4
0.2
0
rs36102158
-log10 p-value
Recombination Rate (cM/Mb)
Chromosome 15 (Mb)
49.80
49.90
50.00
50.10
50.20
GWAS Catalog hits for ASXL1
←ATP8B4
←HDC
SLC27A2→

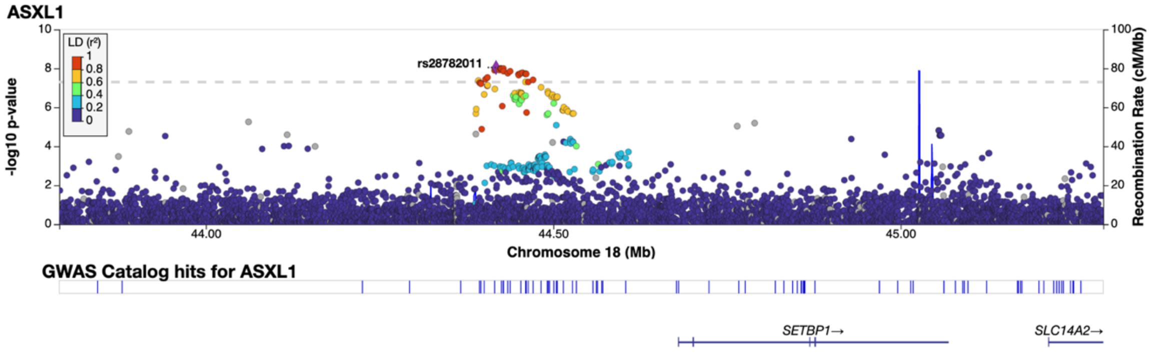

**Extended Data Figure 11 | Locus plots for genomic regions containing new significant signals for CHIP traits in multi-ancestry GWAS meta-analysis.** CHIP: clonal hematopoiesis of indeterminate potential.

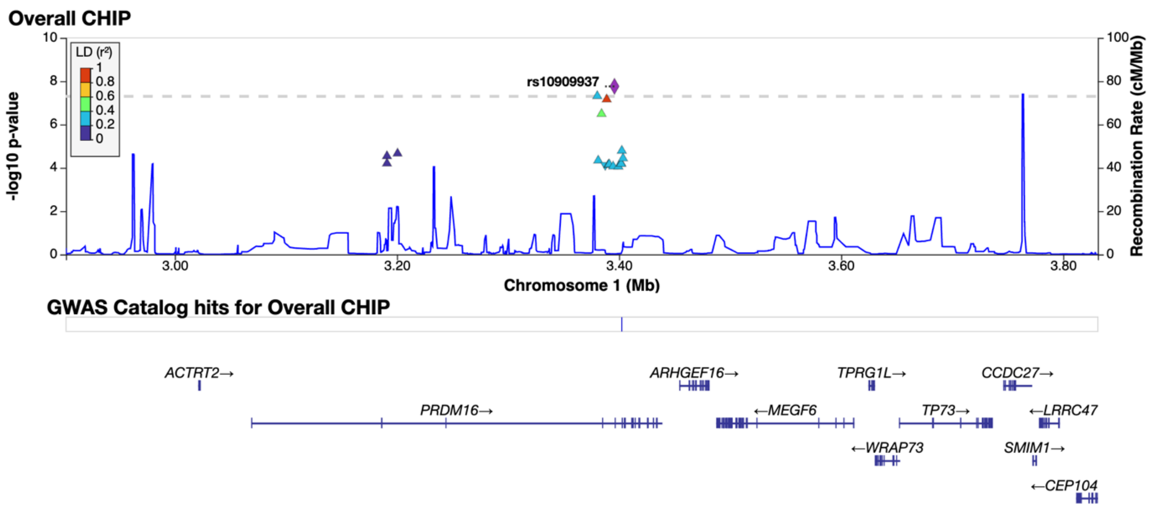
Overall CHIP
LD (r²)
rs10909937
-log10 p-value
Recombination Rate (cM/Mb)
Chromosome 1 (Mb)
GWAS Catalog hits for Overall CHIP
ACTRT2→
PRDM16→
ARHGEF16→
←MEGF6
TPRG1L→
←WRAP73
TP73→
CCDC27→
←LRRC47
SMIM1→
←CEP104

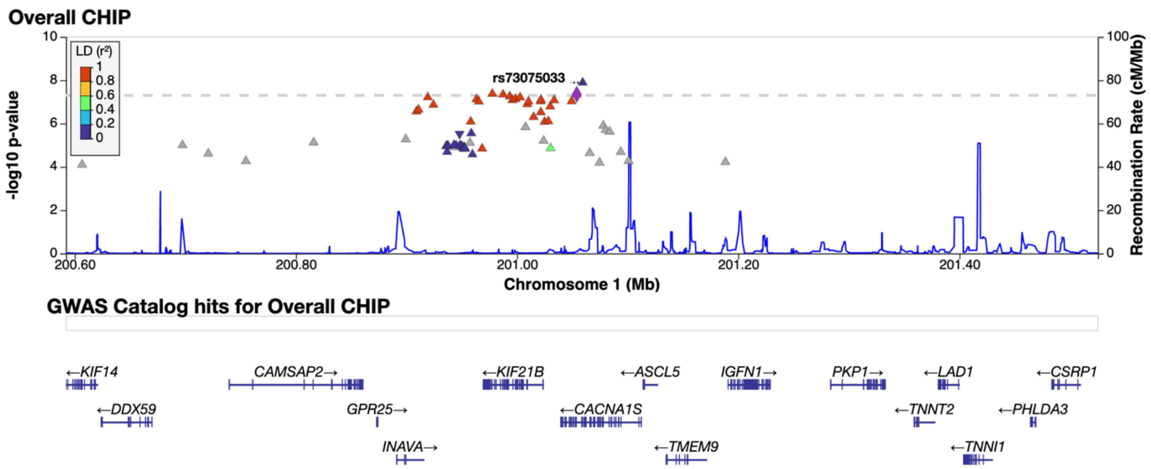
Overall CHIP
LD (r²)
rs73075033
-log10 p-value
Recombination Rate (cM/Mb)
Chromosome 1 (Mb)
GWAS Catalog hits for Overall CHIP
←KIF14
←DDX59
CAMSAP2→
GPR25→
INAVA→
←KIF21B
←CACNA1S
←ASCL5
←TMEM9
IGFN1→
PKP1→
←TNNT2
←LAD1
←TNNI1
←PHLDA3
←CSRP1

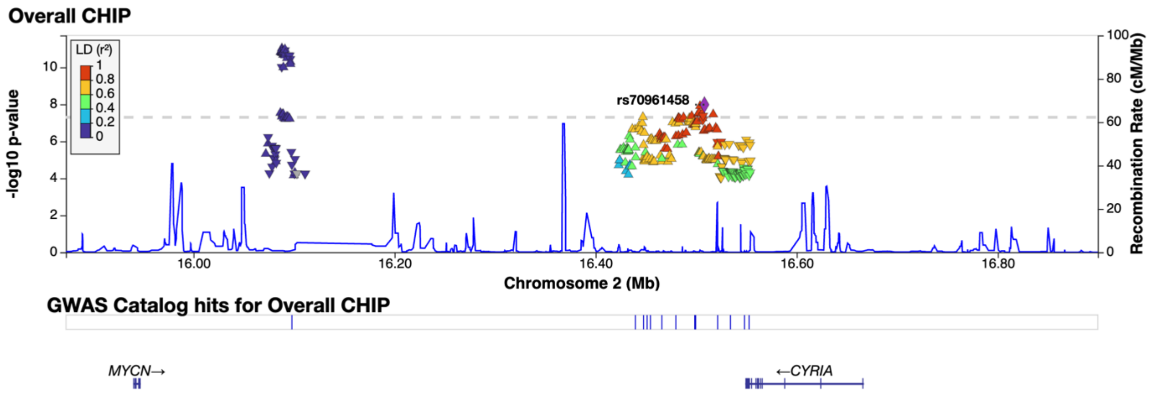
Overall CHIP
LD (r²)
rs70961458
-log10 p-value
Recombination Rate (cM/Mb)
Chromosome 2 (Mb)
GWAS Catalog hits for Overall CHIP
MYCN→
←CYRIA

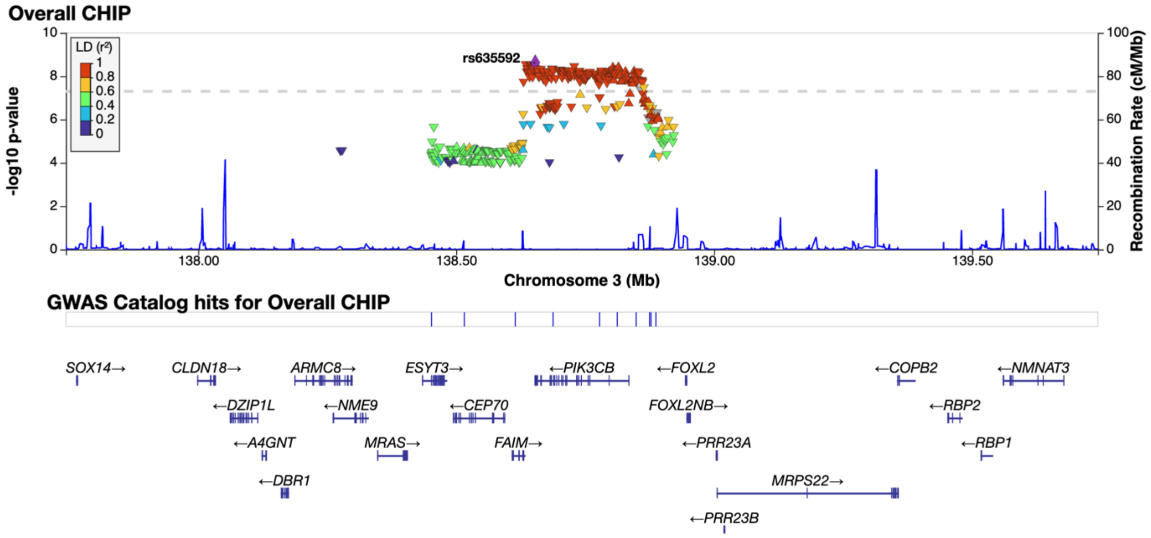
Overall CHIP
rs635592
LD (r²)
1
0.8
0.6
0.4
0.2
0
-log10 p-value
Recombination Rate (cM/Mb)
Chromosome 3 (Mb)
138.00
138.50
139.00
139.50
GWAS Catalog hits for Overall CHIP
SOX14→
CLDN18→
ARMC8→
ESYT3→
←PIK3CB
←FOXL2
←COPB2
←NMNAT3
←DZIP1L
←NME9
←CEP70
FOXL2NB→
←RBP2
←A4GNT
MRAS→
FAIM→
←PRR23A
←RBP1
←DBR1
MRPS22→
←PRR23B

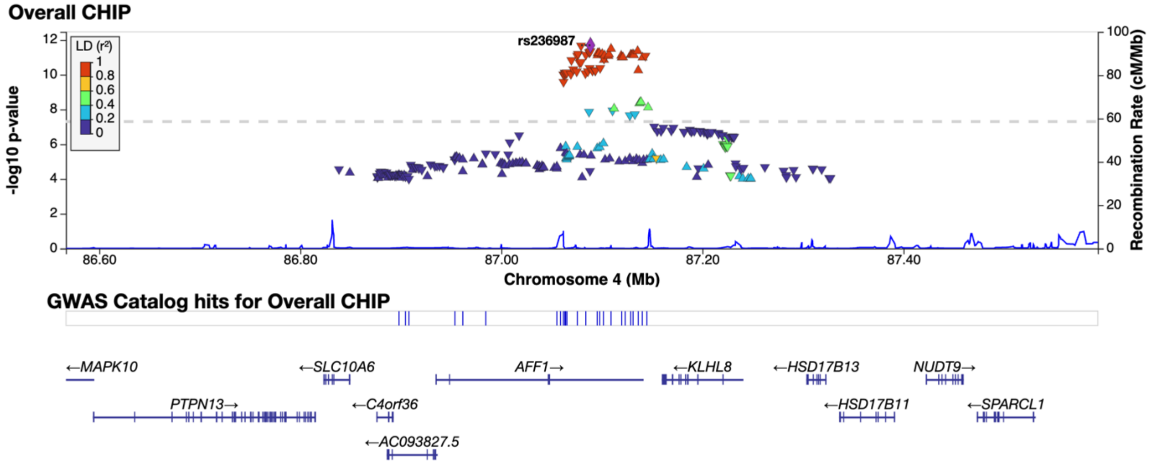
Overall CHIP
rs236987
LD (r²)
1
0.8
0.6
0.4
0.2
0
-log10 p-value
Recombination Rate (cM/Mb)
Chromosome 4 (Mb)
86.60
86.80
87.00
87.20
87.40
GWAS Catalog hits for Overall CHIP
←MAPK10
←SLC10A6
AFF1→
←KLHL8
←HSD17B13
NUDT9→
PTPN13→
←C4orf36
←HSD17B11
←SPARCL1
←AC093827.5

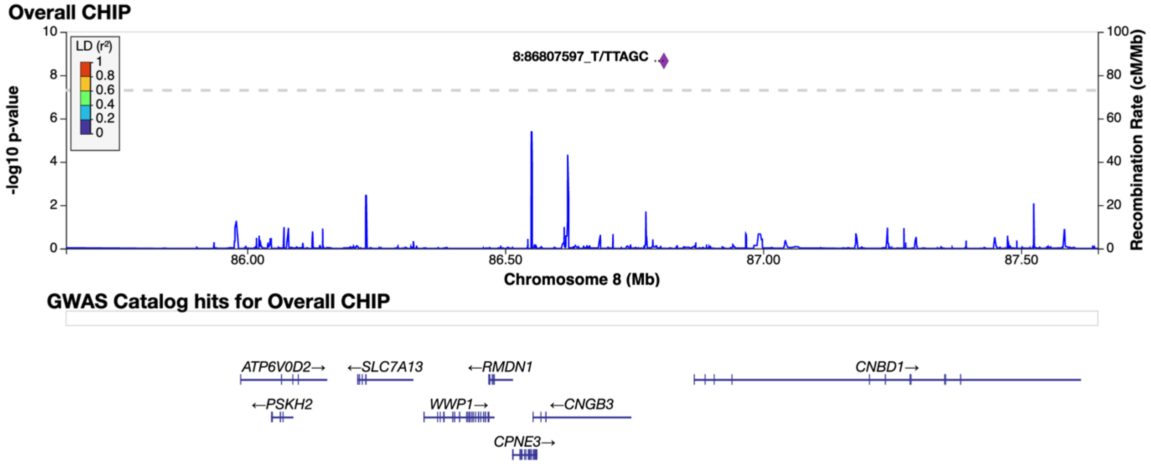
Overall CHIP
8:86807597_T/TTAGC
LD (r²)
1
0.8
0.6
0.4
0.2
0
-log10 p-value
Recombination Rate (cM/Mb)
Chromosome 8 (Mb)
86.00
86.50
87.00
87.50
GWAS Catalog hits for Overall CHIP
ATP6V0D2→
←SLC7A13
←RMDN1
CNBD1→
←PSKH2
WWP1→
←CNGB3
CPNE3→

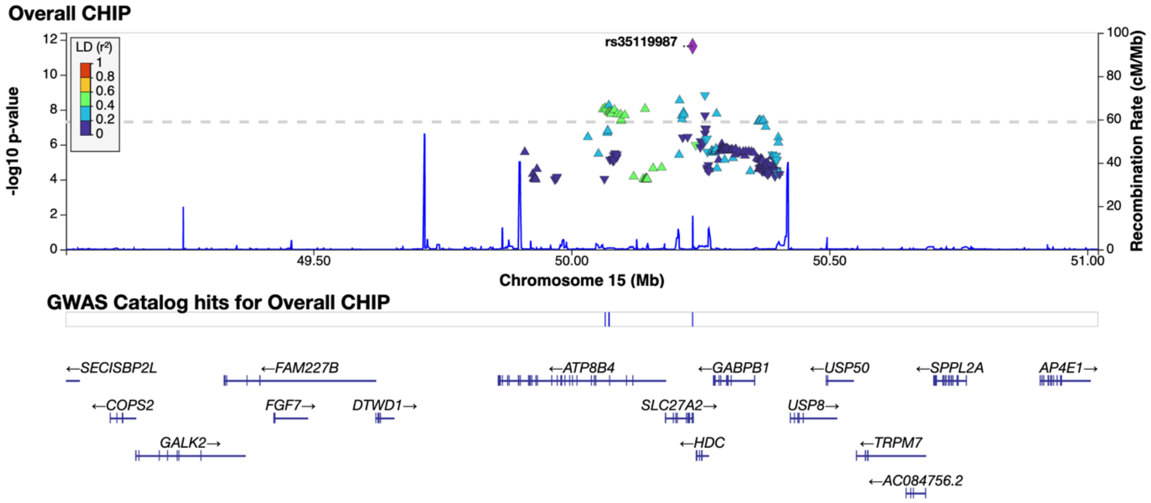

**Extended Data Figure 12 | Locus plots for genomic regions containing new significant signals for overall CHIP in European ancestry-stratified GWAS meta-analysis.** CHIP: clonal hematopoiesis of indeterminate potential.

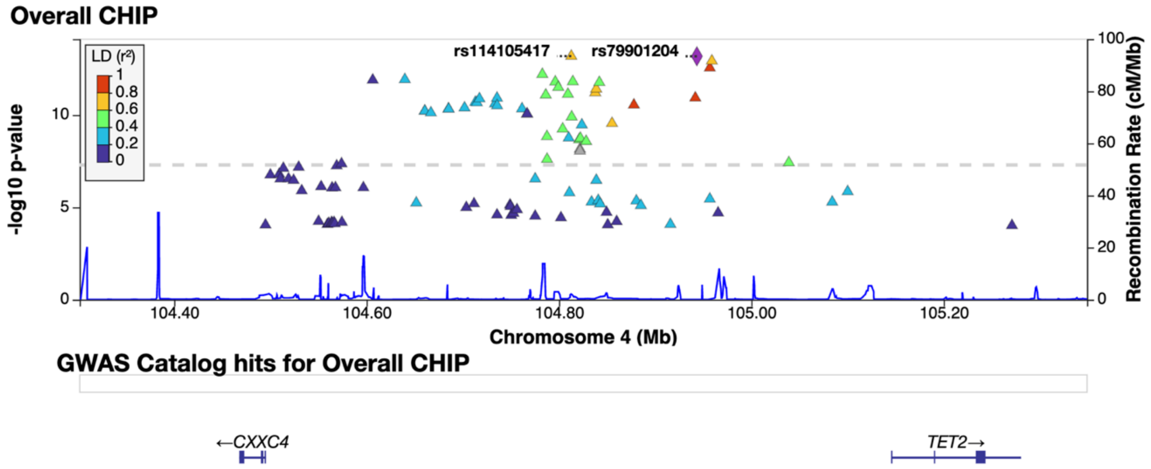
Overall CHIP
rs114105417
rs79901204
LD (r²)
1
0.8
0.6
0.4
0.2
0
-log10 p-value
Recombination Rate (cM/Mb)
Chromosome 4 (Mb)
104.40
104.60
104.80
105.00
105.20
GWAS Catalog hits for Overall CHIP
←CXXC4
TET2→

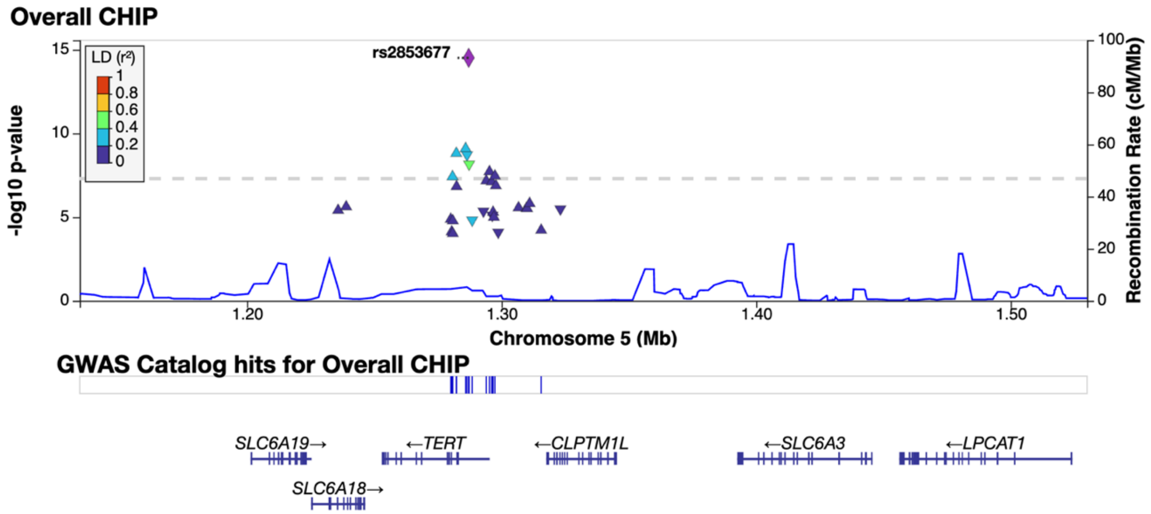
Overall CHIP
rs2853677
LD (r²)
-log10 p-value
Recombination Rate (cM/Mb)
Chromosome 5 (Mb)
1.20
1.30
1.40
1.50
GWAS Catalog hits for Overall CHIP
SLC6A19→
←TERT
←CLPTM1L
←SLC6A3
←LPCAT1
SLC6A18→

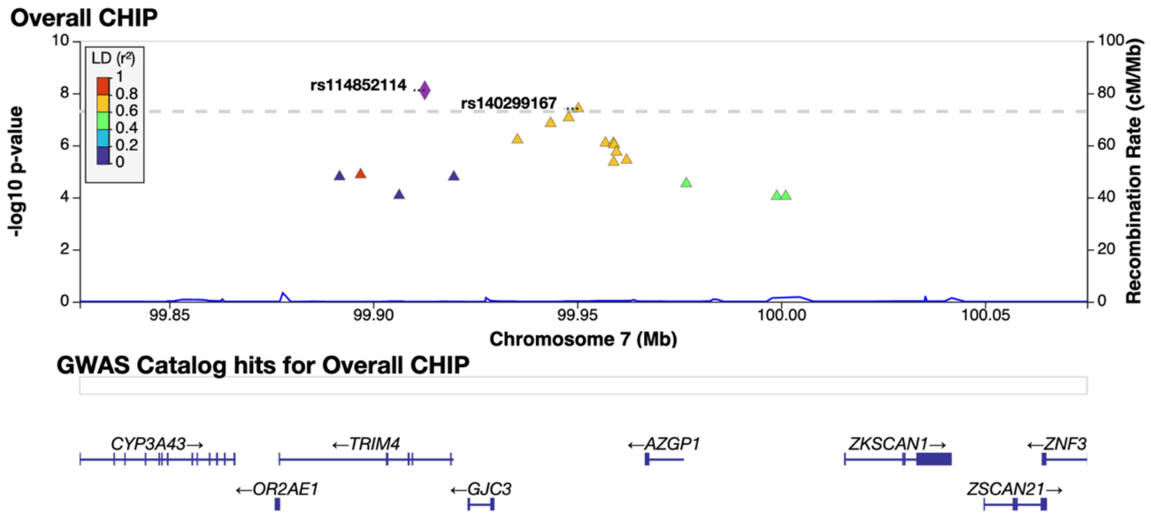
Overall CHIP
rs114852114
rs140299167
LD (r²)
-log10 p-value
Recombination Rate (cM/Mb)
Chromosome 7 (Mb)
99.85
99.90
99.95
100.00
100.05
GWAS Catalog hits for Overall CHIP
CYP3A43→
←TRIM4
←AZGP1
ZKSCAN1→
←ZNF3
←OR2AE1
←GJC3
ZSCAN21→

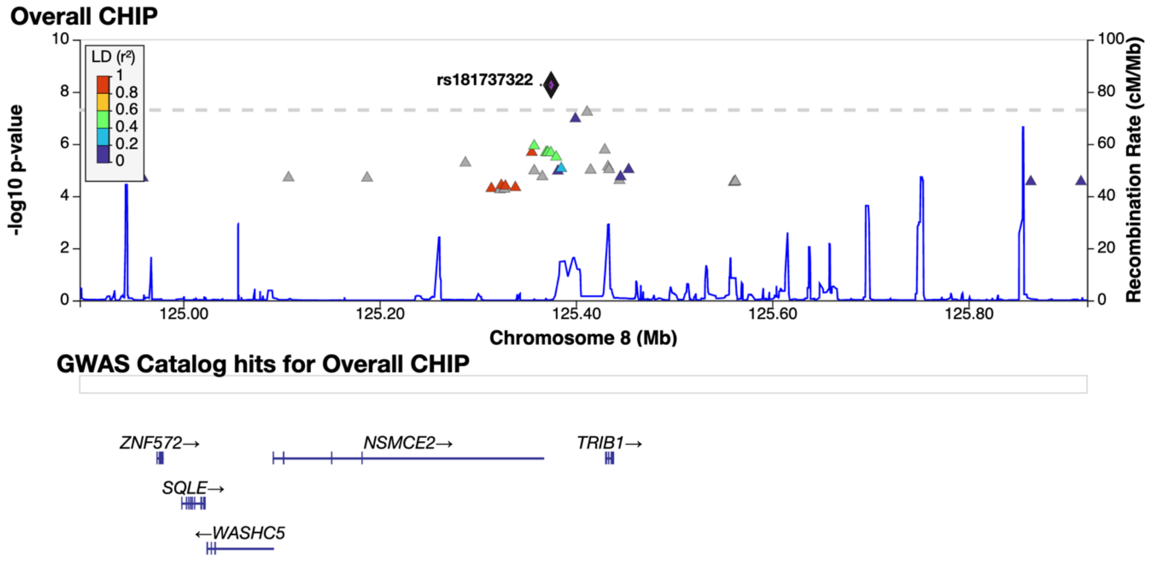
Overall CHIP
LD (r²)
1
0.8
0.6
0.4
0.2
0
rs181737322
-log10 p-value
Recombination Rate (cM/Mb)
125.00
125.20
125.40
125.60
125.80
Chromosome 8 (Mb)
GWAS Catalog hits for Overall CHIP
ZNF572→
NSMCE2→
TRIB1→
SQLE→
←WASHC5

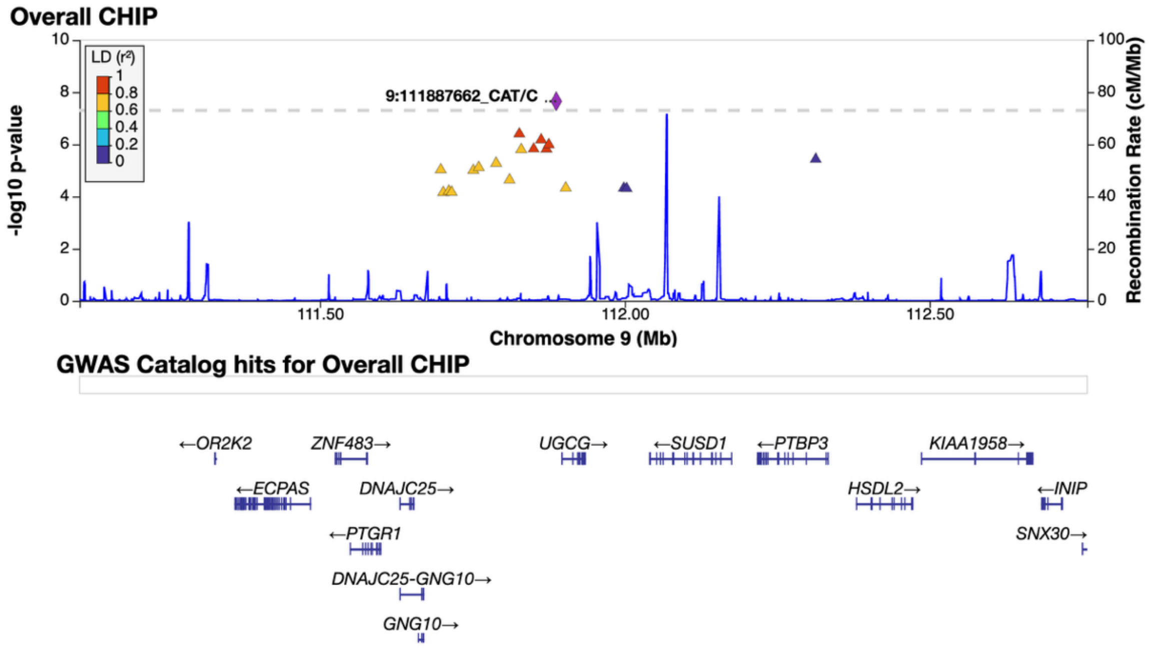
Overall CHIP
LD (r²)
1
0.8
0.6
0.4
0.2
0
9:111887662_CAT/C
-log10 p-value
Recombination Rate (cM/Mb)
111.50
112.00
112.50
Chromosome 9 (Mb)
GWAS Catalog hits for Overall CHIP
←OR2K2
ZNF483→
UGCG→
←SUSD1
←PTBP3
KIAA1958→
←ECPAS
DNAJC25→
HSDL2→
←INIP
←PTGR1
SNX30→
DNAJC25-GNG10→
GNG10→

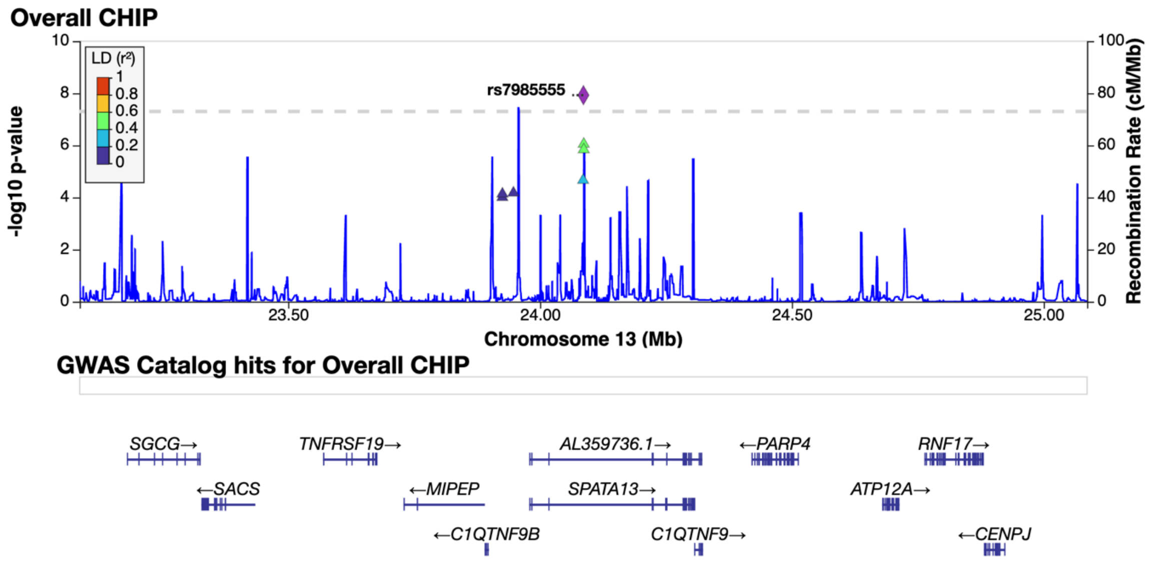
Overall CHIP
LD (r²)
1
0.8
0.6
0.4
0.2
0
rs7985555
-log10 p-value
Recombination Rate (cM/Mb)
23.50
24.00
24.50
25.00
Chromosome 13 (Mb)
GWAS Catalog hits for Overall CHIP
SGCG→
TNFRSF19→
AL359736.1→
←PARP4
RNF17→
←SACS
←MIPEP
SPATA13→
ATP12A→
←C1QTNF9B
C1QTNF9→
←CENPJ

**Extended Data Figure 13 | Locus plots for genomic regions containing significant signals for overall CHIP in African ancestry-stratified GWAS meta-analysis.** CHIP: clonal hematopoiesis of indeterminate potential.

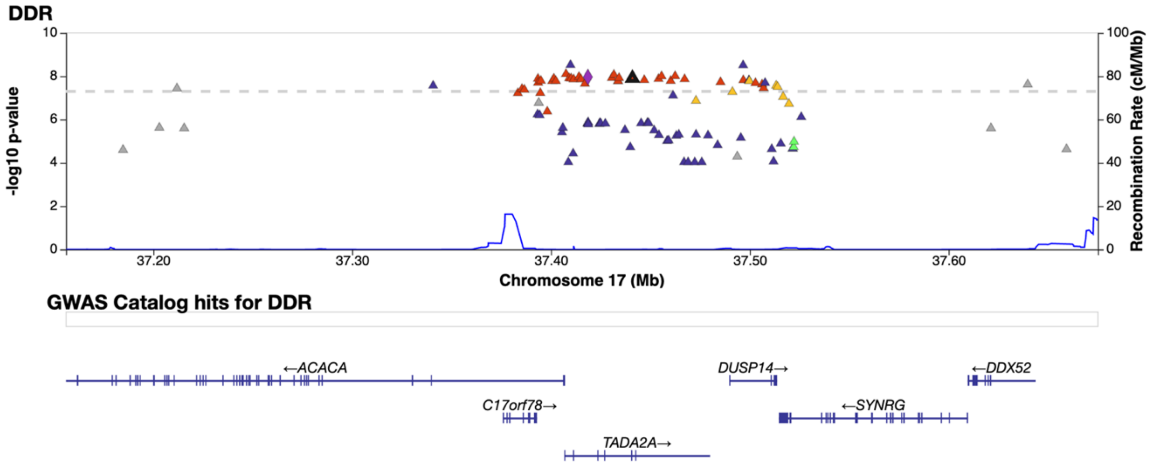

**Extended Data Figure 14 | Locus plots for genomic regions containing significant signals for GWAS meta-analysis for CHIP traits in female.** CHIP: clonal hematopoiesis of indeterminate potential.

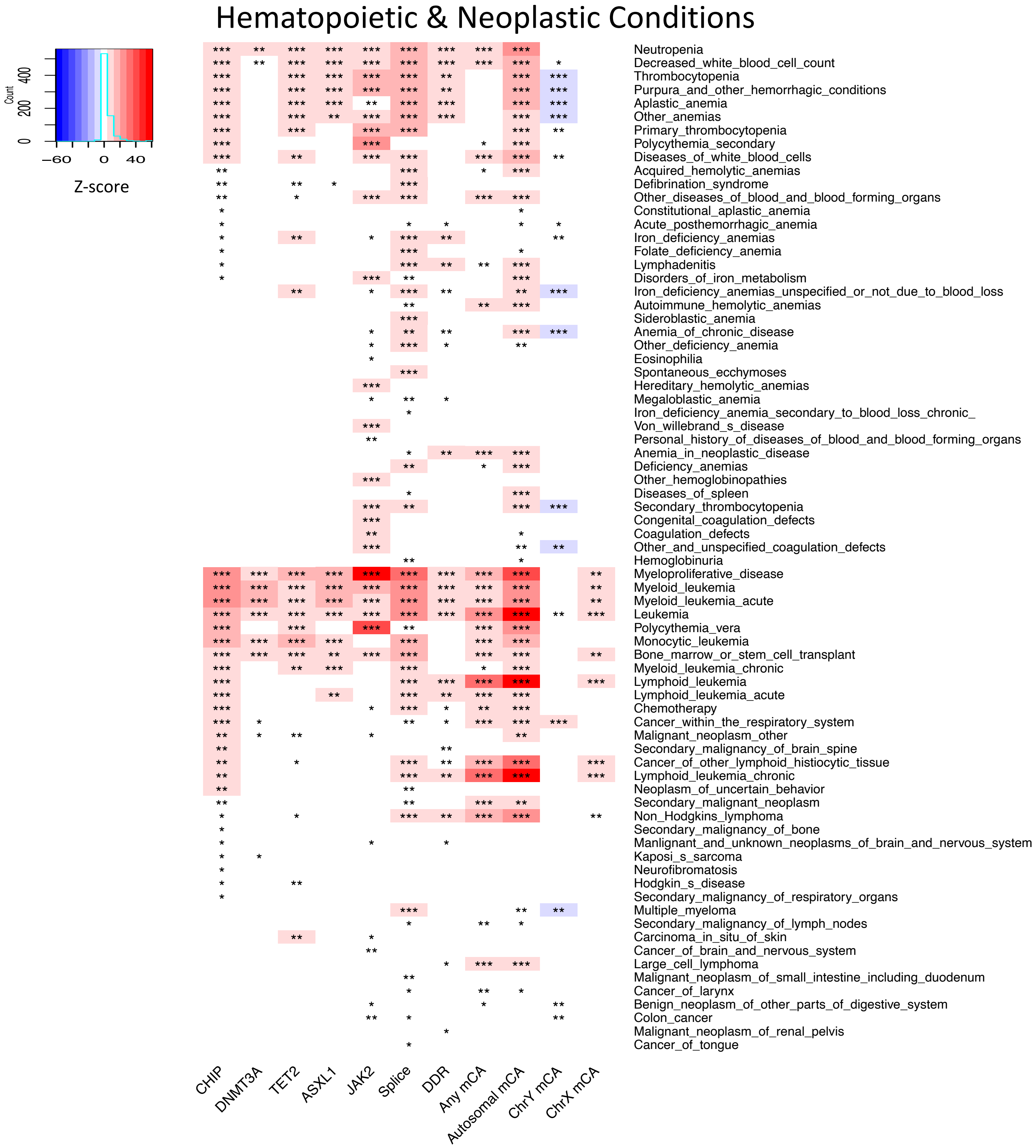

Extended Data Figure 15: Heat map of significant (FDR<0.05) associations between CHIP or mCAs and incident hematopoietic conditions and neoplasms from phenome-wide association analysis, and their respective associations with mCAs. PheCode phenotypes were filtered to those with CHIP FDR<0.05 among incident phenotypes. Analyses. The mCA analyses were conducted among the same subset of individuals in the UK Biobank with CHIP calls available. All analyses were adjusted for age, age$^2$, sex (not used for ChrY and Chr X mCA analysis), smoking status (using a 25-factor smoking status adjustment in the UK Biobank and current/prior/never smoker status in other cohorts), tobacco use disorder, and principal components 1-10 of genetic ancestry. Colors in heat map reflect z-score (beta/se) of associations. *: 0.01≤FDR≤0.05; **: 0.0001<FDR≤0.01; ***: FDR<0.0001

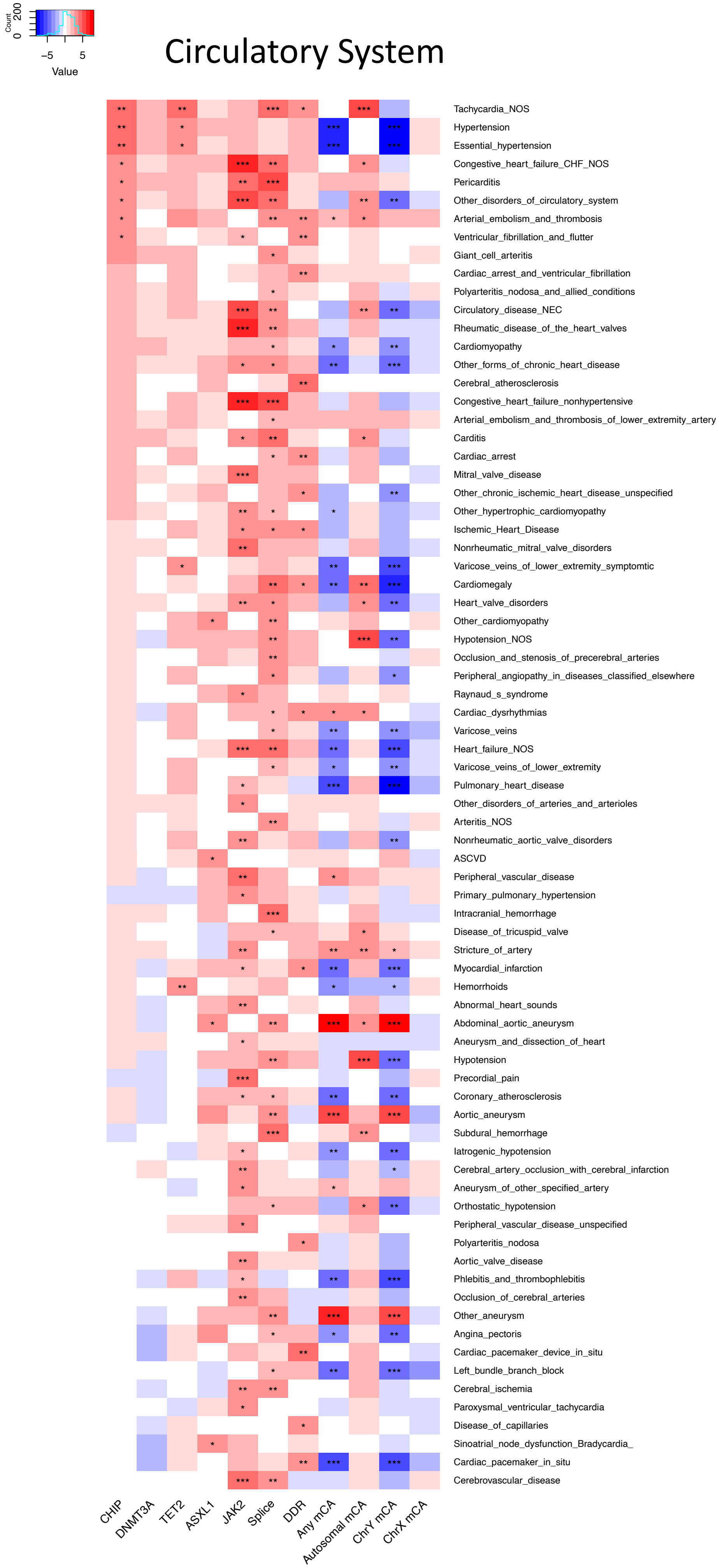

Extended Data Figure 16: Heat map of significant (FDR<0.05) associations between CHIP or mCAs and incident circulatory system conditions from phenome-wide association analysis, and their respective associations with mCAs. PheCode phenotypes were filtered to those with CHIP FDR<0.05 among incident phenotypes.

### Infectious Diseases, Injuries, Poisonings, and Symptoms

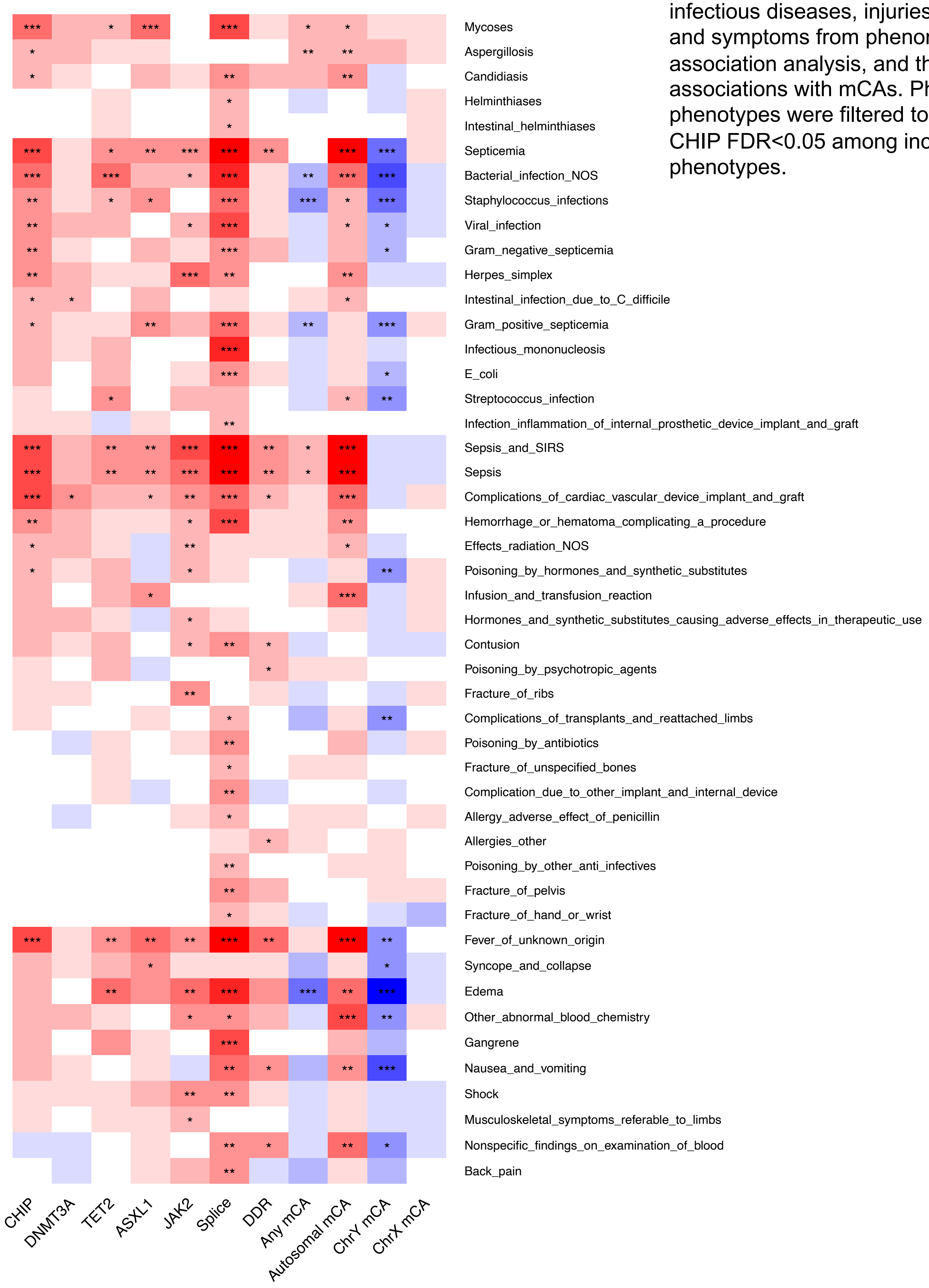

Extended Data Figure 17: Heat map of significant (FDR<0.05) associations between CHIP or mCAs and incident infectious diseases, injuries, poisonings, and symptoms from phenome-wide association analysis, and their respective associations with mCAs. PheCode phenotypes were filtered to those with CHIP FDR<0.05 among incident phenotypes.

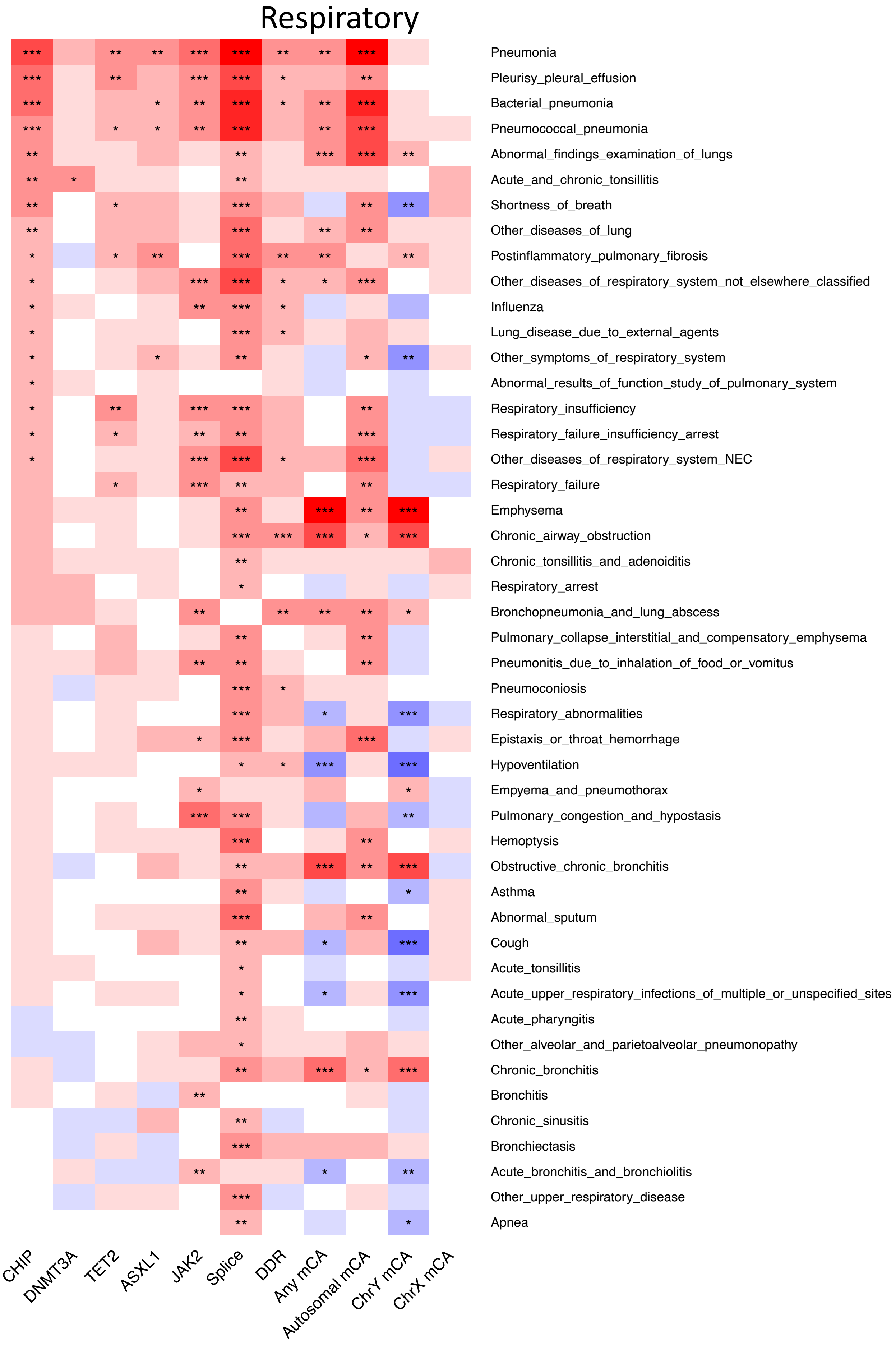

Extended Data Figure 18: Heat map of significant (FDR<0.05) associations between CHIP or mCAs and incident respiratory conditions from phenome-wide association analysis, and their respective associations with mCAs. PheCode phenotypes were filtered to those with CHIP FDR<0.05 among incident phenotypes.

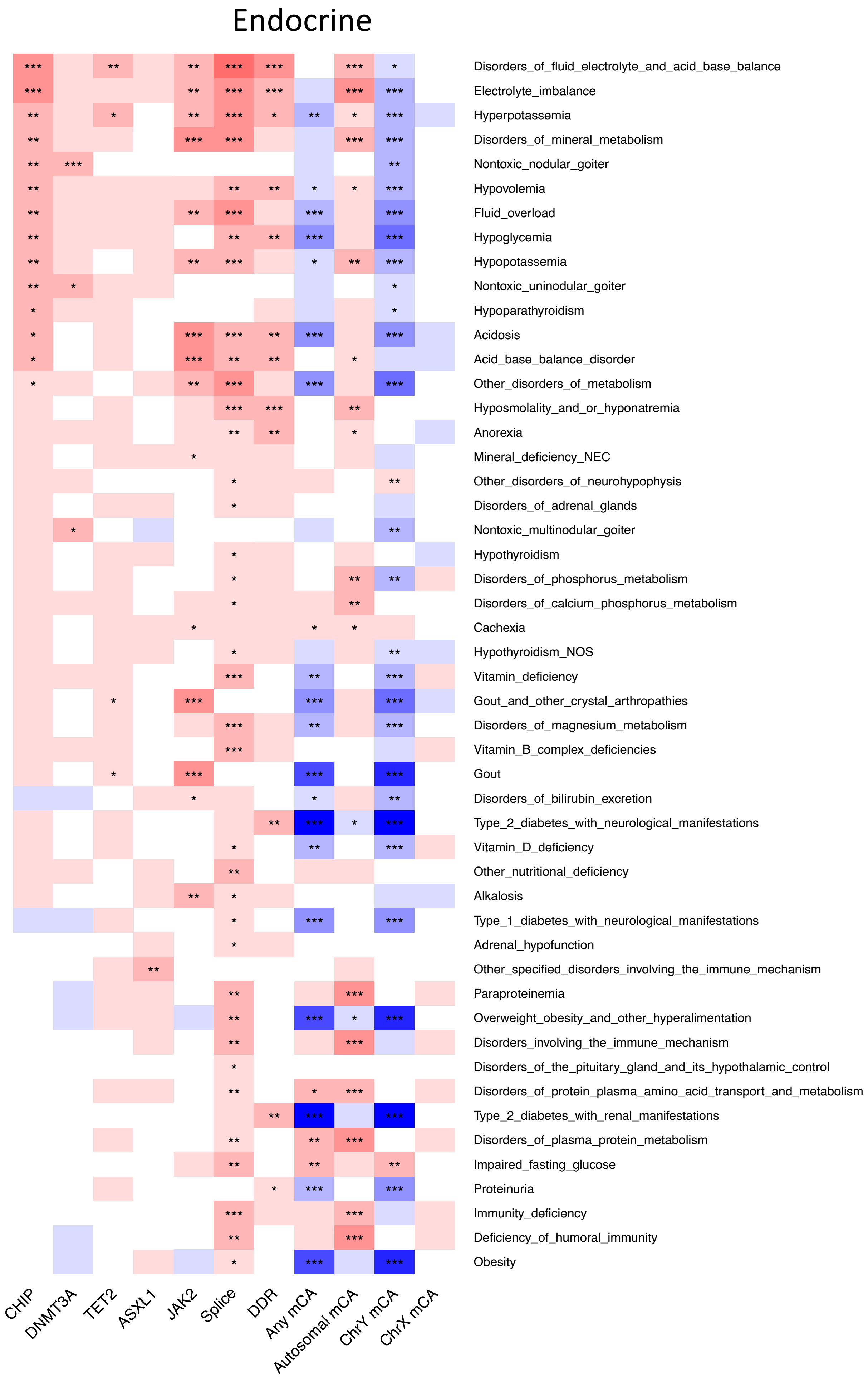

Extended Data Figure 19: Heat map of significant (FDR<0.05) associations between CHIP or mCAs and incident endocrine conditions from phenome-wide association analysis, and their respective associations with mCAs. PheCode phenotypes were filtered to those with CHIP FDR<0.05 among incident phenotypes.

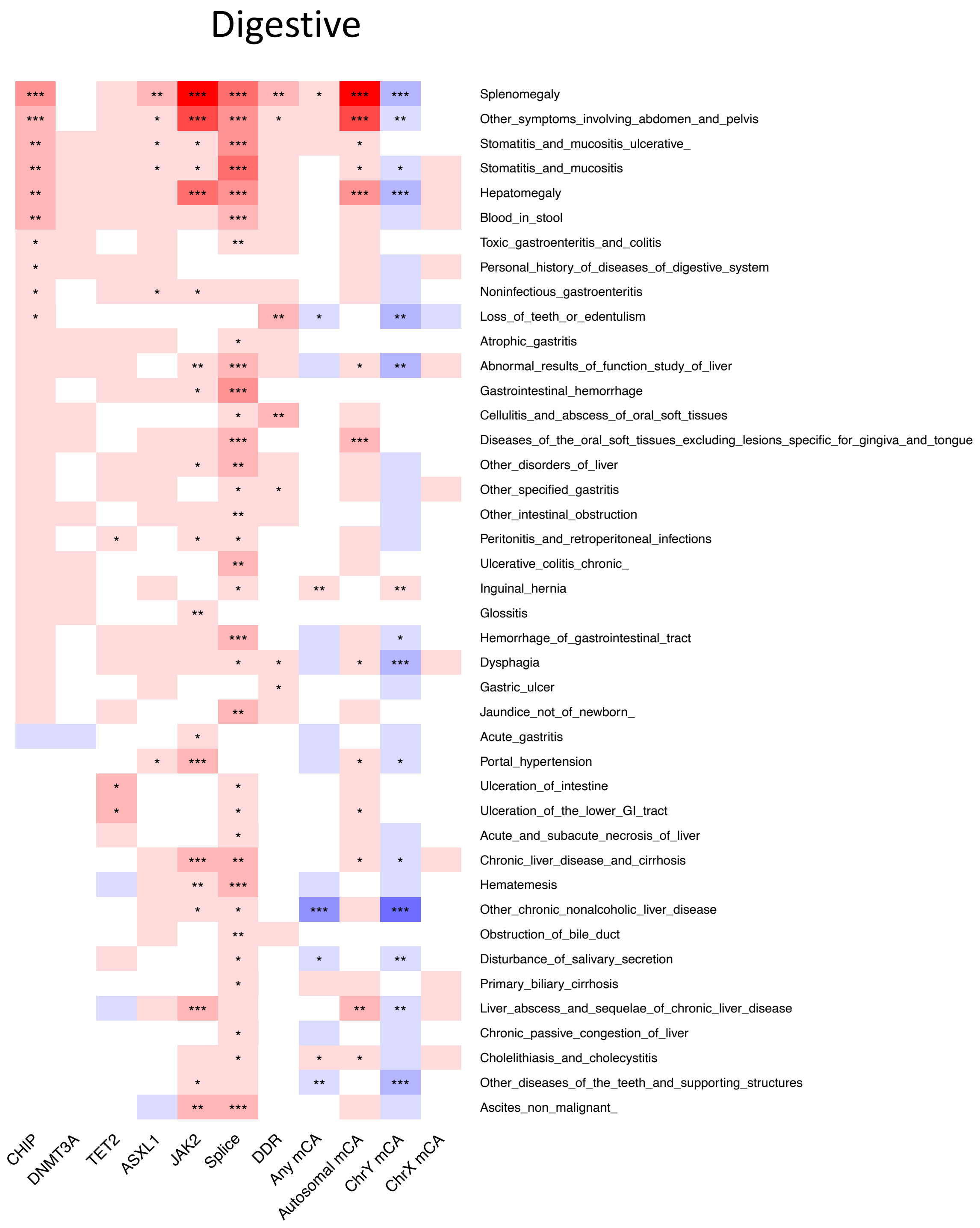

Extended Data Figure 20: Heat map of significant (FDR<0.05) associations between CHIP or mCAs and incident digestive conditions from phenome-wide association analysis, and their respective associations with mCAs. PheCode phenotypes were filtered to those with CHIP FDR<0.05 among incident phenotypes.

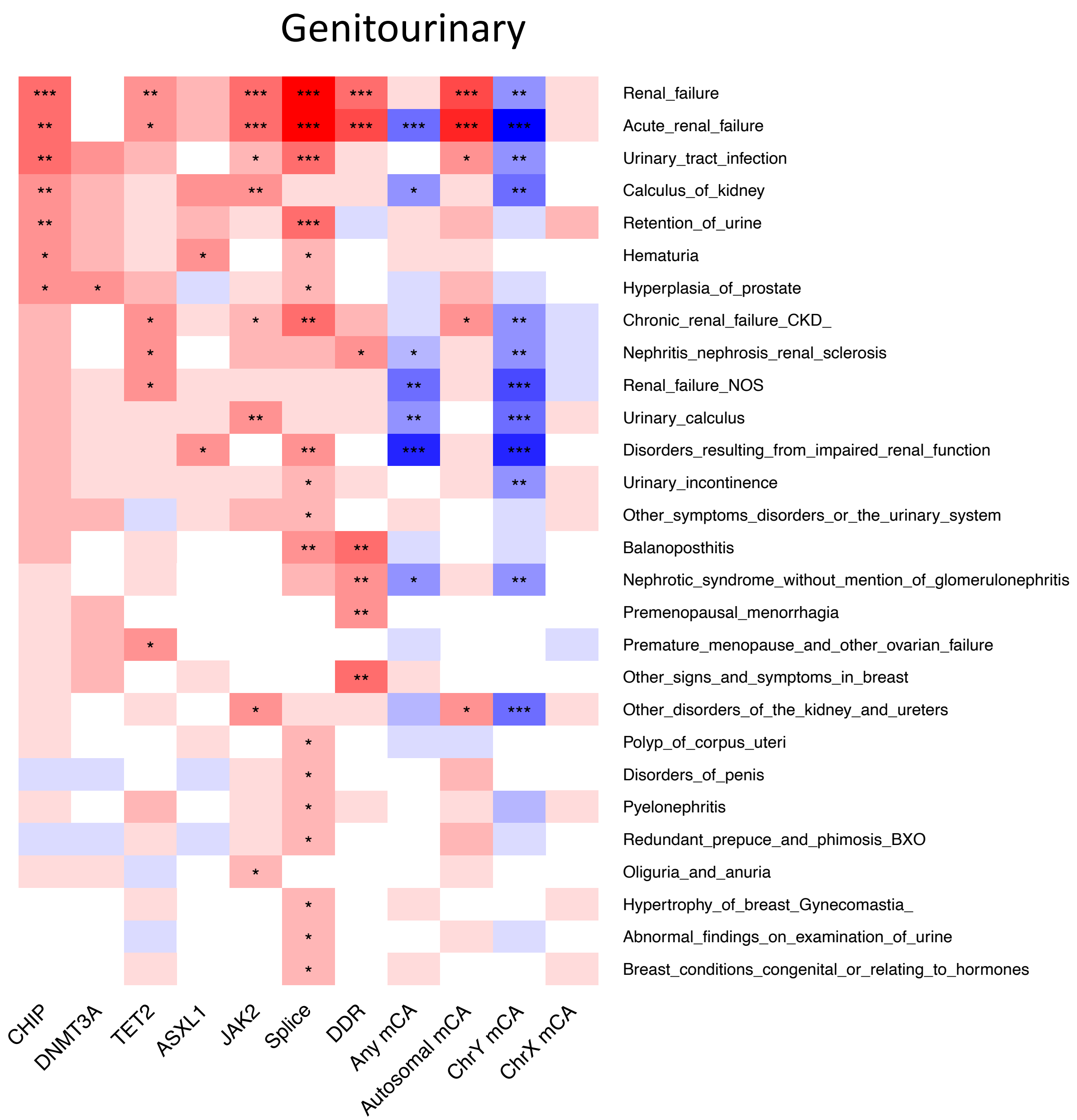

Extended Data Figure 21: Heat map of significant (FDR<0.05) associations between CHIP or mCAs and incident genitourinary conditions from phenome-wide association analysis, and their respective associations with mCAs. PheCode phenotypes were filtered to those with CHIP FDR<0.05 among incident phenotypes.

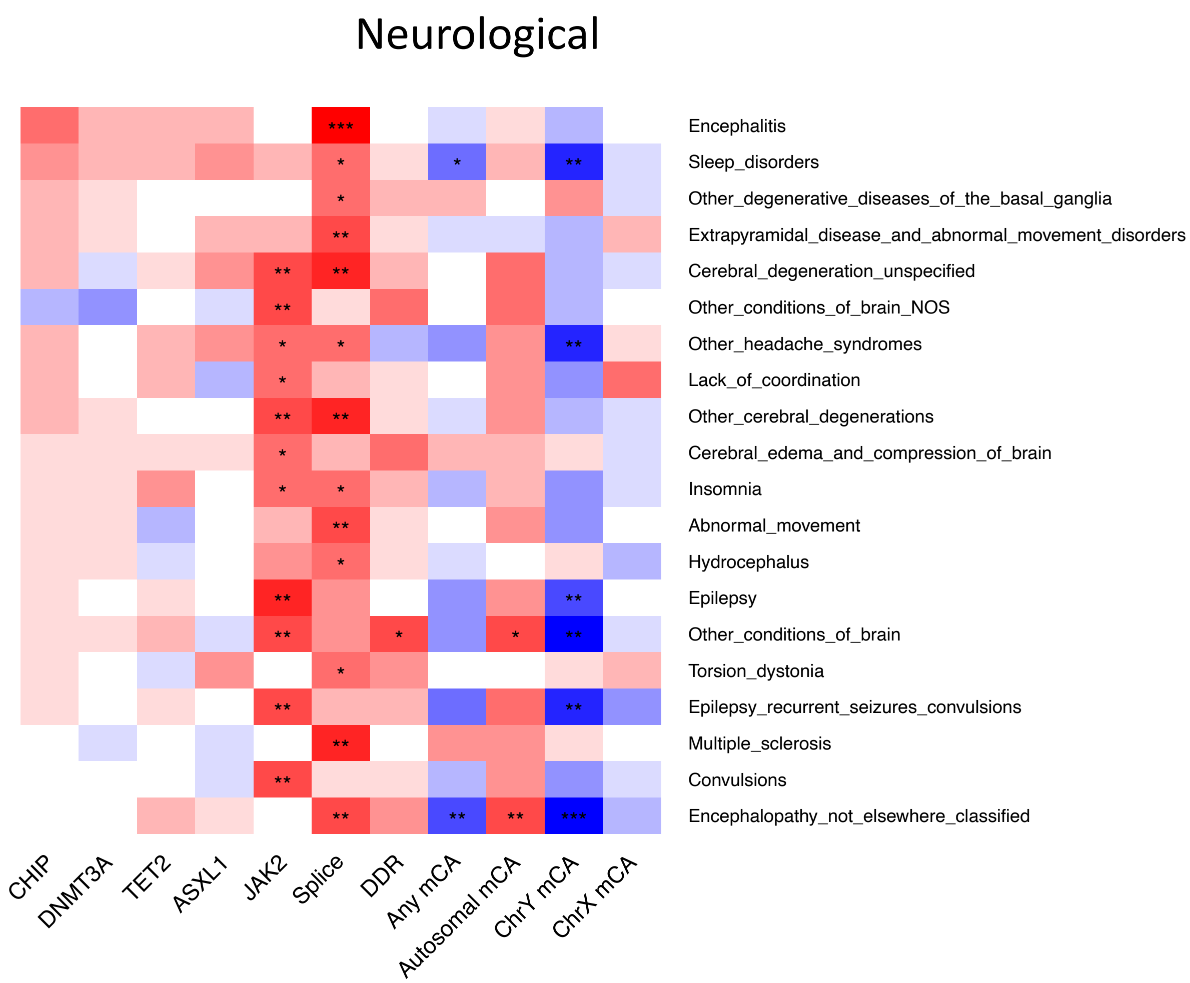

Extended Data Figure 22: Heat map of significant (FDR<0.05) associations between CHIP or mCAs and incident neurological conditions from phenome-wide association analysis, and their respective associations with mCAs. PheCode phenotypes were filtered to those with CHIP FDR<0.05 among incident phenotypes.

Musculoskeletal

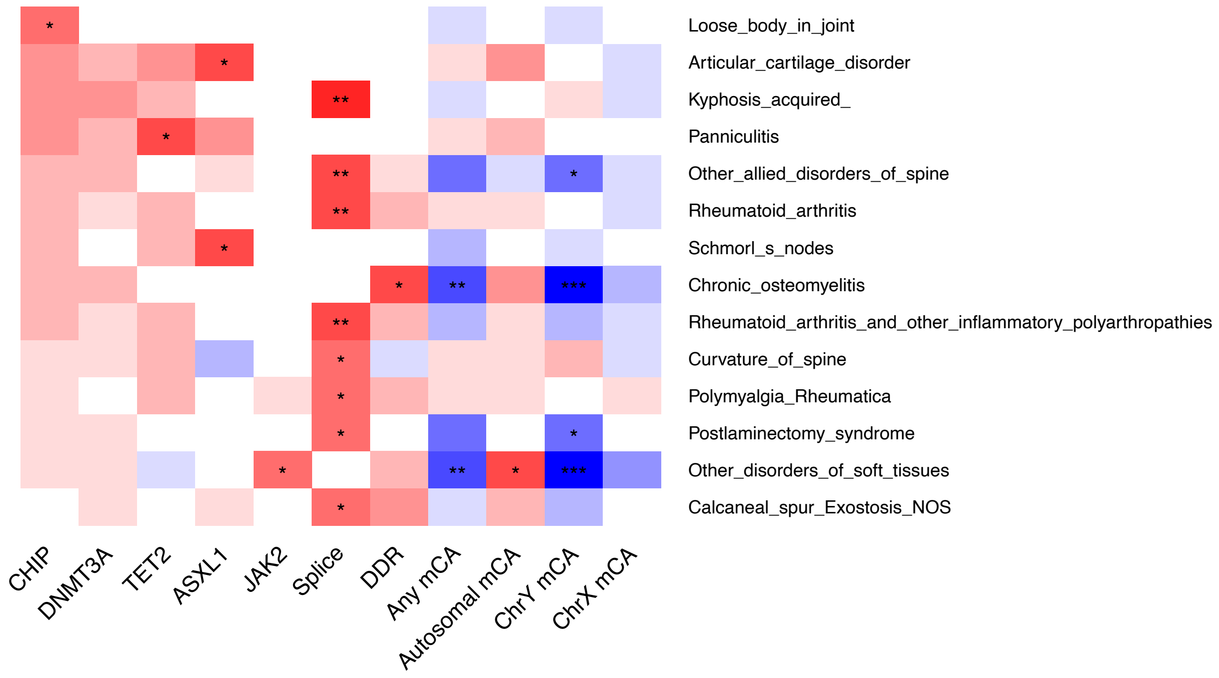

Extended Data Figure 23: Heat map of significant (FDR<0.05) associations between CHIP or mCAs and incident musculoskeletal conditions from phenome-wide association analysis, and their respective associations with mCAs. PheCode phenotypes were filtered to those with CHIP FDR<0.05 among incident phenotypes.

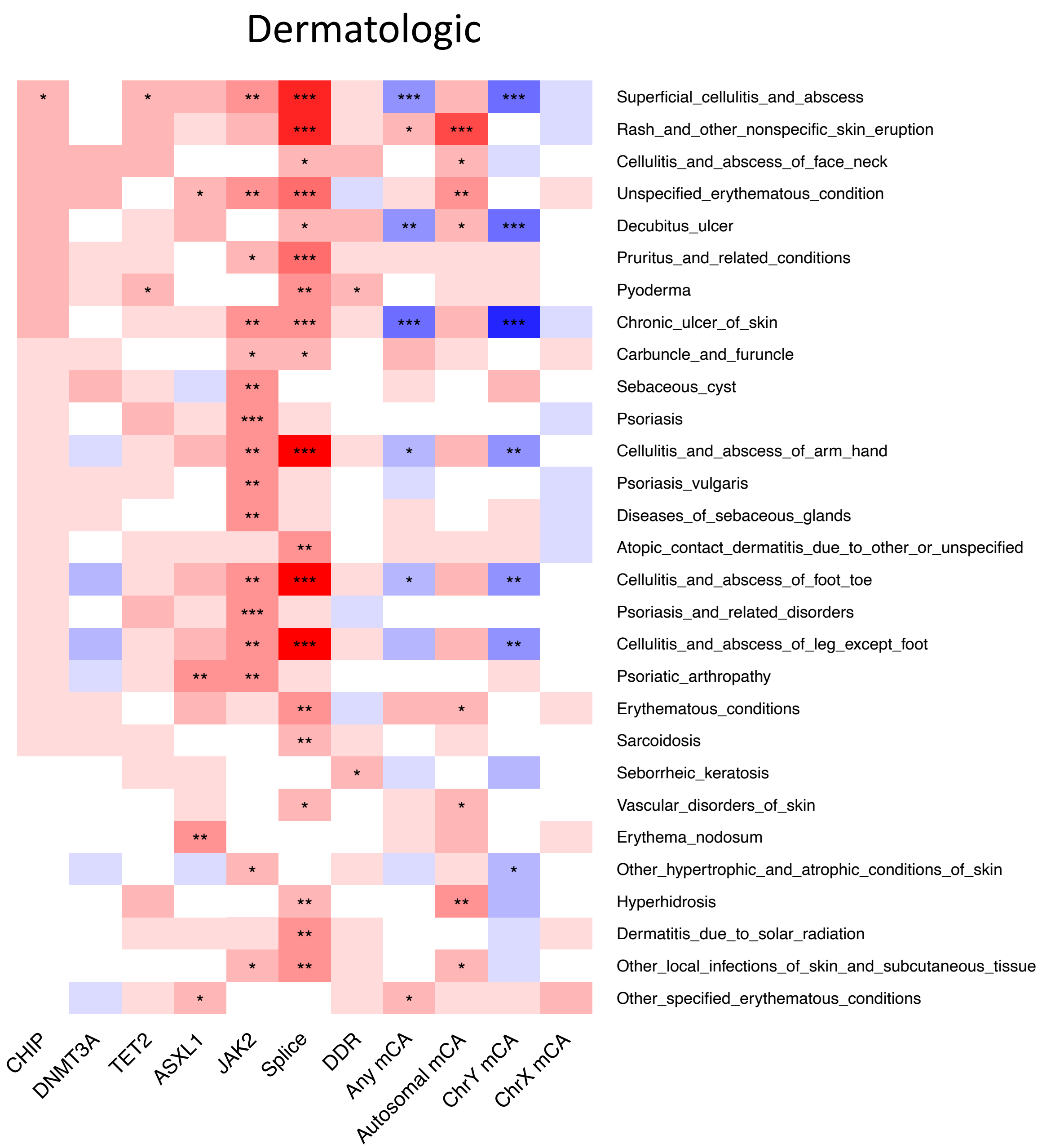

Extended Data Figure 24: Heat map of significant (FDR<0.05) associations between CHIP or mCAs and incident dermatologic conditions from phenome-wide association analysis, and their respective associations with mCAs. PheCode phenotypes were filtered to those with CHIP FDR<0.05 among incident phenotypes.

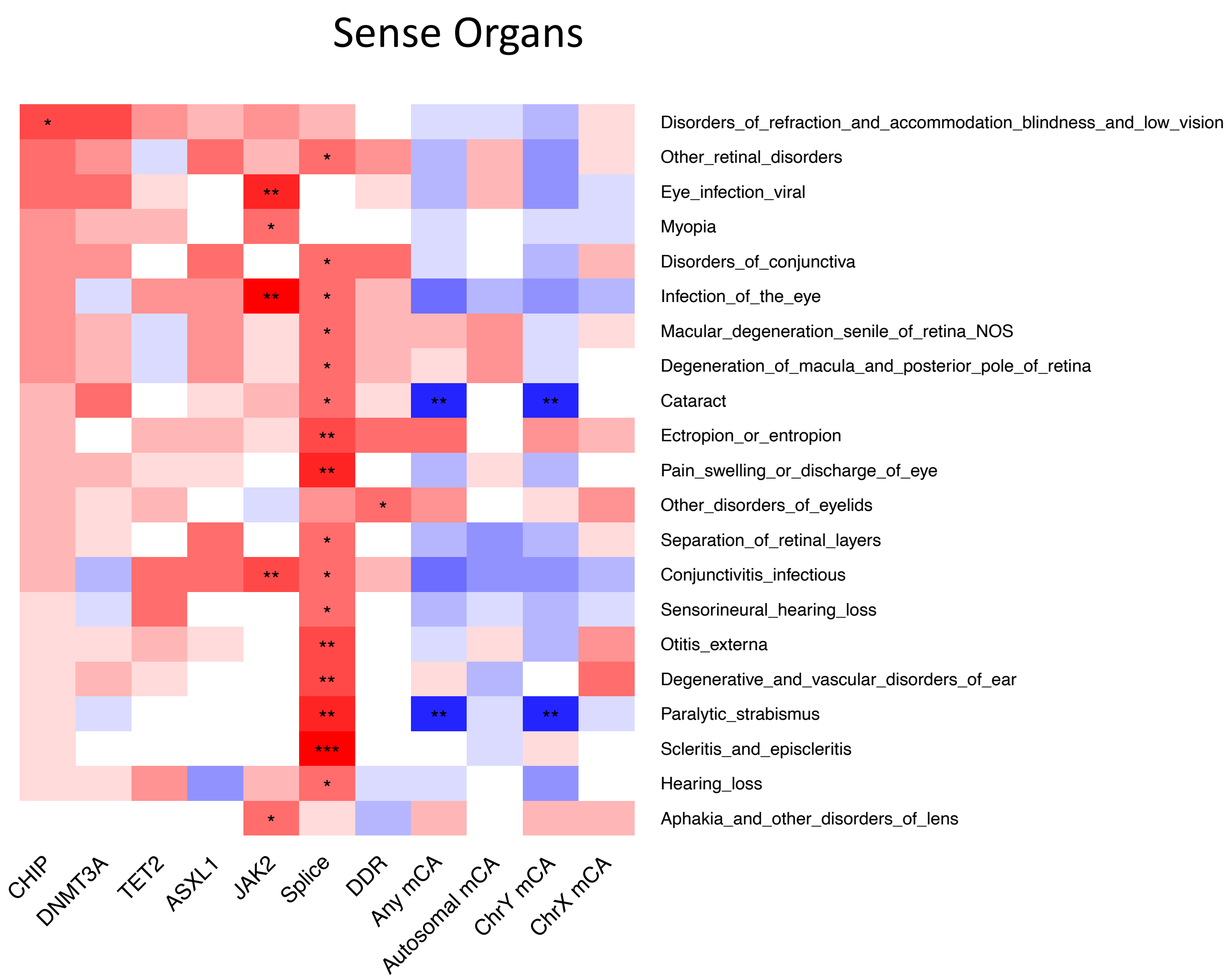

Extended Data Figure 25: Heat map of significant (FDR<0.05) associations between CHIP or mCAs and incident disorders of sense organs (eyes & ears) from phenome-wide association analysis, and their respective associations with mCAs. PheCode phenotypes were filtered to those with CHIP FDR<0.05 among incident phenotypes.

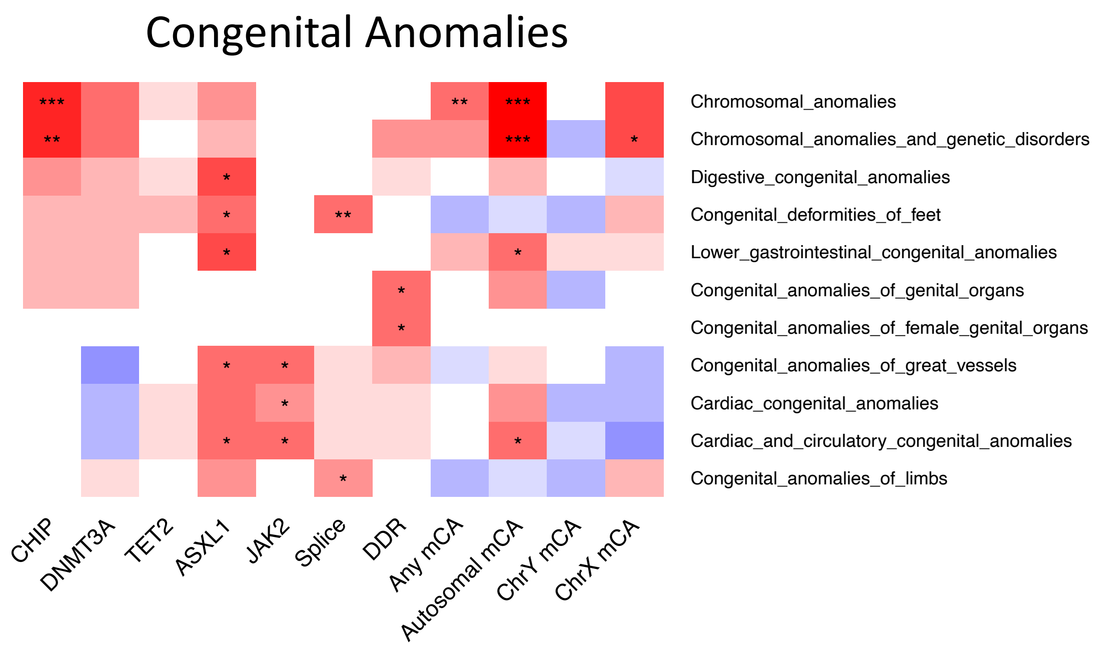

Extended Data Figure 26: Heat map of significant (FDR<0.05) associations between CHIP or mCAs and incident congenital anomalies from phenome-wide association analysis, and their respective associations with mCAs. PheCode phenotypes were filtered to those with CHIP FDR<0.05 among incident phenotypes.

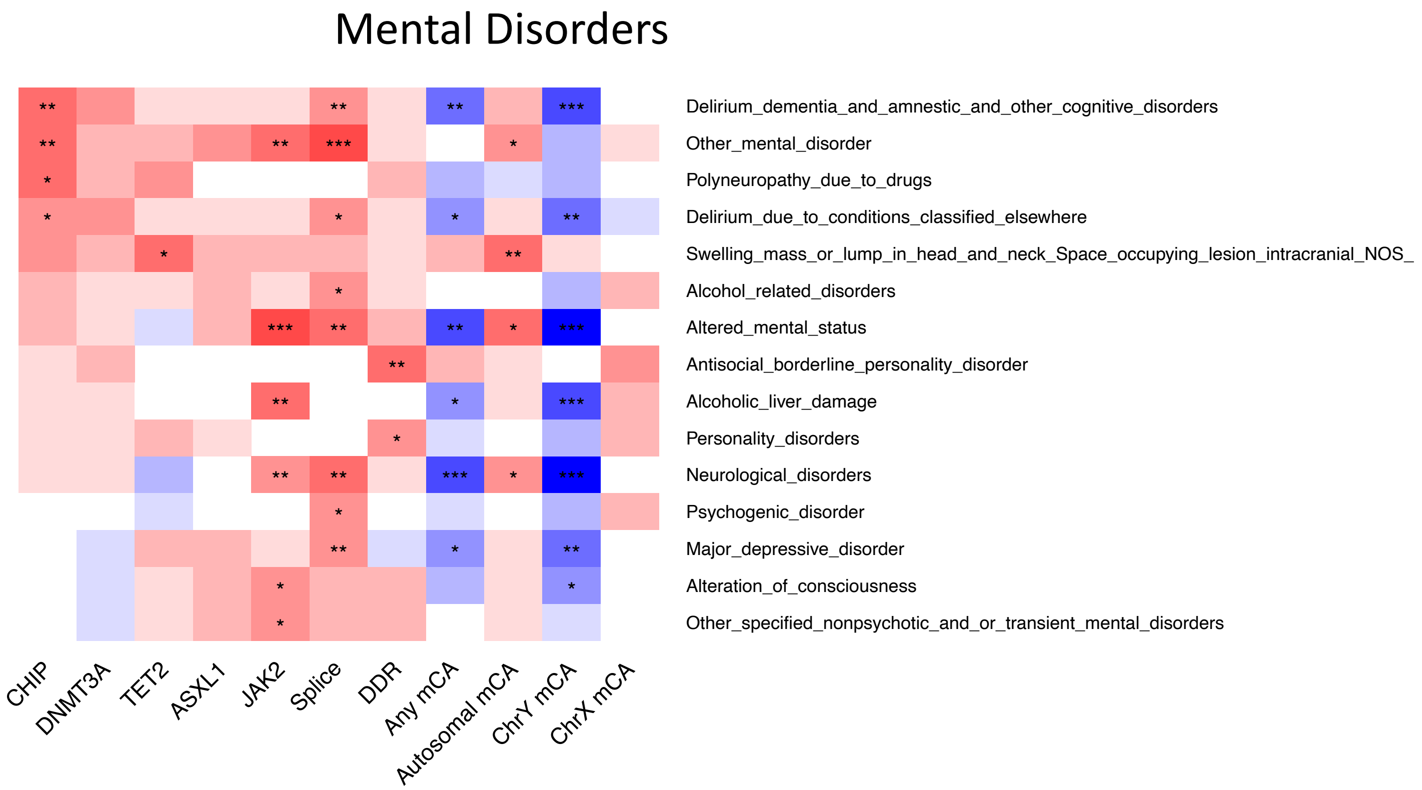

Extended Data Figure 27: Heat map of significant (FDR<0.05) associations between CHIP or mCAs and incident mental disorders from phenome-wide association analysis, and their respective associations with mCAs. PheCode phenotypes were filtered to those with CHIP FDR<0.05 among incident phenotypes.

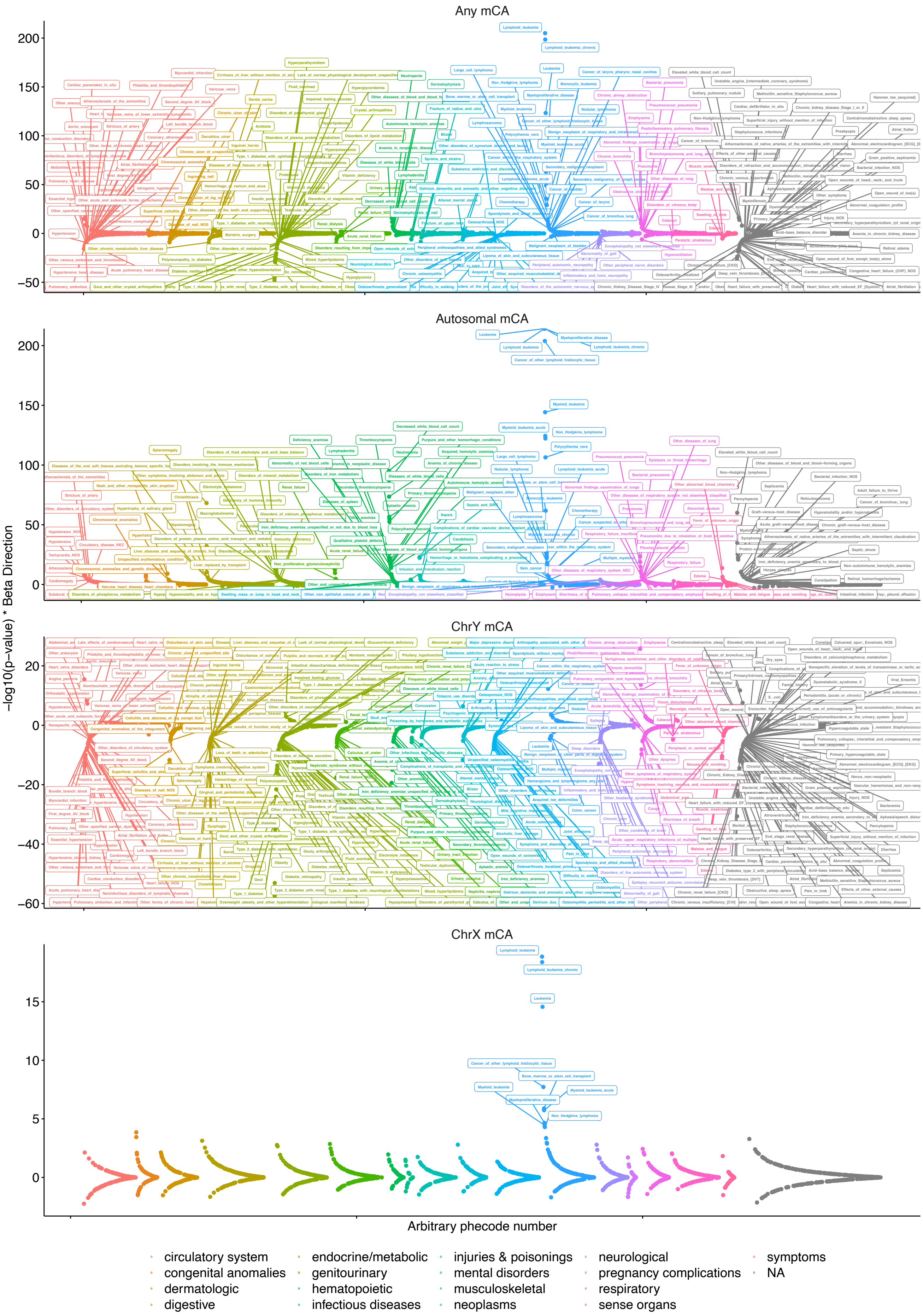

Extended Data Figure 28: mCA Phenome-wide association study results. mCA associations are meta-analyzed across the UK Biobank, Mass-General-Brigham Biobank, BioVU, and Million Veterans Program. All analyses were adjusted for age, age$^2$, sex (not used for ChrY and Chr X mCA analysis), smoking status (using a 25-factor smoking status adjustment in the UK Biobank and current/prior/never smoker status in other cohorts), tobacco use disorder, and principal components 1-10 of genetic ancestry. Labels reflect associations with FDR<0.01.

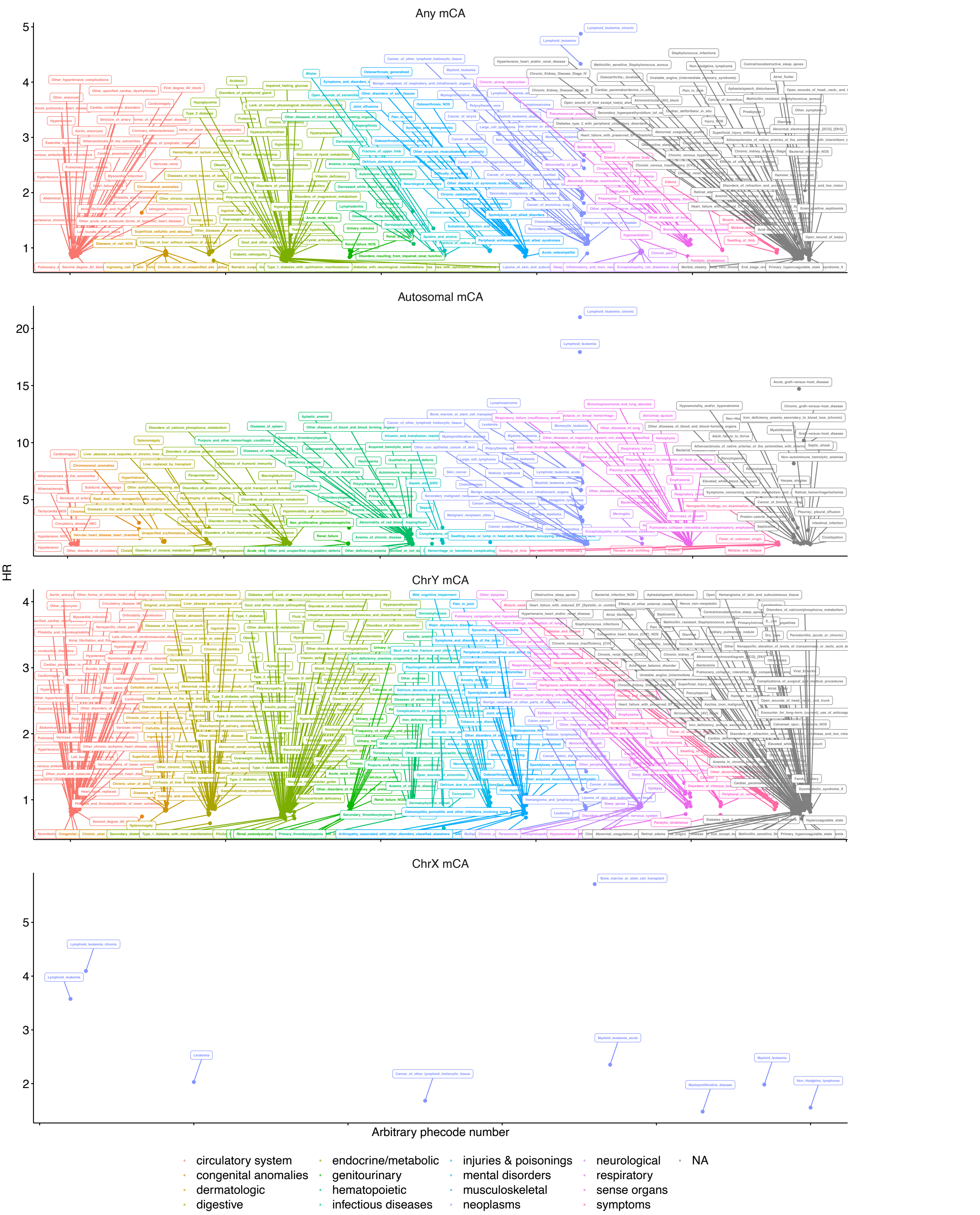

Extended Data Figure 29: mCA Phenome-wide association study results. mCA associations are meta-analyzed across the UK Biobank, Mass-General-Brigham Biobank, BioVU, and Million Veterans Program. All analyses were adjusted for age, age$^2$, sex (not used for ChrY and Chr X mCA analysis), smoking status (using a 25-factor smoking status adjustment in the UK Biobank and current/prior/never smoker status in other cohorts), tobacco use disorder, and principal components 1-10 of genetic ancestry. Y-axis reflects the HR. Labels reflect associations with FDR<0.01.

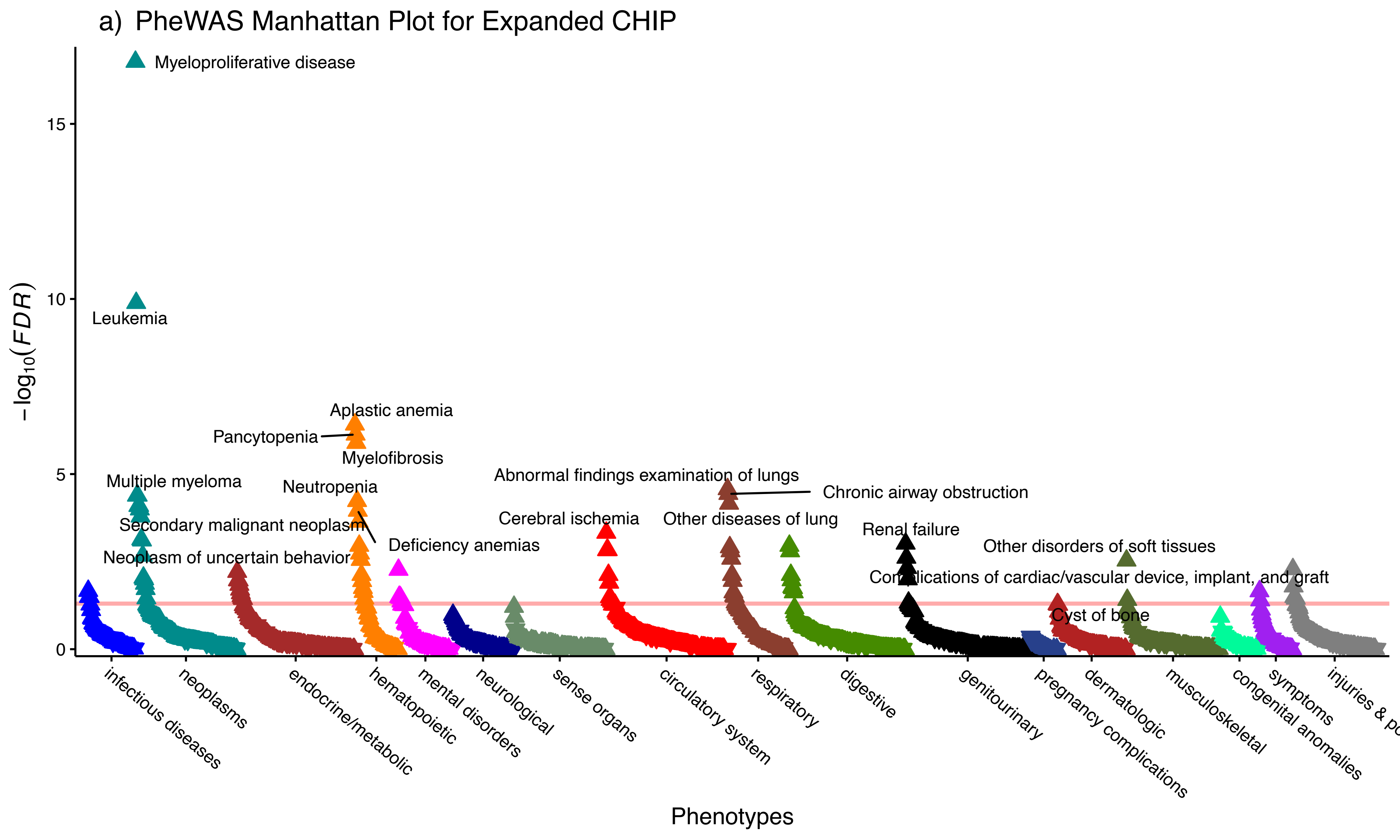
a) PheWAS Manhattan Plot for Expanded CHIP
Myeloproliferative disease
Leukemia
Aplastic anemia
Pancytopenia
Myelofibrosis
Multiple myeloma
Neutropenia
Abnormal findings examination of lungs
Chronic airway obstruction
Secondary malignant neoplasm
Cerebral ischemia
Other diseases of lung
Renal failure
Deficiency anemias
Neoplasm of uncertain behavior
Other disorders of soft tissues
Complications of cardiac/vascular device, implant, and graft
Cyst of bone
15
10
5
0
$-\log_{10}(FDR)$
infectious diseases
neoplasms
endocrine/metabolic
hematopoietic
mental disorders
neurological
sense organs
circulatory system
respiratory
digestive
genitourinary
pregnancy complications
dermatologic
musculoskeletal
congenital anomalies
symptoms
Phenotypes

b) PheWAS Manhattan Plot for *DNMT3A* CHIP

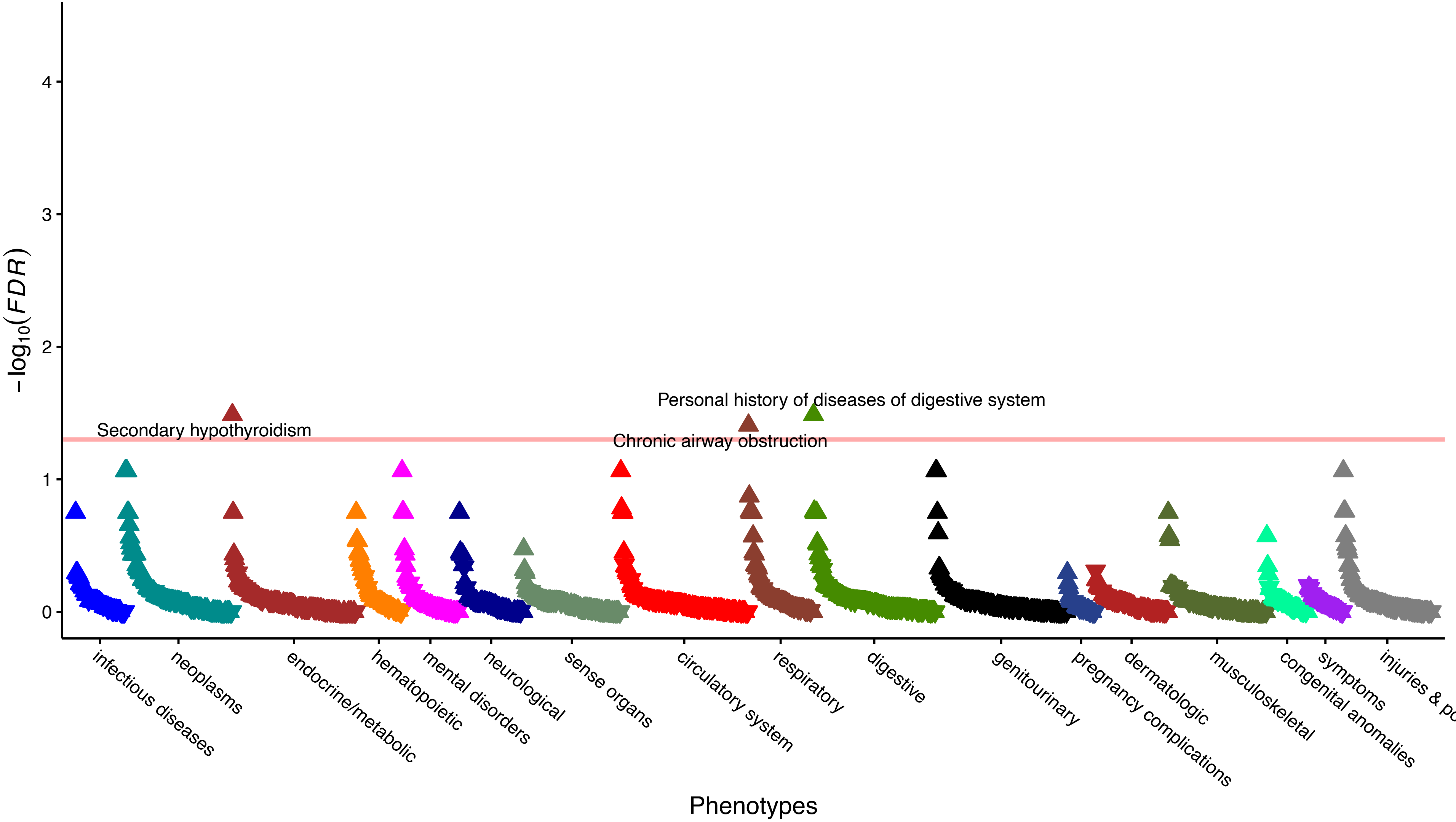

c) PheWAS Manhattan Plot for *TET2* CHIP

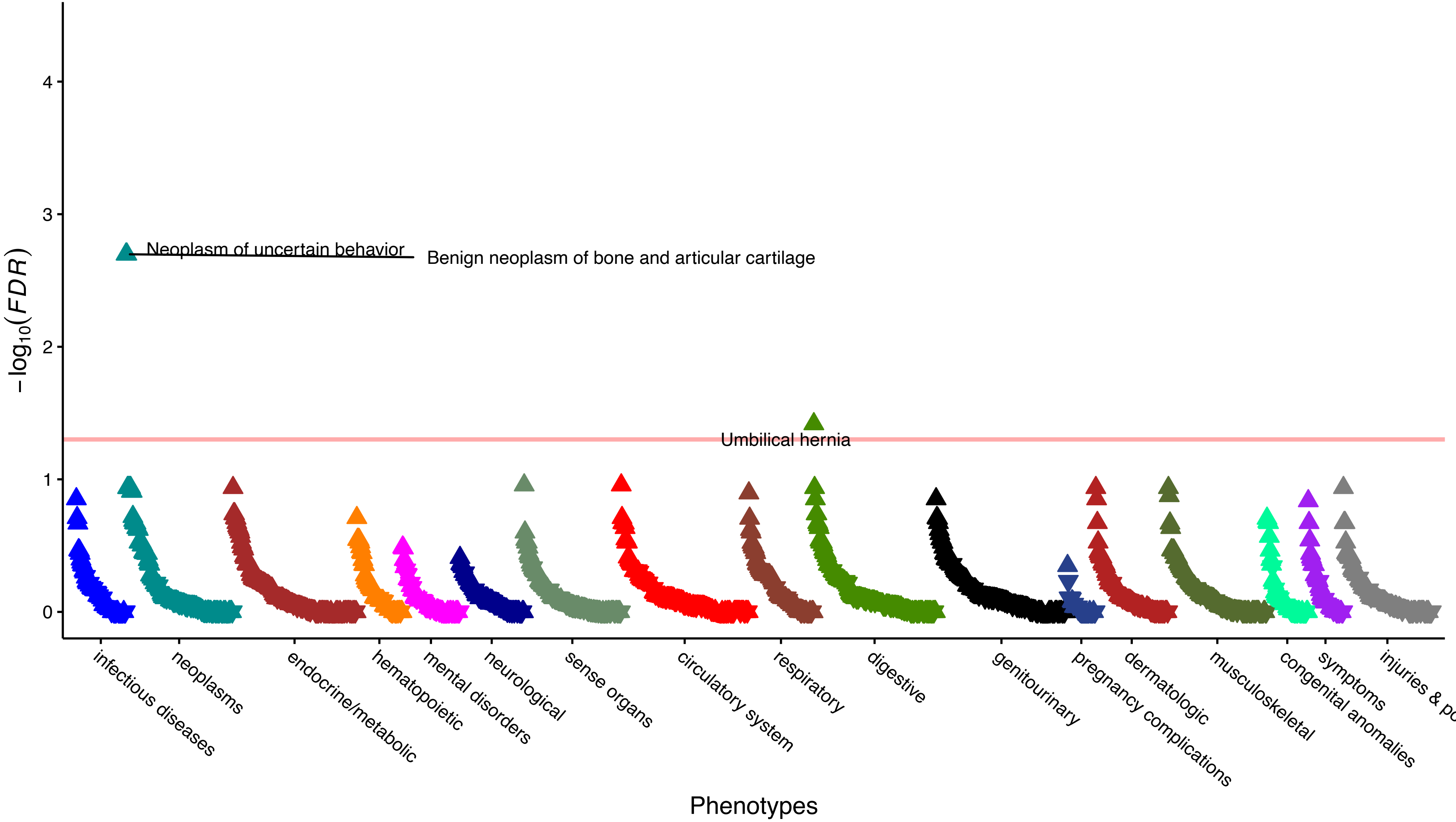

d) PheWAS Manhattan Plot for *ASXL1* CHIP

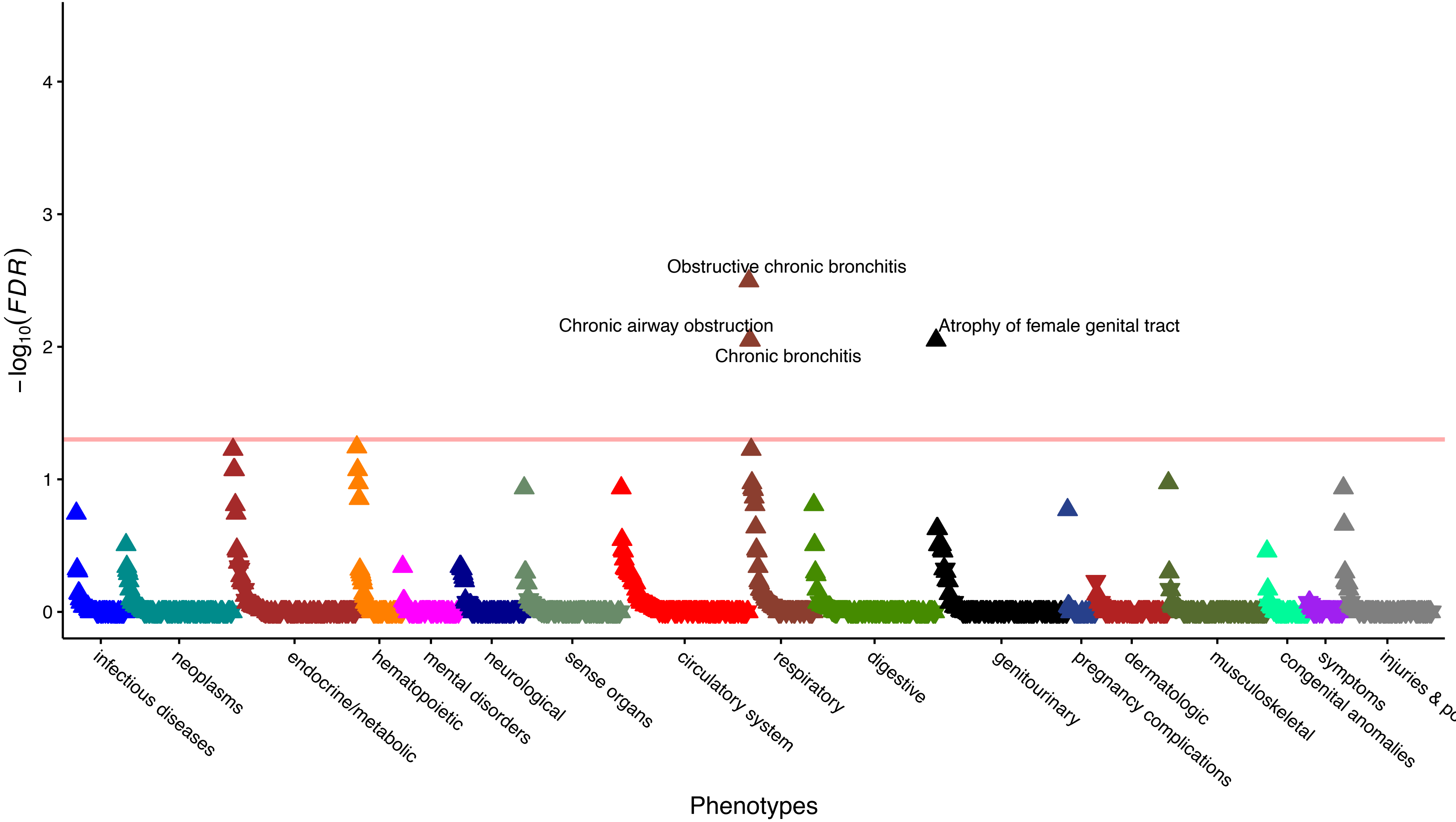

e) PheWAS Manhattan Plot for *JAK2* CHIP

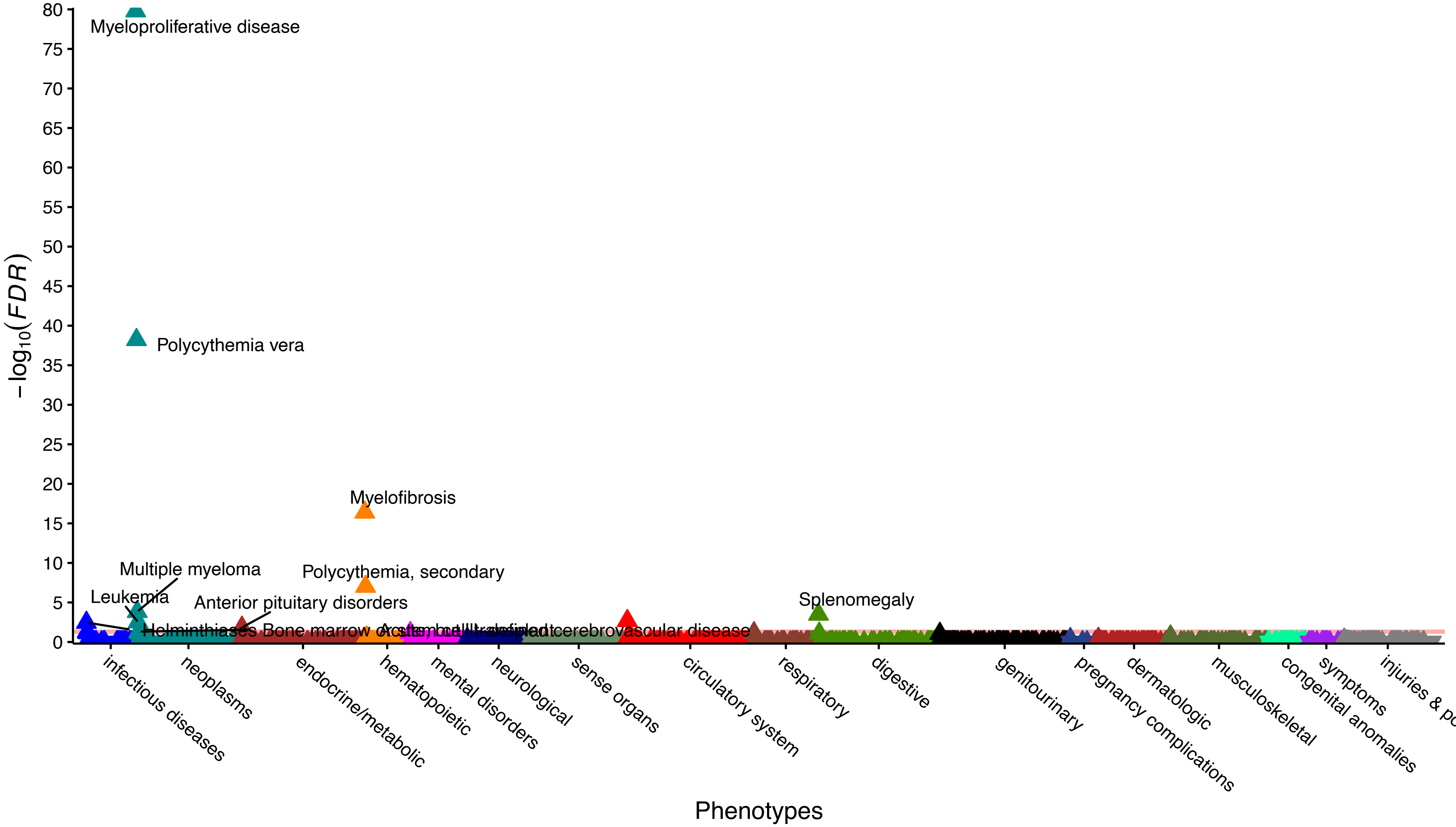

f) PheWAS Manhattan Plot for *PPM1D* CHIP

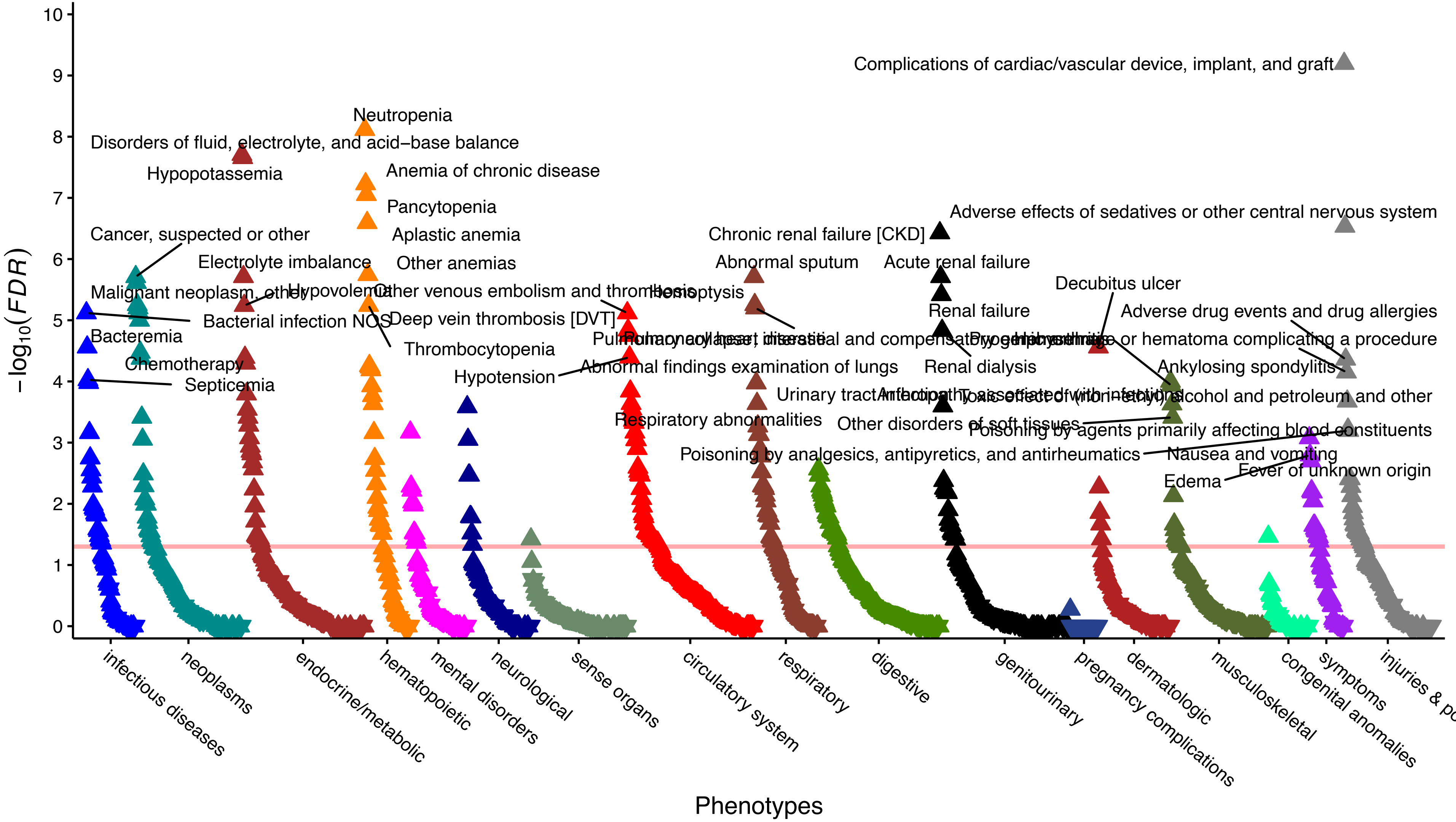

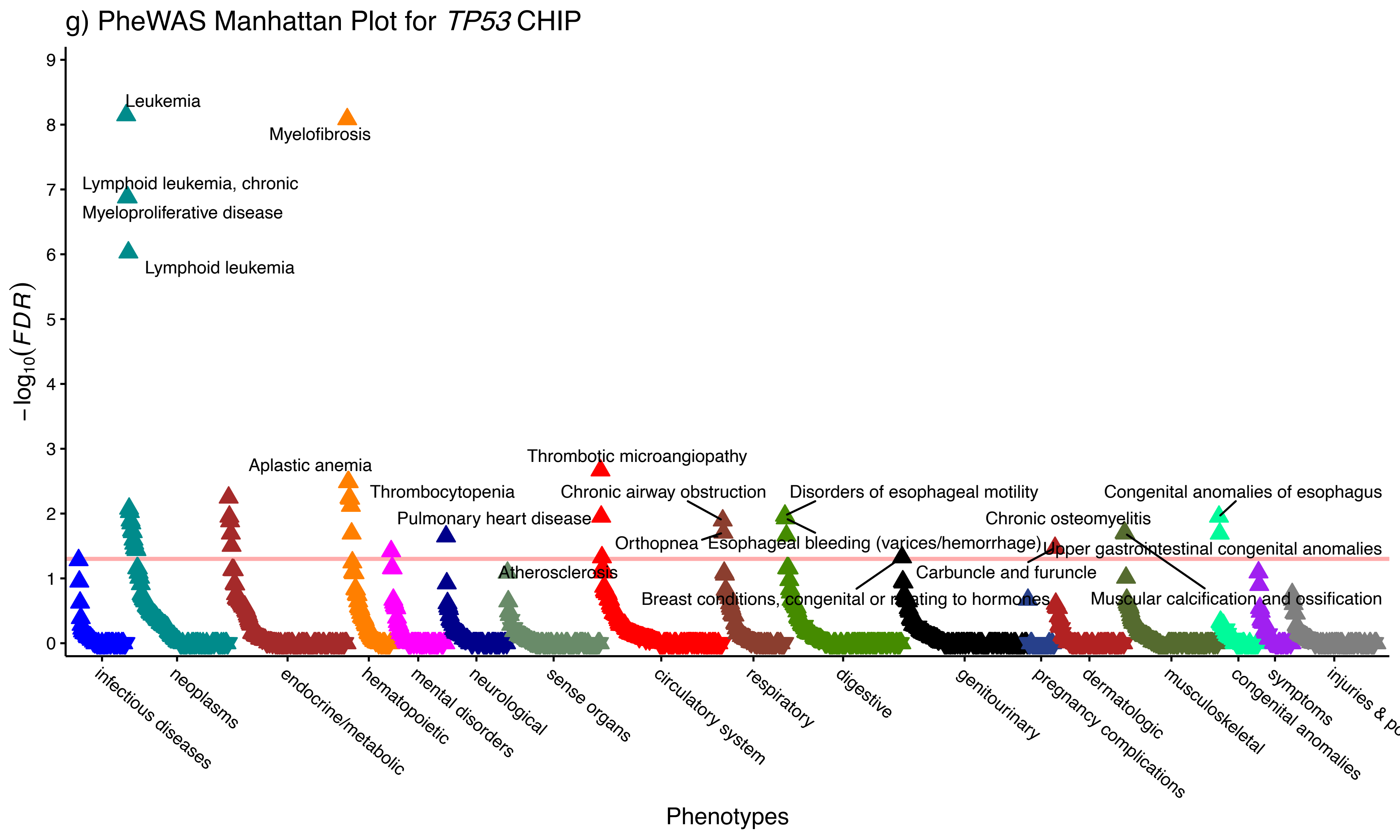

g) PheWAS Manhattan Plot for TP53 CHIP
−log10(FDR)
0
1
2
3
4
5
6
7
8
9
Leukemia
Myelofibrosis
Lymphoid leukemia, chronic
Myeloproliferative disease
Lymphoid leukemia
Aplastic anemia
Thrombotic microangiopathy
Thrombocytopenia
Chronic airway obstruction
Disorders of esophageal motility
Congenital anomalies of esophagus
Pulmonary heart disease
Chronic osteomyelitis
Orthopnea
Esophageal bleeding (varices/hemorrhage)
Upper gastrointestinal congenital anomalies
Atherosclerosis
Carbuncle and furuncle
Breast conditions, congenital or relating to hormones
Muscular calcification and ossification
infectious diseases
neoplasms
endocrine/metabolic
hematopoietic
mental disorders
neurological
sense organs
circulatory system
respiratory
digestive
genitourinary
pregnancy complications
dermatologic
musculoskeletal
congenital anomalies
symptoms
Phenotypes

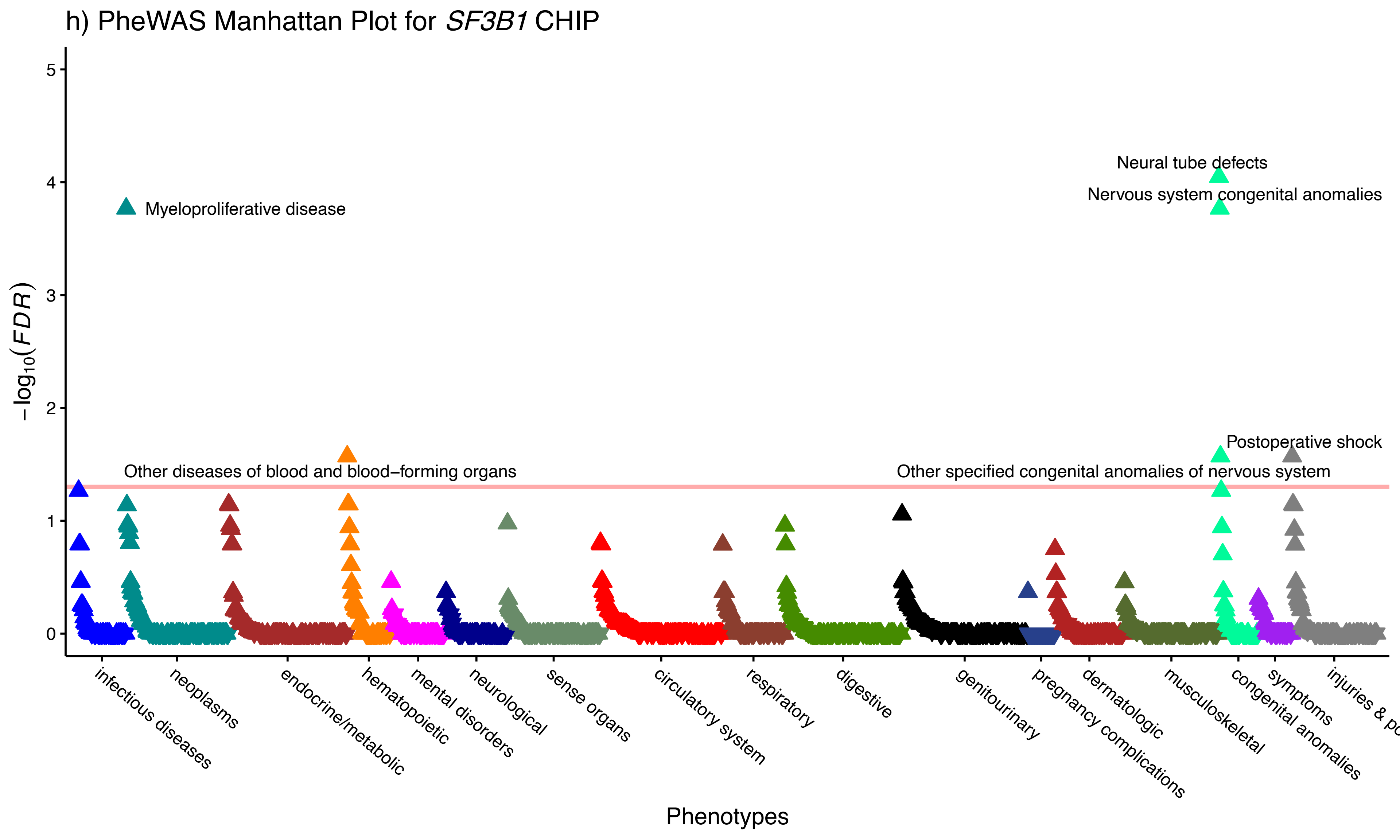
h) PheWAS Manhattan Plot for SF3B1 CHIP
Neural tube defects
Nervous system congenital anomalies
Myeloproliferative disease
Postoperative shock
Other diseases of blood and blood−forming organs
Other specified congenital anomalies of nervous system
−log10(FDR)
0
1
2
3
4
5
infectious diseases
neoplasms
endocrine/metabolic
hematopoietic
mental disorders
neurological
sense organs
circulatory system
respiratory
digestive
genitourinary
pregnancy complications
dermatologic
musculoskeletal
congenital anomalies
symptoms
Phenotypes

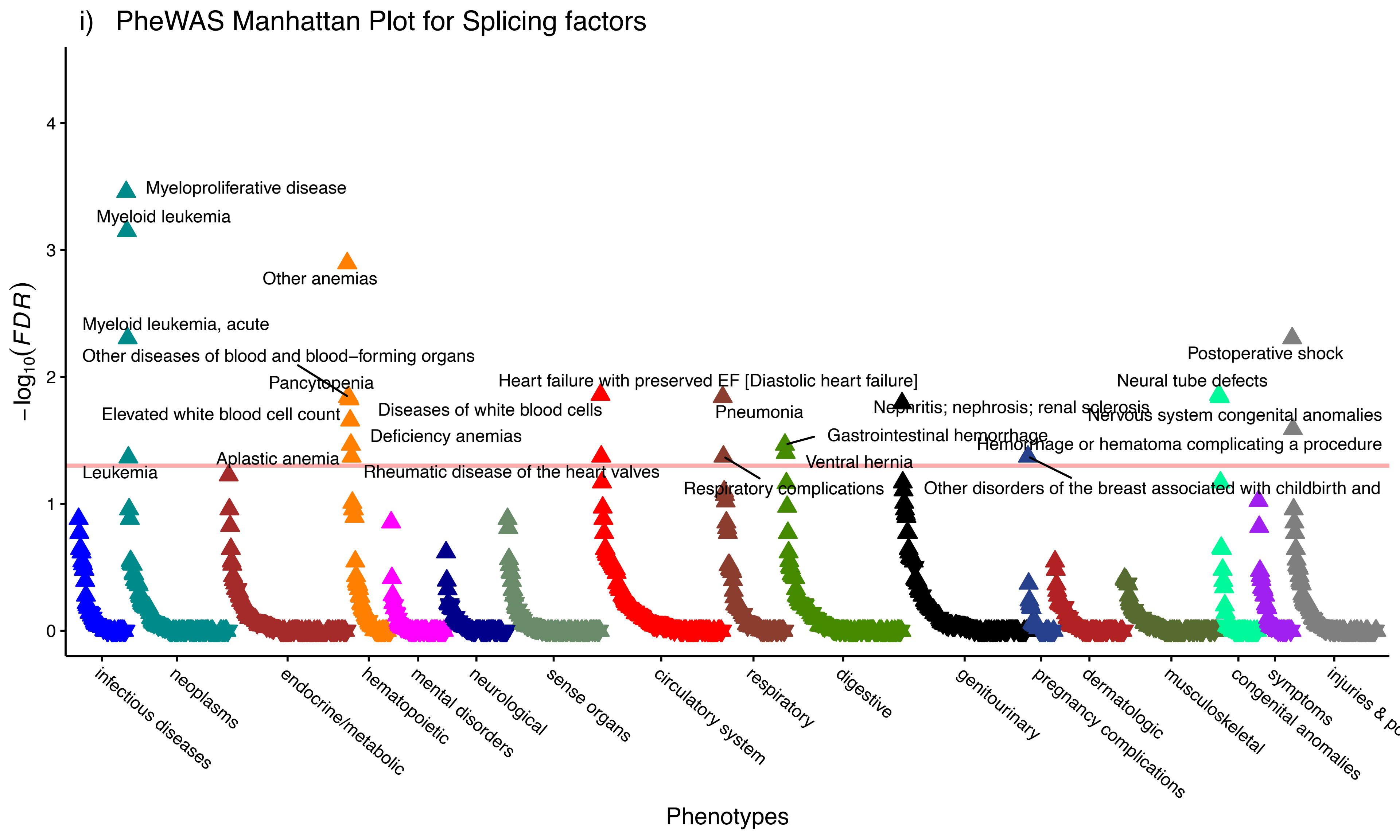
i) PheWAS Manhattan Plot for Splicing factors
−log10(FDR)
Phenotypes
Myeloproliferative disease
Myeloid leukemia
Other anemias
Myeloid leukemia, acute
Other diseases of blood and blood−forming organs
Pancytopenia
Elevated white blood cell count
Diseases of white blood cells
Deficiency anemias
Aplastic anemia
Leukemia
Rheumatic disease of the heart valves
Heart failure with preserved EF [Diastolic heart failure]
Pneumonia
Respiratory complications
Gastrointestinal hemorrhage
Ventral hernia
Nephritis; nephrosis; renal sclerosis
Postoperative shock
Neural tube defects
Nervous system congenital anomalies
Hemorrhage or hematoma complicating a procedure
Other disorders of the breast associated with childbirth and
0
1
2
3
4
infectious diseases
neoplasms
endocrine/metabolic
hematopoietic
mental disorders
neurological
sense organs
circulatory system
respiratory
digestive
genitourinary
pregnancy complications
dermatologic
musculoskeletal
congenital anomalies
symptoms
injuries & po

j) PheWAS Manhattan Plot for DNA damage response genes

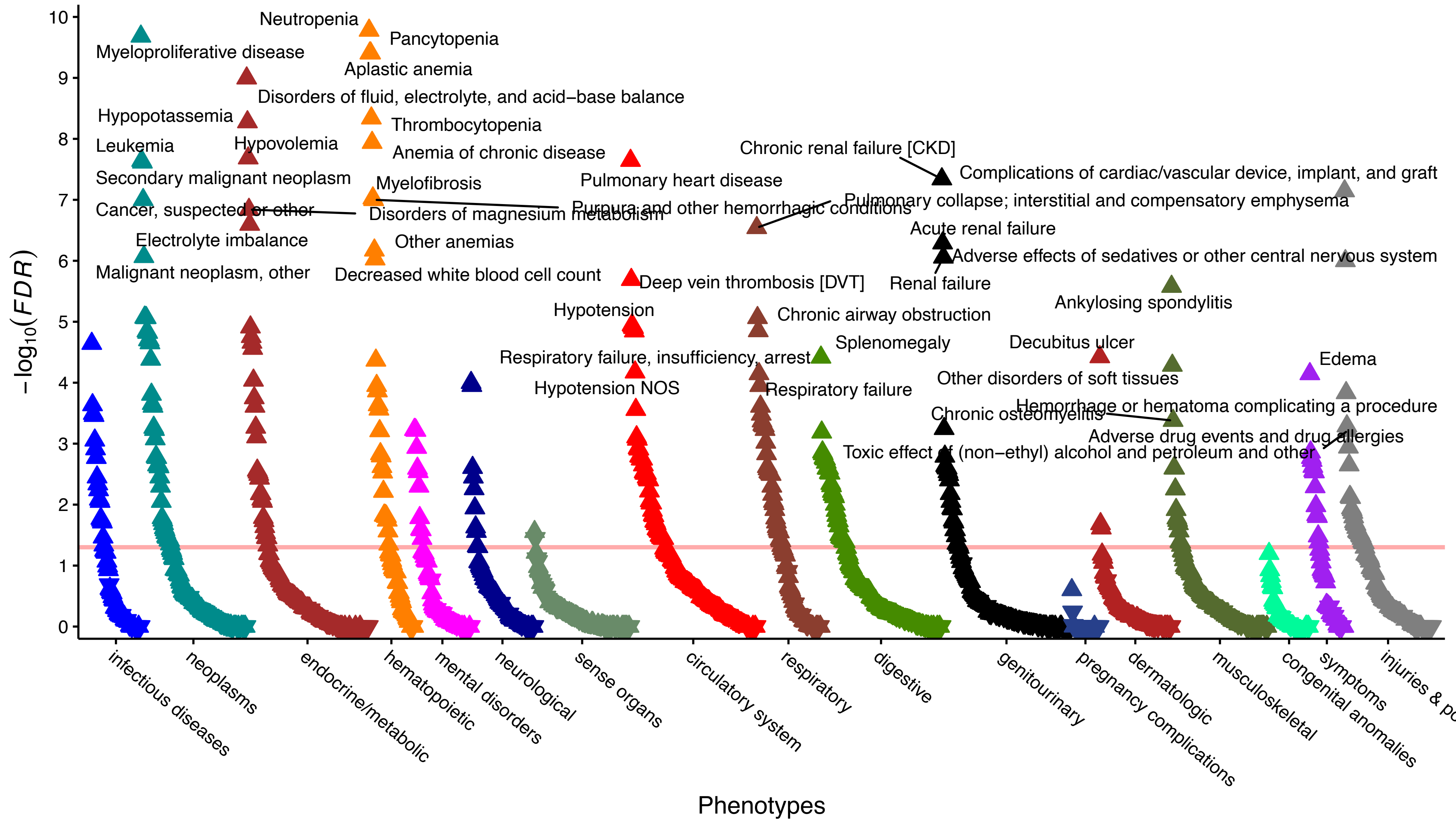

Extended Data Figure 30: PheWAS of CHIP subtypes in the All of Us Research cohort. a) PheWAS Manhattan plot for expanded CHIP (VAF>=10%); b) PheWAS Manhattan plot for *DNMT3A* CHIP; c) PheWAS Manhattan plot for *TET2* CHIP; d) PheWAS Manhattan plot for *ASXL1* CHIP; e) PheWAS Manhattan plot for *JAK2* CHIP; f) PheWAS Manhattan plot for *PPM1D* CHIP; g) PheWAS Manhattan plot for *TP53* CHIP; h) PheWAS Manhattan plot for *SF3B1* CHIP; i) PheWAS Manhattan plot for splicing factors (*SF3B1/SRSF2/U2AF1/ZRSR2*); and j) PheWAS Manhattan plot for DNA damage response genes (*TP53*/*PPM1D*). Associations between CHIP subtypes and any phecode phenotypes (i.e. prevalent and/or incident) are presented here. Multivariable adjusted logistic regression included age, age2, genetic sex, genetic ancestry, first five genetic principal components, status of smoked 100 cigarettes lifetime, batch effects for WGS sequencing center and CHIP calling. Full summary statistics for CHIP and CHIP subtypes are available in Supplementary Data Table 29.
