## Supplemental Author List for "Genomic and phenomic landscape of clonal hematopoiesis in over a million ancestrally diverse participants"

**VA Million Veteran Program**
**Core Acknowledgements for Publications**
**October 2025**

### MVP Program Office

- Sumitra Muralidhar, Ph.D., Program Director
  US Department of Veterans Affairs, 810 Vermont Avenue NW, Washington, DC 20420
- Jennifer Moser, Ph.D., Associate Director, Scientific Programs
  US Department of Veterans Affairs, 810 Vermont Avenue NW, Washington, DC 20420
- Jennifer E. Deen, B.S., Associate Director, Cohort & Public Relations
  US Department of Veterans Affairs, 810 Vermont Avenue NW, Washington, DC 20420

### MVP Steering Committee

- Co-Chair: Philip S. Tsao, Ph.D.
  VA Palo Alto Health Care System, 3801 Miranda Avenue, Palo Alto, CA 94304
- Co-Chair: Sumitra Muralidhar, Ph.D.
  US Department of Veterans Affairs, 810 Vermont Avenue NW, Washington, DC 20420
- J. Michael Gaziano, M.D., M.P.H.
  VA Boston Healthcare System, 150 S. Huntington Avenue, Boston, MA 02130
- Adriana Hung, M.D., M.P.H.,
  VA Tennessee Valley Healthcare System, 1310 24th Avenue, South Nashville, TN 37212
- Dave Oslin, M.D.
  Philadelphia VA Medical Center, 3900 Woodland Avenue, Philadelphia, PA 19104
- Deepak Voora, M.D.
  Durham VA Medical Center, 508 Fulton Street, Durham, NC 27705

### MVP Co-Principal Investigators

- J. Michael Gaziano, M.D., M.P.H.
  VA Boston Healthcare System, 150 S. Huntington Avenue, Boston, MA 02130
- Philip S. Tsao, Ph.D.
  VA Palo Alto Health Care System, 3801 Miranda Avenue, Palo Alto, CA 94304

### MVP Core Operations

- Jessica V. Brewer, M.P.H., Director, MVP Cohort Operations
  VA Boston Healthcare System, 150 S. Huntington Avenue, Boston, MA 02130
- Mary T. Brophy M.D., M.P.H., Director, VA Central Biorepository
  VA Boston Healthcare System, 150 S. Huntington Avenue, Boston, MA 02130
- Kelly Cho, M.P.H, Ph.D., Director, MVP Phenomics
  VA Boston Healthcare System, 150 S. Huntington Avenue, Boston, MA 02130
- Lori Churby, B.S., Director, MVP Regulatory Affairs
  VA Palo Alto Health Care System, 3801 Miranda Avenue, Palo Alto, CA 94304
- Jacob T. Kean, Ph.D., Acting Director, VA Informatics and Computing Infrastructure (VINCI)

VA Salt Lake City Health Care System, 500 Foothill Drive, Salt Lake City, UT 84148
- Saiju Pyarajan Ph.D., Director, Data and Computational Sciences
  VA Boston Healthcare System, 150 S. Huntington Avenue, Boston, MA 02130
- Robert Ringer, Pharm.D., Director, VA Albuquerque Central Biorepository
  New Mexico VA Health Care System, 1501 San Pedro Drive SE, Albuquerque, NM 87108
- Luis E. Selva, Ph.D., Director, MVP Biorepository Coordination
  VA Boston Healthcare System, 150 S. Huntington Avenue, Boston, MA 02130
- Shahpoor (Alex) Shayan, M.S., Director, MVP PRE Informatics
  VA Boston Healthcare System, 150 S. Huntington Avenue, Boston, MA 02130
- Brady Stephens, M.S., Principal Investigator, MVP Information Center
  Canandaigua VA Medical Center, 400 Fort Hill Avenue, Canandaigua, NY 14424
- Stacey B. Whitbourne, Ph.D., Director, MVP Cohort Development and Management
  VA Boston Healthcare System, 150 S. Huntington Avenue, Boston, MA 02130

NHLBI Trans-Omics for Precision Medicine (TOPMed) Consortium

Gonçalo Abecasis[81], Francois Aguet[82], Christine Albert[83], Laura Almasy[84], Alvaro Alonso[85], Seth Ament[86], Peter Anderson[87], Pramod Anugu[88], Kristin Ardlie[82], Dan Arking[89], Donna K Arnett[90], Allison Ashley-Koch[91], Stella Aslibekyan[92], Tim Assimes[93], Paul Auer[94], Dimitrios Avramopoulos[89], Najib Ayas[95], John Barnard[96], Kathleen Barnes[97], R. Graham Barr[98], Emily Barron-Casella[89], Terri Beaty[89], Gerald Beck[96], Diane Becker[89], Lewis Becker[89], Amber Beitelshees[86], Marcos Bezerra[99], Larry Bielak[81], Joshua Bis[87], John Blangero[100], Nathan Blue[101], Donald W. Bowden[102], Russell Bowler[103], Jennifer Brody[87], Ulrich Broeckel[104], Deborah Brown[105], Esteban Burchard[106], Carlos Bustamante[93], Brian Cade[107], Jonathan Cardwell[108], Vincent Carey[107], April P. Carson[109], Cara Carty[110], Richard Casaburi[111], James Casella[89], Peter Castaldi[107], Mark Chaffin[82], Christy Chang[86], Yi-Cheng Chang[112], Daniel Chasman[113], Wei-Min Chen[114], Yii-Der Ida Chen[115], Seung Hoan Choi[82], Lee-Ming Chuang[112], Mina Chung[96], Ren-Hua Chung[116], Clary Clish[82], Suzy Comhair[96], Matthew Conomos[87], Elaine Cornell[117], Carolyn Crandall[111], James Crapo[118], L. Adrienne Cupples[119], Joanne Curran[100], Jeffrey Curtis[81], Brian Custer[120], Coleen Damcott[86], Dawood Darbar[121], Colleen Davis[87], Michelle Daya[108], Michael DeBaun[122], Dawn DeMeo[107], Ranjan Deka[123], Scott Devine[86], Harsha Doddapaneni[124], Qing Duan[125], Shannon Dugan-Perez[126], Ravi Duggirala[127], Jon Peter Durda[117], Susan K. Dutcher[128], Charles Eaton[129], Lynette Ekunwe[88], Adel El Boueiz[130], Patrick Ellinor[131], Serpil Erzurum[96], Charles Farber[114], Tasha Fingerlin[118], Myriam Fornage[132], Nora Franceschini[125], Chris Frazar[87], Mao Fu[86], Stephanie M. Fullerton[87], Lucinda Fulton[133], Stacey Gabriel[82], Weiniu Gan[134], Shanshan Gao[108], Margery Gass[135], Heather Geiger[136], Bruce Gelb[137], Mark Geraci[138], Soren Germer[139], Robert Gerszten[140], Auyon Ghosh[107], Richard Gibbs[126], Mark Gladwin[141], David Glahn[142], Stephanie Gogarten[87], Harald Goring[143], Sharon Graw[144], Kathryn J. Gray[87], C. Charles Gu[133], Xiuqing Guo[115], Namrata Gupta[82], Jeff Haessler[135], Michael Hall[145], Patrick Hanly[146], Daniel Harris[147], Nicola L. Hawley[148], Jiang He[149], Ben Heavner[87], Ryan Hernandez[106], David Herrington[102], Craig Hersh[107], Bertha Hidalgo[92], James Hixson[132], Brian Hobbs[107], John Hokanson[108], Karin Hoth[150], Chao (Agnes) Hsiung[116], Chii Min Hwu[151], Marguerite Ryan Irvin[92], Cashell Jaquish[134], Jill Johnsen[152], Andrew Johnson[134], Craig Johnson[87], Rich Johnston[85], Kimberly Jones[89], Robert Kaplan[153], Sharon Kardia[81], Shannon Kelly[154], Eimear Kenny[137], Michael Kessler[86], Alyna Khan[87], Wonji Kim[130], Greg Kinney[155], Barbara Konkle[156], Charles Kooperberg[135], Holly Kramer[157], Christoph Lange[158], Ethan Lange[108], Cathy Laurie[87], Meryl LeBoff[107], Jonathon LeFaive[81], Wen-Jane Lee[151], Dan Levy[134], Joshua Lewis[86], Xiaohui Li[115], Yun Li[125], Henry Lin[115], Honghuang Lin[159], Xihong Lin[158], Simin Liu[160], Yongmei Liu[91], Yu Liu[93], Ruth J.F. Loos[137], Steven Lubitz[131], Kathryn Lunetta[161], James Luo[134], Ulysses Magalang[162], Michael Mahaney[100], Barry Make[89], Ani Manichaikul[114], Alisa Manning[163], JoAnn Manson[107], Lisa Martin[164], Melissa Marton[136], Susan Mathai[108], Rasika Mathias[165], Patrick McArdle[86], Merry-Lynn McDonald[92], Becky McNeil[166], Hao Mei[88], James Meigs[131], Luisa Mestroni[144], Ginger Metcalf[126], Deborah A Meyers[167], Emmanuel Mignot[168], Julie Mikulla[134], Yuan-I Min[109], Ryan L Minster[141], Braxton D. Mitchell[86], Matt Moll[107], May Montasser[169], Courtney Montgomery[170], Donna Muzny[126], Josyf C Mychaleckyj[114], Girish Nadkarni[137], Rakhi Naik[89], Pradeep Natarajan[82], Sergei

Nekhai[171], Sarah C. Nelson[87], Kari North[125], Jeff O'Connell[172], Tim O'Connor[86], Heather Ochs-Balcom[173], Allan Pack[174], Nicholette Palmer[102], James Pankow[175], Gina Peloso[176], Juan Manuel Peralta[127], Marco Perez[93], James Perry[86], Ulrike Peters[135], Patricia Peyser[81], Lawrence S Phillips[85], Toni Pollin[86], Wendy Post[177], Julia Powers Becker[108], Meher Preethi Boorgula[108], Michael Preuss[137], Bruce Psaty[87], Pankaj Qasba[178], Dandi Qiao[107], Zhaohui Qin[85], Nicholas Rafaels[179], Laura Raffield[125], D.C. Rao[133], Laura Rasmussen-Torvik[180], Aakrosh Ratan[114], Susan Redline[107], Robert Reed[86], Catherine Reeves[136], Elizabeth Regan[118], Alex Reiner[181], Rebecca Robillard[182], Nicolas Robine[136], Dan Roden[122], Carolina Roselli[82], Jerome Rotter[115], Ingo Ruczinski[89], Alexi Runnels[136], Sarah Ruuska[156], Kathleen Ryan[86], Ester Cerdeira Sabino[183], Danish Saleheen[184], Shabnam Salimi[185], Steven Salzberg[89], Kevin Sandow[115], Vijay G. Sankaran[186], Richa Saxena[187], Karen Schwander[133], David Schwartz[108], Frank Sciurba[141], Christine Seidman[188], Jonathan Seidman[188], Vivien Sheehan[189], Stephanie L. Sherman[85], Amol Shetty[86], Wayne Hui-Heng Sheu[151], M. Benjamin Shoemaker[122], Brian Silver[190], Albert Vernon Smith[191], Jennifer Smith[81], Josh Smith[87], Nicholas Smith[87], Sylvia Smoller[153], Michael Snyder[93], Tamar Sofer[192], Nona Sotoodehnia[87], Adrienne M. Stilp[87], Garrett Storm[155], Elizabeth Streeten[86], Jessica Lasky Su[107], Yun Ju Sung[133], Adam Szpiro[87], Hua Tang[93], Margaret Taub[89], Kent D. Taylor[115], Matthew Taylor[144], Simeon Taylor[86], Marilyn Telen[91], Timothy A. Thornton[87], Machiko Threlkeld[87], Lesley Tinker[135], David Tirschwell[87], Sarah Tishkoff[193], Hemant Tiwari[92], Catherine Tong[87], Russell Tracy[117], Michael Tsai[175], Dhananjay Vaidya[89], David Van Den Berg[194], Scott Vrieze[175], Tarik Walker[108], Robert Wallace[150], Heming Wang[195], Jiongming Wang[196], Karol Watson[111], Daniel E. Weeks[141], Joshua Weinstock[85], Lu-Chen Weng[131], Jennifer Wessel[197], L. Keoki Williams[198], Scott Williams[199], Carla Wilson[107], Lara Winterkorn[136], Baojun Wu[198], Joseph Wu[93], Huichun Xu[86], Lisa Yanek[89], Ivana Yang[108], Ronit Yarden[200], Seyedeh Maryam Zekavat[82], Yingze Zhang[141], Wei Zhao[81], Xiaofeng Zhu[201], Elad Ziv[106], Michael Zody[139], Mariza de Andrade[202], Paul de Vries[105], Lisa de las Fuentes[203]

81 - University of Michigan, Ann Arbor, Michigan, 48109, United States of America; 82 - Broad Institute, Cambridge, Massachusetts, 02142, United States of America; 83 - Cedars Sinai, Boston, Massachusetts, 02114, United States of America; 84 - Children's Hospital of Philadelphia, University of Pennsylvania, Philadelphia, Pennsylvania, 19104, United States of America; 85 - Emory University, Atlanta, Georgia, 30322, United States of America; 86 - University of Maryland, Baltimore, Maryland, 21201, United States of America; 87 - University of Washington, Seattle, Washington, 98195, United States of America; 88 - University of Mississippi, Jackson, Mississippi, 38677, United States of America; 89 - Johns Hopkins University, Baltimore, Maryland, 21218, United States of America; 90 - University of South Carolina, Columbia, South Carolina, 29208, United States of America; 91 - Duke University, Durham, North Carolina, 27708, United States of America; 92 - University of Alabama, Birmingham, Alabama, 35487, United States of America; 93 - Stanford University, Stanford, California, 94305, United States of America; 94 - Medical College of Wisconsin, Milwaukee, Wisconsin, 53211, United States of America; 95 - Providence Health Care, Vancouver, Canada; 96 - Cleveland Clinic, Cleveland, Ohio, 44195,

United States of America; 97 - Tempus, University of Colorado Anschutz Medical Campus, Aurora, Colorado, 80045, United States of America; 98 - Columbia University, New York, New York, 10032, United States of America; 99 - Fundação de Hematologia e Hemoterapia de Pernambuco - Hemope, Recife, 52011-000, Brazil; 100 - University of Texas Rio Grande Valley School of Medicine, Brownsville, Texas, 78520, United States of America; 101 - University of Utah, Salt Lake City, Utah, 84132, United States of America; 102 - Wake Forest Baptist Health, Winston-Salem, North Carolina, 27157, United States of America; 103 - Cleveland Clinic, Denver, Colorado, 80206, United States of America; 104 - Medical College of Wisconsin, Milwaukee, Wisconsin, 53226, United States of America; 105 - University of Texas Health at Houston, Houston, Texas, 77030, United States of America; 106 - University of California, San Francisco, San Francisco, California, 94143, United States of America; 107 - Brigham & Women's Hospital, Boston, Massachusetts, 02115, United States of America; 108 - University of Colorado at Denver, Denver, Colorado, 80204, United States of America; 109 - University of Mississippi, Jackson, Mississippi, 39213, United States of America; 110 - Washington State University, Pullman, Washington, 99164, United States of America; 111 - University of California, Los Angeles, Los Angeles, California, 90095, United States of America; 112 - National Taiwan University, Taipei, 10617, Taiwan (Province of China); 113 - Brigham & Women's Hospital, Boston, Massachusetts, 02215, United States of America; 114 - University of Virginia, Charlottesville, Virginia, 22903, United States of America; 115 - Lundquist Institute, Torrance, California, 90502, United States of America; 116 - National Health Research Institute Taiwan, Miaoli County, 350, Taiwan (Province of China); 117 - University of Vermont, Burlington, Vermont, 05405, United States of America; 118 - National Jewish Health, Denver, Colorado, 80206, United States of America; 119 - Boston University, Boston, Massachusetts, 02115, United States of America; 120 - Vitalant Research Institute, San Francisco, California, 94118, United States of America; 121 - University of Illinois at Chicago, Chicago, Illinois, 60607, United States of America; 122 - Vanderbilt University, Nashville, Tennessee, 37235, United States of America; 123 - University of Cincinnati, Cincinnati, Ohio, 45220, United States of America; 124 - Baylor College of Medicine Human Genome Sequencing Center, Houston, Texas, 77030; 125 - University of North Carolina, Chapel Hill, North Carolina, 27599, United States of America; 126 - Baylor College of Medicine Human Genome Sequencing Center, Houston, Texas, 77030, United States of America; 127 - University of Texas Rio Grande Valley School of Medicine, Edinburg, Texas, 78539, United States of America; 128 - Washington University in St Louis, St Louis, Missouri, 63110, United States of America; 129 - Brown University, Providence, Rhode Island, 02912, United States of America; 130 - Harvard University, Cambridge, Massachusetts, 02138, United States of America; 131 - Massachusetts General Hospital, Boston, Massachusetts, 02114, United States of America; 132 - University of Texas Health at Houston, Houston, Texas, 77225, United States of America; 133 - Washington University in St Louis, St Louis, Missouri, 63130, United States of America; 134 - National Heart, Lung, and Blood Institute, National Institutes of Health, Bethesda, Maryland, 20892, United States of America; 135 - Fred Hutchinson Cancer Research Center, Seattle, Washington, 98109, United States of America; 136 - New York Genome Center, New York City, New York, 10013, United

States of America; 137 - Icahn School of Medicine at Mount Sinai, New York, New York, 10029, United States of America; 138 - University of Pittsburgh, Pittsburgh, Pennsylvania, United States of America; 139 - New York Genome Center, New York, New York, 10013, United States of America; 140 - Beth Israel Deaconess Medical Center, Boston, Massachusetts, 02215, United States of America; 141 - University of Pittsburgh, Pittsburgh, Pennsylvania, 15260, United States of America; 142 - Boston Children's Hospital, Harvard Medical School, Boston, Massachusetts, 02115, United States of America; 143 - University of Texas Rio Grande Valley School of Medicine, San Antonio, Texas, 78229, United States of America; 144 - University of Colorado Anschutz Medical Campus, Aurora, Colorado, 80045, United States of America; 145 - University of Mississippi, Jackson, Mississippi, 39216, United States of America; 146 - University of Calgary, Calgary, Canada; 147 - University of Maryland, Philadelphia, Pennsylvania, 19104, United States of America; 148 - Yale University, New Haven, Connecticut, 06520, United States of America; 149 - Tulane University, New Orleans, Louisiana, 70118, United States of America; 150 - University of Iowa, Iowa City, Iowa, 52242, United States of America; 151 - Taichung Veterans General Hospital Taiwan, Taichung City, 407, Taiwan (Province of China); 152 - University of Washington, Seattle, Washington, 98109, United States of America; 153 - Albert Einstein College of Medicine, New York, New York, 10461, United States of America; 154 - University of California, San Francisco, San Francisco, California, 94118, United States of America; 155 - University of Colorado at Denver, Aurora, Colorado, 80045, United States of America; 156 - Blood Works Northwest, Seattle, Washington, 98104, United States of America; 157 - Loyola University, Maywood, Illinois, 60153, United States of America; 158 - Harvard School of Public Health, Boston, Massachusetts, 02115, United States of America; 159 - Boston University, Worcester, Massachusetts, 01655, United States of America; 160 - University of California, Irvine, Irvine, California, 92697, United States of America; 161 - Boston University, Boston, Massachusetts, 02215, United States of America; 162 - The Ohio State University, Columbus, Ohio, 43210, United States of America; 163 - Broad Institute, Harvard University, Massachusetts General Hospital; 164 - George Washington University, Washington, District of Columbia, 20037, United States of America; 165 - National Institutes of Health, Rockville, Maryland, 20852, United States of America; 166 - RTI International, United States of America; 167 - Mayo Clinic, United States of America; 168 - Stanford University, Palo Alto, California, 94304, United States of America; 169 - National Heart, Lung, and Blood Institute, Bethesda, Maryland, 20817, United States of America; 170 - Oklahoma Medical Research Foundation, Oklahoma City, Oklahoma, 73104, United States of America; 171 - Howard University, Washington, District of Columbia, 20059, United States of America; 172 - University of Maryland, Balitmore, Maryland, 21201, United States of America; 173 - University at Buffalo, Buffalo, New York, 14260, United States of America; 174 - University of Pennsylvania, Philadelphia, Pennsylvania, 19104-3403, United States of America; 175 - University of Minnesota, Minneapolis, Minnesota, 55455, United States of America; 176 - Boston University, Boston, Massachusetts, 02118, United States of America; 177 - Johns Hopkins University, Baltimore, Maryland, 21287, United States of America; 178 - National Heart, Lung, and Blood Institute, Bethesda, Maryland, 20892, United States of America; 179 - University of

Colorado at Denver, Denver, Colorado, 80045, United States of America; 180 - Northwestern University, Chicago, Illinois, 60208, United States of America; 181 - Fred Hutchinson Cancer Research Center, University of Washington, Seattle, Washington, 98109, United States of America; 182 - University of Ottawa, Ottawa, ON K1Z 7K4, Canada; 183 - Universidade de Sao Paulo, Sao Paulo, 01310000, Brazil; 184 - Columbia University, New York, New York, 10027, United States of America; 185 - University of Maryland, Seattle, Washington, 98195, United States of America; 186 - Harvard University, Boston Children's Hospital, Boston, Massachusetts, 02115, United States of America; 187 - Broad Institute, Mass General Brigham, Massachusetts General Hospital, Boston, Massachusetts, 02114, United States of America; 188 - Harvard Medical School, Boston, Massachusetts, 02115, United States of America; 189 - Emory University, Atlanta, Georgia, 30307, United States of America; 190 - UMass Memorial Medical Center, Worcester, Massachusetts, 01655, United States of America; 191 - University of Michigan; 192 - Beth Israel Deaconess Medical Center, Boston, Massachusetts, 02115, United States of America; 193 - University of Pennsylvania, Philadelphia, Pennsylvania, 19104, United States of America; 194 - University of Southern California, University of Southern California, California, 90033, United States of America; 195 - Brigham & Women's Hospital, Mass General Brigham, Boston, Massachusetts, 02115, United States of America; 196 - University of Michigan, United States of America; 197 - Indiana University, Indianapolis, Indiana, 46202, United States of America; 198 - Henry Ford Health System, Detroit, Michigan, 48202, United States of America; 199 - Case Western Reserve University; 200 - National Institutes of Health, National Heart, Lung, and Blood Institute, Bethesda, Maryland, 20892, United States of America; 201 - Case Western Reserve University, Cleveland, Ohio, 44106, United States of America; 202 - Mayo Clinic, Rochester, Minnesota, 55905, United States of America; 203 - Washington University in St Louis, St. Louis, Missouri, 63110, United States of America
